# Psychometric properties and local normative references of PSC-17, RCADS-25, CATS-2, SNAP-IV, MCHAT-R/F, and CAST: data from a nationwide sample in Greece

**DOI:** 10.1101/2025.03.21.25324416

**Authors:** André Simioni, Julia Luiza Schafer, Lauro Estivalete Marchionatti, Kenneth Schuster, Caio Borba Casella, Katerina Papanikolaou, Efstathia Kapsimalli, Panagiota Balikou, Giorgos Gerostergios, Kalliopi Triantafyllou, Maria Basta, Nikos Zilikis, Lilian Athanasopoulou, Vaios Dafoulis, Aspasia Serdari, Rafael V. S. Bastos, Peter Szatmari, Ioanna Giannopoulou, Anastasia Koumoula, Giovanni Abrahão Salum, Konstantinos Kotsis

## Abstract

Health professionals in Greece face barriers in assessing child and adolescent mental health conditions due to the lack of instruments with evidence of validity in local samples. This study addresses this gap by evaluating the psychometric properties and establishing common norms for six globally recognized mental health tools in Greece: the Child and Adolescent Trauma Screen-2 (CATS-2), Pediatric Symptoms Checklist-17 (PSC-17), Revised Children’s Anxiety and Depression Scale-25 (RCADS-25), Swanson, Nolan, and Pelham Scale (SNAP-IV), Modified Checklist for Autism in Toddlers-Revised (MCHAT-R/F), and Child Autism Spectrum Test (CAST). We drew on a nationwide Greek survey comprising 1,756 caregivers and 1,201 children and adolescents (age groups: 1 to 18 years). Using Item Response Theory, we assessed internal consistency and factor models according to Consensus-based Standards for the Selection of Health Measurement Instruments (COSMIN) criteria for unidimensionality, local independence, monotonicity, and global model fit. Normative references were calculated using standardized metrics recommended by the Patient-Reported Outcomes Measurement Information System (PROMIS). Final sample sizes ranged from 1,356 (PSC-17, caregiver version) to 198 (CATS-2, caregiver version). Internal consistency was rated as good to excellent across all scales. Factor analyses supported all scales except the MCHAT-R/F (failing monotonicity) and CAST (failing monotonicity and unidimensionality). Local normative references were usually consistent with international samples. This toolkit provides essential evidence-based resources for child and adolescent mental health in Greece, offering a scalable model for other underserved settings. Further research with national probabilistic samples is recommended to enhance risk stratification accuracy.

## Introduction

In Greece, significant hurdles for the provision of child and adolescent mental health care still persist. In the aftermath of an economic crisis, there is a shortage and unequal distribution of specialists working in the public sector as well as regional gaps in provision of care [1]. Adding on to that, referral systems and gatekeeping mechanisms are yet to be established, resulting in long waiting times in many child and adolescent mental health services. Within this context of constrained resources, it is of paramount importance to equip health professionals with tools to screen for and stratify the risk of mental health conditions in primary and pediatric care settings. Therefore, interventions and referrals could be oriented by evidence-based assessment of symptom severity [2].

Integrating psychosocial screening tools into pediatric care improves the identification and management of behavioral and emotional conditions, with several guidelines recommending the adoption of standardized core outcome measures [3–5]. In Greece, a recent systematic review highlighted that many widely-used instruments assessing child and adolescent mental health outcomes were either unavailable or lacked validation with local samples, including the Pediatric Symptom Checklist (PSC) and the Swanson, Noland, and Pelham (SNAP-IV) [6]. There was also a shortage of instruments for assessing child abuse and autism spectrum disorders, with key tools such as the Child and Adolescent Trauma Screen-2 (CATS-2), the Child Autism Spectrum Test (CAST), and the Modified Checklist for Autism in Toddlers - Revised (MCHAT-R/F) notably absent [7–10]. The absence of such tools significantly hinders best practices and research in Greece. In the case of autism spectrum disorders, it has prevented the implementation of systematic screening within the healthcare system and restricted prevalence estimates to administrative diagnostic data [11–13]

The aim of this paper is to contribute to the efforts of the Child and Adolescent Mental Health Initiative (CAMHI) in delivering evidence-based resources to enhance child and adolescent mental health care capacity in Greece. As part of this effort, we previously conducted a nationwide survey covering several patient-relevant outcomes, which employed a set of assessment instruments selected for their brevity, availability, and reliability [14]. In the present study, we report the psychometric validation and local normative references of the following instruments: the Child and Adolescent Trauma Screen-2 (CATS-2), Child Autism Spectrum Test (CAST), Modified Checklist for Autism in Toddlers-Revised (MCHAT-R/F), Pediatric Symptoms Checklist-17 (PSC-17), Revised Children’s Anxiety and Depression Scale-25 (RCADS-25), and Swanson, Nolan, and Pelham Scale (SNAP-IV).

## Methods

This study assesses the psychometric validity and local normative references of six screening instruments for child and adolescent mental health into a Greek sample (see **Table 1**). For each tool, symptom severity bands are presented with a dimensional approach (minimal, mild, moderate, and severe), ensuring user-friendly and comparable metrics aligned with standards defined by the Patient-Reported Outcomes Measurement Information System (PROMIS) group [15, 16].

**Table 1.** Instruments’ description, factor models, and proposed cutoffs.

| Domain | Instrument | Description | Informants in our sample<sup>1</sup> | Factor model Subscales | Literature proposed cut-off | Reference for Greek adaptation | Distribution |
| --- | --- | --- | --- | --- | --- | --- | --- |
| General emotional and behavior screening | Pediatric Symptoms Checklist - Shortened Version (PSC-17) [26, 61] | A 17-item measure assessing psychosocial functioning<br>Rated on a 0-to-2 Likert-scale | Caregiver Self-report | Three-factor model [61, 67]:<br>• Externalizing (7 items)<br>• Internalizing (5 items)<br>• Attention (5 items) | Attention subscale $\geq 7$<br>Internalizing subscale $\geq 5$<br>Externalizing subscale $\geq 7$<br>Total score $\geq 15$<br>[25, 61, 62] | [24] | Distributor's webpage [68] |
| Anxiety and depression | Revised Children's Anxiety and Depression Scale - Short Version (RCADS-25) [27, 69] | A 25-item scale that measures levels of low mood and anxiety.<br>Rated on a 0-to-3 Likert-scale | Self-rated<br>Caregiver-rated | Two-factor model [27, 69]:<br>• Anxiety (15 items)<br>• Depression (10 items) | T-score $< 65$: low severity<br>T-score 65 to 70: medium severity (borderline clinical threshold)<br>T-score $> 70$: high severity (above clinical threshold) [27, 69] | [59, 60] | Distributor's webpage [64] |
| Attention/hyperactivity deficit disorder | Swanson, Nolan and Pelham Scale (SNAP-IV) [37] | A 26-item instrument for hyperactivity, inattention, impulsivity, and | Caregiver-rated | Four-factor model adapted from [37]:<br>• Inattention (9 items)<br>• Hyperactivity (5 | Oppositional subscale: [27, 63, 64]<br>• $< 8$ points: Symptoms not clinically significant | [24] | CAMHI's Open Science Framework database [18] |
|  |  | oppositional behavior<br>Rated on a 0-to-4 Likert-scale |  | items)<br>• Impulsivity (4 items)<br>• Oppositionality (8 items) | • 8 to 13 points: Mild symptoms<br>• 14 to 18 points: Moderate symptoms<br>• 19 – 24 points: Severe symptoms<br>Inattention and hyperactivity/impulsivity<sup>2</sup> subscales:<br>• < 13 points: Not clinically significant<br>• 13 to 17 points: Mild symptoms<br>• 18 to 22 points: Moderate symptoms<br>• 23 to 27 points: Severe symptoms |  |  |
| Autism spectrum disorder | Modified Checklist for Autism in Toddlers - Revised (MCHAT-R/F) [7, 8] | A 20-item tool assessing neurodevelopment, social interaction, and repetitive behavior in toddlers<br>Rated on a 0-to-4 Likert-scale | Caregiver-rated | Unidimensional (20 items) [7, 8] | 0 to 2 points: low risk<br>3 to 4 points: cutoffs for risk<br>8 to 20 points: high risk [7, 8] | [70] | Distributor's webpage [71] |
| Autism spectrum disorder | Child Autism Spectrum Test (CAST) [10, 72] | A 37-items to detect manifestations of Autism Spectrum Disorders. | Caregiver-rated | Two-factor model with 28 items [34]<br>• Social contact issues (16 items)<br>• Inflexible and | ≥ 15 points:<sup>3</sup> possible autism spectrum disorder or social-communication difficulties [10, 72] | [73] | Distributor's webpage [73] |
|  |  | **Important:** we used a model with 28 items validated in Chinese that showed the best fit.<br>Yes/no questions rating 1 for each positive answer. |  | repetitive behaviors (12 items) |  |  |  |
| Trauma | Child and Adolescent Trauma Screen-2 (CATS-2) [9] | 20-item<sup>4</sup> scale assessing symptoms of Posttraumatic Stress Disorder (PTSD)<br>Rated on a 0-to-3 Likert-scale | Self-rated Caregiver | Unidimensional (20 items) [9] | ≥ 21 points: positive screening for PTSD<br>≥ 25 points: probable PTSD [9] | [24] | CAMHI's Open Science Framework database [18] |

**Abbreviations:** CAMHI (Child and Adolescent Mental Health Initiative). **Notes:** <sup>1</sup>See Koumoula et al. (2024)[14] <sup>2</sup>In our model, we separated hyperactivity/impulsivity into two subscales <sup>3</sup>This cut-off score refers to the original model; we are using the model proposed by Sun et al. (2014) [34], with cutoff scores still to be established. <sup>4</sup>CATS-2 is preceded by a 15-item checklist assessing traumatic events.

We followed the STrengthening the Reporting of OBservational studies in Epidemiology (STROBE) guidelines (see **Supplementary Table 1**) [17]. The statistical codes, outcomes, and survey dataset are openly available on our Open Science Framework page at https://osf.io/crz6h/ [18].

### Participants

We drew on data from a 2022–2023 nationwide cross-sectional survey, which included a sample of 1,201 children and adolescents (aged 8 to 17 years) and 1,756 caregivers (children aged 1 to 18 years) (detailed in Koumoula et. al (2024) [14]). Instruments were administered to subsamples according to age, group of interest, and random allocation (see **Figure 1**). Caregivers were recruited via a proprietary online respondent panel based on a census frame; invitation was delivered following an algorithm considering residency region, offspring gender, and age quotas. A first group of 400 children/adolescents consisted of the offspring of the surveyed caregivers, who were invited to an online questionnaire with self-report versions of instruments also rated by their parents. Additionally, 801 children/adolescents were recruited via random phone calls following census quotas on region and gender, providing measures of general screening and sensitive topics (namely, substance use and self-harm).

**Figure 1.**
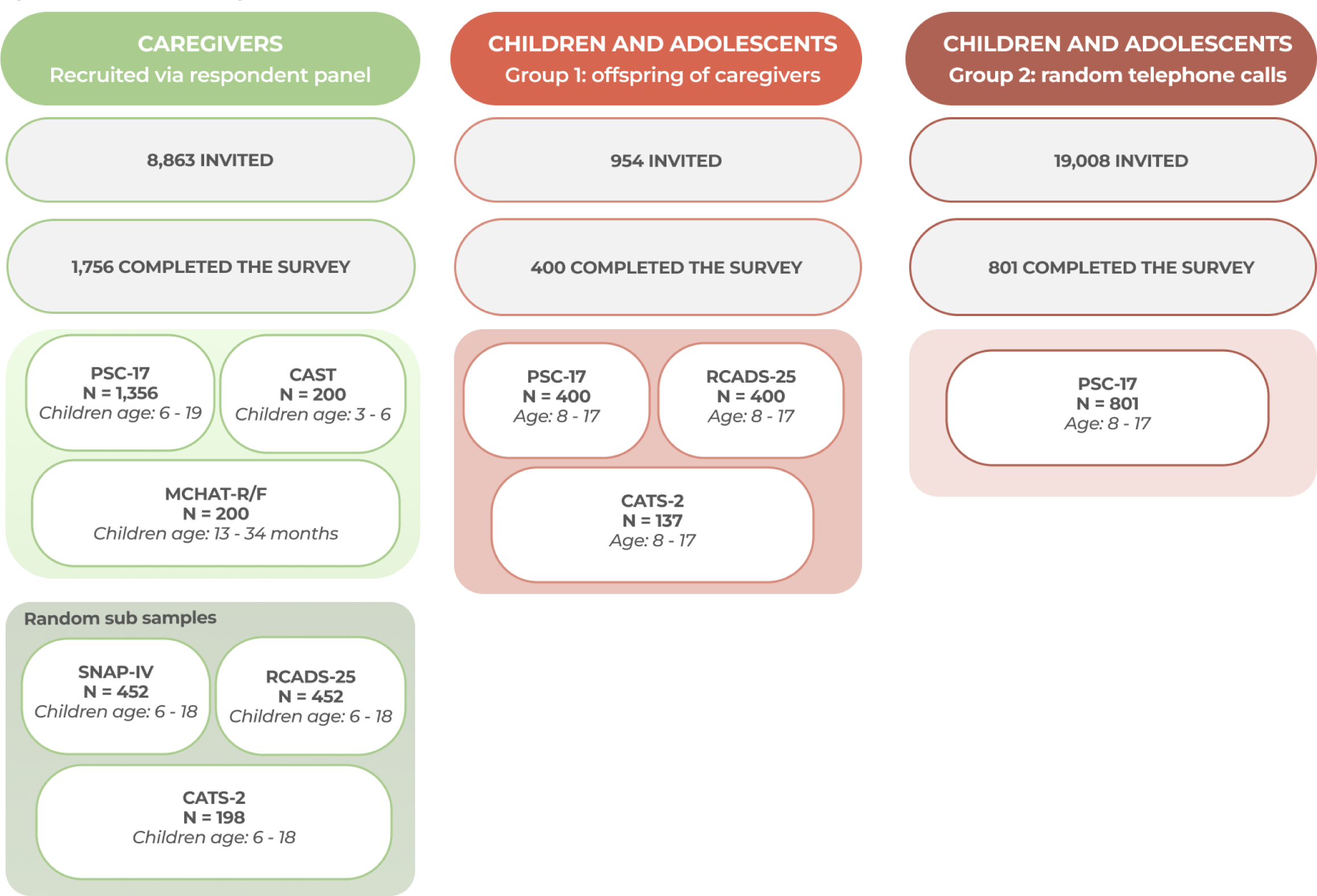
Sampling procedure. **Abbreviations:** CAST (Child Autism Spectrum Test), CATS-2 (Child and Adolescent Trauma Screen-2), MCHAT-R/F (Modified Checklist for Autism in Toddlers - Revised), PSC-17 (Pediatric Symptoms Checklist - 17 items), RCADS-25 (Revised Children’s Anxiety and Depression Scale - 25 items), SNAP-IV (Swanson, Nolan and Pelham Scale).

Data collection was managed with the KoboToolbox [19]. Informed consent and assent were respectively obtained from caregivers and youth participants. Data was collected and preserved according to the General Data Protection Regulation (GDPR) National Policy [20]. The Research Ethics Committee of the Democritus University of Thrace approved the survey [approval number: ΔΠΘ/ΕΗΔΕ/42772/307].

### Selection of instruments

Table 1 outlines the instruments used in the nationwide survey [14]. First, we consulted the International Consortium for Health Outcomes Measurement (ICHOM) on patient-relevant outcomes for child and adolescent mental health [21]. We then reviewed the literature on instruments assessing general and specific symptoms of prevalent mental health conditions. We selected tools based on their brevity, availability, and reliability, as recommended by the Consensus-based Standards for the Selection of Health Measurement Instruments (COSMIN) [22]. Three of the selected instruments (PSC-17, SNAP-IV, and CATS-2) had either not been previously translated into Greek or were not freely available in Greek language. In such cases, we performed a structured procedure for cross-cultural adaptation comprising back-and-forth translation by independent translators, synthesis of versions, expert committee appraisal, and pilot testing with the targeted population [23]; each step has been thoughtfully documented in a previous publication [24].

We employed the brief versions of the Pediatric Symptom Checklist (PSC-17) and the Revised Children’s Anxiety and Depression Scale (RCADS-25) [25–27]. Although the nationwide survey initially used the full-length versions, only the items pertaining to the shortened versions were included in the analysis [14]. For the CATS-2, we validated only the 20-item scale for symptom assessment, which is applied after screening with any positive answer on a 15-item checklist of traumatic events [9].

### Statistical analysis

An Item Response Theory (IRT) approach was conducted to test factor models, internal reliability, and to generate common metrics for each subscale or unidimensional instrument. For normative reference per age group and gender, we converted scores into percentiles, Z-scores, and T-scores, establishing a color-coded classification of symptom severity (minimal, mild, moderate, and severe) [15, 16]. Analysis was performed using the software R version 4.4.3 and the packages *lavaan*, *semTools*, *ltm*, *psych*, and *mirt* [28–33].

#### Selection of factor models

Table 1 shows the factor models tested through Confirmatory Factor Analysis (CFA). Factorial structures were consulted at developers’ distribution pages and supporting literature. For CAST, CATS-2, and SNAP-IV, more than one structure was compared to identify the best fit for our data.

As no developer-recommended structure was available for CAST, we compared models proposed in samples from Spain, Brazil, and China [34–36]. The Chinese two-factor model (“Sociability/Communication” and “Inflexible/Repetitive behaviors”) was selected for its superior fit to our data and alignment to DSM-5 criteria [34].

There are several structures for CATS-2 based on different diagnostic criteria [9]. While a hierarchical model demonstrated the best performance, there are factors with one or few items that may lead to model over rejection, and its clinical utility is limited due to complex scoring. We have chosen an unidimensional model following DSM-5 structure for its practical utility and superior fit to our data.

While the original SNAP-IV suggests a three-factor model, our data demonstrated a significantly better fit when the ‘hyperactivity/impulsivity’ factor was split into two distinct factors [37]. This four-factor structure (inattention, hyperactivity, impulsivity, and opposition) has been supported in the literature and was selected for this analysis [38].

#### Psychometric analysis

We conducted a factor analysis for each subscale or unidimensional tool, evaluating both item-level and model-level fit [32, 39, 40]. Model fit was assessed using the M2 command from the Mirt package, with estimations of the Root Mean Square Error of Approximation (RMSEA), the Standardized Root Mean-square Residual (SRMR), the Comparative Fit Index (CFI), and the Tucker-Lewis Index (TLI) [32, 40–42]. Item performance was analyzed through factor loadings, residual correlations, item scalability, monotonicity violations, fit indices, and discriminative power. Reliability was estimated using Cronbach’s alpha (α) and McDonald’s omega (ω□) [43–45].

#### Evaluation Criteria for Measurement Properties

Criteria were primarily based on the COSMIN manuals and supplemented with additional guidelines as recommended [22, 46, 47]. IRT literature prioritizes a combination of parameters to assess goodness-of-fit at item and test levels as opposed to isolated measures [40, 42, 48, 49].

Internal consistency was considered positive if Cronbach’s alpha (α) or McDonald’s omega (ω□) exceeded 0.7 [22, 43, 50, 51]. Values above 0.8 and 0.9 respectively indicated good and excellent reliability.

Factor analysis was deemed positive if the following criteria were met: 1) unidimensionality (at least two of the following: RMSEA ≤ 0.06, SRMR ≤ 0.08, and TLI/CFI ≥ 0.95; RMSEA ≤ 0.08 is also considered acceptable [47]); 2) local independence (residual correlations among items < 0.2 OR the third quartile of correlations < 0.37); 3) monotonicity (item scalability > 0.3 OR adequate looking graphs); and 4) global model fit (unidimensionality criteria attended AND item infit mean squares ≥ 0.5 and ≤ 1.5).

Additionally, factor loadings above 0.3 were considered positive, with values higher than 0.5 classified as very positive [52, 53]. For monotonicity, we also evaluated the number of violations per item. The absence of violations indicated adequate monotonicity, and violations with critical values between 40 and 90 were also considered acceptable [39, 54].

#### Local normative references

Graded Response Models (GRMs) and Graded Rating Scale Models (GRSMs) were employed for parameterization, assigning an IRT score to each participant [41, 55, 56]. Scores were categorized by age group and gender, with T-scores calculated from factor scores (a T-score of 50 represents the reference population mean, and a 10-point difference corresponds to one standard deviation). Symptom level bands followed PROMIS recommendations for standardized metrics, consisting of four ranges: minimal symptoms (T-score < 55), mild symptoms (T-score 55–59), moderate symptoms (T-score 60–69), and severe symptoms (T-score ≥ 70) [16, 57, 58]. Scores were then rescaled according to the range of each T-score-based severity category, with values truncated within each band. Finally, crude scores were linked to IRT-based scores by grouping factor, T-scores, Z-scores, and percentiles for each summed score value.

## Results

**Table 2** presents the sociodemographic characteristics of caregivers and children/adolescents who completed each instrument. The PSC-17 (proxy-report, school-age children) had the largest sample size, including all 1,356 caregivers of 6- to 18-year-olds participating in the survey. Some instruments were administered to subsamples, with participant numbers ranging from 452 (RCADS-25, caregiver-rated; SNAP-IV, caregiver-rated) to 137 (CATS-2, self-report).

**Table 2.**
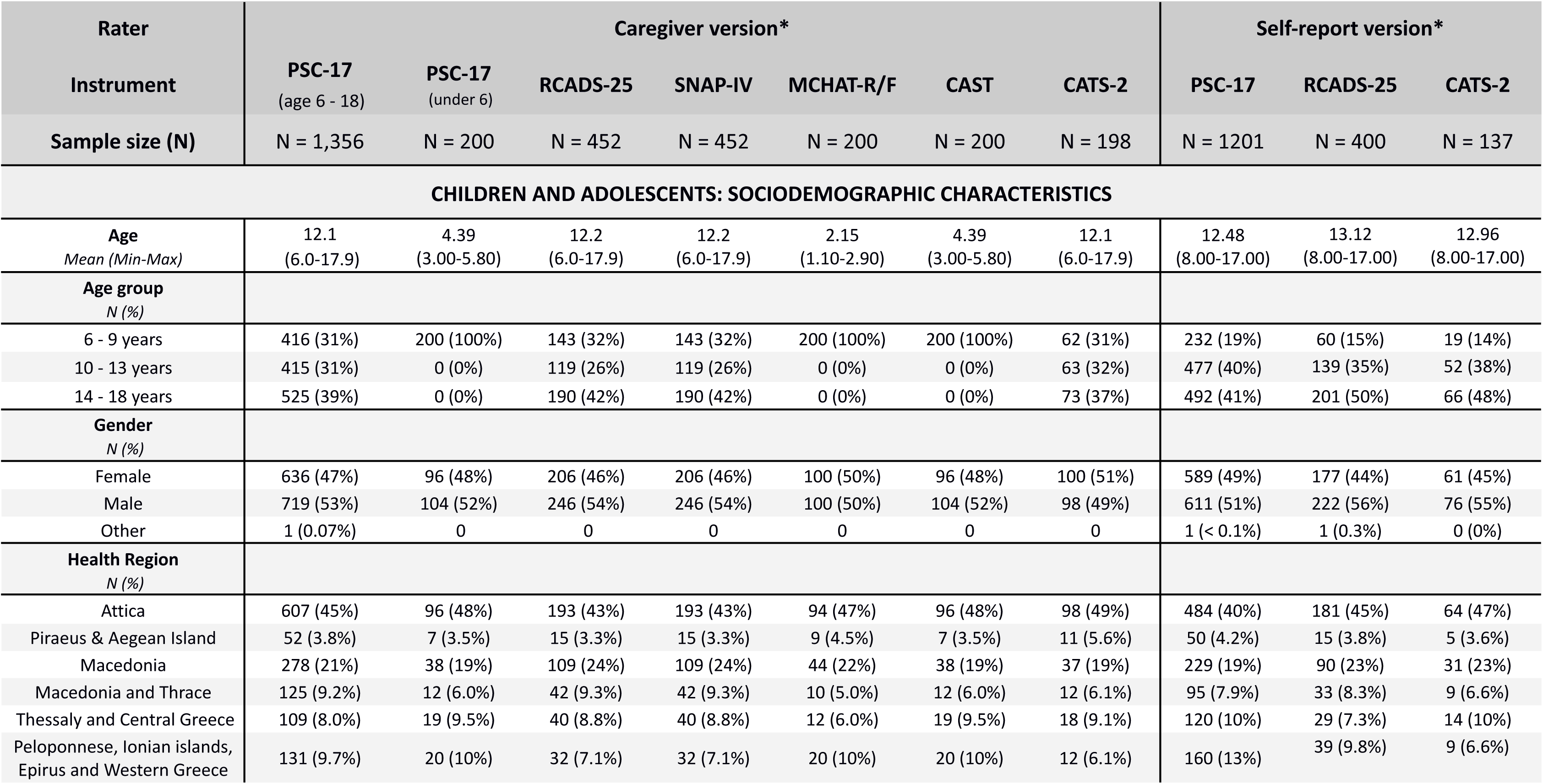
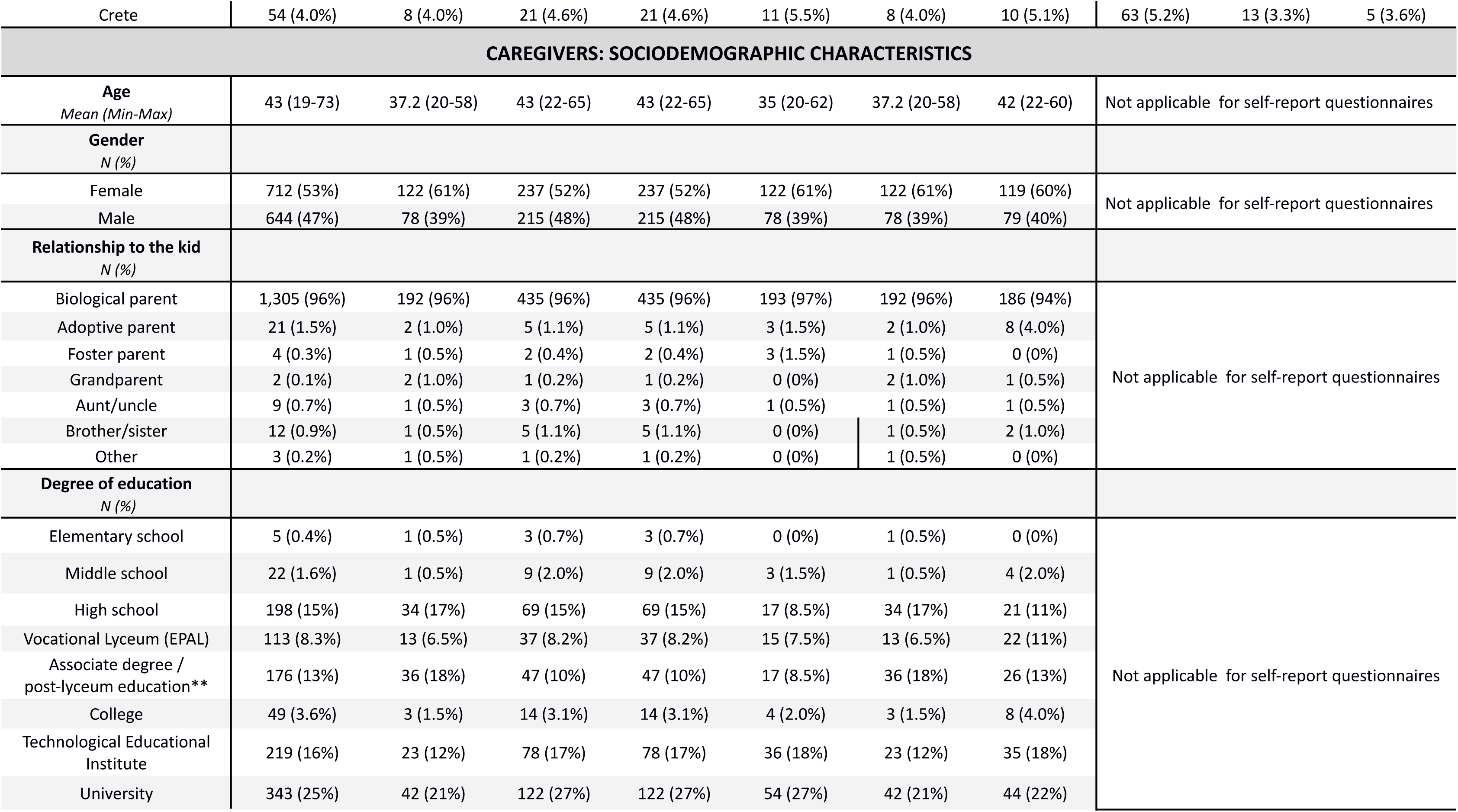
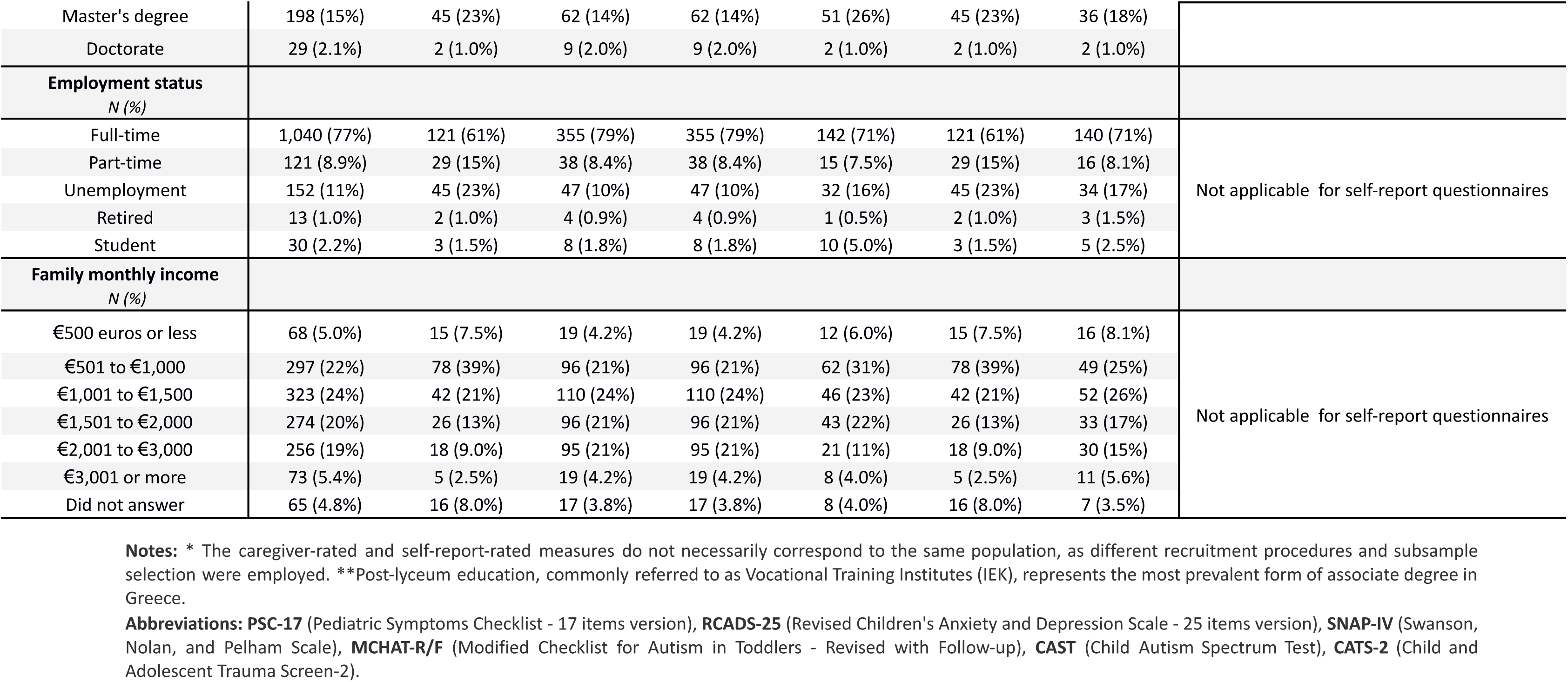
Sociodemographic characteristics of participants.

| Rater | Caregiver version* |  |  |  |  |  |  | Self-report version* |  |  |
| --- | --- | --- | --- | --- | --- | --- | --- | --- | --- | --- |
| Instrument | PSC-17<br>(age 6 - 18) | PSC-17<br>(under 6) | RCADS-25 | SNAP-IV | MCHAT-R/F | CAST | CATS-2 | PSC-17 | RCADS-25 | CATS-2 |
| Sample size (N) | N = 1,356 | N = 200 | N = 452 | N = 452 | N = 200 | N = 200 | N = 198 | N = 1201 | N = 400 | N = 137 |
| CHILDREN AND ADOLESCENTS: SOCIODEMOGRAPHIC CHARACTERISTICS |  |  |  |  |  |  |  |  |  |  |
| Age<br>*Mean (Min-Max)* | 12.1<br>(6.0-17.9) | 4.39<br>(3.00-5.80) | 12.2<br>(6.0-17.9) | 12.2<br>(6.0-17.9) | 2.15<br>(1.10-2.90) | 4.39<br>(3.00-5.80) | 12.1<br>(6.0-17.9) | 12.48<br>(8.00-17.00) | 13.12<br>(8.00-17.00) | 12.96<br>(8.00-17.00) |
| Age group<br>*N (%)* |  |  |  |  |  |  |  |  |  |  |
| 6 - 9 years | 416 (31%) | 200 (100%) | 143 (32%) | 143 (32%) | 200 (100%) | 200 (100%) | 62 (31%) | 232 (19%) | 60 (15%) | 19 (14%) |
| 10 - 13 years | 415 (31%) | 0 (0%) | 119 (26%) | 119 (26%) | 0 (0%) | 0 (0%) | 63 (32%) | 477 (40%) | 139 (35%) | 52 (38%) |
| 14 - 18 years | 525 (39%) | 0 (0%) | 190 (42%) | 190 (42%) | 0 (0%) | 0 (0%) | 73 (37%) | 492 (41%) | 201 (50%) | 66 (48%) |
| Gender<br>*N (%)* |  |  |  |  |  |  |  |  |  |  |
| Female | 636 (47%) | 96 (48%) | 206 (46%) | 206 (46%) | 100 (50%) | 96 (48%) | 100 (51%) | 589 (49%) | 177 (44%) | 61 (45%) |
| Male | 719 (53%) | 104 (52%) | 246 (54%) | 246 (54%) | 100 (50%) | 104 (52%) | 98 (49%) | 611 (51%) | 222 (56%) | 76 (55%) |
| Other | 1 (0.07%) | 0 | 0 | 0 | 0 | 0 | 0 | 1 (< 0.1%) | 1 (0.3%) | 0 (0%) |
| Health Region<br>*N (%)* |  |  |  |  |  |  |  |  |  |  |
| Attica | 607 (45%) | 96 (48%) | 193 (43%) | 193 (43%) | 94 (47%) | 96 (48%) | 98 (49%) | 484 (40%) | 181 (45%) | 64 (47%) |
| Piraeus & Aegean Island | 52 (3.8%) | 7 (3.5%) | 15 (3.3%) | 15 (3.3%) | 9 (4.5%) | 7 (3.5%) | 11 (5.6%) | 50 (4.2%) | 15 (3.8%) | 5 (3.6%) |
| Macedonia | 278 (21%) | 38 (19%) | 109 (24%) | 109 (24%) | 44 (22%) | 38 (19%) | 37 (19%) | 229 (19%) | 90 (23%) | 31 (23%) |
| Macedonia and Thrace | 125 (9.2%) | 12 (6.0%) | 42 (9.3%) | 42 (9.3%) | 10 (5.0%) | 12 (6.0%) | 12 (6.1%) | 95 (7.9%) | 33 (8.3%) | 9 (6.6%) |
| Thessaly and Central Greece | 109 (8.0%) | 19 (9.5%) | 40 (8.8%) | 40 (8.8%) | 12 (6.0%) | 19 (9.5%) | 18 (9.1%) | 120 (10%) | 29 (7.3%) | 14 (10%) |
| Peloponnese, Ionian islands,<br>Epirus and Western Greece | 131 (9.7%) | 20 (10%) | 32 (7.1%) | 32 (7.1%) | 20 (10%) | 20 (10%) | 12 (6.1%) | 160 (13%) | 39 (9.8%) | 9 (6.6%) |
| Crete | 54 (4.0%) | 8 (4.0%) | 21 (4.6%) | 21 (4.6%) | 11 (5.5%) | 8 (4.0%) | 10 (5.1%) | 63 (5.2%) | 13 (3.3%) | 5 (3.6%) |
| **CAREGIVERS: SOCIODEMOGRAPHIC CHARACTERISTICS** |  |  |  |  |  |  |  |  |  |  |
| **Age**<br>*Mean (Min-Max)* | 43 (19-73) | 37.2 (20-58) | 43 (22-65) | 43 (22-65) | 35 (20-62) | 37.2 (20-58) | 42 (22-60) | Not applicable for self-report questionnaires |  |  |
| **Gender**<br>*N (%)* |  |  |  |  |  |  |  |  |  |  |
| Female | 712 (53%) | 122 (61%) | 237 (52%) | 237 (52%) | 122 (61%) | 122 (61%) | 119 (60%) | Not applicable for self-report questionnaires |  |  |
| Male | 644 (47%) | 78 (39%) | 215 (48%) | 215 (48%) | 78 (39%) | 78 (39%) | 79 (40%) |  |  |  |
| **Relationship to the kid**<br>*N (%)* |  |  |  |  |  |  |  |  |  |  |
| Biological parent | 1,305 (96%) | 192 (96%) | 435 (96%) | 435 (96%) | 193 (97%) | 192 (96%) | 186 (94%) | Not applicable for self-report questionnaires |  |  |
| Adoptive parent | 21 (1.5%) | 2 (1.0%) | 5 (1.1%) | 5 (1.1%) | 3 (1.5%) | 2 (1.0%) | 8 (4.0%) |  |  |  |
| Foster parent | 4 (0.3%) | 1 (0.5%) | 2 (0.4%) | 2 (0.4%) | 3 (1.5%) | 1 (0.5%) | 0 (0%) |  |  |  |
| Grandparent | 2 (0.1%) | 2 (1.0%) | 1 (0.2%) | 1 (0.2%) | 0 (0%) | 2 (1.0%) | 1 (0.5%) |  |  |  |
| Aunt/uncle | 9 (0.7%) | 1 (0.5%) | 3 (0.7%) | 3 (0.7%) | 1 (0.5%) | 1 (0.5%) | 1 (0.5%) |  |  |  |
| Brother/sister | 12 (0.9%) | 1 (0.5%) | 5 (1.1%) | 5 (1.1%) | 0 (0%) | 1 (0.5%) | 2 (1.0%) |  |  |  |
| Other | 3 (0.2%) | 1 (0.5%) | 1 (0.2%) | 1 (0.2%) | 0 (0%) | 1 (0.5%) | 0 (0%) |  |  |  |
| **Degree of education**<br>*N (%)* |  |  |  |  |  |  |  |  |  |  |
| Elementary school | 5 (0.4%) | 1 (0.5%) | 3 (0.7%) | 3 (0.7%) | 0 (0%) | 1 (0.5%) | 0 (0%) | Not applicable for self-report questionnaires |  |  |
| Middle school | 22 (1.6%) | 1 (0.5%) | 9 (2.0%) | 9 (2.0%) | 3 (1.5%) | 1 (0.5%) | 4 (2.0%) |  |  |  |
| High school | 198 (15%) | 34 (17%) | 69 (15%) | 69 (15%) | 17 (8.5%) | 34 (17%) | 21 (11%) |  |  |  |
| Vocational Lyceum (EPAL) | 113 (8.3%) | 13 (6.5%) | 37 (8.2%) | 37 (8.2%) | 15 (7.5%) | 13 (6.5%) | 22 (11%) |  |  |  |
| Associate degree /<br>post-lyceum education** | 176 (13%) | 36 (18%) | 47 (10%) | 47 (10%) | 17 (8.5%) | 36 (18%) | 26 (13%) |  |  |  |
| College | 49 (3.6%) | 3 (1.5%) | 14 (3.1%) | 14 (3.1%) | 4 (2.0%) | 3 (1.5%) | 8 (4.0%) |  |  |  |
| Technological Educational<br>Institute | 219 (16%) | 23 (12%) | 78 (17%) | 78 (17%) | 36 (18%) | 23 (12%) | 35 (18%) |  |  |  |
| University | 343 (25%) | 42 (21%) | 122 (27%) | 122 (27%) | 54 (27%) | 42 (21%) | 44 (22%) |  |  |  |

|  |  |  |  |  |  |  |  |  |
| --- | --- | --- | --- | --- | --- | --- | --- | --- |
| Master's degree | 198 (15%) | 45 (23%) | 62 (14%) | 62 (14%) | 51 (26%) | 45 (23%) | 36 (18%) |  |
| Doctorate | 29 (2.1%) | 2 (1.0%) | 9 (2.0%) | 9 (2.0%) | 2 (1.0%) | 2 (1.0%) | 2 (1.0%) |  |
| **Employment status**<br>*N (%)* |  |  |  |  |  |  |  |  |
| Full-time | 1,040 (77%) | 121 (61%) | 355 (79%) | 355 (79%) | 142 (71%) | 121 (61%) | 140 (71%) | Not applicable for self-report questionnaires |
| Part-time | 121 (8.9%) | 29 (15%) | 38 (8.4%) | 38 (8.4%) | 15 (7.5%) | 29 (15%) | 16 (8.1%) |  |
| Unemployment | 152 (11%) | 45 (23%) | 47 (10%) | 47 (10%) | 32 (16%) | 45 (23%) | 34 (17%) |  |
| Retired | 13 (1.0%) | 2 (1.0%) | 4 (0.9%) | 4 (0.9%) | 1 (0.5%) | 2 (1.0%) | 3 (1.5%) |  |
| Student | 30 (2.2%) | 3 (1.5%) | 8 (1.8%) | 8 (1.8%) | 10 (5.0%) | 3 (1.5%) | 5 (2.5%) |  |
| **Family monthly income**<br>*N (%)* |  |  |  |  |  |  |  |  |
| €500 euros or less | 68 (5.0%) | 15 (7.5%) | 19 (4.2%) | 19 (4.2%) | 12 (6.0%) | 15 (7.5%) | 16 (8.1%) | Not applicable for self-report questionnaires |
| €501 to €1,000 | 297 (22%) | 78 (39%) | 96 (21%) | 96 (21%) | 62 (31%) | 78 (39%) | 49 (25%) |  |
| €1,001 to €1,500 | 323 (24%) | 42 (21%) | 110 (24%) | 110 (24%) | 46 (23%) | 42 (21%) | 52 (26%) |  |
| €1,501 to €2,000 | 274 (20%) | 26 (13%) | 96 (21%) | 96 (21%) | 43 (22%) | 26 (13%) | 33 (17%) |  |
| €2,001 to €3,000 | 256 (19%) | 18 (9.0%) | 95 (21%) | 95 (21%) | 21 (11%) | 18 (9.0%) | 30 (15%) |  |
| €3,001 or more | 73 (5.4%) | 5 (2.5%) | 19 (4.2%) | 19 (4.2%) | 8 (4.0%) | 5 (2.5%) | 11 (5.6%) |  |
| Did not answer | 65 (4.8%) | 16 (8.0%) | 17 (3.8%) | 17 (3.8%) | 8 (4.0%) | 16 (8.0%) | 7 (3.5%) |  |

**Notes:** \* The caregiver-rated and self-report-rated measures do not necessarily correspond to the same population, as different recruitment procedures and subsample selection were employed. \*\*Post-lyceum education, commonly referred to as Vocational Training Institutes (IEK), represents the most prevalent form of associate degree in Greece.
**Abbreviations:** **PSC-17** (Pediatric Symptoms Checklist - 17 items version), **RCADS-25** (Revised Children's Anxiety and Depression Scale - 25 items version), **SNAP-IV** (Swanson, Nolan, and Pelham Scale), **MCHAT-R/F** (Modified Checklist for Autism in Toddlers - Revised with Follow-up), **CAST** (Child Autism Spectrum Test), **CATS-2** (Child and Adolescent Trauma Screen-2).

**Table 3** summarizes the psychometric properties of each tool. **Table 4** provides normative references for PSC-17 (self-report). Standardized scores and classification bands for the remaining instruments are detailed in **Supplementary Table 2** through **Supplementary Table 10**. Test information curves, expected total score curves, item probability functions, and item and person infit and outfit statistics for each instrument’s subscale can be consulted in **Supplementary Figure 1.1.1** to **Supplementary Figure 10.4.4**. Comprehensive documentation of codes and measurement properties is also made accessible through our Open Science Framework repository (https://osf.io/crz6h/) [18].

**Table 3.** Psychometric properties of the instruments.

| Instrument - Rater<br>Subscale *[items]* | Items | Sample<br>(N) | $\alpha$ | $\omega$ | Item Scalability<br>(Min - Max) | Violations<br>of<br>monotonicity | Factor<br>loading<br>(Min - Max) | Residual<br>Correlations Among<br>Items<br>3<sup>rd</sup> quartile [Min;<br>Max] | RMSEA | TLI | CFI | SRMR | Item infit<br>mean<br>squares<br>(Min -<br>Max) |
| --- | --- | --- | --- | --- | --- | --- | --- | --- | --- | --- | --- | --- | --- |
| **Pediatric Symptom Checklist - 17 (PSC-17) -**<br>**Caregiver - 6 to 18 years** | 17 | 1,356 |  |  |  |  |  |  |  |  |  |  |  |
| Externalizing *[4, 5, 8, 10, 12, 14, 16]* | 7 |  | 0.89 | 0.89 | 0.41 - 0.57 | 1<sup>a</sup> | 0.65 - 0.81 | 0.05 [-0.10; 0.09] | 0.06 | 0.98 | 0.99 | 0.04 | 0.8 - 0.9 |
| Internalizing *[2, 6, 9, 11, 15]* | 5 |  | 0.89 | 0.89 | 0.5 - 0.6 | 0 | 0.75 - 0.85 | 0.04 [-0.06; 0.08] | 0.04 | 0.99 | 1.00 | 0.02 | 0.8 - 0.9 |
| Attention *[1, 3, 7, 13, 17]* | 5 |  | 0.87 | 0.87 | 0.44 - 0.61 | 0 | 0.57 - 0.93 | 0.05 [-0.10; 0.14] | 0.12 | 0.92 | 0.96 | 0.07 | 0.6 - 0.9 |
| **Pediatric Symptom Checklist - 17 (PSC - 17) -**<br>**Caregiver - 3 to 5 years** | 17 | 200 |  |  |  |  |  |  |  |  |  |  |  |
| Externalizing *[4, 5, 8, 10, 12, 14, 16]* | 7 |  | 0.84 | 0.85 | 0.37 - 0.49 | 0 | 0.58 - 0.9 | 0.12 [-0.22; 0.17]<sup>b</sup> | 0.09 | 0.93 | 0.95 | 0.07 | 0.6 - 0.9 |
| Internalizing *[2, 6, 9, 11, 15]* | 5 |  | 0.89 | 0.89 | 0.45 - 0.5 | 0 | 0.7 - 0.85 | 0.09 [-0.12; 0.13] | 0 | 1 | 1 | 0.05 | 0.9 - 1.0 |
| Attention *[1,3,7,13,17]* | 5 |  | 0.84 | 0.84 | 0.42 - 0.56 | 0 | 0.59 - 0.86 | 0.08 [-0.22; 0.16]<sup>c</sup> | 0.05 | 0.98 | 0.98 | 0.06 | 0.7 - 0.9 |
| **Pediatric Symptom Checklist (PSC) -**<br>**Self-report** | 17 | 400 |  |  |  |  |  |  |  |  |  |  |  |
| Externalizing *[4, 5, 8, 10, 12, 14, 16]* | 7 |  | 0.85 | 0.85 | 0.3 - 0.42 | 0 | 0.52 - 0.8 | 0.05 [-0.07; 0.08] | 0.03 | 0.99 | 0.99 | 0.03 | 0.9 - 1.0 |
| Internalizing *[2, 6, 9, 11, 15]* | 5 |  | 0.88 | 0.88 | 0.46 - 0.56 | 0 | 0.68 - 0.83 | 0.04 [-0.08; 0.09] | 0.04 | 0.99 | 1.0 | 0.03 | 0.8 - 0.9 |
| Attention *[1,3,7,13,17]* | 5 |  | 0.83 | 0.84 | 0.36 - 0.51 | 0 | 0.56 - 0.88 | 0.08 [-0.1; 0.12] | 0.07 | 0.96 | 0.97 | 0.05 | 0.6 - 1.0 |
| **Revised Children's Anxiety and Depression Scale - (RCADS-25) - Caregiver** | 25 | 452 |  |  |  |  |  |  |  |  |  |  |  |
| Anxiety [2, 3, 5, 6, 7, 9, 11, 12, 14, 17, 18, 20, 22, 23, 25] | 15 |  | 0.94 | 0.94 | 0.31 - 0.52 | 0 | 0.45 - 0.89 | 0.10 [-0.14; 0.36]<sup>d</sup> | 0.09 | 0.95 | 0.96 | 0.08 | 0.9 - 1.0 |
| Depression [1, 4, 8, 10, 13, 15, 16, 19, 21, 24] | 10 |  | 0.94 | 0.94 | 0.41 - 0.59 | 0 | 0.64 - 0.9 | 0.07 [-0.14; 0.13] | 0.04 | 0.99 | 0.99 | 0.05 | 0.9 - 1.1 |
| **Revised Children's Anxiety and Depression Scale - (RCADS-25) - Self-report** | 25 | 400 |  |  |  |  |  |  |  |  |  |  |  |
| Anxiety [2, 3, 5, 6, 7, 9, 11, 12, 14, 17, 18, 20, 22, 23, 25] | 15 |  | 0.97 | 0.97 | 0.5 - 0.64 | 0 | 0.71 - 0.94 | 0.08 [-0.16; 0.19] | 0.08 | 0.98 | 0.98 | 0.07 | 0.9 - 1.0 |
| Depression [1, 4, 8, 10, 13, 15, 16, 19, 21, 24] | 10 |  | 0.97 | 0.97 | 0.59 - 0.69 | 0 | 0.76 - 0.92 | 0.09 [-0.15; 0.14] | 0.03 | 0.99 | 0.99 | 0.05 | 0.9 - 1.0 |
| **Swanson, Nolan and Pelham Scale (SNAP-IV) - Caregiver** | 26 | 452 |  |  |  |  |  |  |  |  |  |  |  |
| Inattention [1 - 9] | 9 |  | 0.95 | 0.95 | 0.57 - 0.69 | 0 | 0.76 - 0.91 | -0.03 [-0.17; 0.14] | 0.06 | 0.99 | 0.99 | 0.03 | 0.8 - 0.9 |
| Hyperactivity [10 - 14] | 5 |  | 0.94 | 0.94 | 0.63 - 0.7 | 0 | 0.79 - 0.91 | 0.07 [-0.08; 0.09] | 0.02 | 1 | 1 | 0.02 | 0.8 - 1.0 |
| Impulsivity [15 - 18] | 4 |  | 0.91 | 0.91 | 0.63 - 0.69 | 0 | 0.8 - 0.91 | -0.11 [-0.19; -0.09] | 0.02 | 1 | 1 | 0.02 | 0.7 - 0.9 |
| Oppositional [19 - 26] | 8 |  | 0.95 | 0.96 | 0.65 - 0.69 | 0 | 0.8 - 0.91 | 0.1 [-0.12; 0.14] | 0.08 | 0.98 | 0.98 | 0.05 | 0.9 - 1.0 |
| **Modified Checklist for Autism in Toddlers - Revised (MCHAT-R/F) - Caregiver** | 20 | 200 |  |  |  |  |  |  |  |  |  |  |  |
| Unidimensional [1-20] | 20 |  | 0.92 | 0.93 | -0.01<sup>e</sup> - 0.43 | 0 | -0.39 - 0.94 | 0.04 [-0.19; 0.28]<sup>f</sup> | 0.02 | 0.99 | 0.99 | 0.08 | 1.0 - 1.3 |
| **Child Autism Spectrum Test (CAST) - Caregiver** | 28 | 200 |  |  |  |  |  |  |  |  |  |  |  |
| Social contact issues [1, 2, 5, 8, 10, 11, 13, 15, 16, 17, 21, 23, 24, 27, 31, 35] | 16 |  | 0.88 | 0.88 | 0.11<sup>g</sup> - 0.37 | 0 | 0.18<sup>h</sup> - 0.99 | 0.03 [-0.16; 0.23]<sup>i</sup> | 0.06 | 0.9 | 0.9 | 0.09 | 0.4<sup>j</sup> - 1.2 |
| Inflexible and repetitive behaviors [7, 9, 18, 20, 25, 28, 29, 30, 32, 34, 36, 37] | 12 |  | 0.92 | 0.92 | 0.22<sup>k</sup> - 0.61 | 0 | 0.34 - 0.91 | 0.06 [-0.12; 0.15] | 0 | 1 | 1 | 0.06 | 0.9 - 1.0 |
| **Child and Adolescent Trauma Screen-2 (CATS-2) - Caregiver** | 20 | 198 |  |  |  |  |  |  |  |  |  |  |  |
| Unidimensional scale for PTSD symptoms [16 - 23, 24 (a - d), 25 (a - b), 26 - 29, 30 (a - b), 31 - 35] |  |  | 0.95 | 0.95 | 0.41 - 0.57 | 0 | 0.61 - 0.87 | 0.11 [-0.23; 0.3]<sup>l</sup> | 0.09 | 0.96 | 0.96 | 0.07 | 0.9 - 1.1 |
| **Child and Adolescent Trauma Screen-2 (CATS-2) - Self-report** | 20 | 137 |  |  |  |  |  |  |  |  |  |  |  |
| Unidimensional [16- 23, 24 (a - d), 25 (a - b), 26 - 29, 30 (a - b), 31 - 35] | 20 |  | 0.97 | 0.97 | 0.4 - 0.64 | 0 | 0.56 - 0.92 | 0.13 [-0.26; 0.25]<sup>m</sup> | 0.07 | 0.98 | 0.98 | 0.06 | 0.9 - 1.1 |

**Table 4.**
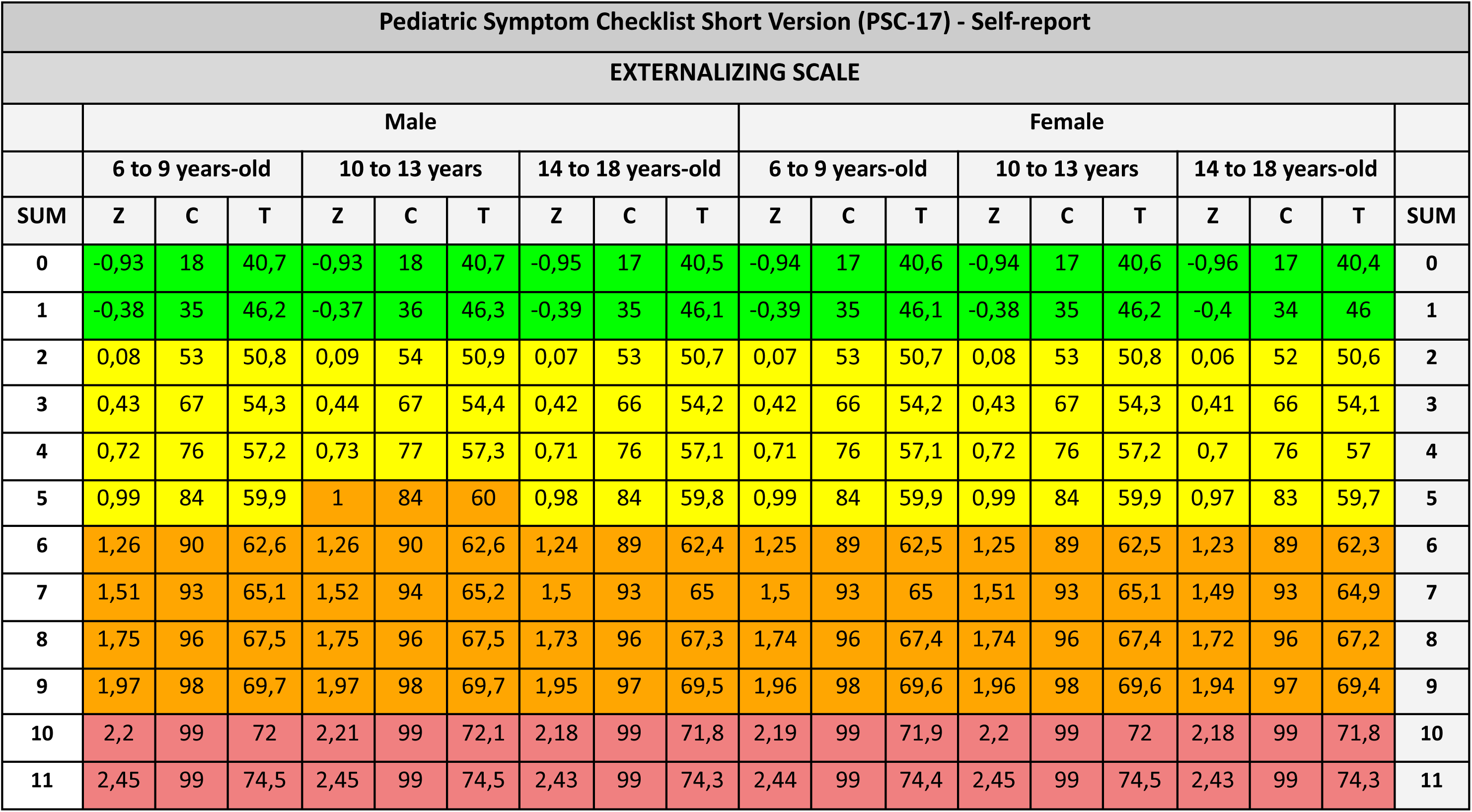
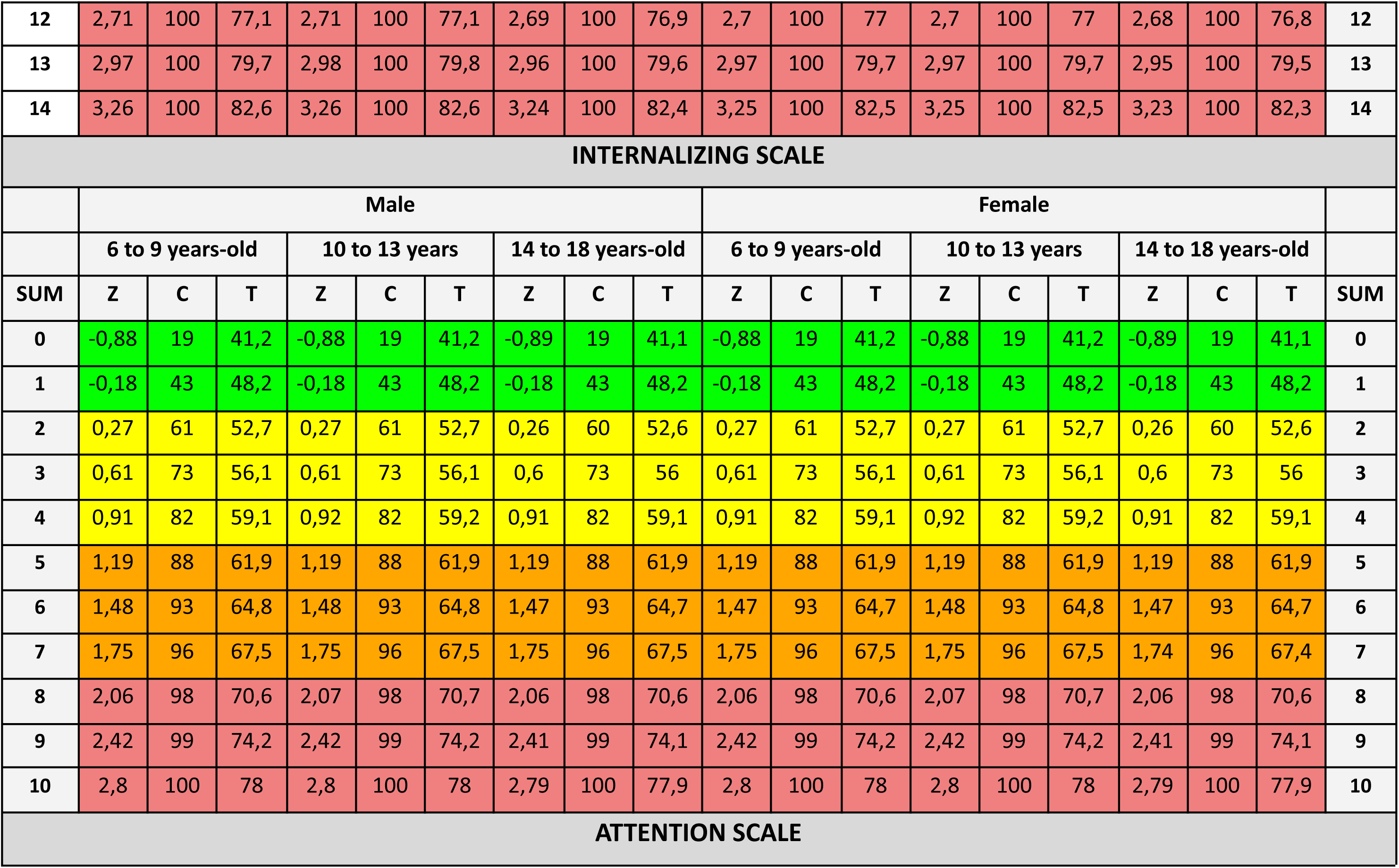
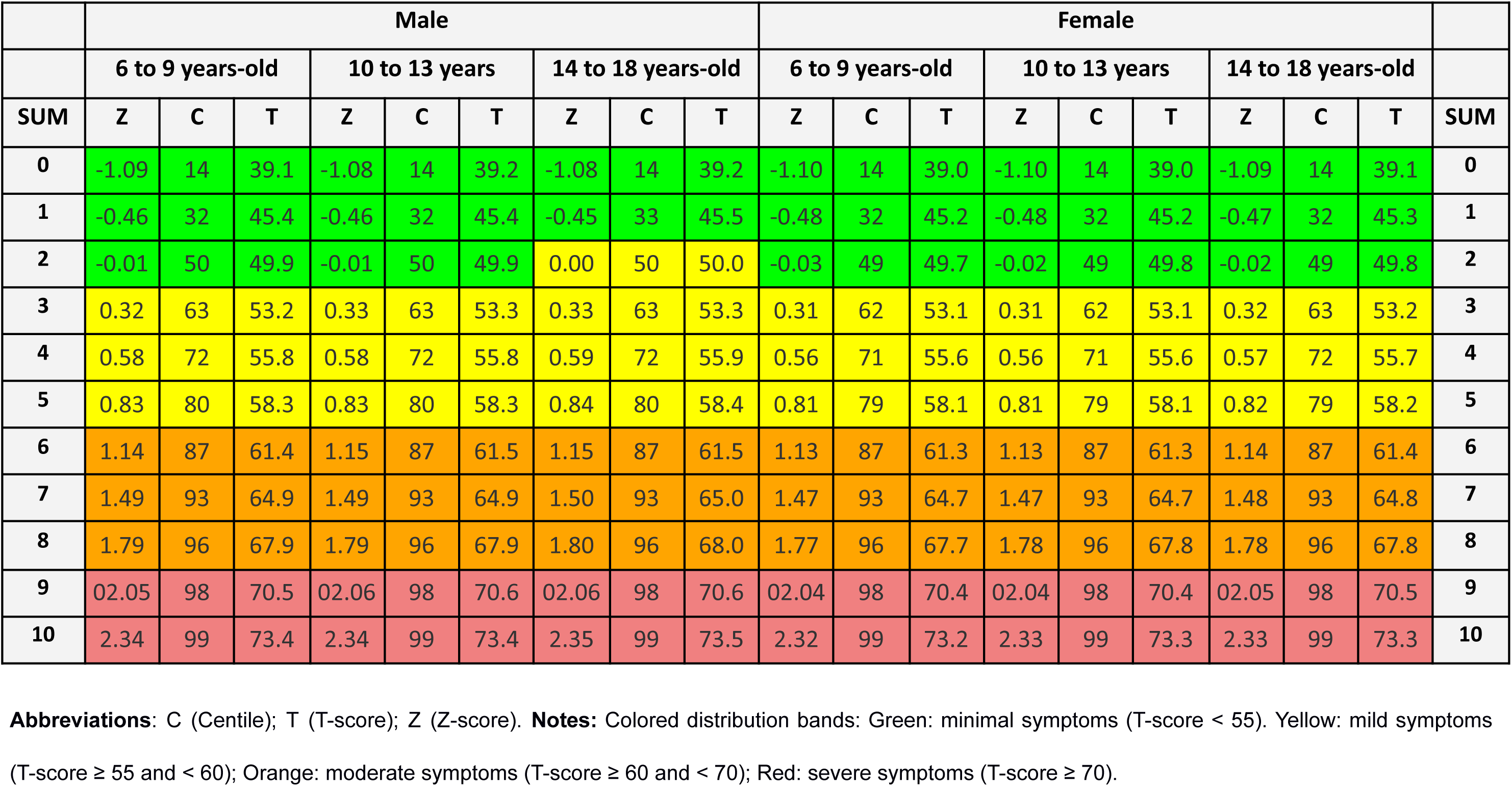
Normative references in Greece: Pediatric Symptom Checklist Short Version (PSC-17), Self-report.

Internal consistency was rated as good to excellent across all subscales, with Cronbach’s alpha (α) values ranging from 0.84 to 0.97 and Omega Total (ω□) values from 0.85 to 0.97. Factor analysis confirmed that all scales met criteria for unidimensionality, monotonicity, local independence, and global model fit, with the exception of MCHAT-R/F (failing monotonicity), CAST - inflexible/repetitive behavior (failing monotonicity), and CAST - social contact issues (failing unidimensionality, monotonicity and model fit). Factor loadings consistently exceeded the 0.3 cutoff across tools, with a single item in CAST - social contact issues - rating 0.18. Except CAST and MCHAT-R/F, all instruments demonstrated adequate item discriminative performance and test scores.

In the CAST, both subscales failed to meet monotonicity (see Supplementary Figure 1.1.5 and Supplementary Figure 1.2.5 for item response functions). In the inflexible/repetitive behavior subscale, three of 15 items exhibited low scalability (<0.3), with items 7 and 36 demonstrating weaker discrimination (see item characteristic curves in Supplementary Figure 1.1.2). In the sociability subscale, 14 of 16 items had inadequate scalability, with item characteristic curves indicating particularly low discrimination for items 23 and 35 (Supplementary Figure 1.2.2). Additionally, the sociability subscale failed to meet unidimensionality, as only RMSEA (0.06) fell within an acceptable range, while TLI (0.90), CFI (0.90), and SRMR (0.09) indicated inadequate global model fit.

MCHAT-R/F failed monotonicity as eight out of 20 items presented scalability values below the 0.3 threshold (the scale’s item response functions are detailed in **Supplementary Figure 4.5**). Item characteristic curves indicate that some questions are unable to discriminate between respondents at both the lower and upper ends of the score spectrum (see **Supplementary Figure 4.2**). For example, item 2 is overly easy to endorse, while items 5 and 12 are excessively difficult to endorse. Test information and expected score plots further indicate that the MCHAT effectively captures individuals with very low ability but fails to adequately assess those with high ability (see **Supplementary Figure 4.1**).

## Discussion

This study validates and establishes local normative references for the Greek versions of six instruments: PSC-17 (caregiver- and self-report), RCADS-25 (caregiver- and self-report), CATS-2 (caregiver- and self-report), SNAP-IV (caregiver-report), MCHAT-R/F (caregiver-report), and CAST (caregiver-report). These scales demonstrated reliable and valid properties in a nationwide community sample of children, adolescents, and caregivers, with the exception of CAST and MCHAT-R/F which did not meet all criteria in factor analysis. To the best of our knowledge, this is the first validation of these tools in Greece apart from the prior national validation of the full-length RCADS-47 [59, 60].

Scores were categorized into symptom severity bands using PROMIS ranges for standardized metrics, with norms generally aligning with cutoffs from other European and North American countries [15, 16]. For example, a crude score of 5 on the PSC-17 internalizing subscales corresponded to a moderate symptom classification (T-score 60–69), consistent with risk cutoffs reported for samples from Spain and the United States [25, 61, 62]. On the RCADS-25 (self-report), anxiety subscale scores of 30 corresponded to a T-score of 70 (severe symptoms). This is consistent with crude-to-T-score conversion tables for boys in 3rd–4th grades and girls in 5th–6th grades from U.S. samples, yet this pattern did not hold for participants beyond the 5th grade [27, 63, 64]. These findings suggest that the scales could benefit clinical practice in Greece by helping professionals distinguish between higher- and lower-symptom patients. As the Greek public mental health system lacks defined patient pathways, these symptom classifications can guide referrals and treatment decisions, supporting a scaled approach to interventions tailored to individual needs [1, 2].

Both scales assessing autism spectrum symptoms (CAST and MCHAT-R/F) did not fully meet performance criteria. This is likely due to their limited sample of 200 community-based caregivers, which is underpowered to represent individuals with more expressive clinical symptoms. Worth noting, the MCHAT-R/F is initially intended for screening children presenting developmental concerns [8]. While there is evidence of validity population-level screening, properties may differ when administered to asymptomatic samples [7]. Items were not dismissed in the scale, yet further validation in clinical samples is crucial to establish the psychometric properties and normative references of such tools.

We analyzed data from a large nationwide sample, achieving consistent psychometric properties through modern IRT approaches and generating accessible metrics that further allows for cross-instrument comparisons. However, there are also some limitations. All 1,756 caregivers were recruited from a proprietary panel of individuals willing to participate in surveys, further leading to an inclusion of 400 children and adolescents cared for by these participants. Although recruitment followed census quotas for sociodemographic variables, this is not a strictly probabilistic approach.

Another group of 801 children/adolescents was recruited through random phone calls, a method known for low response rates and potential bias. Some tools (MCHAT-R/F, CAST, CATS-2) relied on small sample sizes (137 to 200 participants), potentially underrepresenting the full spectrum of symptoms and requiring cautious interpretation of psychometric parameters [47]. While valuable for screening symptoms and orienting referral priorities, our classification bands cannot establish sound risk stratification cutoffs, as we did not include clinical samples and comparisons with gold-standard tools [65, 66].

This toolkit addresses critical gaps in evidence-based resources in Greece by validating a set of widely-used tools, providing a scalable approach that can be applied to other underserved settings. Health professionals and researchers in Greece are now better equipped to reliably screen and assess symptoms across the severity of symptoms in various conditions, including anxiety, ADHD, autism spectrum disorders, depression, and trauma. Future research with clinical samples and instrument comparisons are warranted to confirm normative references and establish construct and criterion validity of these tools in the country.

## Statements and Declarations

## Acknowledgements

The authors would like to thank the Stavros Niarchos Foundation (SNF) for funding the SNF-CMI Child and Adolescent Mental Health Initiative and SNF’s Co-President Andreas C. Dracopoulos for his leadership in creating, launching, and supporting the project. We would also like to thank Ms. Elianna Konialis, Ms. Dimitra Moustaka and Mr. Panos Papoulias for their critical role in multiple steps of the conceptualization and implementation of the SNF-CMI Child and Adolescent Mental Health Initiative. We also thank Samanta Duarte for designing the graphical representations included in this paper.

## Funding

This work is part of The Child and Adolescent Mental Health Initiative (CAMHI) aimed to enhance mental health care capacity in Greece. The CAMHI is funded by the Stavros-Niarchos Foundation (SNF) and led by the Child Mind Institute (CMI) in partnership with multiple institutions and actors in Greece.

## Conflicts of interest

All authors declare no conflicts of interest.

## Data availability statement

Data is openly available in Open Science Framework at http://doi.org/10.17605/OSF.IO/CRZ6H, including the database and the statistical codes.

## Notes

### Competing Interest Statement

The authors have declared no competing interest.

### Author Declarations

The research was approved by the Research Ethics Committee of the Democritus University of Thrace [approval number: ∆ΠΘ/ΕΗ∆Ε/42772/307].

