## Supplementary for "Psychometric properties and local normative references of PSC-17, RCADS-25, CATS-2, SNAP-IV, MCHAT-R/F, and CAST: data from a nationwide sample in Greece"

### Supplementary Material

**Supplementary Table 1 -** Strengthening the reporting of observational studies in epidemiology (STROBE) checklist

| | | Recommendation | Section |
|---|---|---|---|
| Title and abstract | 1 | (*a*) Indicate the study's design with a commonly used term in the title or the abstract | Abstract |
| | | (*b*) Provide in the abstract an informative and balanced summary of what was done and what was found | Abstract |
| Introduction | | | |
| Background/rational e | 2 | Explain the scientific background and rationale for the investigation being reported | Introduction |
| Objectives | 3 | State specific objectives, including any prespecified hypotheses | Introduction |
| Methods | | | |
| Study design | 4 | Present key elements of study design early in the paper | Methods |
| Setting | 5 | Describe the setting, locations, and relevant dates, including periods of recruitment, exposure, follow-up, and data collection | Methods |
| Participants | 6 | (*a*) Give the eligibility criteria, and the sources and methods of selection of participants | Methods<br>Figure 1 |
| Variables | 7 | Clearly define all outcomes, exposures, predictors, potential confounders, and effect modifiers. Give diagnostic criteria, if applicable | Methods |
| Data sources/ measurement | 8* | For each variable of interest, give sources of data and details of methods of assessment (measurement). Describe comparability of assessment methods if there is more than one group | Methods<br>Table 1 |
| Bias | 9 | Describe any efforts to address potential sources of bias | Methods |
| Study size | 10 | Explain how the study size was arrived at | Figure 1 |
| Quantitative variables | 11 | Explain how quantitative variables were handled in the analyses. If applicable, describe which groupings were chosen and why | Methods |
| Statistical methods | 12 | (*a*) Describe all statistical methods, including those used to control for confounding | Methods |
| | | (*b*) Describe any methods used to examine subgroups and interactions | Methods |
| | | (*c*) Explain how missing data were addressed | Not applicable |
| | | (*d*) If applicable, describe analytical methods taking account of sampling strategy | Not applicable |
| | | (*e*) Describe any sensitivity analyses | Not applicable |
| Results | | | |
| Participants | 13* | (a) Report numbers of individuals at each stage of study—eg numbers potentially eligible, examined for eligibility, confirmed eligible, included in the study, completing follow-up, and analysed | Methods<br>Results<br>Figure 1 |

| | | | |
|---|---|---|---|
| | | (b) Give reasons for non-participation at each stage | Results<br>Figure 1 |
| | | (c) Consider use of a flow diagram | Figure 1 |
| Descriptive data | 14* | (a) Give characteristics of study participants (eg demographic, clinical, social) and information on exposures and potential confounders | Results |
| | | (b) Indicate number of participants with missing data for each variable of interest | Results |
| Outcome data | 15* | Report numbers of outcome events or summary measures | Results |
| Main results | 16 | (*a*) Give unadjusted estimates and, if applicable, confounder-adjusted estimates and their precision (eg, 95% confidence interval). Make clear which confounders were adjusted for and why they were included | Results |
| | | (*b*) Report category boundaries when continuous variables were categorized | Methods<br>Results |
| | | (*c*) If relevant, consider translating estimates of relative risk into absolute risk for a meaningful time period | Not applicable |
| Other analyses | 17 | Report other analyses done—eg analyses of subgroups and interactions, and sensitivity analyses | Methods |
| Discussion | | | |
| Key results | 18 | Summarize key results with reference to study objectives | Results |
| Limitations | 19 | Discuss limitations of the study, taking into account sources of potential bias or imprecision. Discuss both direction and magnitude of any potential bias | Discussion |
| Interpretation | 20 | Give a cautious overall interpretation of results considering objectives, limitations, multiplicity of analyses, results from similar studies, and other relevant evidence | Discussion |
| Generalisability | 21 | Discuss the generalisability (external validity) of the study results | Not applicable |
| Other information | | | |
| Funding | 22 | Give the source of funding and the role of the funders for the present study and, if applicable, for the original study on which the present article is based | Statements |

**Supplementary Table 2 -** Child Autism Spectrum Test (CAST), Caregiver-report: normative references in Greece

| CAST - Caregiver-report | | | | | | | | | | | | | | | |
|---|---|---|---|---|---|---|---|---|---|---|---|---|---|---|---|
| Social Contact Issues | | | | | | | | Inflexible Behavior | | | | | | | |
| | Male | | | Female | | | | | Male | | | Female | | | |
| SUM | Z | C | T | Z | C | T | SUM | SUM | Z | C | T | Z | C | T | SUM |
| 0 | -0.90 | 0 | 41.0 | -0.91 | 0 | 40.9 | 0 | 0 | -1.14 | 0 | 38.6 | -1.15 | 0 | 38.5 | 0 |
| 1 | -0.53 | 6 | 44.7 | -0.54 | 6 | 44.6 | 1 | 1 | -0.73 | 8 | 42.7 | -0.74 | 8 | 42.6 | 1 |
| 2 | -0.15 | 12 | 48.5 | -0.16 | 12 | 48.4 | 2 | 2 | -0.33 | 17 | 46.7 | -0.34 | 17 | 46.6 | 2 |
| 3 | 0.24 | 19 | 52.4 | 0.22 | 19 | 52.2 | 3 | 3 | 0.04 | 25 | 50.4 | 0.03 | 25 | 50.3 | 3 |
| 4 | 0.60 | 25 | 56.0 | 0.58 | 25 | 55.8 | 4 | 4 | 0.35 | 33 | 53.5 | 0.34 | 33 | 53.4 | 4 |
| 5 | 0.92 | 31 | 59.2 | 0.91 | 31 | 59.1 | 5 | 5 | 0.61 | 42 | 56.1 | 0.60 | 42 | 56.0 | 5 |
| 6 | 1.22 | 38 | 62.2 | 1.20 | 38 | 62.0 | 6 | 6 | 0.84 | 50 | 58.4 | 0.83 | 50 | 58.3 | 6 |
| 7 | 1.48 | 44 | 64.8 | 1.46 | 44 | 64.6 | 7 | 7 | 01.07 | 58 | 60.7 | 01.06 | 58 | 60.6 | 7 |
| 8 | 1.72 | 50 | 67.2 | 1.70 | 50 | 67.0 | 8 | 8 | 1.30 | 67 | 63.0 | 1.29 | 67 | 62.9 | 8 |
| 9 | 1.95 | 56 | 69.5 | 1.93 | 56 | 69.3 | 9 | 9 | 1.53 | 75 | 65.3 | 1.52 | 75 | 65.2 | 9 |
| 10 | 2.16 | 62 | 71.6 | 2.15 | 62 | 71.5 | 10 | 10 | 1.76 | 83 | 67.6 | 1.75 | 83 | 67.5 | 10 |
| 11 | 2.37 | 69 | 73.7 | 2.35 | 69 | 73.5 | 11 | 11 | 02.01 | 92 | 70.1 | 2.00 | 92 | 70.0 | 11 |
| 12 | 2.56 | 75 | 75.6 | 2.55 | 75 | 75.5 | 12 | 12 | 2.26 | 100 | 72.6 | 2.25 | 100 | 72.5 | 12 |
| 13 | 2.75 | 81 | 77.5 | 2.73 | 81 | 77.3 | 13 | | | | | | | | |
| 14 | 2.93 | 88 | 79.3 | 2.92 | 88 | 79.2 | 14 | | | | | | | | |
| 15 | 3.11 | 94 | 81.1 | 3.10 | 94 | 81.0 | 15 | | | | | | | | |
| 16 | 3.29 | 100 | 82.9 | 3.27 | 100 | 82.7 | 16 | | | | | | | | |

**Abbreviations**: C (Centile); T (T-score); Z (Z-score). **Notes**: Colored distribution bands: Green: minimal symptoms (T-score < 55). Yellow: mild symptoms (T-score ≥ 55 and < 60); Orange: moderate symptoms (T-score ≥ 60 and < 70); Red: severe symptoms (T-score ≥ 70).

**Supplementary Table 3 -** Child and Adolescent Trauma Screen-2 (CATS-2), Caregiver-report: normative references in Greece

| CATS-2 - Caregiver-report | | | | | | | | | | | | | | | | | | | |
|---|---|---|---|---|---|---|---|---|---|---|---|---|---|---|---|---|---|---|---|
| | Male | | | | | | | | | Female | | | | | | | | | |
| | 6 to 9 years-old | | | 10 to 13 years | | | 14 to 18 years-old | | | 6 to 9 years-old | | | 10 to 13 years | | | 14 to 18 years-old | | | |
| SUM | Z | C | T | Z | C | T | Z | C | T | Z | C | T | Z | C | T | Z | C | T | SUM |
| 0 | -1.68 | 5 | 33.2 | -1.70 | 4 | 33.0 | -1.68 | 5 | 33.2 | -1.69 | 5 | 33.1 | -1.70 | 4 | 33.0 | -1.68 | 5 | 33.2 | 0 |
| 1 | -1.49 | 7 | 35.1 | -1.51 | 7 | 34.9 | -1.49 | 7 | 35.1 | -1.50 | 7 | 35.0 | -1.51 | 7 | 34.9 | -1.49 | 7 | 35.1 | 1 |
| 2 | -1.31 | 10 | 36.9 | -1.32 | 9 | 36.8 | -1.30 | 10 | 37.0 | -1.31 | 10 | 36.9 | -1.33 | 9 | 36.7 | -1.30 | 10 | 37.0 | 2 |
| 3 | -1.13 | 13 | 38.7 | -1.14 | 13 | 38.6 | -1.12 | 13 | 38.8 | -1.13 | 13 | 38.7 | -1.15 | 13 | 38.5 | -1.12 | 13 | 38.8 | 3 |
| 4 | -0.95 | 17 | 40.5 | -0.97 | 17 | 40.3 | -0.95 | 17 | 40.5 | -0.96 | 17 | 40.4 | -0.97 | 17 | 40.3 | -0.95 | 17 | 40.5 | 4 |
| 5 | -0.80 | 21 | 42.0 | -0.81 | 21 | 41.9 | -0.79 | 21 | 42.1 | -0.80 | 21 | 42.0 | -0.82 | 21 | 41.8 | -0.79 | 21 | 42.1 | 5 |
| 6 | -0.65 | 26 | 43.5 | -0.67 | 25 | 43.3 | -0.65 | 26 | 43.5 | -0.66 | 25 | 43.4 | -0.67 | 25 | 43.3 | -0.65 | 26 | 43.5 | 6 |
| 7 | -0.53 | 30 | 44.7 | -0.55 | 29 | 44.5 | -0.52 | 30 | 44.8 | -0.53 | 30 | 44.7 | -0.55 | 29 | 44.5 | -0.53 | 30 | 44.7 | 7 |
| 8 | -0.42 | 34 | 45.8 | -0.44 | 33 | 45.6 | -0.42 | 34 | 45.8 | -0.43 | 33 | 45.7 | -0.44 | 33 | 45.6 | -0.42 | 34 | 45.8 | 8 |
| 9 | -0.33 | 37 | 46.7 | -0.35 | 36 | 46.5 | -0.33 | 37 | 46.7 | -0.33 | 37 | 46.7 | -0.35 | 36 | 46.5 | -0.33 | 37 | 46.7 | 9 |
| 10 | -0.25 | 40 | 47.5 | -0.27 | 39 | 47.3 | -0.25 | 40 | 47.5 | -0.26 | 40 | 47.4 | -0.27 | 39 | 47.3 | -0.25 | 40 | 47.5 | 10 |
| 11 | -0.19 | 42 | 48.1 | -0.20 | 42 | 48.0 | -0.18 | 43 | 48.2 | -0.19 | 42 | 48.1 | -0.20 | 42 | 48.0 | -0.18 | 43 | 48.2 | 11 |
| 12 | -0.12 | 45 | 48.8 | -0.14 | 44 | 48.6 | -0.12 | 45 | 48.8 | -0.13 | 45 | 48.7 | -0.14 | 44 | 48.6 | -0.12 | 45 | 48.8 | 12 |
| 13 | -0.07 | 47 | 49.3 | -0.08 | 47 | 49.2 | -0.06 | 48 | 49.4 | -0.07 | 47 | 49.3 | -0.09 | 46 | 49.1 | -0.07 | 47 | 49.3 | 13 |
| 14 | -0.01 | 50 | 49.9 | -0.03 | 49 | 49.7 | -0.01 | 50 | 49.9 | -0.02 | 49 | 49.8 | -0.03 | 49 | 49.7 | -0.01 | 50 | 49.9 | 14 |
| 15 | 0.04 | 52 | 50.4 | 0.03 | 51 | 50.3 | 0.05 | 52 | 50.5 | 0.04 | 52 | 50.4 | 0.02 | 51 | 50.2 | 0.04 | 52 | 50.4 | 15 |
| 16 | 0.10 | 54 | 51.0 | 0.08 | 53 | 50.8 | 0.10 | 54 | 51.0 | 0.10 | 54 | 51.0 | 0.08 | 53 | 50.8 | 0.10 | 54 | 51.0 | 16 |

| | | | | | | | | | | | | | | | | | | |
|---|---|---|---|---|---|---|---|---|---|---|---|---|---|---|---|---|---|---|
| **17** | 0.16 | 56 | 51.6 | 0.14 | 56 | 51.4 | 0.16 | 56 | 51.6 | 0.15 | 56 | 51.5 | 0.14 | 56 | 51.4 | 0.16 | 56 | 51.6 | **17** |
| **18** | 0.22 | 59 | 52.2 | 0.20 | 58 | 52.0 | 0.22 | 59 | 52.2 | 0.21 | 58 | 52.1 | 0.20 | 58 | 52.0 | 0.22 | 59 | 52.2 | **18** |
| **19** | 0.27 | 61 | 52.7 | 0.26 | 60 | 52.6 | 0.28 | 61 | 52.8 | 0.27 | 61 | 52.7 | 0.25 | 60 | 52.5 | 0.28 | 61 | 52.8 | **19** |
| **20** | 0.33 | 63 | 53.3 | 0.31 | 62 | 53.1 | 0.34 | 63 | 53.4 | 0.33 | 63 | 53.3 | 0.31 | 62 | 53.1 | 0.33 | 63 | 53.3 | **20** |
| **21** | 0.39 | 65 | 53.9 | 0.37 | 64 | 53.7 | 0.39 | 65 | 53.9 | 0.38 | 65 | 53.8 | 0.37 | 64 | 53.7 | 0.39 | 65 | 53.9 | **21** |
| **22** | 0.44 | 67 | 54.4 | 0.42 | 66 | 54.2 | 0.44 | 67 | 54.4 | 0.44 | 67 | 54.4 | 0.42 | 66 | 54.2 | 0.44 | 67 | 54.4 | **22** |
| **23** | 0.49 | 69 | 54.9 | 0.47 | 68 | 54.7 | 0.49 | 69 | 54.9 | 0.49 | 69 | 54.9 | 0.47 | 68 | 54.7 | 0.49 | 69 | 54.9 | **23** |
| **24** | 0.54 | 71 | 55.4 | 0.52 | 70 | 55.2 | 0.54 | 71 | 55.4 | 0.53 | 70 | 55.3 | 0.52 | 70 | 55.2 | 0.54 | 71 | 55.4 | **24** |
| **25** | 0.58 | 72 | 55.8 | 0.57 | 72 | 55.7 | 0.59 | 72 | 55.9 | 0.58 | 72 | 55.8 | 0.56 | 71 | 55.6 | 0.58 | 72 | 55.8 | **25** |
| **26** | 0.63 | 74 | 56.3 | 0.61 | 73 | 56.1 | 0.63 | 74 | 56.3 | 0.62 | 73 | 56.2 | 0.61 | 73 | 56.1 | 0.63 | 74 | 56.3 | **26** |
| **27** | 0.67 | 75 | 56.7 | 0.66 | 75 | 56.6 | 0.68 | 75 | 56.8 | 0.67 | 75 | 56.7 | 0.65 | 74 | 56.5 | 0.68 | 75 | 56.8 | **27** |
| **28** | 0.72 | 76 | 57.2 | 0.70 | 76 | 57.0 | 0.73 | 77 | 57.3 | 0.72 | 76 | 57.2 | 0.70 | 76 | 57.0 | 0.72 | 76 | 57.2 | **28** |
| **29** | 0.77 | 78 | 57.7 | 0.75 | 77 | 57.5 | 0.78 | 78 | 57.8 | 0.77 | 78 | 57.7 | 0.75 | 77 | 57.5 | 0.77 | 78 | 57.7 | **29** |
| **30** | 0.82 | 79 | 58.2 | 0.80 | 79 | 58.0 | 0.83 | 80 | 58.3 | 0.82 | 79 | 58.2 | 0.80 | 79 | 58.0 | 0.82 | 79 | 58.2 | **30** |
| **31** | 0.87 | 81 | 58.7 | 0.85 | 80 | 58.5 | 0.88 | 81 | 58.8 | 0.87 | 81 | 58.7 | 0.85 | 80 | 58.5 | 0.87 | 81 | 58.7 | **31** |
| **32** | 0.92 | 82 | 59.2 | 0.91 | 82 | 59.1 | 0.93 | 82 | 59.3 | 0.92 | 82 | 59.2 | 0.90 | 82 | 59.0 | 0.92 | 82 | 59.2 | **32** |
| **33** | 0.97 | 83 | 59.7 | 0.96 | 83 | 59.6 | 0.98 | 84 | 59.8 | 0.97 | 83 | 59.7 | 0.95 | 83 | 59.5 | 0.97 | 83 | 59.7 | **33** |
| **34** | 01.02 | 85 | 60.2 | 1.00 | 84 | 60.0 | 01.02 | 85 | 60.2 | 01.02 | 85 | 60.2 | 1.00 | 84 | 60.0 | 01.02 | 85 | 60.2 | **34** |
| **35** | 01.06 | 86 | 60.6 | 01.05 | 85 | 60.5 | 01.07 | 86 | 60.7 | 01.06 | 86 | 60.6 | 01.04 | 85 | 60.4 | 01.07 | 86 | 60.7 | **35** |
| **36** | 1.10 | 86 | 61.0 | 01.09 | 86 | 60.9 | 1.11 | 87 | 61.1 | 1.10 | 86 | 61.0 | 01.08 | 86 | 60.8 | 1.11 | 87 | 61.1 | **36** |
| **37** | 1.14 | 87 | 61.4 | 1.12 | 87 | 61.2 | 1.14 | 87 | 61.4 | 1.14 | 87 | 61.4 | 1.12 | 87 | 61.2 | 1.14 | 87 | 61.4 | **37** |
| **38** | 1.17 | 88 | 61.7 | 1.16 | 88 | 61.6 | 1.18 | 88 | 61.8 | 1.17 | 88 | 61.7 | 1.15 | 87 | 61.5 | 1.18 | 88 | 61.8 | **38** |

| | | | | | | | | | | | | | | | | | | | |
|---|---|---|---|---|---|---|---|---|---|---|---|---|---|---|---|---|---|---|---|
| **39** | 1.20 | 88 | 62.0 | 1.19 | 88 | 61.9 | 1.21 | 89 | 62.1 | 1.20 | 88 | 62.0 | 1.18 | 88 | 61.8 | 1.21 | 89 | 62.1 | **39** |
| **40** | 1.23 | 89 | 62.3 | 1.22 | 89 | 62.2 | 1.24 | 89 | 62.4 | 1.23 | 89 | 62.3 | 1.21 | 89 | 62.1 | 1.24 | 89 | 62.4 | **40** |
| **41** | 1.26 | 90 | 62.6 | 1.25 | 89 | 62.5 | 1.27 | 90 | 62.7 | 1.26 | 90 | 62.6 | 1.24 | 89 | 62.4 | 1.27 | 90 | 62.7 | **41** |
| **42** | 1.30 | 90 | 63.0 | 1.28 | 90 | 62.8 | 1.30 | 90 | 63.0 | 1.29 | 90 | 62.9 | 1.28 | 90 | 62.8 | 1.30 | 90 | 63.0 | **42** |
| **43** | 1.33 | 91 | 63.3 | 1.32 | 91 | 63.2 | 1.34 | 91 | 63.4 | 1.33 | 91 | 63.3 | 1.31 | 90 | 63.1 | 1.33 | 91 | 63.3 | **43** |
| **44** | 1.37 | 91 | 63.7 | 1.35 | 91 | 63.5 | 1.38 | 92 | 63.8 | 1.37 | 91 | 63.7 | 1.35 | 91 | 63.5 | 1.37 | 91 | 63.7 | **44** |
| **45** | 1.41 | 92 | 64.1 | 1.40 | 92 | 64.0 | 1.42 | 92 | 64.2 | 1.41 | 92 | 64.1 | 1.39 | 92 | 63.9 | 1.42 | 92 | 64.2 | **45** |
| **46** | 1.46 | 93 | 64.6 | 1.44 | 93 | 64.4 | 1.46 | 93 | 64.6 | 1.46 | 93 | 64.6 | 1.44 | 93 | 64.4 | 1.46 | 93 | 64.6 | **46** |
| **47** | 1.51 | 93 | 65.1 | 1.49 | 93 | 64.9 | 1.51 | 93 | 65.1 | 1.50 | 93 | 65.0 | 1.49 | 93 | 64.9 | 1.51 | 93 | 65.1 | **47** |
| **48** | 1.56 | 94 | 65.6 | 1.54 | 94 | 65.4 | 1.56 | 94 | 65.6 | 1.55 | 94 | 65.5 | 1.54 | 94 | 65.4 | 1.56 | 94 | 65.6 | **48** |
| **49** | 1.61 | 95 | 66.1 | 1.59 | 94 | 65.9 | 1.61 | 95 | 66.1 | 1.61 | 95 | 66.1 | 1.59 | 94 | 65.9 | 1.61 | 95 | 66.1 | **49** |
| **50** | 1.66 | 95 | 66.6 | 1.64 | 95 | 66.4 | 1.67 | 95 | 66.7 | 1.66 | 95 | 66.6 | 1.64 | 95 | 66.4 | 1.66 | 95 | 66.6 | **50** |
| **51** | 1.71 | 96 | 67.1 | 1.70 | 96 | 67.0 | 1.72 | 96 | 67.2 | 1.71 | 96 | 67.1 | 1.69 | 95 | 66.9 | 1.72 | 96 | 67.2 | **51** |
| **52** | 1.77 | 96 | 67.7 | 1.75 | 96 | 67.5 | 1.77 | 96 | 67.7 | 1.76 | 96 | 67.6 | 1.75 | 96 | 67.5 | 1.77 | 96 | 67.7 | **52** |
| **53** | 1.82 | 97 | 68.2 | 1.80 | 96 | 68.0 | 1.82 | 97 | 68.2 | 1.82 | 97 | 68.2 | 1.80 | 96 | 68.0 | 1.82 | 97 | 68.2 | **53** |
| **54** | 1.87 | 97 | 68.7 | 1.86 | 97 | 68.6 | 1.88 | 97 | 68.8 | 1.87 | 97 | 68.7 | 1.85 | 97 | 68.5 | 1.88 | 97 | 68.8 | **54** |
| **55** | 1.93 | 97 | 69.3 | 1.91 | 97 | 69.1 | 1.94 | 97 | 69.4 | 1.93 | 97 | 69.3 | 1.91 | 97 | 69.1 | 1.93 | 97 | 69.3 | **55** |
| **56** | 1.99 | 98 | 69.9 | 1.97 | 98 | 69.7 | 2.00 | 98 | 70.0 | 1.99 | 98 | 69.9 | 1.97 | 98 | 69.7 | 1.99 | 98 | 69.9 | **56** |
| **57** | 02.05 | 98 | 70.5 | 02.04 | 98 | 70.4 | 02.06 | 98 | 70.6 | 02.05 | 98 | 70.5 | 02.03 | 98 | 70.3 | 02.06 | 98 | 70.6 | **57** |
| **58** | 2.12 | 98 | 71.2 | 2.10 | 98 | 71.0 | 2.13 | 98 | 71.3 | 2.12 | 98 | 71.2 | 2.10 | 98 | 71.0 | 2.12 | 98 | 71.2 | **58** |
| **59** | 2.19 | 99 | 71.9 | 2.17 | 98 | 71.7 | 2.20 | 99 | 72.0 | 2.19 | 99 | 71.9 | 2.17 | 98 | 71.7 | 2.19 | 99 | 71.9 | **59** |
| **60** | 2.27 | 99 | 72.7 | 2.25 | 99 | 72.5 | 2.27 | 99 | 72.7 | 2.26 | 99 | 72.6 | 2.25 | 99 | 72.5 | 2.27 | 99 | 72.7 | **60** |

| SUM | Z | C | T | Z | C | T | Z | C | T | Z | C | T | Z | C | T | Z | C | T | SUM |
|---|---|---|---|---|---|---|---|---|---|---|---|---|---|---|---|---|---|---|---|
| | 6 to 9 years-old | | | 10 to 13 years | | | 14 to 18 years-old | | | 6 to 9 years-old | | | 10 to 13 years | | | 14 to 18 years-old | | | |
| | Male | | | | | | | | | Female | | | | | | | | | |

**Abbreviations**: C (Centile); T (T-score); Z (Z-score). **Notes**: Colored distribution bands: Green: minimal symptoms (T-score < 55). Yellow: mild symptoms (T-score ≥ 55 and < 60); Orange: moderate symptoms (T-score ≥ 60 and < 70); Red: severe symptoms (T-score ≥ 70).

**Supplementary Table 4 -** Child and Adolescent Trauma Screen-2 (CATS-2), Self-report: normative references in Greece

| CATS-2 - Self-report | | | | | | | | | | | | | | | | | | | |
|---|---|---|---|---|---|---|---|---|---|---|---|---|---|---|---|---|---|---|---|
| | Male | | | | | | | | | Female | | | | | | | | | |
| | 6 to 9 years-old | | | 10 to 13 years | | | 14 to 18 years-old | | | 6 to 9 years-old | | | 10 to 13 years | | | 14 to 18 years-old | | | |
| SUM | Z | C | T | Z | C | T | Z | C | T | Z | C | T | Z | C | T | Z | C | T | SUM |
| 0 | -1.68 | 5 | 33.2 | -1.70 | 4 | 33.0 | -1.68 | 5 | 33.2 | -1.69 | 5 | 33.1 | -1.70 | 4 | 33.0 | -1.68 | 5 | 33.2 | 0 |
| 1 | -1.49 | 7 | 35.1 | -1.51 | 7 | 34.9 | -1.49 | 7 | 35.1 | -1.50 | 7 | 35.0 | -1.51 | 7 | 34.9 | -1.49 | 7 | 35.1 | 1 |
| 2 | -1.31 | 10 | 36.9 | -1.32 | 9 | 36.8 | -1.30 | 10 | 37.0 | -1.31 | 10 | 36.9 | -1.33 | 9 | 36.7 | -1.30 | 10 | 37.0 | 2 |
| 3 | -1.13 | 13 | 38.7 | -1.14 | 13 | 38.6 | -1.12 | 13 | 38.8 | -1.13 | 13 | 38.7 | -1.15 | 13 | 38.5 | -1.12 | 13 | 38.8 | 3 |
| 4 | -0.95 | 17 | 40.5 | -0.97 | 17 | 40.3 | -0.95 | 17 | 40.5 | -0.96 | 17 | 40.4 | -0.97 | 17 | 40.3 | -0.95 | 17 | 40.5 | 4 |
| 5 | -0.80 | 21 | 42.0 | -0.81 | 21 | 41.9 | -0.79 | 21 | 42.1 | -0.80 | 21 | 42.0 | -0.82 | 21 | 41.8 | -0.79 | 21 | 42.1 | 5 |
| 6 | -0.65 | 26 | 43.5 | -0.67 | 25 | 43.3 | -0.65 | 26 | 43.5 | -0.66 | 25 | 43.4 | -0.67 | 25 | 43.3 | -0.65 | 26 | 43.5 | 6 |
| 7 | -0.53 | 30 | 44.7 | -0.55 | 29 | 44.5 | -0.52 | 30 | 44.8 | -0.53 | 30 | 44.7 | -0.55 | 29 | 44.5 | -0.53 | 30 | 44.7 | 7 |
| 8 | -0.42 | 34 | 45.8 | -0.44 | 33 | 45.6 | -0.42 | 34 | 45.8 | -0.43 | 33 | 45.7 | -0.44 | 33 | 45.6 | -0.42 | 34 | 45.8 | 8 |
| 9 | -0.33 | 37 | 46.7 | -0.35 | 36 | 46.5 | -0.33 | 37 | 46.7 | -0.33 | 37 | 46.7 | -0.35 | 36 | 46.5 | -0.33 | 37 | 46.7 | 9 |
| 10 | -0.25 | 40 | 47.5 | -0.27 | 39 | 47.3 | -0.25 | 40 | 47.5 | -0.26 | 40 | 47.4 | -0.27 | 39 | 47.3 | -0.25 | 40 | 47.5 | 10 |
| 11 | -0.19 | 42 | 48.1 | -0.20 | 42 | 48.0 | -0.18 | 43 | 48.2 | -0.19 | 42 | 48.1 | -0.20 | 42 | 48.0 | -0.18 | 43 | 48.2 | 11 |
| 12 | -0.12 | 45 | 48.8 | -0.14 | 44 | 48.6 | -0.12 | 45 | 48.8 | -0.13 | 45 | 48.7 | -0.14 | 44 | 48.6 | -0.12 | 45 | 48.8 | 12 |
| 13 | -0.07 | 47 | 49.3 | -0.08 | 47 | 49.2 | -0.06 | 48 | 49.4 | -0.07 | 47 | 49.3 | -0.09 | 46 | 49.1 | -0.07 | 47 | 49.3 | 13 |
| 14 | -0.01 | 50 | 49.9 | -0.03 | 49 | 49.7 | -0.01 | 50 | 49.9 | -0.02 | 49 | 49.8 | -0.03 | 49 | 49.7 | -0.01 | 50 | 49.9 | 14 |
| 15 | 0.04 | 52 | 50.4 | 0.03 | 51 | 50.3 | 0.05 | 52 | 50.5 | 0.04 | 52 | 50.4 | 0.02 | 51 | 50.2 | 0.04 | 52 | 50.4 | 15 |
| 16 | 0.10 | 54 | 51.0 | 0.08 | 53 | 50.8 | 0.10 | 54 | 51.0 | 0.10 | 54 | 51.0 | 0.08 | 53 | 50.8 | 0.10 | 54 | 51.0 | 16 |

| | | | | | | | | | | | | | | | | | | | |
|---|---|---|---|---|---|---|---|---|---|---|---|---|---|---|---|---|---|---|---|
| **17** | 0.16 | 56 | 51.6 | 0.14 | 56 | 51.4 | 0.16 | 56 | 51.6 | 0.15 | 56 | 51.5 | 0.14 | 56 | 51.4 | 0.16 | 56 | 51.6 | **17** |
| **18** | 0.22 | 59 | 52.2 | 0.20 | 58 | 52.0 | 0.22 | 59 | 52.2 | 0.21 | 58 | 52.1 | 0.20 | 58 | 52.0 | 0.22 | 59 | 52.2 | **18** |
| **19** | 0.27 | 61 | 52.7 | 0.26 | 60 | 52.6 | 0.28 | 61 | 52.8 | 0.27 | 61 | 52.7 | 0.25 | 60 | 52.5 | 0.28 | 61 | 52.8 | **19** |
| **20** | 0.33 | 63 | 53.3 | 0.31 | 62 | 53.1 | 0.34 | 63 | 53.4 | 0.33 | 63 | 53.3 | 0.31 | 62 | 53.1 | 0.33 | 63 | 53.3 | **20** |
| **21** | 0.39 | 65 | 53.9 | 0.37 | 64 | 53.7 | 0.39 | 65 | 53.9 | 0.38 | 65 | 53.8 | 0.37 | 64 | 53.7 | 0.39 | 65 | 53.9 | **21** |
| **22** | 0.44 | 67 | 54.4 | 0.42 | 66 | 54.2 | 0.44 | 67 | 54.4 | 0.44 | 67 | 54.4 | 0.42 | 66 | 54.2 | 0.44 | 67 | 54.4 | **22** |
| **23** | 0.49 | 69 | 54.9 | 0.47 | 68 | 54.7 | 0.49 | 69 | 54.9 | 0.49 | 69 | 54.9 | 0.47 | 68 | 54.7 | 0.49 | 69 | 54.9 | **23** |
| **24** | 0.54 | 71 | 55.4 | 0.52 | 70 | 55.2 | 0.54 | 71 | 55.4 | 0.53 | 70 | 55.3 | 0.52 | 70 | 55.2 | 0.54 | 71 | 55.4 | **24** |
| **25** | 0.58 | 72 | 55.8 | 0.57 | 72 | 55.7 | 0.59 | 72 | 55.9 | 0.58 | 72 | 55.8 | 0.56 | 71 | 55.6 | 0.58 | 72 | 55.8 | **25** |
| **26** | 0.63 | 74 | 56.3 | 0.61 | 73 | 56.1 | 0.63 | 74 | 56.3 | 0.62 | 73 | 56.2 | 0.61 | 73 | 56.1 | 0.63 | 74 | 56.3 | **26** |
| **27** | 0.67 | 75 | 56.7 | 0.66 | 75 | 56.6 | 0.68 | 75 | 56.8 | 0.67 | 75 | 56.7 | 0.65 | 74 | 56.5 | 0.68 | 75 | 56.8 | **27** |
| **28** | 0.72 | 76 | 57.2 | 0.70 | 76 | 57.0 | 0.73 | 77 | 57.3 | 0.72 | 76 | 57.2 | 0.70 | 76 | 57.0 | 0.72 | 76 | 57.2 | **28** |
| **29** | 0.77 | 78 | 57.7 | 0.75 | 77 | 57.5 | 0.78 | 78 | 57.8 | 0.77 | 78 | 57.7 | 0.75 | 77 | 57.5 | 0.77 | 78 | 57.7 | **29** |
| **30** | 0.82 | 79 | 58.2 | 0.80 | 79 | 58.0 | 0.83 | 80 | 58.3 | 0.82 | 79 | 58.2 | 0.80 | 79 | 58.0 | 0.82 | 79 | 58.2 | **30** |
| **31** | 0.87 | 81 | 58.7 | 0.85 | 80 | 58.5 | 0.88 | 81 | 58.8 | 0.87 | 81 | 58.7 | 0.85 | 80 | 58.5 | 0.87 | 81 | 58.7 | **31** |
| **32** | 0.92 | 82 | 59.2 | 0.91 | 82 | 59.1 | 0.93 | 82 | 59.3 | 0.92 | 82 | 59.2 | 0.90 | 82 | 59.0 | 0.92 | 82 | 59.2 | **32** |
| **33** | 0.97 | 83 | 59.7 | 0.96 | 83 | 59.6 | 0.98 | 84 | 59.8 | 0.97 | 83 | 59.7 | 0.95 | 83 | 59.5 | 0.97 | 83 | 59.7 | **33** |
| **34** | 01.02 | 85 | 60.2 | 1.00 | 84 | 60.0 | 01.02 | 85 | 60.2 | 01.02 | 85 | 60.2 | 1.00 | 84 | 60.0 | 01.02 | 85 | 60.2 | **34** |
| **35** | 01.06 | 86 | 60.6 | 01.05 | 85 | 60.5 | 01.07 | 86 | 60.7 | 01.06 | 86 | 60.6 | 01.04 | 85 | 60.4 | 01.07 | 86 | 60.7 | **35** |
| **36** | 1.10 | 86 | 61.0 | 01.09 | 86 | 60.9 | 1.11 | 87 | 61.1 | 1.10 | 86 | 61.0 | 01.08 | 86 | 60.8 | 1.11 | 87 | 61.1 | **36** |
| **37** | 1.14 | 87 | 61.4 | 1.12 | 87 | 61.2 | 1.14 | 87 | 61.4 | 1.14 | 87 | 61.4 | 1.12 | 87 | 61.2 | 1.14 | 87 | 61.4 | **37** |
| **38** | 1.17 | 88 | 61.7 | 1.16 | 88 | 61.6 | 1.18 | 88 | 61.8 | 1.17 | 88 | 61.7 | 1.15 | 87 | 61.5 | 1.18 | 88 | 61.8 | **38** |

| | | | | | | | | | | | | | | | | | | | |
|---|---|---|---|---|---|---|---|---|---|---|---|---|---|---|---|---|---|---|---|
| **39** | 1.20 | 88 | 62.0 | 1.19 | 88 | 61.9 | 1.21 | 89 | 62.1 | 1.20 | 88 | 62.0 | 1.18 | 88 | 61.8 | 1.21 | 89 | 62.1 | **39** |
| **40** | 1.23 | 89 | 62.3 | 1.22 | 89 | 62.2 | 1.24 | 89 | 62.4 | 1.23 | 89 | 62.3 | 1.21 | 89 | 62.1 | 1.24 | 89 | 62.4 | **40** |
| **41** | 1.26 | 90 | 62.6 | 1.25 | 89 | 62.5 | 1.27 | 90 | 62.7 | 1.26 | 90 | 62.6 | 1.24 | 89 | 62.4 | 1.27 | 90 | 62.7 | **41** |
| **42** | 1.30 | 90 | 63.0 | 1.28 | 90 | 62.8 | 1.30 | 90 | 63.0 | 1.29 | 90 | 62.9 | 1.28 | 90 | 62.8 | 1.30 | 90 | 63.0 | **42** |
| **43** | 1.33 | 91 | 63.3 | 1.32 | 91 | 63.2 | 1.34 | 91 | 63.4 | 1.33 | 91 | 63.3 | 1.31 | 90 | 63.1 | 1.33 | 91 | 63.3 | **43** |
| **44** | 1.37 | 91 | 63.7 | 1.35 | 91 | 63.5 | 1.38 | 92 | 63.8 | 1.37 | 91 | 63.7 | 1.35 | 91 | 63.5 | 1.37 | 91 | 63.7 | **44** |
| **45** | 1.41 | 92 | 64.1 | 1.40 | 92 | 64.0 | 1.42 | 92 | 64.2 | 1.41 | 92 | 64.1 | 1.39 | 92 | 63.9 | 1.42 | 92 | 64.2 | **45** |
| **46** | 1.46 | 93 | 64.6 | 1.44 | 93 | 64.4 | 1.46 | 93 | 64.6 | 1.46 | 93 | 64.6 | 1.44 | 93 | 64.4 | 1.46 | 93 | 64.6 | **46** |
| **47** | 1.51 | 93 | 65.1 | 1.49 | 93 | 64.9 | 1.51 | 93 | 65.1 | 1.50 | 93 | 65.0 | 1.49 | 93 | 64.9 | 1.51 | 93 | 65.1 | **47** |
| **48** | 1.56 | 94 | 65.6 | 1.54 | 94 | 65.4 | 1.56 | 94 | 65.6 | 1.55 | 94 | 65.5 | 1.54 | 94 | 65.4 | 1.56 | 94 | 65.6 | **48** |
| **49** | 1.61 | 95 | 66.1 | 1.59 | 94 | 65.9 | 1.61 | 95 | 66.1 | 1.61 | 95 | 66.1 | 1.59 | 94 | 65.9 | 1.61 | 95 | 66.1 | **49** |
| **50** | 1.66 | 95 | 66.6 | 1.64 | 95 | 66.4 | 1.67 | 95 | 66.7 | 1.66 | 95 | 66.6 | 1.64 | 95 | 66.4 | 1.66 | 95 | 66.6 | **50** |
| **51** | 1.71 | 96 | 67.1 | 1.70 | 96 | 67.0 | 1.72 | 96 | 67.2 | 1.71 | 96 | 67.1 | 1.69 | 95 | 66.9 | 1.72 | 96 | 67.2 | **51** |
| **52** | 1.77 | 96 | 67.7 | 1.75 | 96 | 67.5 | 1.77 | 96 | 67.7 | 1.76 | 96 | 67.6 | 1.75 | 96 | 67.5 | 1.77 | 96 | 67.7 | **52** |
| **53** | 1.82 | 97 | 68.2 | 1.80 | 96 | 68.0 | 1.82 | 97 | 68.2 | 1.82 | 97 | 68.2 | 1.80 | 96 | 68.0 | 1.82 | 97 | 68.2 | **53** |
| **54** | 1.87 | 97 | 68.7 | 1.86 | 97 | 68.6 | 1.88 | 97 | 68.8 | 1.87 | 97 | 68.7 | 1.85 | 97 | 68.5 | 1.88 | 97 | 68.8 | **54** |
| **55** | 1.93 | 97 | 69.3 | 1.91 | 97 | 69.1 | 1.94 | 97 | 69.4 | 1.93 | 97 | 69.3 | 1.91 | 97 | 69.1 | 1.93 | 97 | 69.3 | **55** |
| **56** | 1.99 | 98 | 69.9 | 1.97 | 98 | 69.7 | 2.00 | 98 | 70.0 | 1.99 | 98 | 69.9 | 1.97 | 98 | 69.7 | 1.99 | 98 | 69.9 | **56** |
| **57** | 02.05 | 98 | 70.5 | 02.04 | 98 | 70.4 | 02.06 | 98 | 70.6 | 02.05 | 98 | 70.5 | 02.03 | 98 | 70.3 | 02.06 | 98 | 70.6 | **57** |
| **58** | 2.12 | 98 | 71.2 | 2.10 | 98 | 71.0 | 2.13 | 98 | 71.3 | 2.12 | 98 | 71.2 | 2.10 | 98 | 71.0 | 2.12 | 98 | 71.2 | **58** |
| **59** | 2.19 | 99 | 71.9 | 2.17 | 98 | 71.7 | 2.20 | 99 | 72.0 | 2.19 | 99 | 71.9 | 2.17 | 98 | 71.7 | 2.19 | 99 | 71.9 | **59** |
| **60** | 2.27 | 99 | 72.7 | 2.25 | 99 | 72.5 | 2.27 | 99 | 72.7 | 2.26 | 99 | 72.6 | 2.25 | 99 | 72.5 | 2.27 | 99 | 72.7 | **60** |

| SUM | Z | C | T | Z | C | T | Z | C | T | Z | C | T | Z | C | T | Z | C | T | SUM |
|---|---|---|---|---|---|---|---|---|---|---|---|---|---|---|---|---|---|---|---|
| | 6 to 9 years-old | | | 10 to 13 years | | | 14 to 18 years-old | | | 6 to 9 years-old | | | 10 to 13 years | | | 14 to 18 years-old | | | |
| | Male | | | | | | | | | Female | | | | | | | | | |

**Abbreviations**: C (Centile); T (T-score); Z (Z-score). **Notes:** Colored distribution bands: Green: minimal symptoms (T-score < 55). Yellow: mild symptoms (T-score ≥ 55 and < 60); Orange: moderate symptoms (T-score ≥ 60 and < 70); Red: severe symptoms (T-score ≥ 70).

**Supplementary Table 5 -** Modified Checklist for Autism in Toddlers (M-CHAT-R), Caregiver-report: normative references in Greece

| M-CHAT-R - Caregiver-report | | | | | | | |
|---|---|---|---|---|---|---|---|
| | Male | | | Female | | | |
| SUM | Z | C | T | Z | C | T | SUM |
| 0 | -0,67 | 0 | 43,3 | -0,68 | 0 | 43,2 | 0 |
| 1 | -0,33 | 5 | 46,7 | -0,34 | 5 | 46,6 | 1 |
| 2 | 0,08 | 10 | 50,8 | 0,07 | 10 | 50,7 | 2 |
| 3 | 0,51 | 15 | 55,1 | 0,50 | 15 | 55 | 3 |
| 4 | 0,88 | 20 | 58,8 | 0,87 | 20 | 58,7 | 4 |
| 5 | 1,16 | 25 | 61,6 | 1,15 | 25 | 61,5 | 5 |
| 6 | 1,38 | 30 | 63,8 | 1,37 | 30 | 63,7 | 6 |
| 7 | 1,54 | 35 | 65,4 | 1,53 | 35 | 65,3 | 7 |
| 8 | 1,67 | 40 | 66,7 | 1,66 | 40 | 66,6 | 8 |
| 9 | 1,80 | 45 | 68 | 1,79 | 45 | 67,9 | 9 |
| 10 | 1,92 | 50 | 69,2 | 1,91 | 50 | 69,1 | 10 |
| 11 | 2,05 | 55 | 70,5 | 2,04 | 55 | 70,4 | 11 |
| 12 | 2,19 | 60 | 71,9 | 2,18 | 60 | 71,8 | 12 |
| 13 | 2,34 | 65 | 73,4 | 2,33 | 65 | 73,3 | 13 |
| 14 | 2,51 | 70 | 75,1 | 2,50 | 70 | 75 | 14 |
| 15 | 2,69 | 75 | 76,9 | 2,68 | 75 | 76,8 | 15 |
| 16 | 2,87 | 80 | 78,7 | 2,86 | 80 | 78,6 | 16 |
| 17 | 3,05 | 85 | 80,5 | 3,04 | 85 | 80,4 | 17 |
| 18 | 3,23 | 90 | 82,3 | 3,22 | 90 | 82,2 | 18 |
| 19 | 3,41 | 95 | 84,1 | 3,40 | 95 | 84 | 19 |
| 20 | 3,59 | 100 | 85,9 | 3,58 | 100 | 85,8 | 20 |

**Abbreviations**: C (Centile); T (T-score); Z (Z-score). **Notes:** Colored distribution bands: Green: minimal symptoms (T-score < 55). Yellow: mild symptoms (T-score ≥ 55 and < 60); Orange: moderate symptoms (T-score ≥ 60 and < 70); Red: severe symptoms (T-score ≥ 70).

**Supplementary Table 6 -**Pediatric Symptom Checklist short version (PSC-17), age under 6 years, Caregiver-report: normative references in Greece

**PSC-17 (3 to 5 years-old) - Caregiver-report**

**Externalizing**

| | Male | | | Female | | | |
|---|---|---|---|---|---|---|---|
| SUM | Z | C | T | Z | C | T | SUM |
| 0 | -1,44 | 0 | 35,6 | -1,45 | 0 | 35,5 | 0 |
| 1 | -0,95 | 7 | 40,5 | -0,96 | 7 | 40,4 | 1 |
| 2 | -0,57 | 14 | 44,3 | -0,58 | 14 | 44,2 | 2 |
| 3 | -0,24 | 21 | 47,6 | -0,25 | 21 | 47,5 | 3 |
| 4 | 0,12 | 29 | 51,2 | 0,11 | 29 | 51,1 | 4 |
| 5 | 0,43 | 36 | 54,3 | 0,42 | 36 | 54,2 | 5 |
| 6 | 0,65 | 43 | 56,5 | 0,64 | 43 | 56,4 | 6 |
| 7 | 0,9 | 50 | 59 | 0,89 | 50 | 58,9 | 7 |
| 8 | 1,21 | 57 | 62,1 | 1,2 | 57 | 62 | 8 |
| 9 | 1,54 | 64 | 65,4 | 1,53 | 64 | 65,3 | 9 |
| 10 | 1,83 | 71 | 68,3 | 1,82 | 71 | 68,2 | 10 |
| 11 | 2,11 | 79 | 71,1 | 2,1 | 79 | 71 | 11 |
| 12 | 2,38 | 86 | 73,8 | 2,37 | 86 | 73,7 | 12 |
| 13 | 2,65 | 93 | 76,5 | 2,65 | 93 | 76,5 | 13 |

**Internalizing**

| | Male | | | Female | | | |
|---|---|---|---|---|---|---|---|
| SUM | Z | C | T | Z | C | T | SUM |
| 0 | -0,65 | 0 | 43,5 | -0,66 | 0 | 43,4 | 0 |
| 1 | 0,14 | 10 | 51,4 | 0,14 | 10 | 51,4 | 1 |
| 2 | 0,64 | 20 | 56,4 | 0,63 | 20 | 56,3 | 2 |
| 3 | 0,97 | 30 | 59,7 | 0,96 | 30 | 59,6 | 3 |
| 4 | 1,23 | 40 | 62,3 | 1,22 | 40 | 62,2 | 4 |
| 5 | 1,49 | 50 | 64,9 | 1,48 | 50 | 64,8 | 5 |
| 6 | 1,73 | 60 | 67,3 | 1,73 | 60 | 67,3 | 6 |
| 7 | 2,09 | 70 | 70,9 | 2,08 | 70 | 70,8 | 7 |
| 8 | 2,57 | 80 | 75,7 | 2,57 | 80 | 75,7 | 8 |
| 9 | 2,71 | 90 | 77,1 | 2,7 | 90 | 77 | 9 |
| 10 | 2,74 | 100 | 77,4 | 2,74 | 100 | 77,4 | 10 |

**Attention**

| | Male | | | Female | | | |
|---|---|---|---|---|---|---|---|
| SUM | Z | C | T | Z | C | T | SUM |
| 0 | -1.29 | 0 | 37.1 | -1.30 | 0 | 37.0 | 0 |
| 1 | -0.70 | 10 | 43.0 | -0.71 | 10 | 42.9 | 1 |
| 2 | -0.29 | 20 | 47.1 | -0.29 | 20 | 47.1 | 2 |
| 3 | 0.13 | 30 | 51.3 | 0.13 | 30 | 51.3 | 3 |
| 4 | 0.45 | 40 | 54.5 | 0.44 | 40 | 54.4 | 4 |
| 5 | 0.71 | 50 | 57.1 | 0.70 | 50 | 57.0 | 5 |
| 6 | 01.07 | 60 | 60.7 | 01.06 | 60 | 60.6 | 6 |
| 7 | 1.39 | 70 | 63.9 | 1.38 | 70 | 63.8 | 7 |
| 8 | 1.70 | 80 | 67.0 | 1.69 | 80 | 66.9 | 8 |
| 9 | 02.05 | 90 | 70.5 | 2.05 | 90 | 70.5 | 9 |
| 10 | 2.41 | 100 | 74.1 | 2.41 | 100 | 74.1 | 10 |

| | | | | | | | |
|---|---|---|---|---|---|---|---|
| **14** | 2,93 | 100 | 79,3 | 2,92 | 100 | 79,2 | **14** |

**Abbreviations**: C (Centile); T (T-score); Z (Z-score). **Notes:** Colored distribution bands: Green: minimal symptoms (T-score < 55). Yellow: mild symptoms (T-score ≥ 55 and < 60); Orange: moderate symptoms (T-score ≥ 60 and < 70); Red: severe symptoms (T-score ≥ 70).

**Supplementary Table 7 -** Pediatric Symptom Checklist Short Version (PSC-17), 6- to 18-year-olds, Caregiver-report: normative references in Greece

| PSC-17 (6 to 18 years-old) - Caregiver-report | | | | | | | | | | | | | | | | | | | |
|---|---|---|---|---|---|---|---|---|---|---|---|---|---|---|---|---|---|---|---|
| EXTERNALIZING SCALE | | | | | | | | | | | | | | | | | | | |
| | Male | | | | | | | | | Female | | | | | | | | | |
| | 6 to 9 years-old | | | 10 to 13 years | | | 14 to 18 years-old | | | 6 to 9 years-old | | | 10 to 13 years | | | 14 to 18 years-old | | | |
| SUM | Z | C | T | Z | C | T | Z | C | T | Z | C | T | Z | C | T | Z | C | T | SUM |
| 0 | -1,15 | 13 | 38,5 | -1,14 | 13 | 38,6 | -1,15 | 13 | 38,5 | -1,14 | 13 | 38,6 | -1,13 | 13 | 38,7 | -1,13 | 13 | 38,7 | 0 |
| 1 | -0,58 | 28 | 44,2 | -0,57 | 28 | 44,3 | -0,57 | 28 | 44,3 | -0,56 | 29 | 44,4 | -0,55 | 29 | 44,5 | -0,56 | 29 | 44,4 | 1 |
| 2 | -0,15 | 44 | 48,5 | -0,15 | 44 | 48,5 | -0,15 | 44 | 48,5 | -0,14 | 44 | 48,6 | -0,13 | 45 | 48,7 | -0,14 | 44 | 48,6 | 2 |
| 3 | 0,14 | 56 | 51,4 | 0,15 | 56 | 51,5 | 0,15 | 56 | 51,5 | 0,16 | 56 | 51,6 | 0,17 | 57 | 51,7 | 0,16 | 56 | 51,6 | 3 |
| 4 | 0,42 | 66 | 54,2 | 0,43 | 67 | 54,3 | 0,42 | 66 | 54,2 | 0,44 | 67 | 54,4 | 0,45 | 67 | 54,5 | 0,44 | 67 | 54,4 | 4 |
| 5 | 0,69 | 75 | 56,9 | 0,7 | 76 | 57 | 0,69 | 75 | 56,9 | 0,7 | 76 | 57 | 0,71 | 76 | 57,1 | 0,71 | 76 | 57,1 | 5 |
| 6 | 0,94 | 83 | 59,4 | 0,95 | 83 | 59,5 | 0,94 | 83 | 59,4 | 0,95 | 83 | 59,5 | 0,96 | 83 | 59,6 | 0,96 | 83 | 59,6 | 6 |
| 7 | 1,19 | 88 | 61,9 | 1,2 | 88 | 62 | 1,2 | 88 | 62 | 1,21 | 89 | 62,1 | 1,22 | 89 | 62,2 | 1,21 | 89 | 62,1 | 7 |
| 8 | 1,44 | 93 | 64,4 | 1,45 | 93 | 64,5 | 1,44 | 93 | 64,4 | 1,45 | 93 | 64,5 | 1,46 | 93 | 64,6 | 1,46 | 93 | 64,6 | 8 |
| 9 | 1,67 | 95 | 66,7 | 1,68 | 95 | 66,8 | 1,67 | 95 | 66,7 | 1,68 | 95 | 66,8 | 1,69 | 95 | 66,9 | 1,69 | 95 | 66,9 | 9 |
| 10 | 1,92 | 97 | 69,2 | 1,92 | 97 | 69,2 | 1,92 | 97 | 69,2 | 1,93 | 97 | 69,3 | 1,94 | 97 | 69,4 | 1,93 | 97 | 69,3 | 10 |
| 11 | 2,19 | 99 | 71,9 | 2,2 | 99 | 72 | 2,19 | 99 | 71,9 | 2,2 | 99 | 72 | 2,21 | 99 | 72,1 | 2,21 | 99 | 72,1 | 11 |
| 12 | 2,47 | 99 | 74,7 | 2,48 | 99 | 74,8 | 2,47 | 99 | 74,7 | 2,48 | 99 | 74,8 | 2,49 | 99 | 74,9 | 2,48 | 99 | 74,8 | 12 |
| 13 | 2,75 | 100 | 77,5 | 2,76 | 100 | 77,6 | 2,75 | 100 | 77,5 | 2,76 | 100 | 77,6 | 2,77 | 100 | 77,7 | 2,76 | 100 | 77,6 | 13 |
| 14 | 3,05 | 100 | 80,5 | 3,06 | 100 | 80,6 | 3,05 | 100 | 80,5 | 3,06 | 100 | 80,6 | 3,07 | 100 | 80,7 | 3,06 | 100 | 80,6 | 14 |
| INTERNALIZING SCALE | | | | | | | | | | | | | | | | | | | |

| | Male | | | | | | | | | Female | | | | | | | | | |
|---|---|---|---|---|---|---|---|---|---|---|---|---|---|---|---|---|---|---|---|
| | 6 to 9 years-old | | | 10 to 13 years | | | 14 to 18 years-old | | | 6 to 9 years-old | | | 10 to 13 years | | | 14 to 18 years-old | | | |
| **SUM** | **Z** | **C** | **T** | **Z** | **C** | **T** | **Z** | **C** | **T** | **Z** | **C** | **T** | **Z** | **C** | **T** | **Z** | **C** | **T** | **SUM** |
| **0** | -0,91 | 18 | 40,9 | -0,92 | 18 | 40,8 | -0,92 | 18 | 40,8 | -0,91 | 18 | 40,9 | -0,92 | 18 | 40,8 | -0,92 | 18 | 40,8 | **0** |
| **1** | -0,21 | 42 | 47,9 | -0,21 | 42 | 47,9 | -0,21 | 42 | 47,9 | -0,21 | 42 | 47,9 | -0,21 | 42 | 47,9 | -0,21 | 42 | 47,9 | **1** |
| **2** | 0,22 | 59 | 52,2 | 0,21 | 58 | 52,1 | 0,21 | 58 | 52,1 | 0,22 | 59 | 52,2 | 0,21 | 58 | 52,1 | 0,21 | 58 | 52,1 | **2** |
| **3** | 0,58 | 72 | 55,8 | 0,57 | 72 | 55,7 | 0,57 | 72 | 55,7 | 0,58 | 72 | 55,8 | 0,57 | 72 | 55,7 | 0,58 | 72 | 55,8 | **3** |
| **4** | 0,89 | 81 | 58,9 | 0,89 | 81 | 58,9 | 0,89 | 81 | 58,9 | 0,9 | 82 | 59 | 0,89 | 81 | 58,9 | 0,89 | 81 | 58,9 | **4** |
| **5** | 1,2 | 88 | 62 | 1,19 | 88 | 61,9 | 1,2 | 88 | 62 | 1,2 | 88 | 62 | 1,2 | 88 | 62 | 1,2 | 88 | 62 | **5** |
| **6** | 1,52 | 94 | 65,2 | 1,51 | 93 | 65,1 | 1,52 | 94 | 65,2 | 1,52 | 94 | 65,2 | 1,52 | 94 | 65,2 | 1,52 | 94 | 65,2 | **6** |
| **7** | 1,81 | 96 | 68,1 | 1,8 | 96 | 68 | 1,8 | 96 | 68 | 1,81 | 96 | 68,1 | 1,8 | 96 | 68 | 1,8 | 96 | 68 | **7** |
| **8** | 2,13 | 98 | 71,3 | 2,13 | 98 | 71,3 | 2,13 | 98 | 71,3 | 2,14 | 98 | 71,4 | 2,13 | 98 | 71,3 | 2,13 | 98 | 71,3 | **8** |
| **9** | 2,48 | 99 | 74,8 | 2,48 | 99 | 74,8 | 2,48 | 99 | 74,8 | 2,48 | 99 | 74,8 | 2,48 | 99 | 74,8 | 2,48 | 99 | 74,8 | **9** |
| **10** | 2,86 | 100 | 78,6 | 2,86 | 100 | 78,6 | 2,86 | 100 | 78,6 | 2,86 | 100 | 78,6 | 2,86 | 100 | 78,6 | 2,86 | 100 | 78,6 | **10** |

**ATTENTION SCALE**

| | Male | | | | | | | | | Female | | | | | | | | | |
|---|---|---|---|---|---|---|---|---|---|---|---|---|---|---|---|---|---|---|---|
| | 6 to 9 years-old | | | 10 to 13 years | | | 14 to 18 years-old | | | 6 to 9 years-old | | | 10 to 13 years | | | 14 to 18 years-old | | | |
| **SUM** | **Z** | **C** | **T** | **Z** | **C** | **T** | **Z** | **C** | **T** | **Z** | **C** | **T** | **Z** | **C** | **T** | **Z** | **C** | **T** | **SUM** |
| **0** | -1.10 | 14 | 39.0 | -1.12 | 13 | 38.8 | -1.13 | 13 | 38.7 | -1.13 | 13 | 38.7 | -1.14 | 13 | 38.6 | -1.15 | 13 | 38.5 | **0** |
| **1** | -0.49 | 31 | 45.1 | -0.50 | 31 | 45.0 | -0.51 | 31 | 44.9 | -0.51 | 31 | 44.9 | -0.53 | 30 | 44.7 | -0.54 | 29 | 44.6 | **1** |
| **2** | 0.00 | 50 | 50.0 | -0.01 | 50 | 49.9 | -0.02 | 49 | 49.8 | -0.02 | 49 | 49.8 | -0.04 | 48 | 49.6 | -0.05 | 48 | 49.5 | **2** |
| **3** | 0.32 | 63 | 53.2 | 0.31 | 62 | 53.1 | 0.30 | 62 | 53.0 | 0.30 | 62 | 53.0 | 0.28 | 61 | 52.8 | 0.27 | 61 | 52.7 | **3** |
| **4** | 0.61 | 73 | 56.1 | 0.60 | 73 | 56.0 | 0.59 | 72 | 55.9 | 0.58 | 72 | 55.8 | 0.57 | 72 | 55.7 | 0.56 | 71 | 55.6 | **4** |

| | | | | | | | | | | | | | | | | | | |
|---|---|---|---|---|---|---|---|---|---|---|---|---|---|---|---|---|---|---|
| **5** | 0.94 | 83 | 59.4 | 0.92 | 82 | 59.2 | 0.91 | 82 | 59.1 | 0.91 | 82 | 59.1 | 0.89 | 81 | 58.9 | 0.88 | 81 | 58.8 | **5** |
| **6** | 1.26 | 90 | 62.6 | 1.24 | 89 | 62.4 | 1.23 | 89 | 62.3 | 1.23 | 89 | 62.3 | 1.22 | 89 | 62.2 | 1.21 | 89 | 62.1 | **6** |
| **7** | 1.56 | 94 | 65.6 | 1.55 | 94 | 65.5 | 1.54 | 94 | 65.4 | 1.53 | 94 | 65.3 | 1.52 | 94 | 65.2 | 1.51 | 93 | 65.1 | **7** |
| **8** | 1.84 | 97 | 68.4 | 1.83 | 97 | 68.3 | 1.82 | 97 | 68.2 | 1.81 | 96 | 68.1 | 1.80 | 96 | 68.0 | 1.79 | 96 | 67.9 | **8** |
| **9** | 2.12 | 98 | 71.2 | 2.11 | 98 | 71.1 | 2.10 | 98 | 71.0 | 02.09 | 98 | 70.9 | 02.08 | 98 | 70.8 | 02.07 | 98 | 70.7 | **9** |
| **10** | 2.46 | 99 | 74.6 | 2.44 | 99 | 74.4 | 2.43 | 99 | 74.3 | 2.43 | 99 | 74.3 | 2.42 | 99 | 74.2 | 2.41 | 99 | 74.1 | **10** |

**Abbreviations**: C (Centile); T (T-score); Z (Z-score). **Notes:** Colored distribution bands: Green: minimal symptoms (T-score < 55). Yellow: mild symptoms (T-score ≥ 55 and < 60); Orange: moderate symptoms (T-score ≥ 60 and < 70); Red: severe symptoms (T-score ≥ 70).

**Supplementary Table 8 -** Revised Children's Anxiety and Depression Scale short-version (RCADS-25), Caregiver-report: normative references in Greece

| RCADS-25 - Caregiver-report | | | | | | | | | | | | | | | | | | | |
|---|---|---|---|---|---|---|---|---|---|---|---|---|---|---|---|---|---|---|---|
| ANXIETY SCALE | | | | | | | | | | | | | | | | | | | |
| | Male | | | | | | | | | Female | | | | | | | | | |
| | 6 to 9 years-old | | | 10 to 13 years | | | 14 to 18 years-old | | | 6 to 9 years-old | | | 10 to 13 years | | | 14 to 18 years-old | | | |
| SUM | Z | C | T | Z | C | T | Z | C | T | Z | C | T | Z | C | T | Z | C | T | SUM |
| 0 | -1.29 | 10 | 37.1 | -1.22 | 11 | 37.8 | -1.36 | 9 | 36.4 | -1.30 | 10 | 37.0 | -1.22 | 11 | 37.8 | -1.37 | 9 | 36.3 | 0 |
| 1 | -0.89 | 19 | 41.1 | -0.82 | 21 | 41.8 | -0.97 | 17 | 40.3 | -0.90 | 18 | 41.0 | -0.83 | 20 | 41.7 | -0.98 | 16 | 40.2 | 1 |
| 2 | -0.52 | 30 | 44.8 | -0.45 | 33 | 45.5 | -0.60 | 27 | 44.0 | -0.53 | 30 | 44.7 | -0.46 | 32 | 45.4 | -0.61 | 27 | 43.9 | 2 |
| 3 | -0.20 | 42 | 48.0 | -0.13 | 45 | 48.7 | -0.28 | 39 | 47.2 | -0.21 | 42 | 47.9 | -0.14 | 44 | 48.6 | -0.29 | 39 | 47.1 | 3 |
| 4 | 0.05 | 52 | 50.5 | 0.12 | 55 | 51.2 | -0.03 | 49 | 49.7 | 0.04 | 52 | 50.4 | 0.11 | 54 | 51.1 | -0.04 | 48 | 49.6 | 4 |
| 5 | 0.24 | 59 | 52.4 | 0.31 | 62 | 53.1 | 0.16 | 56 | 51.6 | 0.23 | 59 | 52.3 | 0.30 | 62 | 53.0 | 0.15 | 56 | 51.5 | 5 |
| 6 | 0.39 | 65 | 53.9 | 0.46 | 68 | 54.6 | 0.31 | 62 | 53.1 | 0.38 | 65 | 53.8 | 0.45 | 67 | 54.5 | 0.30 | 62 | 53.0 | 6 |
| 7 | 0.52 | 70 | 55.2 | 0.59 | 72 | 55.9 | 0.44 | 67 | 54.4 | 0.51 | 69 | 55.1 | 0.58 | 72 | 55.8 | 0.44 | 67 | 54.4 | 7 |
| 8 | 0.65 | 74 | 56.5 | 0.72 | 76 | 57.2 | 0.58 | 72 | 55.8 | 0.64 | 74 | 56.4 | 0.72 | 76 | 57.2 | 0.57 | 72 | 55.7 | 8 |
| 9 | 0.78 | 78 | 57.8 | 0.86 | 81 | 58.6 | 0.71 | 76 | 57.1 | 0.78 | 78 | 57.8 | 0.85 | 80 | 58.5 | 0.70 | 76 | 57.0 | 9 |
| 10 | 0.92 | 82 | 59.2 | 0.99 | 84 | 59.9 | 0.84 | 80 | 58.4 | 0.91 | 82 | 59.1 | 0.98 | 84 | 59.8 | 0.83 | 80 | 58.3 | 10 |

| | | | | | | | | | | | | | | | | | | |
|---|---|---|---|---|---|---|---|---|---|---|---|---|---|---|---|---|---|---|
| **11** | 01.03 | 85 | 60.3 | 1.11 | 87 | 61.1 | 0.96 | 83 | 59.6 | 01.03 | 85 | 60.3 | 1.10 | 86 | 61.0 | 0.95 | 83 | 59.5 | **11** |
| **12** | 1.13 | 87 | 61.3 | 1.20 | 88 | 62.0 | 01.06 | 86 | 60.6 | 1.12 | 87 | 61.2 | 1.20 | 88 | 62.0 | 01.05 | 85 | 60.5 | **12** |
| **13** | 1.21 | 89 | 62.1 | 1.28 | 90 | 62.8 | 1.14 | 87 | 61.4 | 1.20 | 88 | 62.0 | 1.28 | 90 | 62.8 | 1.13 | 87 | 61.3 | **13** |
| **14** | 1.28 | 90 | 62.8 | 1.35 | 91 | 63.5 | 1.20 | 88 | 62.0 | 1.27 | 90 | 62.7 | 1.34 | 91 | 63.4 | 1.19 | 88 | 61.9 | **14** |
| **15** | 1.34 | 91 | 63.4 | 1.42 | 92 | 64.2 | 1.27 | 90 | 62.7 | 1.34 | 91 | 63.4 | 1.41 | 92 | 64.1 | 1.26 | 90 | 62.6 | **15** |
| **16** | 1.41 | 92 | 64.1 | 1.49 | 93 | 64.9 | 1.34 | 91 | 63.4 | 1.41 | 92 | 64.1 | 1.48 | 93 | 64.8 | 1.33 | 91 | 63.3 | **16** |
| **17** | 1.49 | 93 | 64.9 | 1.57 | 94 | 65.7 | 1.42 | 92 | 64.2 | 1.49 | 93 | 64.9 | 1.56 | 94 | 65.6 | 1.41 | 92 | 64.1 | **17** |
| **18** | 1.58 | 94 | 65.8 | 1.65 | 95 | 66.5 | 1.51 | 93 | 65.1 | 1.57 | 94 | 65.7 | 1.65 | 95 | 66.5 | 1.50 | 93 | 65.0 | **18** |
| **19** | 1.68 | 95 | 66.8 | 1.75 | 96 | 67.5 | 1.60 | 95 | 66.0 | 1.67 | 95 | 66.7 | 1.74 | 96 | 67.4 | 1.59 | 94 | 65.9 | **19** |
| **20** | 1.77 | 96 | 67.7 | 1.84 | 97 | 68.4 | 1.70 | 96 | 67.0 | 1.76 | 96 | 67.6 | 1.84 | 97 | 68.4 | 1.69 | 95 | 66.9 | **20** |
| **21** | 1.87 | 97 | 68.7 | 1.94 | 97 | 69.4 | 1.79 | 96 | 67.9 | 1.86 | 97 | 68.6 | 1.93 | 97 | 69.3 | 1.79 | 96 | 67.9 | **21** |
| **22** | 1.97 | 98 | 69.7 | 02.04 | 98 | 70.4 | 1.89 | 97 | 68.9 | 1.96 | 98 | 69.6 | 02.03 | 98 | 70.3 | 1.88 | 97 | 68.8 | **22** |
| **23** | 02.06 | 98 | 70.6 | 2.14 | 98 | 71.4 | 1.99 | 98 | 69.9 | 02.06 | 98 | 70.6 | 2.13 | 98 | 71.3 | 1.98 | 98 | 69.8 | **23** |
| **24** | 2.16 | 98 | 71.6 | 2.23 | 99 | 72.3 | 02.08 | 98 | 70.8 | 2.15 | 98 | 71.5 | 2.22 | 99 | 72.2 | 02.07 | 98 | 70.7 | **24** |
| **25** | 2.25 | 99 | 72.5 | 2.32 | 99 | 73.2 | 2.17 | 98 | 71.7 | 2.24 | 99 | 72.4 | 2.31 | 99 | 73.1 | 2.16 | 98 | 71.6 | **25** |
| **26** | 2.33 | 99 | 73.3 | 2.40 | 99 | 74.0 | 2.25 | 99 | 72.5 | 2.32 | 99 | 73.2 | 2.39 | 99 | 73.9 | 2.24 | 99 | 72.4 | **26** |
| **27** | 2.39 | 99 | 73.9 | 2.47 | 99 | 74.7 | 2.32 | 99 | 73.2 | 2.39 | 99 | 73.9 | 2.46 | 99 | 74.6 | 2.31 | 99 | 73.1 | **27** |
| **28** | 2.46 | 99 | 74.6 | 2.53 | 99 | 75.3 | 2.38 | 99 | 73.8 | 2.45 | 99 | 74.5 | 2.52 | 99 | 75.2 | 2.37 | 99 | 73.7 | **28** |

| | | | | | | | | | | | | | | | | | | | |
|---|---|---|---|---|---|---|---|---|---|---|---|---|---|---|---|---|---|---|---|
| **29** | 2.51 | 99 | 75.1 | 2.59 | 100 | 75.9 | 2.44 | 99 | 74.4 | 2.51 | 99 | 75.1 | 2.58 | 100 | 75.8 | 2.43 | 99 | 74.3 | **29** |
| **30** | 2.57 | 99 | 75.7 | 2.65 | 100 | 76.5 | 2.50 | 99 | 75.0 | 2.57 | 99 | 75.7 | 2.64 | 100 | 76.4 | 2.49 | 99 | 74.9 | **30** |
| **31** | 2.64 | 100 | 76.4 | 2.71 | 100 | 77.1 | 2.56 | 99 | 75.6 | 2.63 | 100 | 76.3 | 2.70 | 100 | 77.0 | 2.55 | 99 | 75.5 | **31** |
| **32** | 2.71 | 100 | 77.1 | 2.78 | 100 | 77.8 | 2.63 | 100 | 76.3 | 2.70 | 100 | 77.0 | 2.77 | 100 | 77.7 | 2.62 | 100 | 76.2 | **32** |
| **33** | 2.78 | 100 | 77.8 | 2.86 | 100 | 78.6 | 2.71 | 100 | 77.1 | 2.78 | 100 | 77.8 | 2.85 | 100 | 78.5 | 2.70 | 100 | 77.0 | **33** |
| **34** | 2.86 | 100 | 78.6 | 2.93 | 100 | 79.3 | 2.78 | 100 | 77.8 | 2.85 | 100 | 78.5 | 2.93 | 100 | 79.3 | 2.78 | 100 | 77.8 | **34** |
| **35** | 2.94 | 100 | 79.4 | 03.01 | 100 | 80.1 | 2.86 | 100 | 78.6 | 2.93 | 100 | 79.3 | 3.00 | 100 | 80.0 | 2.85 | 100 | 78.5 | **35** |
| **36** | 03.02 | 100 | 80.2 | 03.09 | 100 | 80.9 | 2.94 | 100 | 79.4 | 03.01 | 100 | 80.1 | 03.08 | 100 | 80.8 | 2.93 | 100 | 79.3 | **36** |
| **37** | 03.09 | 100 | 80.9 | 3.17 | 100 | 81.7 | 03.02 | 100 | 80.2 | 03.09 | 100 | 80.9 | 3.16 | 100 | 81.6 | 03.01 | 100 | 80.1 | **37** |
| **38** | 3.17 | 100 | 81.7 | 3.24 | 100 | 82.4 | 3.10 | 100 | 81.0 | 3.16 | 100 | 81.6 | 3.24 | 100 | 82.4 | 03.09 | 100 | 80.9 | **38** |
| **39** | 3.25 | 100 | 82.5 | 3.32 | 100 | 83.2 | 3.17 | 100 | 81.7 | 3.24 | 100 | 82.4 | 3.31 | 100 | 83.1 | 3.17 | 100 | 81.7 | **39** |
| **40** | 3.33 | 100 | 83.3 | 3.40 | 100 | 84.0 | 3.25 | 100 | 82.5 | 3.32 | 100 | 83.2 | 3.39 | 100 | 83.9 | 3.24 | 100 | 82.4 | **40** |
| **41** | 3.40 | 100 | 84.0 | 3.48 | 100 | 84.8 | 3.33 | 100 | 83.3 | 3.40 | 100 | 84.0 | 3.47 | 100 | 84.7 | 3.32 | 100 | 83.2 | **41** |
| **42** | 3.48 | 100 | 84.8 | 3.55 | 100 | 85.5 | 3.41 | 100 | 84.1 | 3.47 | 100 | 84.7 | 3.55 | 100 | 85.5 | 3.40 | 100 | 84.0 | **42** |
| **43** | 3.56 | 100 | 85.6 | 3.63 | 100 | 86.3 | 3.48 | 100 | 84.8 | 3.55 | 100 | 85.5 | 3.62 | 100 | 86.2 | 3.48 | 100 | 84.8 | **43** |
| **44** | 3.64 | 100 | 86.4 | 3.71 | 100 | 87.1 | 3.56 | 100 | 85.6 | 3.63 | 100 | 86.3 | 3.70 | 100 | 87.0 | 3.55 | 100 | 85.5 | **44** |
| **45** | 3.72 | 100 | 87.2 | 3.79 | 100 | 87.9 | 3.64 | 100 | 86.4 | 3.71 | 100 | 87.1 | 3.78 | 100 | 87.8 | 3.63 | 100 | 86.3 | **45** |
| **DEPRESSION SCALE** | | | | | | | | | | | | | | | | | | | |
| | **Male** | | | | | | | | | **Female** | | | | | | | | | |

| | 6 to 9 years-old | | | 10 to 13 years | | | 14 to 18 years-old | | | 6 to 9 years-old | | | 10 to 13 years | | | 14 to 18 years-old | | | |
|---|---|---|---|---|---|---|---|---|---|---|---|---|---|---|---|---|---|---|---|
| **SUM** | **Z** | **C** | **T** | **Z** | **C** | **T** | **Z** | **C** | **T** | **Z** | **C** | **T** | **Z** | **C** | **T** | **Z** | **C** | **T** | **SUM** |
| **0** | -0.93 | 18 | 40.7 | -0.94 | 17 | 40.6 | -0.95 | 17 | 40.5 | -0.91 | 18 | 40.9 | -0.92 | 18 | 40.8 | -0.93 | 18 | 40.7 | **0** |
| **1** | -0.41 | 34 | 45.9 | -0.42 | 34 | 45.8 | -0.43 | 33 | 45.7 | -0.39 | 35 | 46.1 | -0.40 | 34 | 46.0 | -0.41 | 34 | 45.9 | **1** |
| **2** | 0.03 | 51 | 50.3 | 0.02 | 51 | 50.2 | 0.01 | 50 | 50.1 | 0.05 | 52 | 50.5 | 0.04 | 52 | 50.4 | 0.03 | 51 | 50.3 | **2** |
| **3** | 0.33 | 63 | 53.3 | 0.31 | 62 | 53.1 | 0.31 | 62 | 53.1 | 0.34 | 63 | 53.4 | 0.33 | 63 | 53.3 | 0.32 | 63 | 53.2 | **3** |
| **4** | 0.50 | 69 | 55.0 | 0.49 | 69 | 54.9 | 0.48 | 68 | 54.8 | 0.52 | 70 | 55.2 | 0.50 | 69 | 55.0 | 0.50 | 69 | 55.0 | **4** |
| **5** | 0.62 | 73 | 56.2 | 0.61 | 73 | 56.1 | 0.60 | 73 | 56.0 | 0.64 | 74 | 56.4 | 0.63 | 74 | 56.3 | 0.62 | 73 | 56.2 | **5** |
| **6** | 0.76 | 78 | 57.6 | 0.74 | 77 | 57.4 | 0.74 | 77 | 57.4 | 0.77 | 78 | 57.7 | 0.76 | 78 | 57.6 | 0.75 | 77 | 57.5 | **6** |
| **7** | 0.92 | 82 | 59.2 | 0.91 | 82 | 59.1 | 0.90 | 82 | 59.0 | 0.94 | 83 | 59.4 | 0.93 | 82 | 59.3 | 0.92 | 82 | 59.2 | **7** |
| **8** | 01.09 | 86 | 60.9 | 01.08 | 86 | 60.8 | 01.07 | 86 | 60.7 | 1.11 | 87 | 61.1 | 1.10 | 86 | 61.0 | 01.09 | 86 | 60.9 | **8** |
| **9** | 1.22 | 89 | 62.2 | 1.21 | 89 | 62.1 | 1.20 | 88 | 62.0 | 1.24 | 89 | 62.4 | 1.23 | 89 | 62.3 | 1.22 | 89 | 62.2 | **9** |
| **10** | 1.32 | 91 | 63.2 | 1.31 | 90 | 63.1 | 1.30 | 90 | 63.0 | 1.34 | 91 | 63.4 | 1.33 | 91 | 63.3 | 1.32 | 91 | 63.2 | **10** |
| **11** | 1.42 | 92 | 64.2 | 1.41 | 92 | 64.1 | 1.40 | 92 | 64.0 | 1.44 | 93 | 64.4 | 1.42 | 92 | 64.2 | 1.42 | 92 | 64.2 | **11** |
| **12** | 1.54 | 94 | 65.4 | 1.53 | 94 | 65.3 | 1.52 | 94 | 65.2 | 1.56 | 94 | 65.6 | 1.55 | 94 | 65.5 | 1.54 | 94 | 65.4 | **12** |
| **13** | 1.69 | 95 | 66.9 | 1.68 | 95 | 66.8 | 1.67 | 95 | 66.7 | 1.71 | 96 | 67.1 | 1.70 | 96 | 67.0 | 1.69 | 95 | 66.9 | **13** |
| **14** | 1.82 | 97 | 68.2 | 1.81 | 96 | 68.1 | 1.81 | 96 | 68.1 | 1.84 | 97 | 68.4 | 1.83 | 97 | 68.3 | 1.82 | 97 | 68.2 | **14** |
| **15** | 1.92 | 97 | 69.2 | 1.90 | 97 | 69.0 | 1.90 | 97 | 69.0 | 1.93 | 97 | 69.3 | 1.92 | 97 | 69.2 | 1.91 | 97 | 69.1 | **15** |
| **16** | 1.97 | 98 | 69.7 | 1.96 | 98 | 69.6 | 1.95 | 97 | 69.5 | 1.99 | 98 | 69.9 | 1.98 | 98 | 69.8 | 1.97 | 98 | 69.7 | **16** |

| | | | | | | | | | | | | | | | | | | | |
|---|---|---|---|---|---|---|---|---|---|---|---|---|---|---|---|---|---|---|---|
| **17** | 02.04 | 98 | 70.4 | 02.03 | 98 | 70.3 | 02.02 | 98 | 70.2 | 02.05 | 98 | 70.5 | 02.04 | 98 | 70.4 | 02.04 | 98 | 70.4 | **17** |
| **18** | 2.15 | 98 | 71.5 | 2.14 | 98 | 71.4 | 2.14 | 98 | 71.4 | 2.17 | 98 | 71.7 | 2.16 | 98 | 71.6 | 2.15 | 98 | 71.5 | **18** |
| **19** | 2.32 | 99 | 73.2 | 2.31 | 99 | 73.1 | 2.31 | 99 | 73.1 | 2.34 | 99 | 73.4 | 2.33 | 99 | 73.3 | 2.32 | 99 | 73.2 | **19** |
| **20** | 2.50 | 99 | 75.0 | 2.49 | 99 | 74.9 | 2.48 | 99 | 74.8 | 2.52 | 99 | 75.2 | 2.51 | 99 | 75.1 | 2.50 | 99 | 75.0 | **20** |
| **21** | 2.62 | 100 | 76.2 | 2.61 | 100 | 76.1 | 2.60 | 100 | 76.0 | 2.64 | 100 | 76.4 | 2.63 | 100 | 76.3 | 2.62 | 100 | 76.2 | **21** |
| **22** | 2.68 | 100 | 76.8 | 2.67 | 100 | 76.7 | 2.66 | 100 | 76.6 | 2.70 | 100 | 77.0 | 2.69 | 100 | 76.9 | 2.68 | 100 | 76.8 | **22** |
| **23** | 2.72 | 100 | 77.2 | 2.71 | 100 | 77.1 | 2.70 | 100 | 77.0 | 2.74 | 100 | 77.4 | 2.72 | 100 | 77.2 | 2.72 | 100 | 77.2 | **23** |
| **24** | 2.79 | 100 | 77.9 | 2.78 | 100 | 77.8 | 2.77 | 100 | 77.7 | 2.81 | 100 | 78.1 | 2.79 | 100 | 77.9 | 2.79 | 100 | 77.9 | **24** |
| **25** | 2.92 | 100 | 79.2 | 2.91 | 100 | 79.1 | 2.90 | 100 | 79.0 | 2.94 | 100 | 79.4 | 2.93 | 100 | 79.3 | 2.92 | 100 | 79.2 | **25** |
| **26** | 3.10 | 100 | 81.0 | 03.09 | 100 | 80.9 | 03.08 | 100 | 80.8 | 3.12 | 100 | 81.2 | 3.11 | 100 | 81.1 | 3.10 | 100 | 81.0 | **26** |
| **27** | 3.30 | 100 | 83.0 | 3.29 | 100 | 82.9 | 3.29 | 100 | 82.9 | 3.32 | 100 | 83.2 | 3.31 | 100 | 83.1 | 3.30 | 100 | 83.0 | **27** |
| **28** | 3.51 | 100 | 85.1 | 3.50 | 100 | 85.0 | 3.49 | 100 | 84.9 | 3.53 | 100 | 85.3 | 3.52 | 100 | 85.2 | 3.51 | 100 | 85.1 | **28** |
| **29** | 3.72 | 100 | 87.2 | 3.71 | 100 | 87.1 | 3.70 | 100 | 87.0 | 3.74 | 100 | 87.4 | 3.73 | 100 | 87.3 | 3.72 | 100 | 87.2 | **29** |
| **30** | 3.93 | 100 | 89.3 | 3.92 | 100 | 89.2 | 3.91 | 100 | 89.1 | 3.95 | 100 | 89.5 | 3.93 | 100 | 89.3 | 3.93 | 100 | 89.3 | **30** |

Abbreviations: C (Centile); T (T-score); Z (Z-score). Notes: Colored distribution bands: Green: minimal symptoms (T-score < 55). Yellow: mild symptoms (T-score ≥ 55 and < 60); Orange: moderate symptoms (T-score ≥ 60 and < 70); Red: severe symptoms (T-score ≥ 70).

Supplementary Table 9 - Shortened Revised Children's Anxiety and Depression Scale (RCADS-25), Self-report: normative references in Greece

| RCADS-25 - Self-report | | | | | | | | | | | | | | | | | | | |
|---|---|---|---|---|---|---|---|---|---|---|---|---|---|---|---|---|---|---|---|
| ANXIETY SCALE | | | | | | | | | | | | | | | | | | | |
| | Male | | | | | | | | | Female | | | | | | | | | |
| | 6 to 9 years-old | | | 10 to 13 years | | | 14 to 18 years-old | | | 6 to 9 years-old | | | 10 to 13 years | | | 14 to 18 years-old | | | |
| SUM | Z | C | T | Z | C | T | Z | C | T | Z | C | T | Z | C | T | Z | C | T | SUM |
| 0 | -1.25 | 11 | 37.5 | -1.22 | 11 | 37.8 | -1.24 | 11 | 37.6 | -1.27 | 10 | 37.3 | -1.24 | 11 | 37.6 | -1.26 | 10 | 37.4 | 0 |
| 1 | -0.86 | 19 | 41.4 | -0.83 | 20 | 41.7 | -0.84 | 20 | 41.6 | -0.88 | 19 | 41.2 | -0.85 | 20 | 41.5 | -0.86 | 19 | 41.4 | 1 |
| 2 | -0.49 | 31 | 45.1 | -0.46 | 32 | 45.4 | -0.47 | 32 | 45.3 | -0.51 | 31 | 44.9 | -0.48 | 32 | 45.2 | -0.49 | 31 | 45.1 | 2 |
| 3 | -0.17 | 43 | 48.3 | -0.14 | 44 | 48.6 | -0.16 | 44 | 48.4 | -0.19 | 42 | 48.1 | -0.16 | 44 | 48.4 | -0.18 | 43 | 48.2 | 3 |
| 4 | 0.07 | 53 | 50.7 | 0.10 | 54 | 51.0 | 0.08 | 53 | 50.8 | 0.05 | 52 | 50.5 | 0.08 | 53 | 50.8 | 0.06 | 52 | 50.6 | 4 |
| 5 | 0.23 | 59 | 52.3 | 0.26 | 60 | 52.6 | 0.25 | 60 | 52.5 | 0.21 | 58 | 52.1 | 0.24 | 59 | 52.4 | 0.23 | 59 | 52.3 | 5 |
| 6 | 0.33 | 63 | 53.3 | 0.36 | 64 | 53.6 | 0.35 | 64 | 53.5 | 0.31 | 62 | 53.1 | 0.34 | 63 | 53.4 | 0.33 | 63 | 53.3 | 6 |
| 7 | 0.40 | 66 | 54.0 | 0.43 | 67 | 54.3 | 0.42 | 66 | 54.2 | 0.38 | 65 | 53.8 | 0.41 | 66 | 54.1 | 0.40 | 66 | 54.0 | 7 |
| 8 | 0.47 | 68 | 54.7 | 0.49 | 69 | 54.9 | 0.48 | 68 | 54.8 | 0.45 | 67 | 54.5 | 0.48 | 68 | 54.8 | 0.46 | 68 | 54.6 | 8 |
| 9 | 0.55 | 71 | 55.5 | 0.57 | 72 | 55.7 | 0.56 | 71 | 55.6 | 0.53 | 70 | 55.3 | 0.55 | 71 | 55.5 | 0.54 | 71 | 55.4 | 9 |
| 10 | 0.65 | 74 | 56.5 | 0.68 | 75 | 56.8 | 0.66 | 75 | 56.6 | 0.63 | 74 | 56.3 | 0.66 | 75 | 56.6 | 0.64 | 74 | 56.4 | 10 |
| 11 | 0.76 | 78 | 57.6 | 0.79 | 79 | 57.9 | 0.78 | 78 | 57.8 | 0.74 | 77 | 57.4 | 0.77 | 78 | 57.7 | 0.76 | 78 | 57.6 | 11 |

| | | | | | | | | | | | | | | | | | | |
|---|---|---|---|---|---|---|---|---|---|---|---|---|---|---|---|---|---|---|
| **12** | 0.88 | 81 | 58.8 | 0.91 | 82 | 59.1 | 0.89 | 81 | 58.9 | 0.86 | 81 | 58.6 | 0.89 | 81 | 58.9 | 0.87 | 81 | 58.7 | **12** |
| **13** | 0.98 | 84 | 59.8 | 01.01 | 84 | 60.1 | 0.99 | 84 | 59.9 | 0.96 | 83 | 59.6 | 0.99 | 84 | 59.9 | 0.97 | 83 | 59.7 | **13** |
| **14** | 01.05 | 85 | 60.5 | 01.07 | 86 | 60.7 | 01.06 | 86 | 60.6 | 01.03 | 85 | 60.3 | 01.05 | 85 | 60.5 | 01.04 | 85 | 60.4 | **14** |
| **15** | 01.09 | 86 | 60.9 | 1.12 | 87 | 61.2 | 1.10 | 86 | 61.0 | 01.07 | 86 | 60.7 | 1.10 | 86 | 61.0 | 01.08 | 86 | 60.8 | **15** |
| **16** | 1.11 | 87 | 61.1 | 1.14 | 87 | 61.4 | 1.13 | 87 | 61.3 | 01.09 | 86 | 60.9 | 1.12 | 87 | 61.2 | 1.11 | 87 | 61.1 | **16** |
| **17** | 1.14 | 87 | 61.4 | 1.16 | 88 | 61.6 | 1.15 | 87 | 61.5 | 1.12 | 87 | 61.2 | 1.14 | 87 | 61.4 | 1.13 | 87 | 61.3 | **17** |
| **18** | 1.18 | 88 | 61.8 | 1.21 | 89 | 62.1 | 1.19 | 88 | 61.9 | 1.16 | 88 | 61.6 | 1.19 | 88 | 61.9 | 1.17 | 88 | 61.7 | **18** |
| **19** | 1.24 | 89 | 62.4 | 1.27 | 90 | 62.7 | 1.26 | 90 | 62.6 | 1.22 | 89 | 62.2 | 1.25 | 89 | 62.5 | 1.24 | 89 | 62.4 | **19** |
| **20** | 1.33 | 91 | 63.3 | 1.36 | 91 | 63.6 | 1.35 | 91 | 63.5 | 1.31 | 90 | 63.1 | 1.34 | 91 | 63.4 | 1.33 | 91 | 63.3 | **20** |
| **21** | 1.44 | 93 | 64.4 | 1.47 | 93 | 64.7 | 1.45 | 93 | 64.5 | 1.42 | 92 | 64.2 | 1.45 | 93 | 64.5 | 1.43 | 92 | 64.3 | **21** |
| **22** | 1.54 | 94 | 65.4 | 1.57 | 94 | 65.7 | 1.55 | 94 | 65.5 | 1.52 | 94 | 65.2 | 1.55 | 94 | 65.5 | 1.53 | 94 | 65.3 | **22** |
| **23** | 1.62 | 95 | 66.2 | 1.65 | 95 | 66.5 | 1.64 | 95 | 66.4 | 1.61 | 95 | 66.1 | 1.63 | 95 | 66.3 | 1.62 | 95 | 66.2 | **23** |
| **24** | 1.69 | 95 | 66.9 | 1.72 | 96 | 67.2 | 1.71 | 96 | 67.1 | 1.67 | 95 | 66.7 | 1.70 | 96 | 67.0 | 1.69 | 95 | 66.9 | **24** |
| **25** | 1.74 | 96 | 67.4 | 1.77 | 96 | 67.7 | 1.76 | 96 | 67.6 | 1.72 | 96 | 67.2 | 1.75 | 96 | 67.5 | 1.74 | 96 | 67.4 | **25** |
| **26** | 1.79 | 96 | 67.9 | 1.82 | 97 | 68.2 | 1.80 | 96 | 68.0 | 1.77 | 96 | 67.7 | 1.80 | 96 | 68.0 | 1.78 | 96 | 67.8 | **26** |
| **27** | 1.84 | 97 | 68.4 | 1.87 | 97 | 68.7 | 1.85 | 97 | 68.5 | 1.82 | 97 | 68.2 | 1.85 | 97 | 68.5 | 1.83 | 97 | 68.3 | **27** |
| **28** | 1.90 | 97 | 69.0 | 1.93 | 97 | 69.3 | 1.91 | 97 | 69.1 | 1.88 | 97 | 68.8 | 1.91 | 97 | 69.1 | 1.89 | 97 | 68.9 | **28** |
| **29** | 1.97 | 98 | 69.7 | 2.00 | 98 | 70.0 | 1.99 | 98 | 69.9 | 1.95 | 97 | 69.5 | 1.98 | 98 | 69.8 | 1.97 | 98 | 69.7 | **29** |

| | | | | | | | | | | | | | | | | | | | |
|---|---|---|---|---|---|---|---|---|---|---|---|---|---|---|---|---|---|---|---|
| **30** | 02.05 | 98 | 70.5 | 02.08 | 98 | 70.8 | 02.07 | 98 | 70.7 | 02.03 | 98 | 70.3 | 02.06 | 98 | 70.6 | 02.05 | 98 | 70.5 | **30** |
| **31** | 2.12 | 98 | 71.2 | 2.15 | 98 | 71.5 | 2.14 | 98 | 71.4 | 2.10 | 98 | 71.0 | 2.13 | 98 | 71.3 | 2.12 | 98 | 71.2 | **31** |
| **32** | 2.18 | 99 | 71.8 | 2.21 | 99 | 72.1 | 2.20 | 99 | 72.0 | 2.16 | 98 | 71.6 | 2.19 | 99 | 71.9 | 2.18 | 99 | 71.8 | **32** |
| **33** | 2.22 | 99 | 72.2 | 2.25 | 99 | 72.5 | 2.23 | 99 | 72.3 | 2.20 | 99 | 72.0 | 2.23 | 99 | 72.3 | 2.21 | 99 | 72.1 | **33** |
| **34** | 2.24 | 99 | 72.4 | 2.27 | 99 | 72.7 | 2.26 | 99 | 72.6 | 2.22 | 99 | 72.2 | 2.25 | 99 | 72.5 | 2.24 | 99 | 72.4 | **34** |
| **35** | 2.25 | 99 | 72.5 | 2.28 | 99 | 72.8 | 2.27 | 99 | 72.7 | 2.23 | 99 | 72.3 | 2.26 | 99 | 72.6 | 2.25 | 99 | 72.5 | **35** |
| **36** | 2.26 | 99 | 72.6 | 2.29 | 99 | 72.9 | 2.28 | 99 | 72.8 | 2.25 | 99 | 72.5 | 2.27 | 99 | 72.7 | 2.26 | 99 | 72.6 | **36** |
| **37** | 2.29 | 99 | 72.9 | 2.32 | 99 | 73.2 | 2.31 | 99 | 73.1 | 2.27 | 99 | 72.7 | 2.30 | 99 | 73.0 | 2.29 | 99 | 72.9 | **37** |
| **38** | 2.34 | 99 | 73.4 | 2.37 | 99 | 73.7 | 2.35 | 99 | 73.5 | 2.32 | 99 | 73.2 | 2.35 | 99 | 73.5 | 2.33 | 99 | 73.3 | **38** |
| **39** | 2.42 | 99 | 74.2 | 2.45 | 99 | 74.5 | 2.43 | 99 | 74.3 | 2.40 | 99 | 74.0 | 2.43 | 99 | 74.3 | 2.41 | 99 | 74.1 | **39** |
| **40** | 2.53 | 99 | 75.3 | 2.56 | 99 | 75.6 | 2.54 | 99 | 75.4 | 2.51 | 99 | 75.1 | 2.54 | 99 | 75.4 | 2.52 | 99 | 75.2 | **40** |
| **41** | 2.66 | 100 | 76.6 | 2.69 | 100 | 76.9 | 2.67 | 100 | 76.7 | 2.64 | 100 | 76.4 | 2.67 | 100 | 76.7 | 2.65 | 100 | 76.5 | **41** |
| **42** | 2.81 | 100 | 78.1 | 2.84 | 100 | 78.4 | 2.82 | 100 | 78.2 | 2.79 | 100 | 77.9 | 2.82 | 100 | 78.2 | 2.80 | 100 | 78.0 | **42** |
| **43** | 2.97 | 100 | 79.7 | 3.00 | 100 | 80.0 | 2.99 | 100 | 79.9 | 2.95 | 100 | 79.5 | 2.98 | 100 | 79.8 | 2.97 | 100 | 79.7 | **43** |
| **44** | 3.15 | 100 | 81.5 | 3.18 | 100 | 81.8 | 3.16 | 100 | 81.6 | 3.13 | 100 | 81.3 | 3.16 | 100 | 81.6 | 3.14 | 100 | 81.4 | **44** |
| **45** | 3.33 | 100 | 83.3 | 3.35 | 100 | 83.5 | 3.34 | 100 | 83.4 | 3.31 | 100 | 83.1 | 3.34 | 100 | 83.4 | 3.32 | 100 | 83.2 | **45** |
| **DEPRESSION SCALE** | | | | | | | | | | | | | | | | | | | |
| | **Male** | | | | | | | | | **Female** | | | | | | | | | |
| | **6 to 9 years-old** | | | **10 to 13 years** | | | **14 to 18 years-old** | | | **6 to 9 years-old** | | | **10 to 13 years** | | | **14 to 18 years-old** | | | |

| SUM | Z | C | T | Z | C | T | Z | C | T | Z | C | T | Z | C | T | Z | C | T | SUM |
|---|---|---|---|---|---|---|---|---|---|---|---|---|---|---|---|---|---|---|---|
| **0** | -0.98 | 16 | 40.2 | -0.97 | 17 | 40.3 | -0.97 | 17 | 40.3 | -0.98 | 16 | 40.2 | -0.97 | 17 | 40.3 | -0.97 | 17 | 40.3 | **0** |
| **1** | -0.44 | 33 | 45.6 | -0.43 | 33 | 45.7 | -0.43 | 33 | 45.7 | -0.44 | 33 | 45.6 | -0.43 | 33 | 45.7 | -0.43 | 33 | 45.7 | **1** |
| **2** | -0.01 | 50 | 49.9 | 0.00 | 50 | 50.0 | 0.00 | 50 | 50.0 | -0.01 | 50 | 49.9 | 0.00 | 50 | 50.0 | 0.00 | 50 | 50.0 | **2** |
| **3** | 0.25 | 60 | 52.5 | 0.26 | 60 | 52.6 | 0.26 | 60 | 52.6 | 0.25 | 60 | 52.5 | 0.26 | 60 | 52.6 | 0.25 | 60 | 52.5 | **3** |
| **4** | 0.36 | 64 | 53.6 | 0.37 | 64 | 53.7 | 0.37 | 64 | 53.7 | 0.35 | 64 | 53.5 | 0.36 | 64 | 53.6 | 0.36 | 64 | 53.6 | **4** |
| **5** | 0.42 | 66 | 54.2 | 0.43 | 67 | 54.3 | 0.43 | 67 | 54.3 | 0.41 | 66 | 54.1 | 0.42 | 66 | 54.2 | 0.42 | 66 | 54.2 | **5** |
| **6** | 0.52 | 70 | 55.2 | 0.53 | 70 | 55.3 | 0.53 | 70 | 55.3 | 0.52 | 70 | 55.2 | 0.53 | 70 | 55.3 | 0.53 | 70 | 55.3 | **6** |
| **7** | 0.69 | 75 | 56.9 | 0.70 | 76 | 57.0 | 0.70 | 76 | 57.0 | 0.69 | 75 | 56.9 | 0.70 | 76 | 57.0 | 0.69 | 75 | 56.9 | **7** |
| **8** | 0.87 | 81 | 58.7 | 0.88 | 81 | 58.8 | 0.88 | 81 | 58.8 | 0.87 | 81 | 58.7 | 0.88 | 81 | 58.8 | 0.88 | 81 | 58.8 | **8** |
| **9** | 01.01 | 84 | 60.1 | 01.02 | 85 | 60.2 | 01.02 | 85 | 60.2 | 1.00 | 84 | 60.0 | 01.01 | 84 | 60.1 | 01.01 | 84 | 60.1 | **9** |
| **10** | 01.07 | 86 | 60.7 | 01.09 | 86 | 60.9 | 01.08 | 86 | 60.8 | 01.07 | 86 | 60.7 | 01.08 | 86 | 60.8 | 01.08 | 86 | 60.8 | **10** |
| **11** | 1.12 | 87 | 61.2 | 1.13 | 87 | 61.3 | 1.13 | 87 | 61.3 | 1.11 | 87 | 61.1 | 1.12 | 87 | 61.2 | 1.12 | 87 | 61.2 | **11** |
| **12** | 1.20 | 88 | 62.0 | 1.21 | 89 | 62.1 | 1.21 | 89 | 62.1 | 1.20 | 88 | 62.0 | 1.21 | 89 | 62.1 | 1.21 | 89 | 62.1 | **12** |
| **13** | 1.35 | 91 | 63.5 | 1.36 | 91 | 63.6 | 1.36 | 91 | 63.6 | 1.34 | 91 | 63.4 | 1.35 | 91 | 63.5 | 1.35 | 91 | 63.5 | **13** |
| **14** | 1.52 | 94 | 65.2 | 1.53 | 94 | 65.3 | 1.53 | 94 | 65.3 | 1.52 | 94 | 65.2 | 1.53 | 94 | 65.3 | 1.52 | 94 | 65.2 | **14** |
| **15** | 1.66 | 95 | 66.6 | 1.67 | 95 | 66.7 | 1.67 | 95 | 66.7 | 1.65 | 95 | 66.5 | 1.66 | 95 | 66.6 | 1.66 | 95 | 66.6 | **15** |
| **16** | 1.72 | 96 | 67.2 | 1.73 | 96 | 67.3 | 1.73 | 96 | 67.3 | 1.72 | 96 | 67.2 | 1.73 | 96 | 67.3 | 1.73 | 96 | 67.3 | **16** |
| **17** | 1.74 | 96 | 67.4 | 1.75 | 96 | 67.5 | 1.75 | 96 | 67.5 | 1.74 | 96 | 67.4 | 1.75 | 96 | 67.5 | 1.75 | 96 | 67.5 | **17** |

| | | | | | | | | | | | | | | | | | | | |
|---|---|---|---|---|---|---|---|---|---|---|---|---|---|---|---|---|---|---|---|
| **18** | 1.78 | 96 | 67.8 | 1.79 | 96 | 67.9 | 1.79 | 96 | 67.9 | 1.77 | 96 | 67.7 | 1.78 | 96 | 67.8 | 1.78 | 96 | 67.8 | **18** |
| **19** | 1.87 | 97 | 68.7 | 1.88 | 97 | 68.8 | 1.88 | 97 | 68.8 | 1.87 | 97 | 68.7 | 1.88 | 97 | 68.8 | 1.88 | 97 | 68.8 | **19** |
| **20** | 02.01 | 98 | 70.1 | 02.02 | 98 | 70.2 | 02.02 | 98 | 70.2 | 2.00 | 98 | 70.0 | 02.01 | 98 | 70.1 | 02.01 | 98 | 70.1 | **20** |
| **21** | 2.15 | 98 | 71.5 | 2.16 | 98 | 71.6 | 2.16 | 98 | 71.6 | 2.15 | 98 | 71.5 | 2.16 | 98 | 71.6 | 2.16 | 98 | 71.6 | **21** |
| **22** | 2.26 | 99 | 72.6 | 2.27 | 99 | 72.7 | 2.27 | 99 | 72.7 | 2.26 | 99 | 72.6 | 2.27 | 99 | 72.7 | 2.27 | 99 | 72.7 | **22** |
| **23** | 2.32 | 99 | 73.2 | 2.33 | 99 | 73.3 | 2.33 | 99 | 73.3 | 2.31 | 99 | 73.1 | 2.32 | 99 | 73.2 | 2.32 | 99 | 73.2 | **23** |
| **24** | 2.36 | 99 | 73.6 | 2.37 | 99 | 73.7 | 2.36 | 99 | 73.6 | 2.35 | 99 | 73.5 | 2.36 | 99 | 73.6 | 2.36 | 99 | 73.6 | **24** |
| **25** | 2.41 | 99 | 74.1 | 2.42 | 99 | 74.2 | 2.42 | 99 | 74.2 | 2.41 | 99 | 74.1 | 2.42 | 99 | 74.2 | 2.42 | 99 | 74.2 | **25** |
| **26** | 2.52 | 99 | 75.2 | 2.53 | 99 | 75.3 | 2.53 | 99 | 75.3 | 2.52 | 99 | 75.2 | 2.53 | 99 | 75.3 | 2.53 | 99 | 75.3 | **26** |
| **27** | 2.68 | 100 | 76.8 | 2.69 | 100 | 76.9 | 2.69 | 100 | 76.9 | 2.68 | 100 | 76.8 | 2.69 | 100 | 76.9 | 2.69 | 100 | 76.9 | **27** |
| **28** | 2.88 | 100 | 78.8 | 2.89 | 100 | 78.9 | 2.89 | 100 | 78.9 | 2.87 | 100 | 78.7 | 2.88 | 100 | 78.8 | 2.88 | 100 | 78.8 | **28** |
| **29** | 03.09 | 100 | 80.9 | 3.11 | 100 | 81.1 | 3.10 | 100 | 81.0 | 03.09 | 100 | 80.9 | 3.10 | 100 | 81.0 | 3.10 | 100 | 81.0 | **29** |
| **30** | 3.33 | 100 | 83.3 | 3.34 | 100 | 83.4 | 3.33 | 100 | 83.3 | 3.32 | 100 | 83.2 | 3.33 | 100 | 83.3 | 3.33 | 100 | 83.3 | **30** |

**Abbreviations**: C (Centile); T (T-score); Z (Z-score). **Notes:** Colored distribution bands: Green: minimal symptoms (T-score < 55). Yellow: mild symptoms (T-score ≥ 55 and < 60); Orange: moderate symptoms (T-score ≥ 60 and < 70); Red: severe symptoms (T-score ≥ 70).

**Supplementary Table 10 -** Swanson, Nolan and Pelham Scale (SNAP-IV), Caregiver-report: normative references in Greece

| SNAP-IV - Caregiver-report | | | | | | | | | | | | | | | | | | | |
|---|---|---|---|---|---|---|---|---|---|---|---|---|---|---|---|---|---|---|---|
| INATTENTION SCALE | | | | | | | | | | | | | | | | | | | |
| | Male | | | | | | | | | Female | | | | | | | | | |
| | 6 to 9 years-old | | | 10 to 13 years | | | 14 to 18 years-old | | | 6 to 9 years-old | | | 10 to 13 years | | | 14 to 18 years-old | | | |
| SUM | Z | C | T | Z | C | T | Z | C | T | Z | C | T | Z | C | T | Z | C | T | SUM |
| 0 | -1.42 | 8 | 35.8 | -1.41 | 8 | 35.9 | -1.43 | 8 | 35.7 | -1.44 | 7 | 35.6 | -1.43 | 8 | 35.7 | -1.45 | 7 | 35.5 | 0 |
| 1 | -1.00 | 16 | 40.0 | -0.98 | 16 | 40.2 | -1.00 | 16 | 40.0 | -1.01 | 16 | 39.9 | -1.00 | 16 | 40.0 | -1.02 | 15 | 39.8 | 1 |
| 2 | -0.62 | 27 | 43.8 | -0.61 | 27 | 43.9 | -0.63 | 26 | 43.7 | -0.64 | 26 | 43.6 | -0.63 | 26 | 43.7 | -0.65 | 26 | 43.5 | 2 |
| 3 | -0.35 | 36 | 46.5 | -0.34 | 37 | 46.6 | -0.36 | 36 | 46.4 | -0.36 | 36 | 46.4 | -0.35 | 36 | 46.5 | -0.37 | 36 | 46.3 | 3 |
| 4 | -0.17 | 43 | 48.3 | -0.16 | 44 | 48.4 | -0.18 | 43 | 48.2 | -0.18 | 43 | 48.2 | -0.17 | 43 | 48.3 | -0.19 | 42 | 48.1 | 4 |
| 5 | -0.05 | 48 | 49.5 | -0.03 | 49 | 49.7 | -0.05 | 48 | 49.5 | -0.06 | 48 | 49.4 | -0.05 | 48 | 49.5 | -0.07 | 47 | 49.3 | 5 |
| 6 | 0.07 | 53 | 50.7 | 0.08 | 53 | 50.8 | 0.06 | 52 | 50.6 | 0.05 | 52 | 50.5 | 0.07 | 53 | 50.7 | 0.04 | 52 | 50.4 | 6 |
| 7 | 0.20 | 58 | 52.0 | 0.21 | 58 | 52.1 | 0.19 | 58 | 51.9 | 0.19 | 58 | 51.9 | 0.20 | 58 | 52.0 | 0.18 | 57 | 51.8 | 7 |
| 8 | 0.35 | 64 | 53.5 | 0.36 | 64 | 53.6 | 0.34 | 63 | 53.4 | 0.33 | 63 | 53.3 | 0.34 | 63 | 53.4 | 0.32 | 63 | 53.2 | 8 |
| 9 | 0.49 | 69 | 54.9 | 0.50 | 69 | 55.0 | 0.48 | 68 | 54.8 | 0.47 | 68 | 54.7 | 0.49 | 69 | 54.9 | 0.46 | 68 | 54.6 | 9 |
| 10 | 0.62 | 73 | 56.2 | 0.63 | 74 | 56.3 | 0.61 | 73 | 56.1 | 0.60 | 73 | 56.0 | 0.62 | 73 | 56.2 | 0.60 | 73 | 56.0 | 10 |
| 11 | 0.74 | 77 | 57.4 | 0.76 | 78 | 57.6 | 0.74 | 77 | 57.4 | 0.73 | 77 | 57.3 | 0.74 | 77 | 57.4 | 0.72 | 76 | 57.2 | 11 |

| | | | | | | | | | | | | | | | | | | | |
|---|---|---|---|---|---|---|---|---|---|---|---|---|---|---|---|---|---|---|---|
| **12** | 0.88 | 81 | 58.8 | 0.89 | 81 | 58.9 | 0.87 | 81 | 58.7 | 0.86 | 81 | 58.6 | 0.87 | 81 | 58.7 | 0.85 | 80 | 58.5 | **12** |
| **13** | 1.03 | 85 | 60.3 | 01.04 | 85 | 60.4 | 1.02 | 85 | 60.2 | 1.01 | 84 | 60.1 | 1.02 | 85 | 60.2 | 1.00 | 84 | 60.0 | **13** |
| **14** | 1.18 | 88 | 61.8 | 1.19 | 88 | 61.9 | 1.17 | 88 | 61.7 | 1.16 | 88 | 61.6 | 1.17 | 88 | 61.7 | 1.15 | 87 | 61.5 | **14** |
| **15** | 1.31 | 90 | 63.1 | 1.32 | 91 | 63.2 | 1.30 | 90 | 63.0 | 1.29 | 90 | 62.9 | 1.30 | 90 | 63.0 | 1.28 | 90 | 62.8 | **15** |
| **16** | 1.41 | 92 | 64.1 | 1.42 | 92 | 64.2 | 1.40 | 92 | 64.0 | 1.40 | 92 | 64.0 | 1.41 | 92 | 64.1 | 1.39 | 92 | 63.9 | **16** |
| **17** | 1.50 | 93 | 65.0 | 1.51 | 93 | 65.1 | 1.49 | 93 | 64.9 | 1.48 | 93 | 64.8 | 1.50 | 93 | 65.0 | 1.48 | 93 | 64.8 | **17** |
| **18** | 1.60 | 95 | 66.0 | 1.61 | 95 | 66.1 | 1.59 | 94 | 65.9 | 1.58 | 94 | 65.8 | 1.59 | 94 | 65.9 | 1.57 | 94 | 65.7 | **18** |
| **19** | 1.72 | 96 | 67.2 | 1.73 | 96 | 67.3 | 1.71 | 96 | 67.1 | 1.71 | 96 | 67.1 | 1.72 | 96 | 67.2 | 1.70 | 96 | 67.0 | **19** |
| **20** | 1.87 | 97 | 68.7 | 1.88 | 97 | 68.8 | 1.86 | 97 | 68.6 | 1.86 | 97 | 68.6 | 1.87 | 97 | 68.7 | 1.85 | 97 | 68.5 | **20** |
| **21** | 02.04 | 98 | 70.4 | 02.05 | 98 | 70.5 | 02.03 | 98 | 70.3 | 02.02 | 98 | 70.2 | 02.03 | 98 | 70.3 | 02.01 | 98 | 70.1 | **21** |
| **22** | 2.19 | 99 | 71.9 | 2.20 | 99 | 72.0 | 2.18 | 99 | 71.8 | 2.18 | 99 | 71.8 | 2.19 | 99 | 71.9 | 2.17 | 98 | 71.7 | **22** |
| **23** | 2.34 | 99 | 73.4 | 2.35 | 99 | 73.5 | 2.33 | 99 | 73.3 | 2.33 | 99 | 73.3 | 2.34 | 99 | 73.4 | 2.32 | 99 | 73.2 | **23** |
| **24** | 2.50 | 99 | 75.0 | 2.51 | 99 | 75.1 | 2.49 | 99 | 74.9 | 2.49 | 99 | 74.9 | 2.50 | 99 | 75.0 | 2.48 | 99 | 74.8 | **24** |
| **25** | 2.69 | 100 | 76.9 | 2.70 | 100 | 77.0 | 2.68 | 100 | 76.8 | 2.67 | 100 | 76.7 | 2.69 | 100 | 76.9 | 2.66 | 100 | 76.6 | **25** |
| **26** | 2.91 | 100 | 79.1 | 2.92 | 100 | 79.2 | 2.90 | 100 | 79.0 | 2.89 | 100 | 78.9 | 2.90 | 100 | 79.0 | 2.88 | 100 | 78.8 | **26** |
| **27** | 3.14 | 100 | 81.4 | 3.15 | 100 | 81.5 | 3.13 | 100 | 81.3 | 3.12 | 100 | 81.2 | 3.14 | 100 | 81.4 | 3.11 | 100 | 81.1 | **27** |
| **HYPERACTIVITY SCALE** | | | | | | | | | | | | | | | | | | | |
| | **Male** | | | | | | | | | **Female** | | | | | | | | | |
| | **6 to 9 years-old** | | | **10 to 13 years** | | | **14 to 18 years-old** | | | **6 to 9 years-old** | | | **10 to 13 years** | | | **14 to 18 years-old** | | | |

| SUM | Z | C | T | Z | C | T | Z | C | T | Z | C | T | Z | C | T | Z | C | T | SUM |
|---|---|---|---|---|---|---|---|---|---|---|---|---|---|---|---|---|---|---|---|
| 0 | -0.69 | 25 | 43.1 | -0.70 | 24 | 43.0 | -0.69 | 25 | 43.1 | -0.70 | 24 | 43.0 | -0.71 | 24 | 42.9 | -0.69 | 25 | 43.1 | 0 |
| 1 | 0.08 | 53 | 50.8 | 0.07 | 53 | 50.7 | 0.08 | 53 | 50.8 | 0.07 | 53 | 50.7 | 0.06 | 52 | 50.6 | 0.08 | 53 | 50.8 | 1 |
| 2 | 0.48 | 68 | 54.8 | 0.47 | 68 | 54.7 | 0.49 | 69 | 54.9 | 0.48 | 68 | 54.8 | 0.47 | 68 | 54.7 | 0.48 | 68 | 54.8 | 2 |
| 3 | 0.64 | 74 | 56.4 | 0.63 | 74 | 56.3 | 0.64 | 74 | 56.4 | 0.63 | 74 | 56.3 | 0.62 | 73 | 56.2 | 0.64 | 74 | 56.4 | 3 |
| 4 | 0.84 | 80 | 58.4 | 0.83 | 80 | 58.3 | 0.85 | 80 | 58.5 | 0.83 | 80 | 58.3 | 0.83 | 80 | 58.3 | 0.84 | 80 | 58.4 | 4 |
| 5 | 1.09 | 86 | 60.9 | 1.08 | 86 | 60.8 | 1.09 | 86 | 60.9 | 1.08 | 86 | 60.8 | 1.07 | 86 | 60.7 | 1.09 | 86 | 60.9 | 5 |
| 6 | 1.26 | 90 | 62.6 | 1.25 | 89 | 62.5 | 1.26 | 90 | 62.6 | 1.25 | 89 | 62.5 | 1.24 | 89 | 62.4 | 1.26 | 90 | 62.6 | 6 |
| 7 | 1.40 | 92 | 64.0 | 1.39 | 92 | 63.9 | 1.41 | 92 | 64.1 | 1.39 | 92 | 63.9 | 1.39 | 92 | 63.9 | 1.40 | 92 | 64.0 | 7 |
| 8 | 1.58 | 94 | 65.8 | 1.57 | 94 | 65.7 | 1.58 | 94 | 65.8 | 1.57 | 94 | 65.7 | 1.56 | 94 | 65.6 | 1.58 | 94 | 65.8 | 8 |
| 9 | 1.72 | 96 | 67.2 | 1.71 | 96 | 67.1 | 1.73 | 96 | 67.3 | 1.71 | 96 | 67.1 | 1.70 | 96 | 67.0 | 1.72 | 96 | 67.2 | 9 |
| 10 | 1.83 | 97 | 68.3 | 1.82 | 97 | 68.2 | 1.84 | 97 | 68.4 | 1.83 | 97 | 68.3 | 1.82 | 97 | 68.2 | 1.83 | 97 | 68.3 | 10 |
| 11 | 02.01 | 98 | 70.1 | 2.00 | 98 | 70.0 | 02.02 | 98 | 70.2 | 02.01 | 98 | 70.1 | 2.00 | 98 | 70.0 | 02.01 | 98 | 70.1 | 11 |
| 12 | 2.21 | 99 | 72.1 | 2.21 | 99 | 72.1 | 2.22 | 99 | 72.2 | 2.21 | 99 | 72.1 | 2.20 | 99 | 72.0 | 2.21 | 99 | 72.1 | 12 |
| 13 | 2.37 | 99 | 73.7 | 2.36 | 99 | 73.6 | 2.38 | 99 | 73.8 | 2.36 | 99 | 73.6 | 2.35 | 99 | 73.5 | 2.37 | 99 | 73.7 | 13 |
| 14 | 2.59 | 100 | 75.9 | 2.59 | 100 | 75.9 | 2.60 | 100 | 76.0 | 2.59 | 100 | 75.9 | 2.58 | 100 | 75.8 | 2.59 | 100 | 75.9 | 14 |
| 15 | 2.97 | 100 | 79.7 | 2.96 | 100 | 79.6 | 2.98 | 100 | 79.8 | 2.97 | 100 | 79.7 | 2.96 | 100 | 79.6 | 2.97 | 100 | 79.7 | 15 |

### IMPULSIVITY SCALE

| | Male | | | | | | | | | Female | | | | | | | | | |
|---|---|---|---|---|---|---|---|---|---|---|---|---|---|---|---|---|---|---|---|
| | 6 to 9 years-old | | | 10 to 13 years | | | 14 to 18 years-old | | | 6 to 9 years-old | | | 10 to 13 years | | | 14 to 18 years-old | | | |
| SUM | Z | C | T | Z | C | T | Z | C | T | Z | C | T | Z | C | T | Z | C | T | SUM |
| 0 | -1.04 | 15 | 39.6 | -1.02 | 15 | 39.8 | -1.03 | 15 | 39.7 | -1.05 | 15 | 39.5 | -1.03 | 15 | 39.7 | -1.04 | 15 | 39.6 | 0 |

| | | | | | | | | | | | | | | | | | | | |
|---|---|---|---|---|---|---|---|---|---|---|---|---|---|---|---|---|---|---|---|
| 1 | -0.38 | 35 | 46.2 | -0.36 | 36 | 46.4 | -0.37 | 36 | 46.3 | -0.39 | 35 | 46.1 | -0.36 | 36 | 46.4 | -0.38 | 35 | 46.2 | 1 |
| 2 | 0.02 | 51 | 50.2 | 0.04 | 52 | 50.4 | 0.03 | 51 | 50.3 | 0.01 | 50 | 50.1 | 0.04 | 52 | 50.4 | 0.02 | 51 | 50.2 | 2 |
| 3 | 0.28 | 61 | 52.8 | 0.30 | 62 | 53.0 | 0.28 | 61 | 52.8 | 0.27 | 61 | 52.7 | 0.29 | 61 | 52.9 | 0.28 | 61 | 52.8 | 3 |
| 4 | 0.52 | 70 | 55.2 | 0.54 | 71 | 55.4 | 0.52 | 70 | 55.2 | 0.51 | 69 | 55.1 | 0.53 | 70 | 55.3 | 0.52 | 70 | 55.2 | 4 |
| 5 | 0.75 | 77 | 57.5 | 0.77 | 78 | 57.7 | 0.76 | 78 | 57.6 | 0.74 | 77 | 57.4 | 0.76 | 78 | 57.6 | 0.75 | 77 | 57.5 | 5 |
| 6 | 01.01 | 84 | 60.1 | 1.03 | 85 | 60.3 | 01.02 | 85 | 60.2 | 1.01 | 84 | 60.1 | 1.03 | 85 | 60.3 | 1.01 | 84 | 60.1 | 6 |
| 7 | 1.26 | 90 | 62.6 | 1.28 | 90 | 62.8 | 1.27 | 90 | 62.7 | 1.25 | 89 | 62.5 | 1.28 | 90 | 62.8 | 1.26 | 90 | 62.6 | 7 |
| 8 | 1.48 | 93 | 64.8 | 1.50 | 93 | 65.0 | 1.48 | 93 | 64.8 | 1.47 | 93 | 64.7 | 1.49 | 93 | 64.9 | 1.48 | 93 | 64.8 | 8 |
| 9 | 1.74 | 96 | 67.4 | 1.76 | 96 | 67.6 | 1.74 | 96 | 67.4 | 1.73 | 96 | 67.3 | 1.75 | 96 | 67.5 | 1.74 | 96 | 67.4 | 9 |
| 10 | 02.02 | 98 | 70.2 | 02.04 | 98 | 70.4 | 02.02 | 98 | 70.2 | 02.01 | 98 | 70.1 | 02.03 | 98 | 70.3 | 02.02 | 98 | 70.2 | 10 |
| 11 | 2.31 | 99 | 73.1 | 2.33 | 99 | 73.3 | 2.32 | 99 | 73.2 | 2.30 | 99 | 73.0 | 2.32 | 99 | 73.2 | 2.31 | 99 | 73.1 | 11 |
| 12 | 2.69 | 100 | 76.9 | 2.71 | 100 | 77.1 | 2.69 | 100 | 76.9 | 2.68 | 100 | 76.8 | 2.70 | 100 | 77.0 | 2.69 | 100 | 76.9 | 12 |

### OPPOSITIONALITY SCALE

| | Male | | | | | | | | | Female | | | | | | | | | |
|---|---|---|---|---|---|---|---|---|---|---|---|---|---|---|---|---|---|---|---|
| | 6 to 9 years-old | | | 10 to 13 years | | | 14 to 18 years-old | | | 6 to 9 years-old | | | 10 to 13 years | | | 14 to 18 years-old | | | |
| SUM | Z | C | T | Z | C | T | Z | C | T | Z | C | T | Z | C | T | Z | C | T | SUM |
| 0 | -1.01 | 16 | 39.9 | -1.01 | 16 | 39.9 | -1.00 | 16 | 40.0 | -1.00 | 16 | 40.0 | -1.00 | 16 | 40.0 | -1.00 | 16 | 40.0 | 0 |
| 1 | -0.43 | 33 | 45.7 | -0.43 | 33 | 45.7 | -0.42 | 34 | 45.8 | -0.42 | 34 | 45.8 | -0.42 | 34 | 45.8 | -0.42 | 34 | 45.8 | 1 |
| 2 | -0.01 | 50 | 49.9 | -0.01 | 50 | 49.9 | 0.00 | 50 | 50.0 | 0.00 | 50 | 50.0 | 0.00 | 50 | 50.0 | 0.00 | 50 | 50.0 | 2 |
| 3 | 0.20 | 58 | 52.0 | 0.20 | 58 | 52.0 | 0.21 | 58 | 52.1 | 0.21 | 58 | 52.1 | 0.21 | 58 | 52.1 | 0.21 | 58 | 52.1 | 3 |
| 4 | 0.29 | 61 | 52.9 | 0.29 | 61 | 52.9 | 0.30 | 62 | 53.0 | 0.30 | 62 | 53.0 | 0.30 | 62 | 53.0 | 0.30 | 62 | 53.0 | 4 |

| | | | | | | | | | | | | | | | | | | | |
|---|---|---|---|---|---|---|---|---|---|---|---|---|---|---|---|---|---|---|---|
| **5** | 0.41 | 66 | 54.1 | 0.41 | 66 | 54.1 | 0.41 | 66 | 54.1 | 0.41 | 66 | 54.1 | 0.41 | 66 | 54.1 | 0.42 | 66 | 54.2 | **5** |
| **6** | 0.58 | 72 | 55.8 | 0.58 | 72 | 55.8 | 0.59 | 72 | 55.9 | 0.59 | 72 | 55.9 | 0.59 | 72 | 55.9 | 0.59 | 72 | 55.9 | **6** |
| **7** | 0.76 | 78 | 57.6 | 0.76 | 78 | 57.6 | 0.77 | 78 | 57.7 | 0.77 | 78 | 57.7 | 0.77 | 78 | 57.7 | 0.77 | 78 | 57.7 | **7** |
| **8** | 0.88 | 81 | 58.8 | 0.88 | 81 | 58.8 | 0.89 | 81 | 58.9 | 0.89 | 81 | 58.9 | 0.89 | 81 | 58.9 | 0.89 | 81 | 58.9 | **8** |
| **9** | 0.96 | 83 | 59.6 | 0.96 | 83 | 59.6 | 0.96 | 83 | 59.6 | 0.96 | 83 | 59.6 | 0.96 | 83 | 59.6 | 0.97 | 83 | 59.7 | **9** |
| **10** | 01.07 | 86 | 60.7 | 01.07 | 86 | 60.7 | 01.07 | 86 | 60.7 | 1.07 | 86 | 60.7 | 01.07 | 86 | 60.7 | 01.08 | 86 | 60.8 | **10** |
| **11** | 1.23 | 89 | 62.3 | 1.23 | 89 | 62.3 | 1.24 | 89 | 62.4 | 1.24 | 89 | 62.4 | 1.24 | 89 | 62.4 | 1.24 | 89 | 62.4 | **11** |
| **12** | 1.40 | 92 | 64.0 | 1.40 | 92 | 64.0 | 1.40 | 92 | 64.0 | 1.40 | 92 | 64.0 | 1.40 | 92 | 64.0 | 1.41 | 92 | 64.1 | **12** |
| **13** | 1.50 | 93 | 65.0 | 1.50 | 93 | 65.0 | 1.51 | 93 | 65.1 | 1.51 | 93 | 65.1 | 1.51 | 93 | 65.1 | 1.51 | 93 | 65.1 | **13** |
| **14** | 1.57 | 94 | 65.7 | 1.57 | 94 | 65.7 | 1.57 | 94 | 65.7 | 1.57 | 94 | 65.7 | 1.57 | 94 | 65.7 | 1.58 | 94 | 65.8 | **14** |
| **15** | 1.66 | 95 | 66.6 | 1.66 | 95 | 66.6 | 1.66 | 95 | 66.6 | 1.66 | 95 | 66.6 | 1.66 | 95 | 66.6 | 1.67 | 95 | 66.7 | **15** |
| **16** | 1.81 | 96 | 68.1 | 1.81 | 96 | 68.1 | 1.82 | 97 | 68.2 | 1.81 | 96 | 68.1 | 1.82 | 97 | 68.2 | 1.82 | 97 | 68.2 | **16** |
| **17** | 1.98 | 98 | 69.8 | 1.98 | 98 | 69.8 | 1.99 | 98 | 69.9 | 1.99 | 98 | 69.9 | 1.99 | 98 | 69.9 | 1.99 | 98 | 69.9 | **17** |
| **18** | 2.10 | 98 | 71.0 | 2.10 | 98 | 71.0 | 2.11 | 98 | 71.1 | 2.11 | 98 | 71.1 | 2.11 | 98 | 71.1 | 2.11 | 98 | 71.1 | **18** |
| **19** | 2.16 | 98 | 71.6 | 2.16 | 98 | 71.6 | 2.16 | 98 | 71.6 | 2.16 | 98 | 71.6 | 2.17 | 98 | 71.7 | 2.17 | 98 | 71.7 | **19** |
| **20** | 2.24 | 99 | 72.4 | 2.24 | 99 | 72.4 | 2.25 | 99 | 72.5 | 2.25 | 99 | 72.5 | 2.25 | 99 | 72.5 | 2.25 | 99 | 72.5 | **20** |
| **21** | 2.42 | 99 | 74.2 | 2.42 | 99 | 74.2 | 2.43 | 99 | 74.3 | 2.43 | 99 | 74.3 | 2.43 | 99 | 74.3 | 2.43 | 99 | 74.3 | **21** |
| **22** | 2.69 | 100 | 76.9 | 2.69 | 100 | 76.9 | 2.69 | 100 | 76.9 | 2.69 | 100 | 76.9 | 2.69 | 100 | 76.9 | 2.70 | 100 | 77.0 | **22** |

| | | | | | | | | | | | | | | | | | | | |
|---|---|---|---|---|---|---|---|---|---|---|---|---|---|---|---|---|---|---|---|
| **23** | 2.97 | 100 | 79.7 | 2.97 | 100 | 79.7 | 2.98 | 100 | 79.8 | 2.98 | 100 | 79.8 | 2.98 | 100 | 79.8 | 2.98 | 100 | 79.8 | **23** |
| **24** | 3.26 | 100 | 82.6 | 3.26 | 100 | 82.6 | 3.26 | 100 | 82.6 | 3.26 | 100 | 82.6 | 3.26 | 100 | 82.6 | 3.27 | 100 | 82.7 | **24** |

**Abbreviations**: C (Centile); T (T-score); Z (Z-score). **Notes:** Colored distribution bands: Green: minimal symptoms (T-score < 55). Yellow: mild symptoms (T-score ≥ 55 and < 60); Orange: moderate symptoms (T-score ≥ 60 and < 70); Red: severe symptoms (T-score ≥ 70).

**Supplementary Figure 1.1.1 -** Child Autism Spectrum Test (CAST), Caregiver-report (inflexible/repetitive behaviors): test information and expected scores

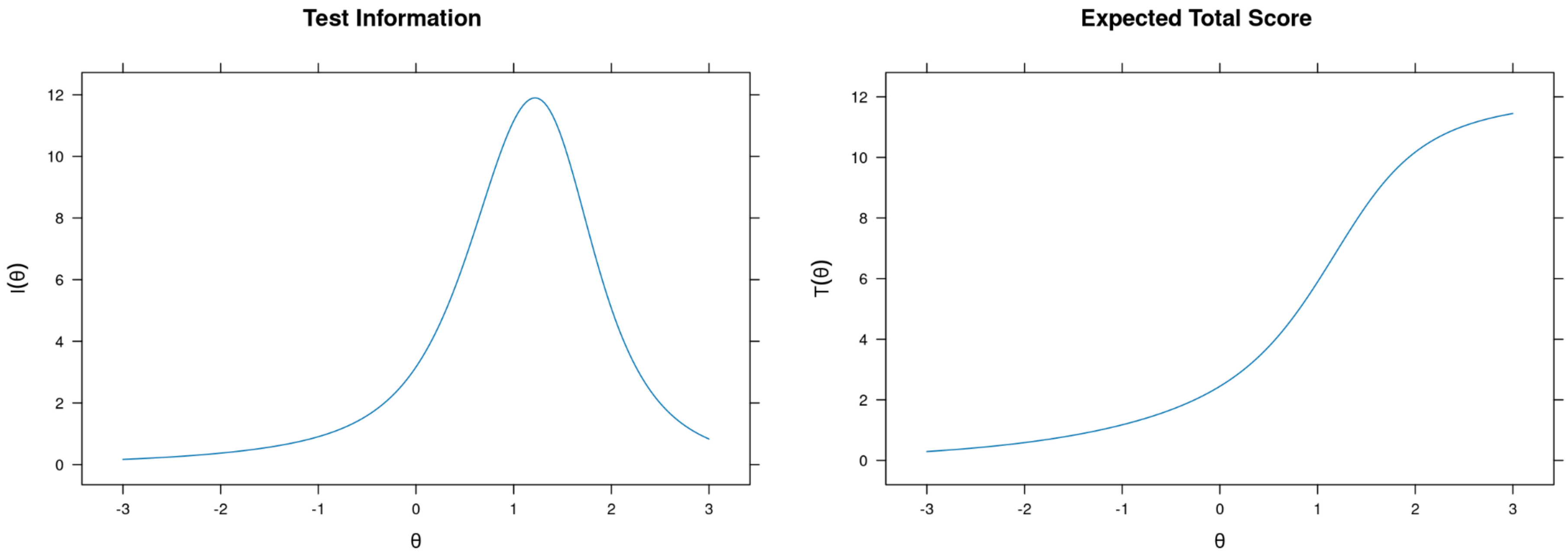

**Supplementary Figure 1.1.2 -** Child Autism Spectrum Test (CAST), Caregiver-report (inflexible/repetitive behaviors): item probability functions

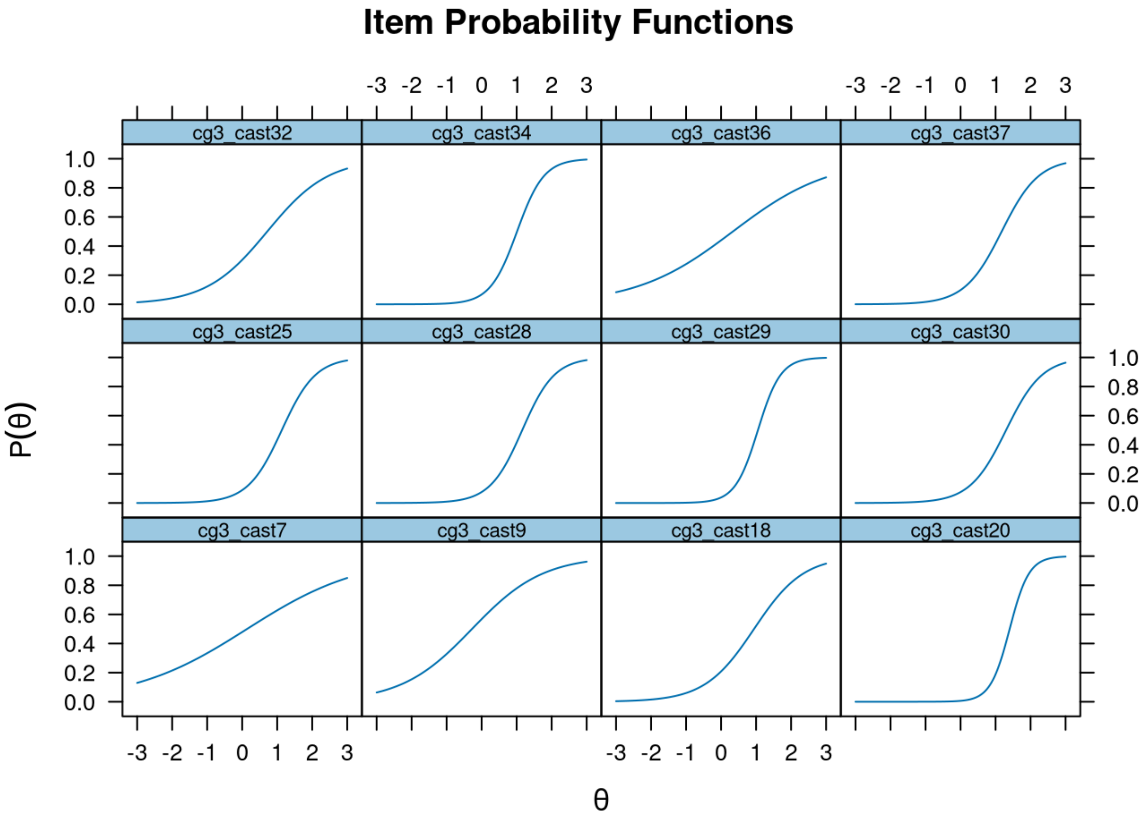

**Supplementary Figure 1.1.3 -** Child Autism Spectrum Test (CAST), Caregiver-report (inflexible/repetitive behaviors): item infit and outfit statistics

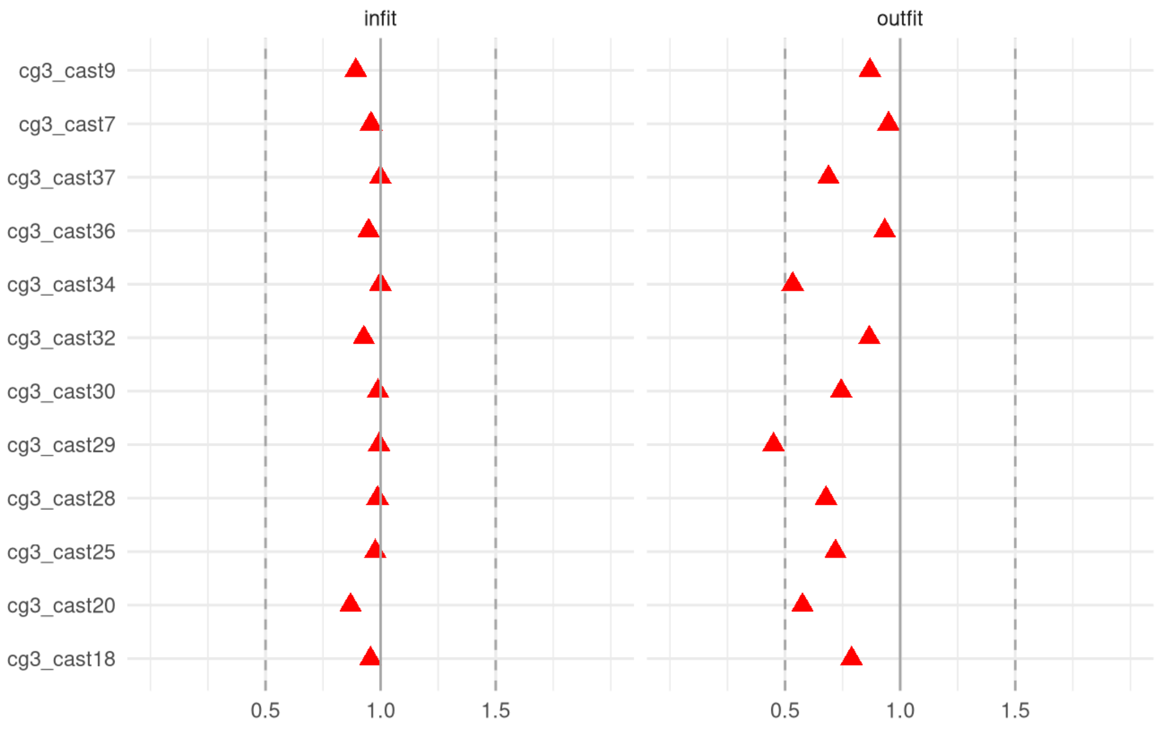

**Supplementary Figure 1.1.4 -** Child Autism Spectrum Test (CAST), Caregiver-report (inflexible/repetitive behaviors): person infit and outfit statistics

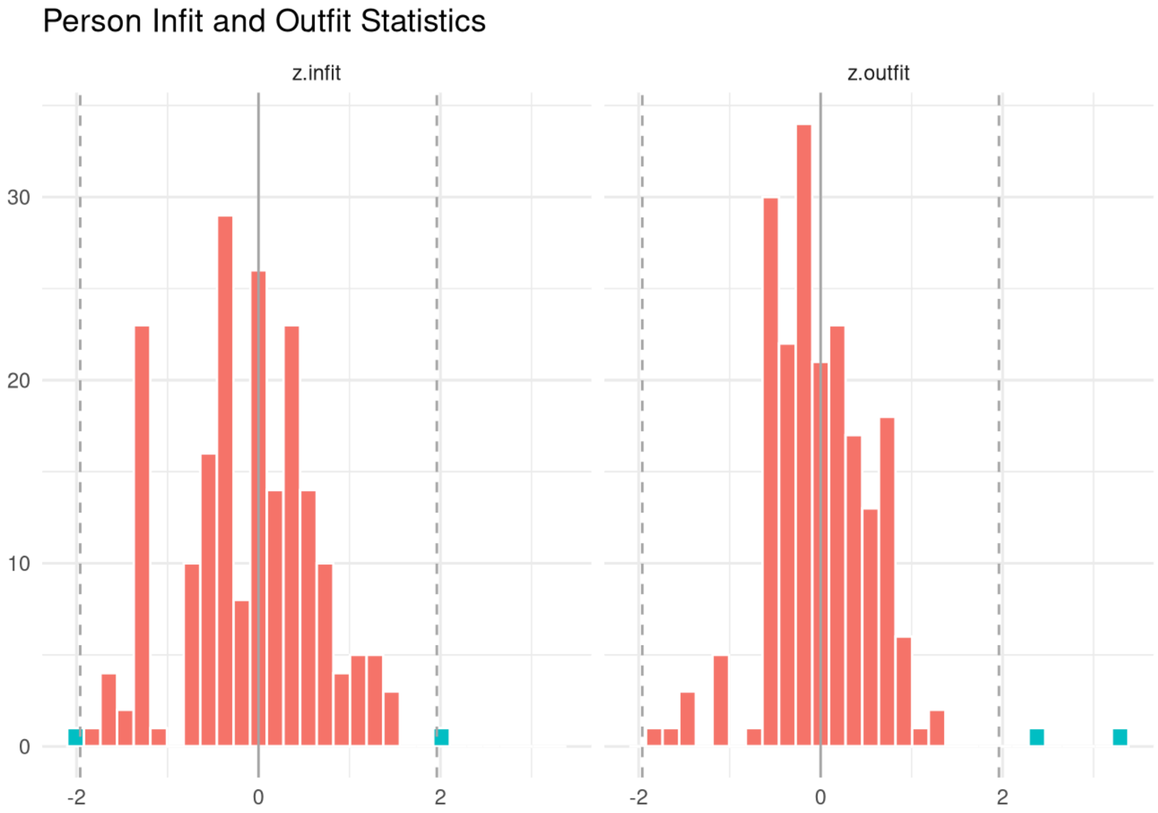

**Supplementary Figure 1.1.5 -** Child Autism Spectrum Test (CAST), Caregiver-report (inflexible/repetitive behaviors): item response functions

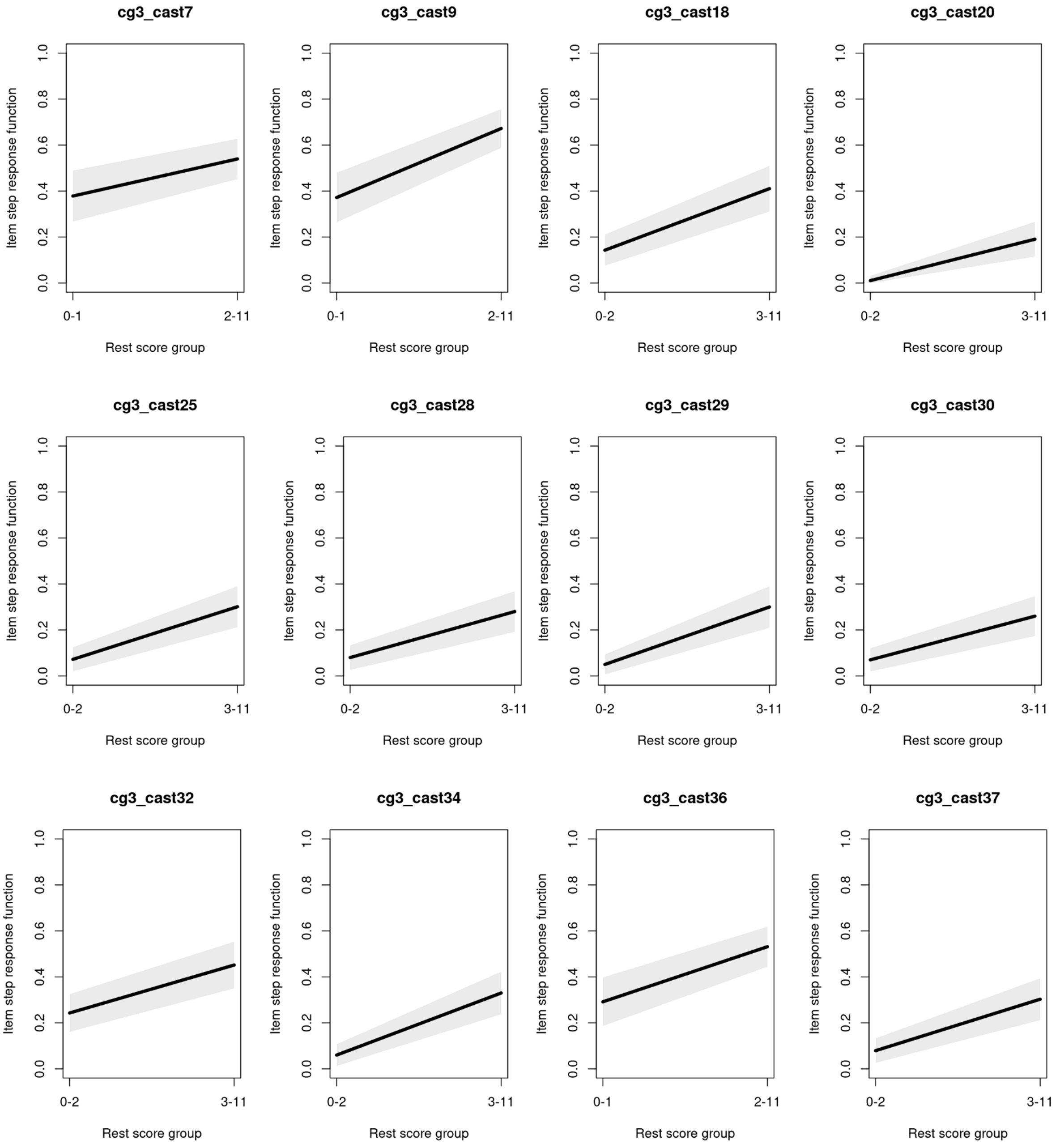

**Supplementary Figure 1.2.1 -** Child Autism Spectrum Test (CAST), Caregiver-report (Sociability/communication): test information and expected scores

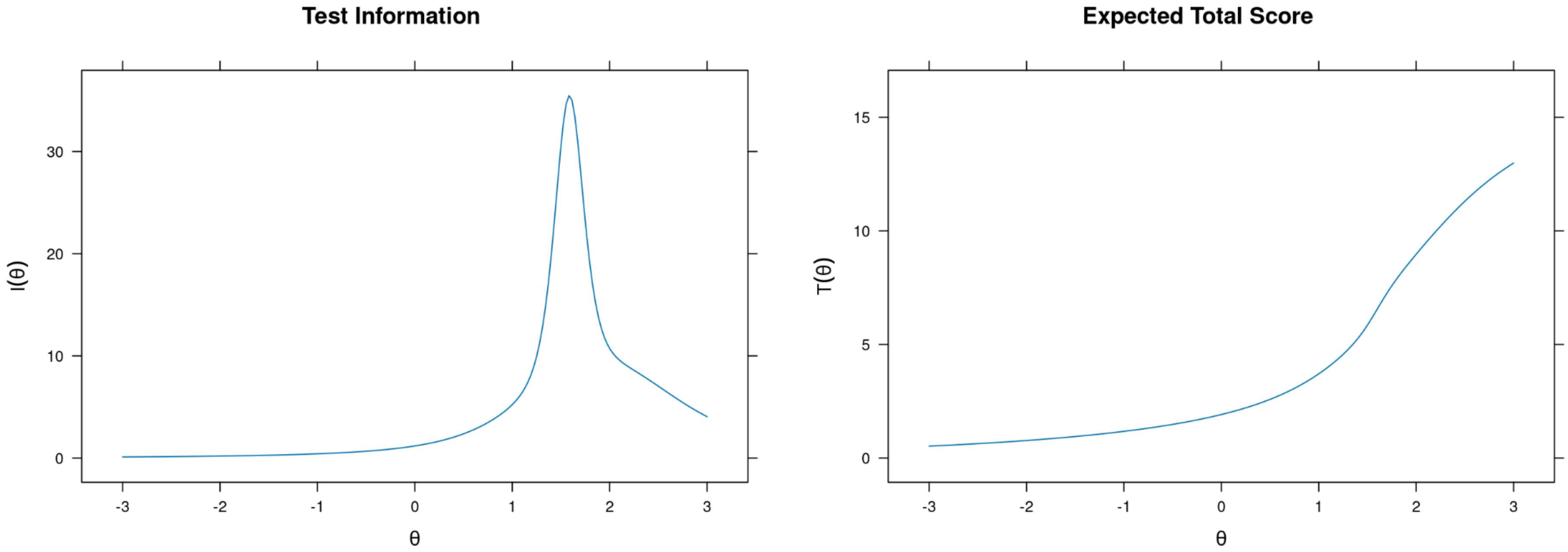

**Supplementary Figure 1.2.2 -** Child Autism Spectrum Test (CAST), Caregiver-report (Sociability/communication): item probability functions

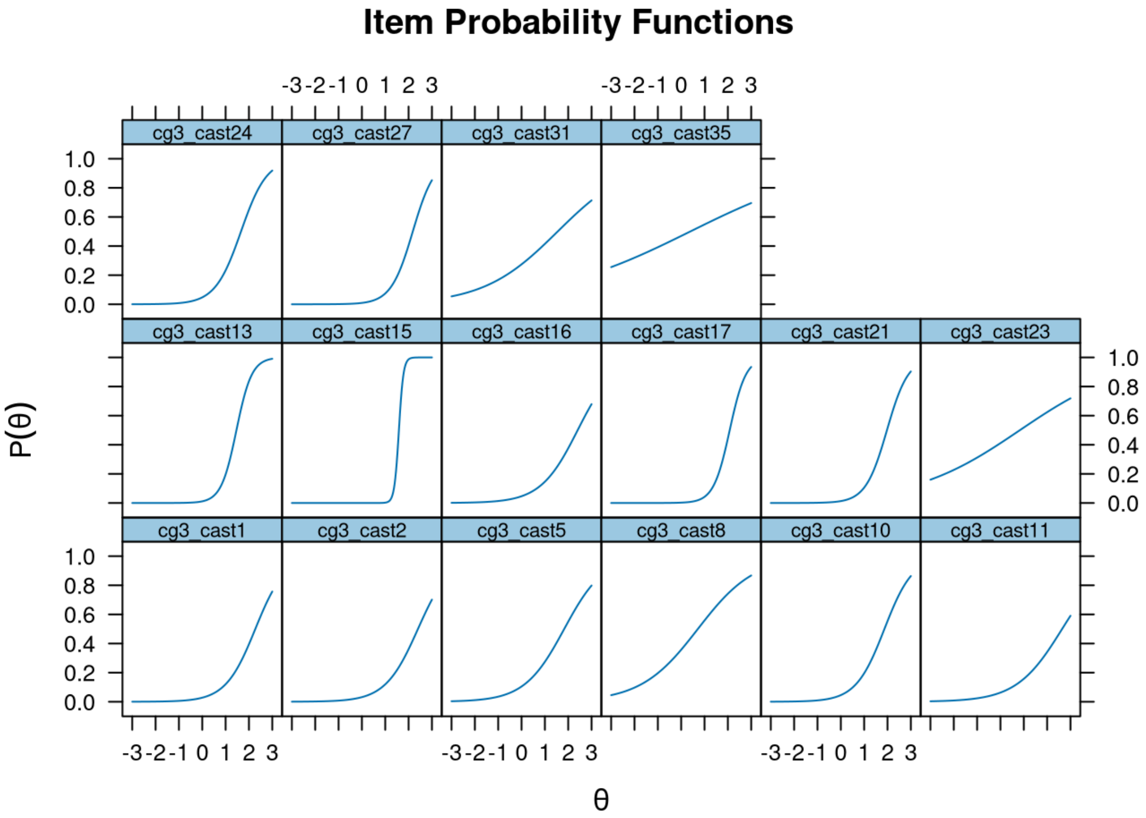

**Supplementary Figure 1.2.3 -** Child Autism Spectrum Test (CAST), Caregiver-report (Sociability/communication): item infit and outfit statistics

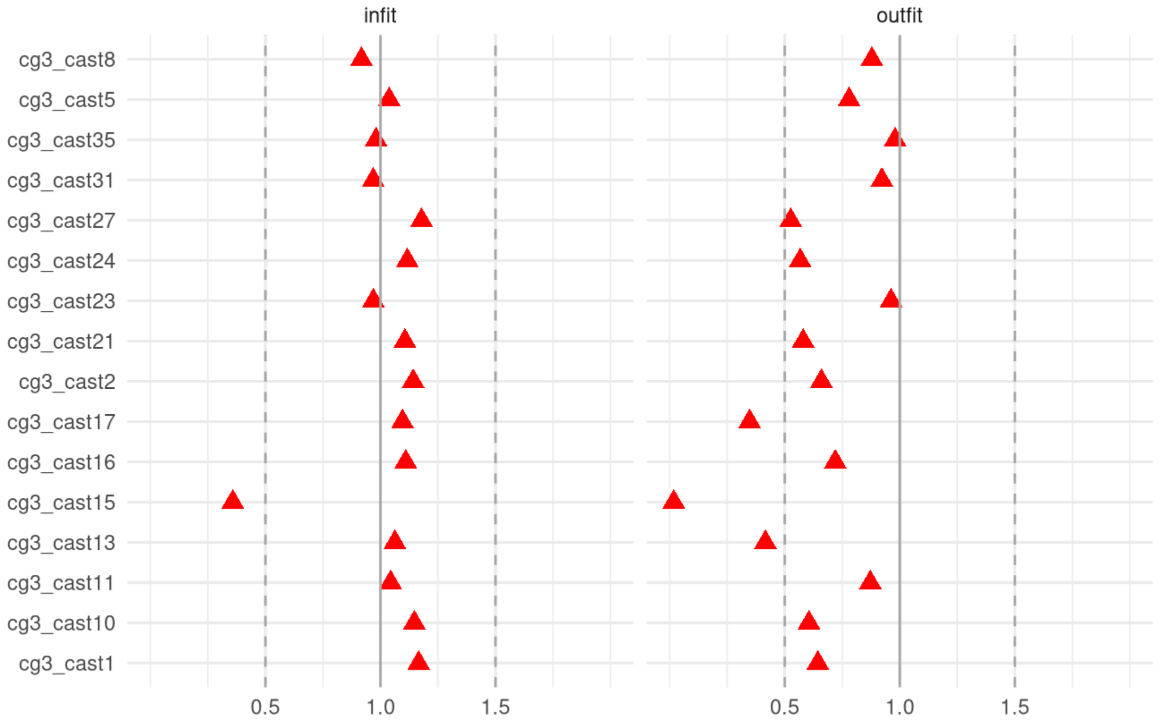

**Supplementary Figure 1.2.4 -** Child Autism Spectrum Test (CAST), Caregiver-report (Sociability/communication): person infit and outfit statistics

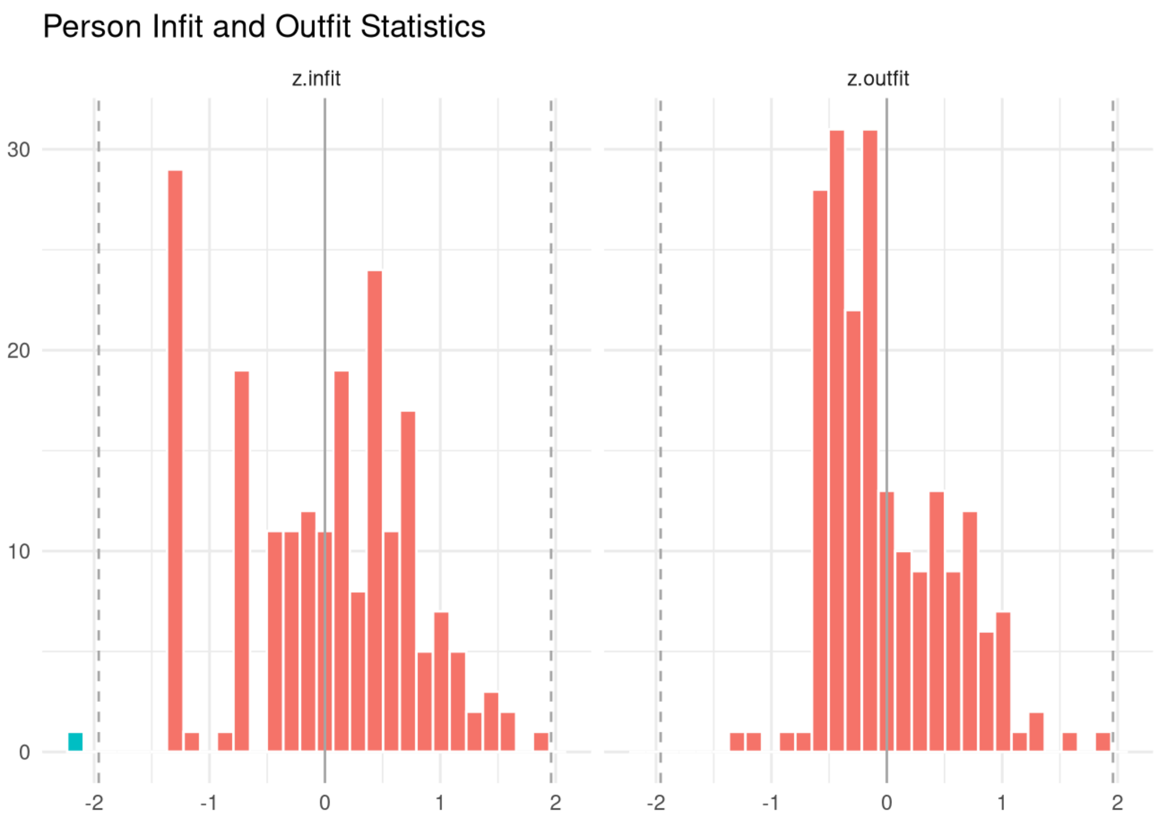

**Supplementary Figure 1.2.5 -** Child Autism Spectrum Test (CAST), Caregiver-report (Sociability/communication): item response functions

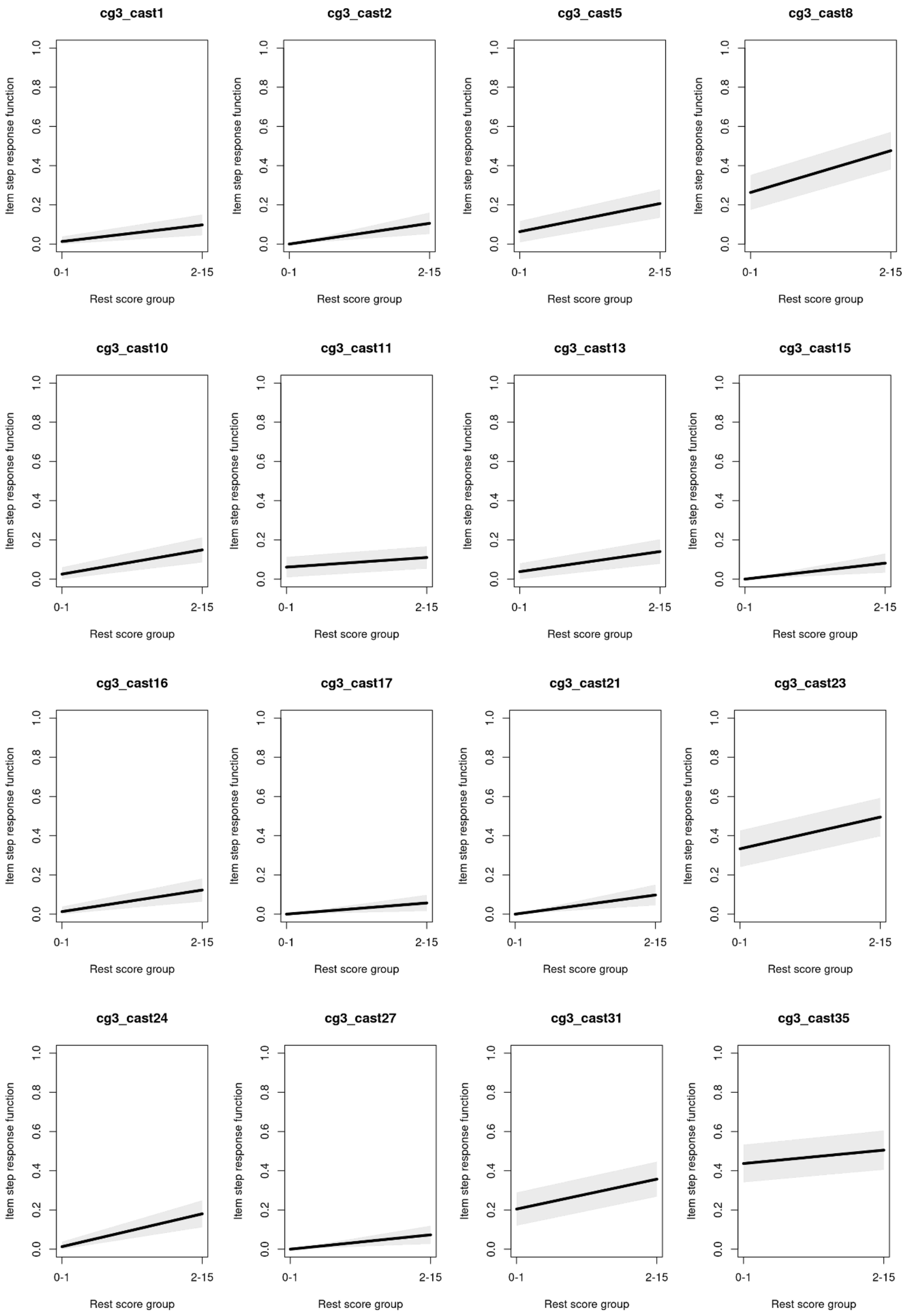

**Supplementary Figure 2.1** : Child and Adolescent Trauma Screen-2 (CATS-2), Caregiver-report: test information and expected scores

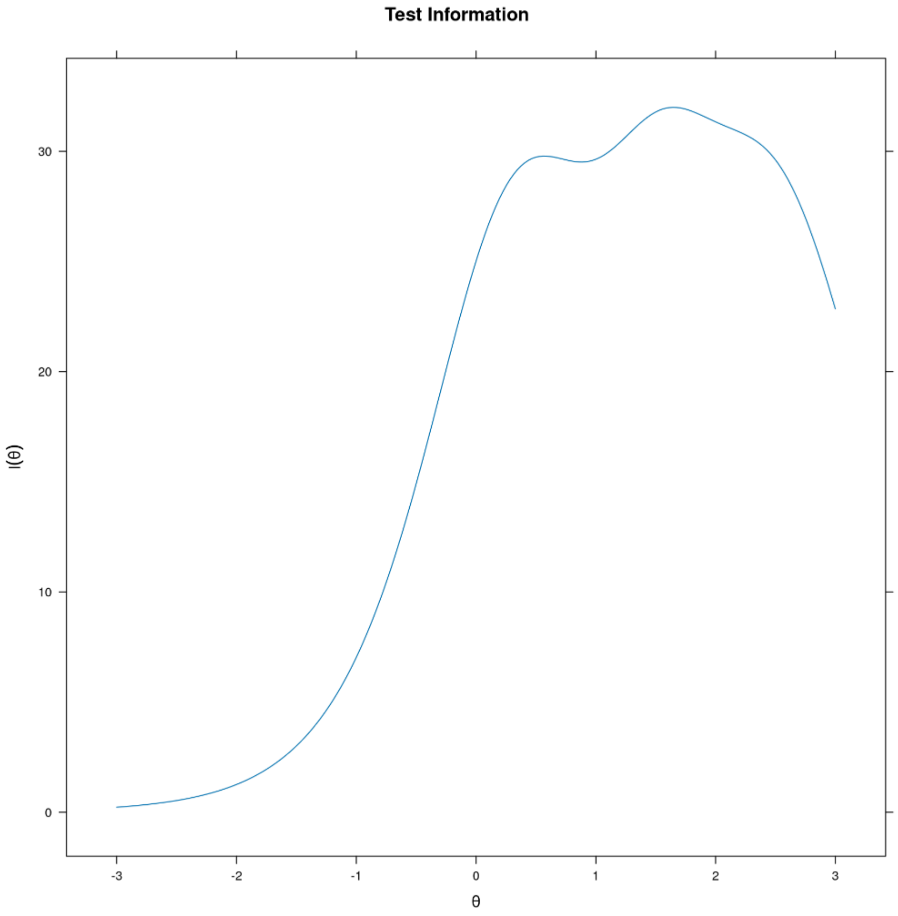

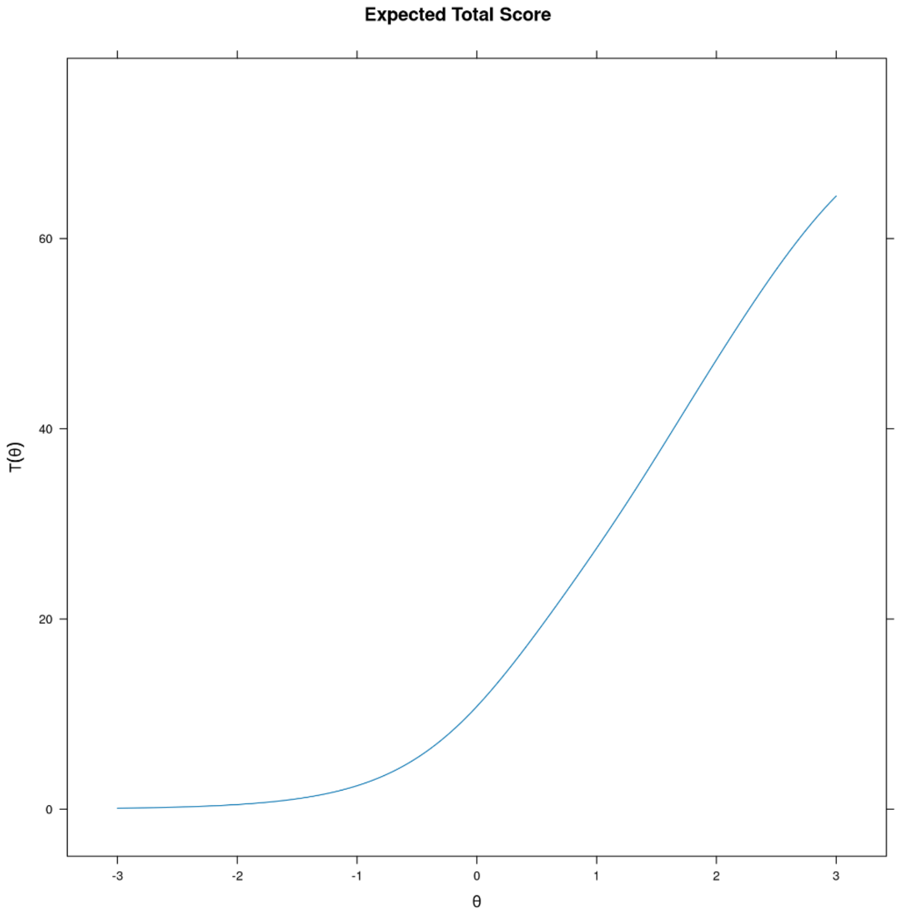

**Supplementary Figure 2.2**: Child and Adolescent Trauma Screen-2 (CATS-2), Caregiver-report: item probability functions

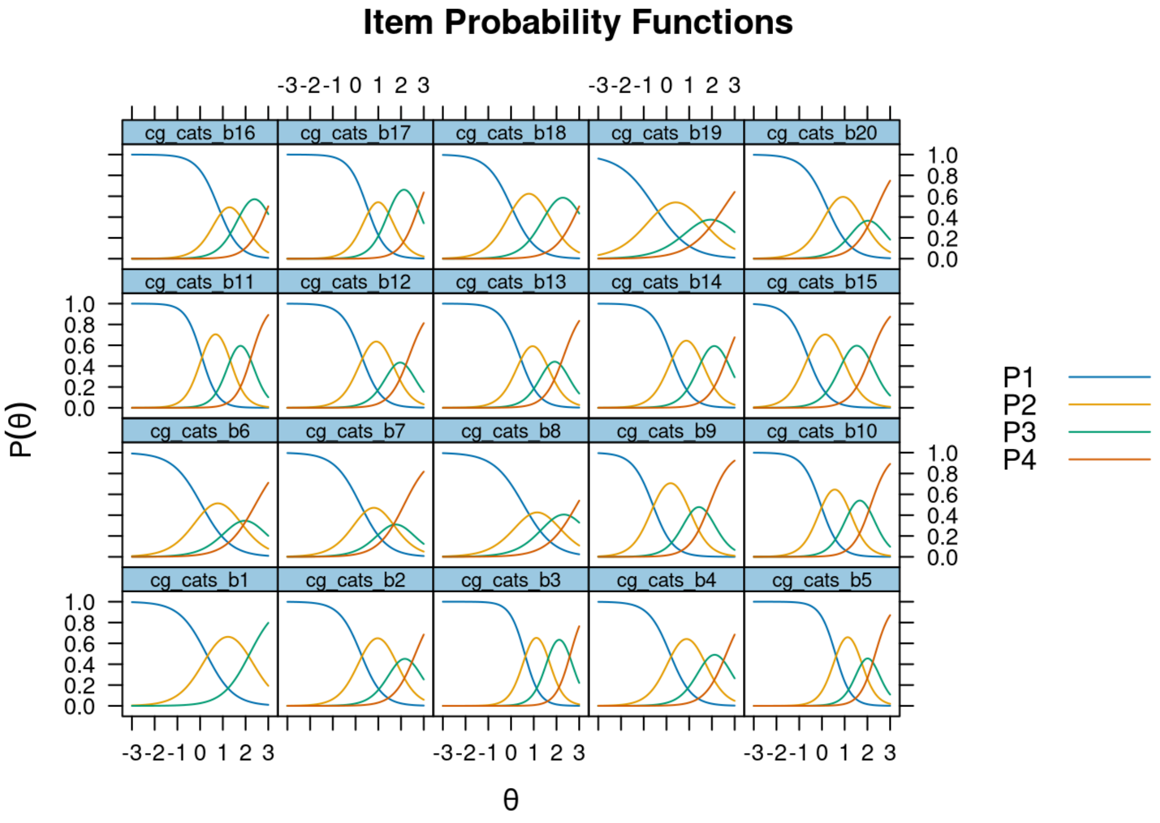

**Supplementary Figure 2.3**: Child and Adolescent Trauma Screen-2 (CATS-2), Caregiver-report: item infit and outfit statistics

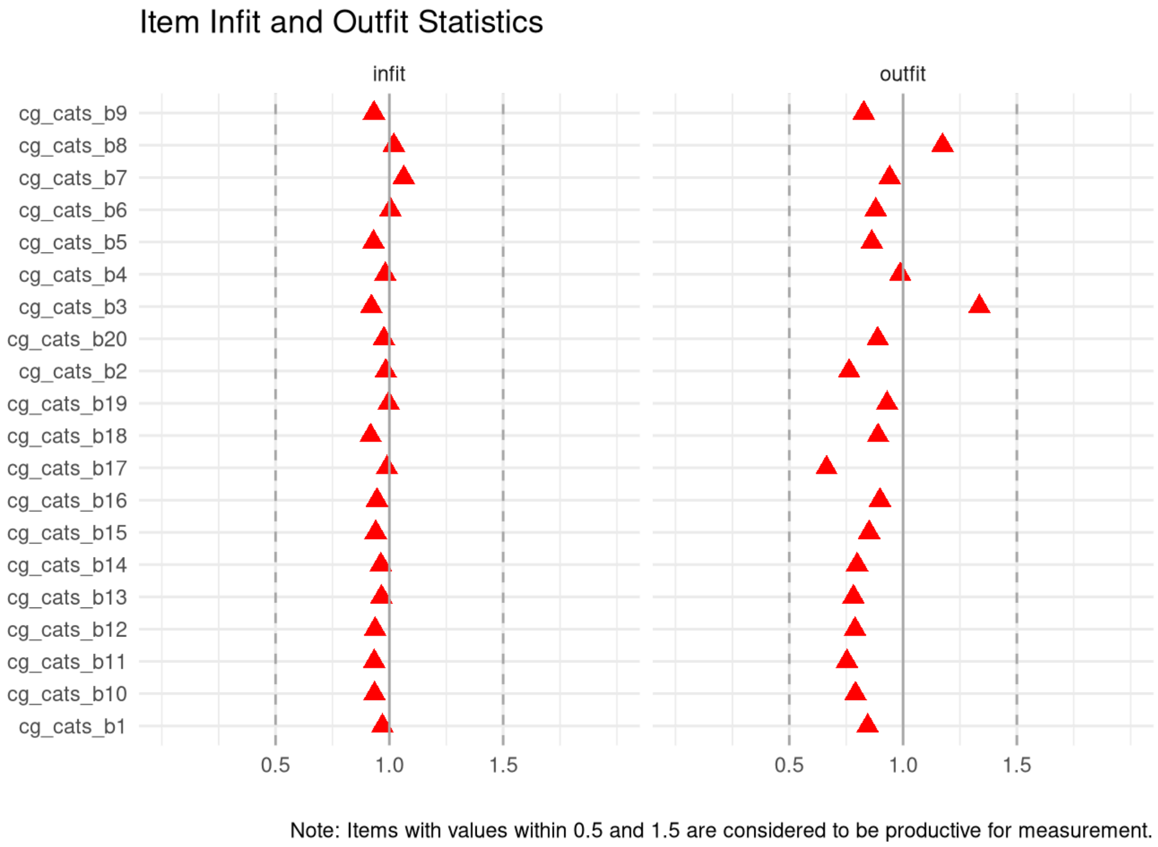

**Supplementary Figure 2.4**: Child and Adolescent Trauma Screen-2 (CATS-2), Caregiver-report: person infit and outfit statistics

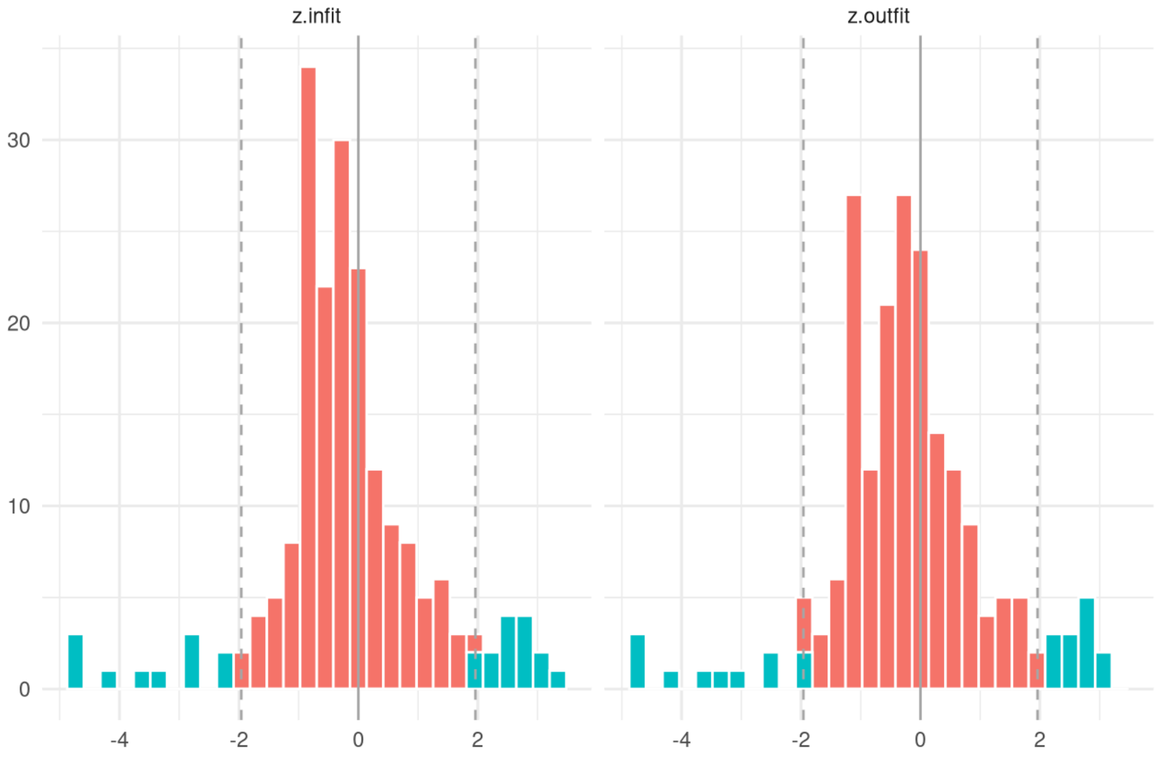

**Supplementary Figure 3.1** : Child and Adolescent Trauma Screen-2 (CATS-2), Self-report: test information and expected scores

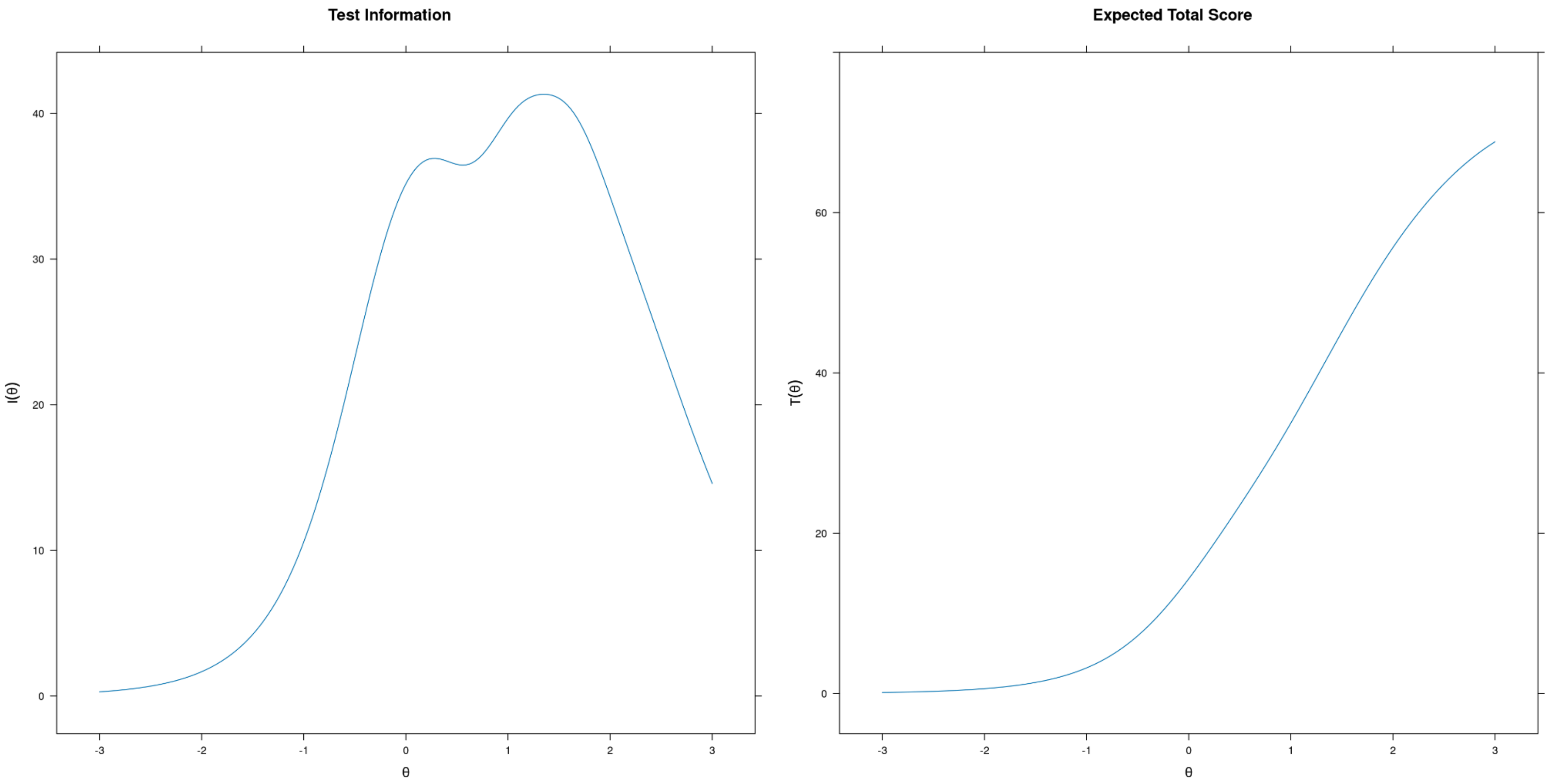

**Supplementary Figure 3.2**: Child and Adolescent Trauma Screen-2 (CATS-2), Self-report: item probability functions

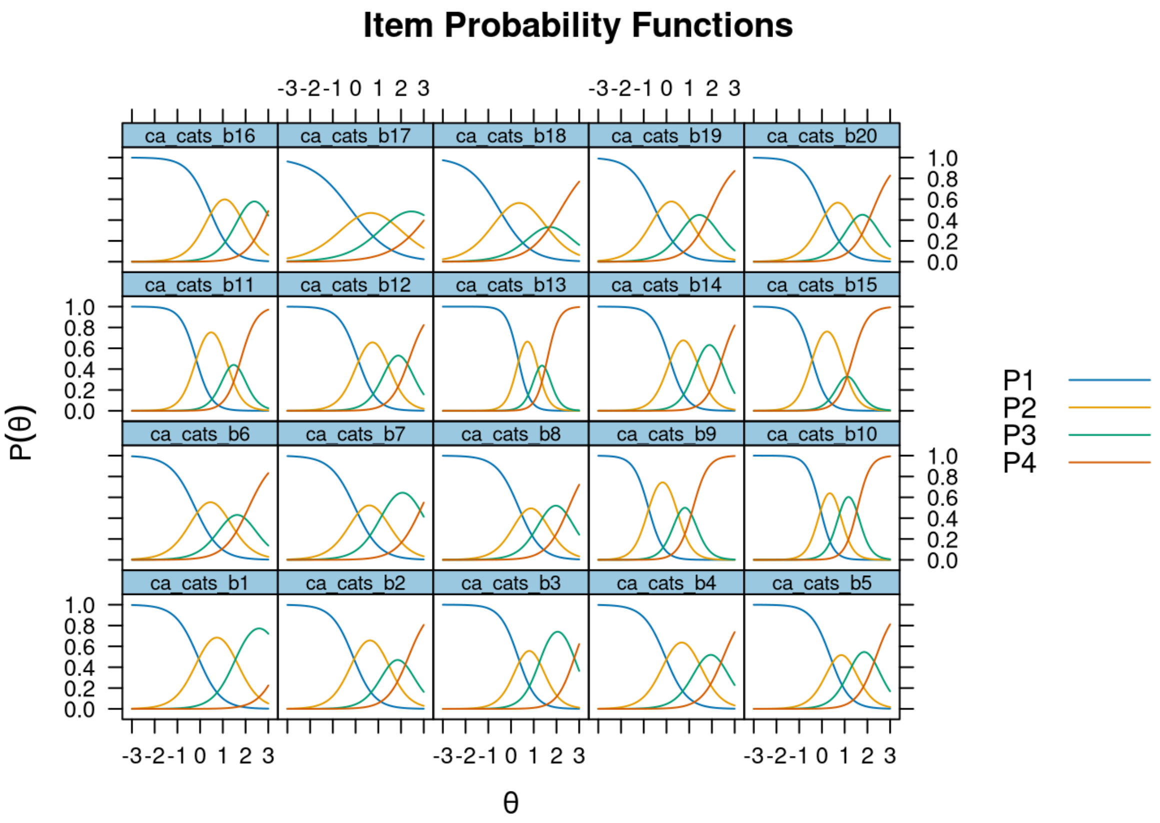

**Supplementary Figure 3.3**: Child and Adolescent Trauma Screen-2 (CATS-2), Self-report: item infit and outfit statistics

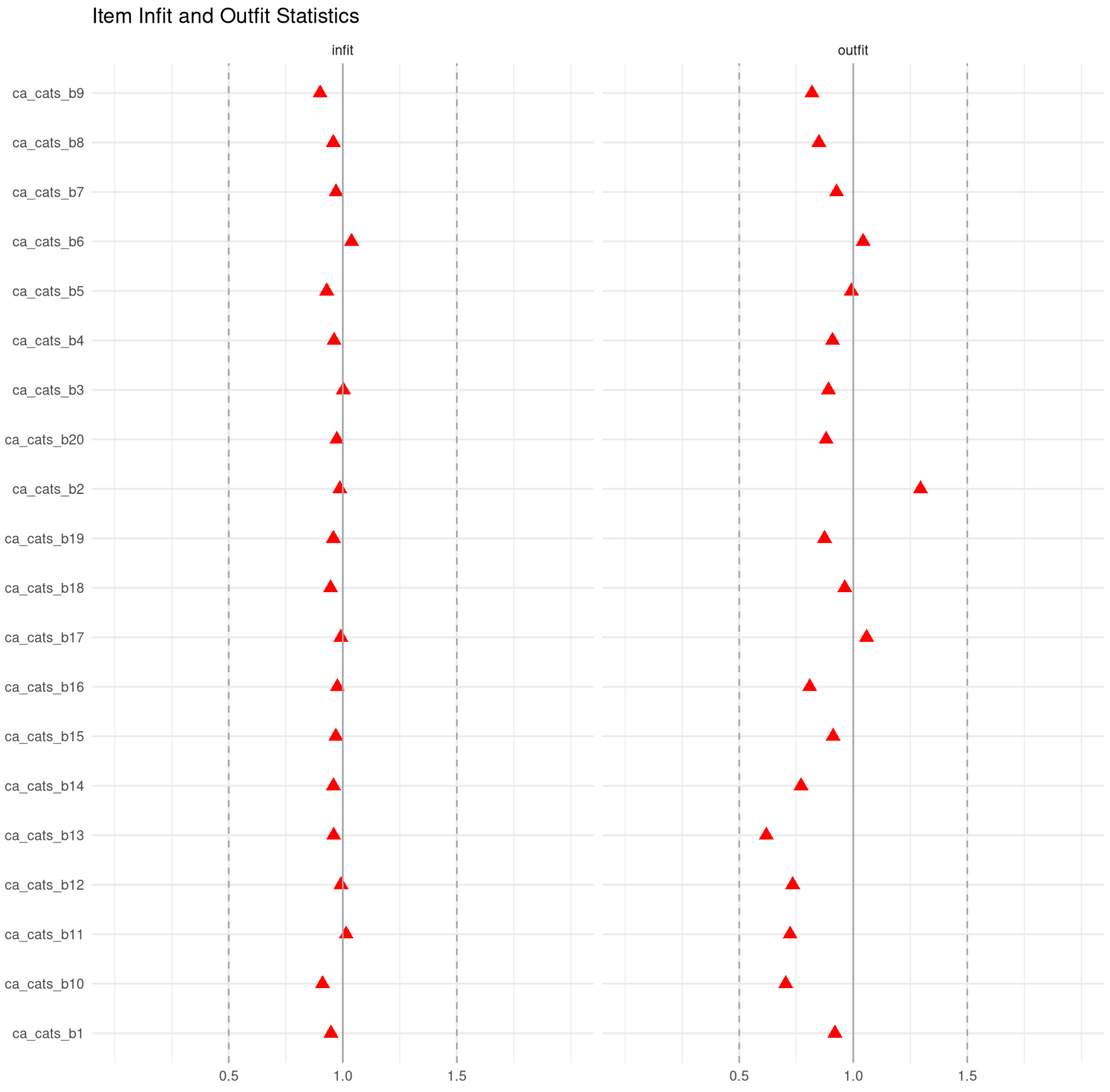

Note: Items with values within 0.5 and 1.5 are considered to be productive for measurement.

**Supplementary Figure 3.4**: Child and Adolescent Trauma Screen-2 (CATS-2), Self-report: person infit and outfit statistics

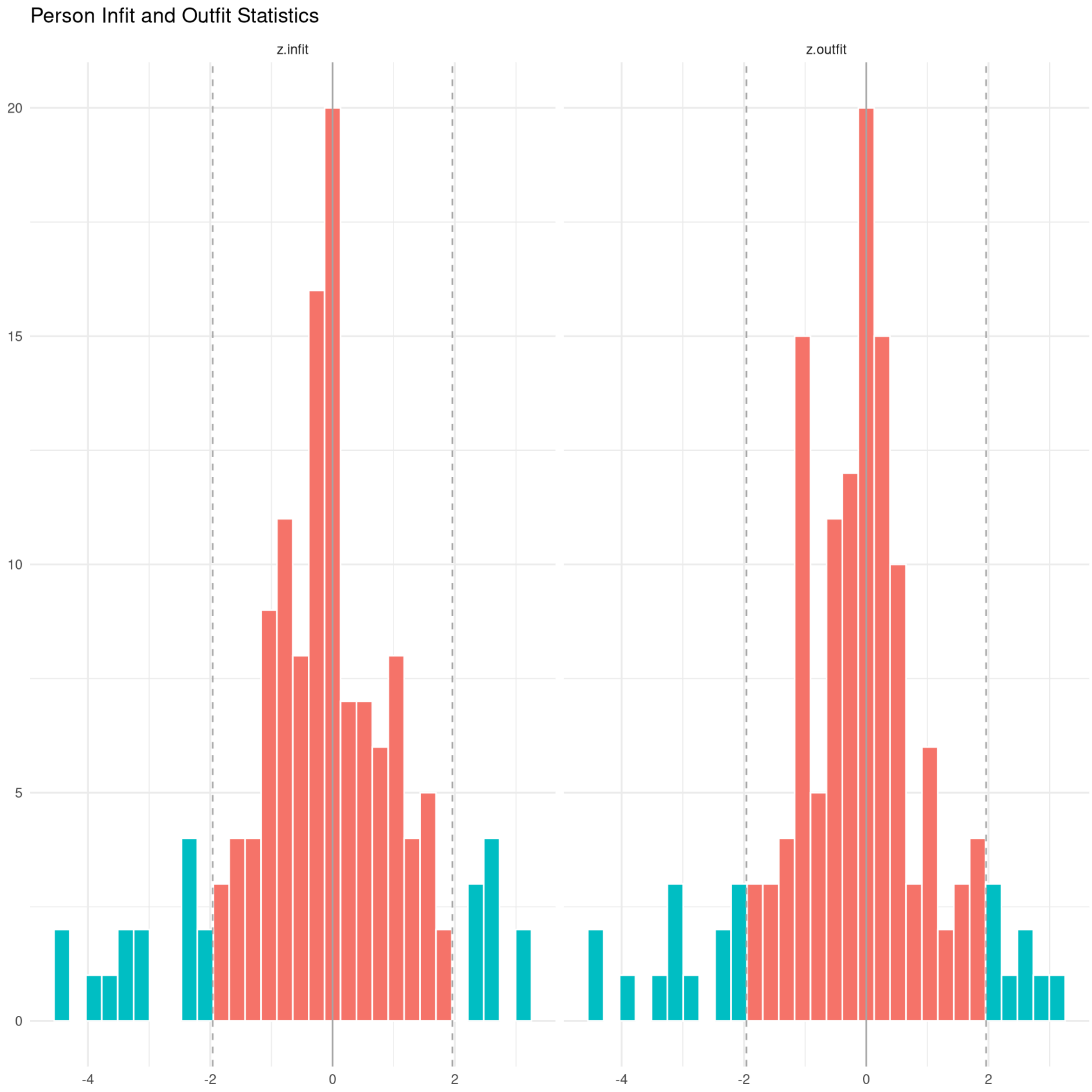

**Supplementary Figure 4.1 -** Modified Checklist for Autism in Toddlers (M-CHAT-R), Caregiver-report: test information and expected scores

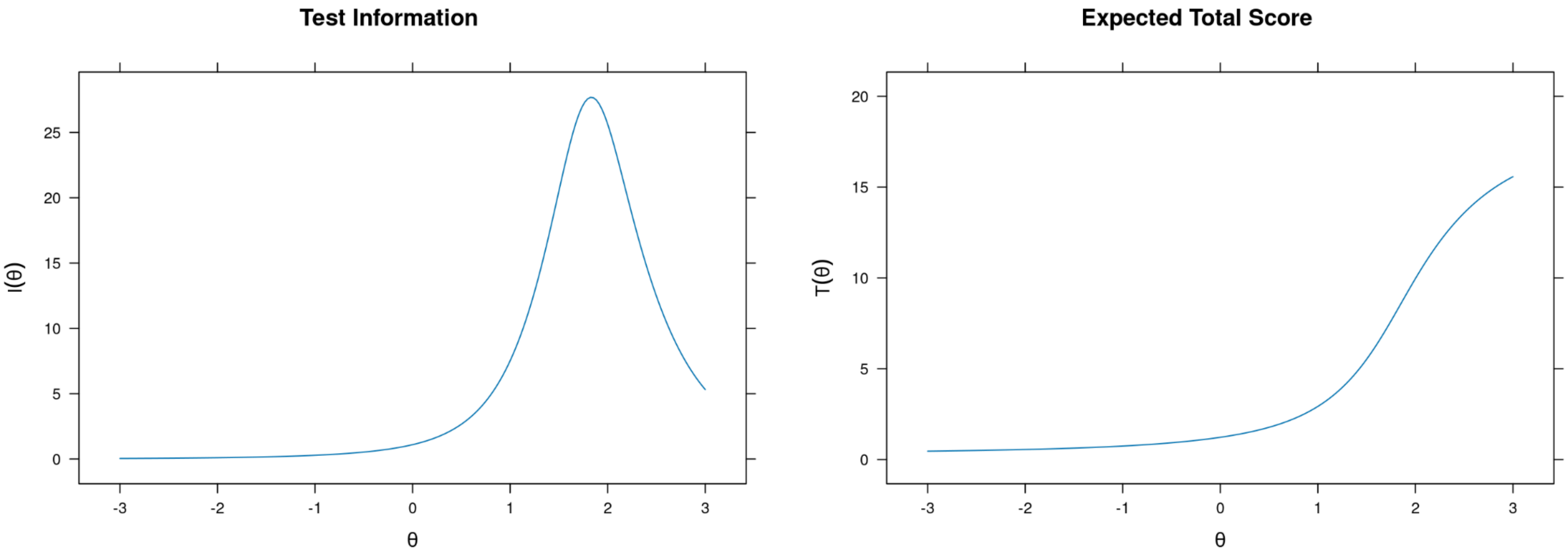

**Supplementary Figure 4.2 -** Modified Checklist for Autism in Toddlers (M-CHAT-R), Caregiver-report: item probability functions

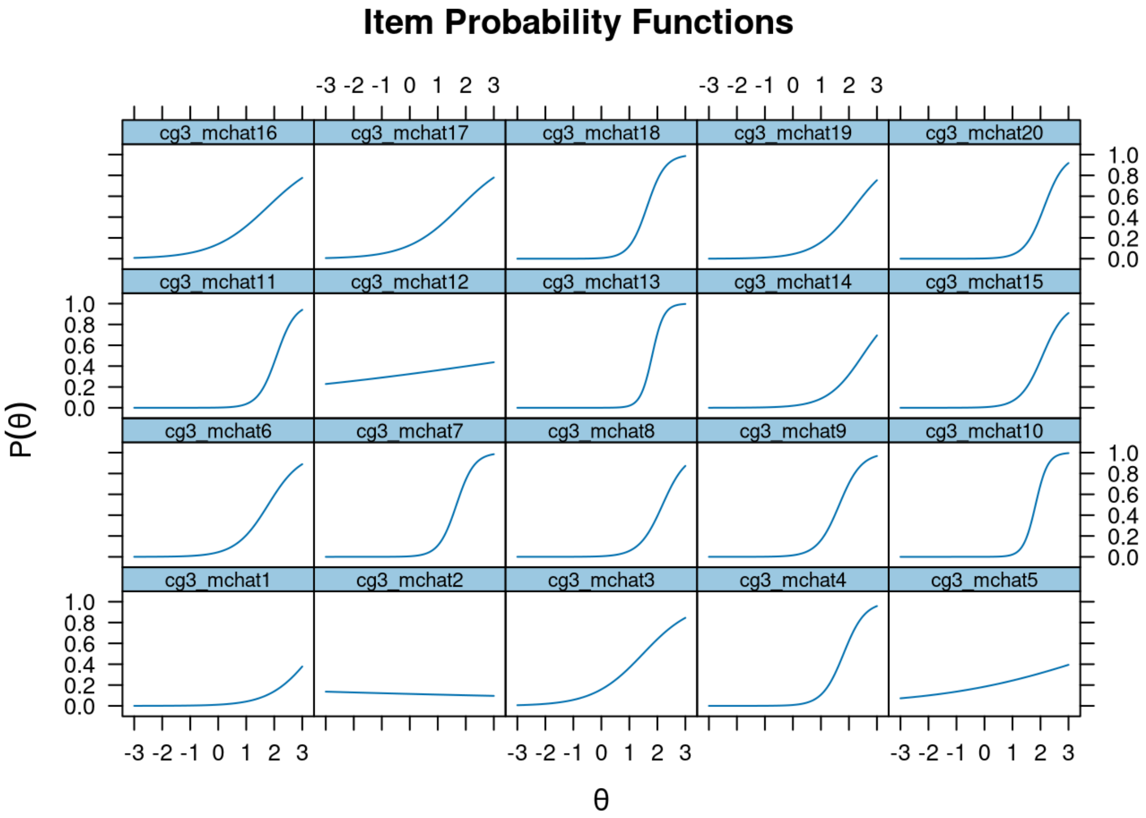

**Supplementary Figure 4.3 -** Modified Checklist for Autism in Toddlers (M-CHAT-R), Caregiver-report: item infit and outfit statistics

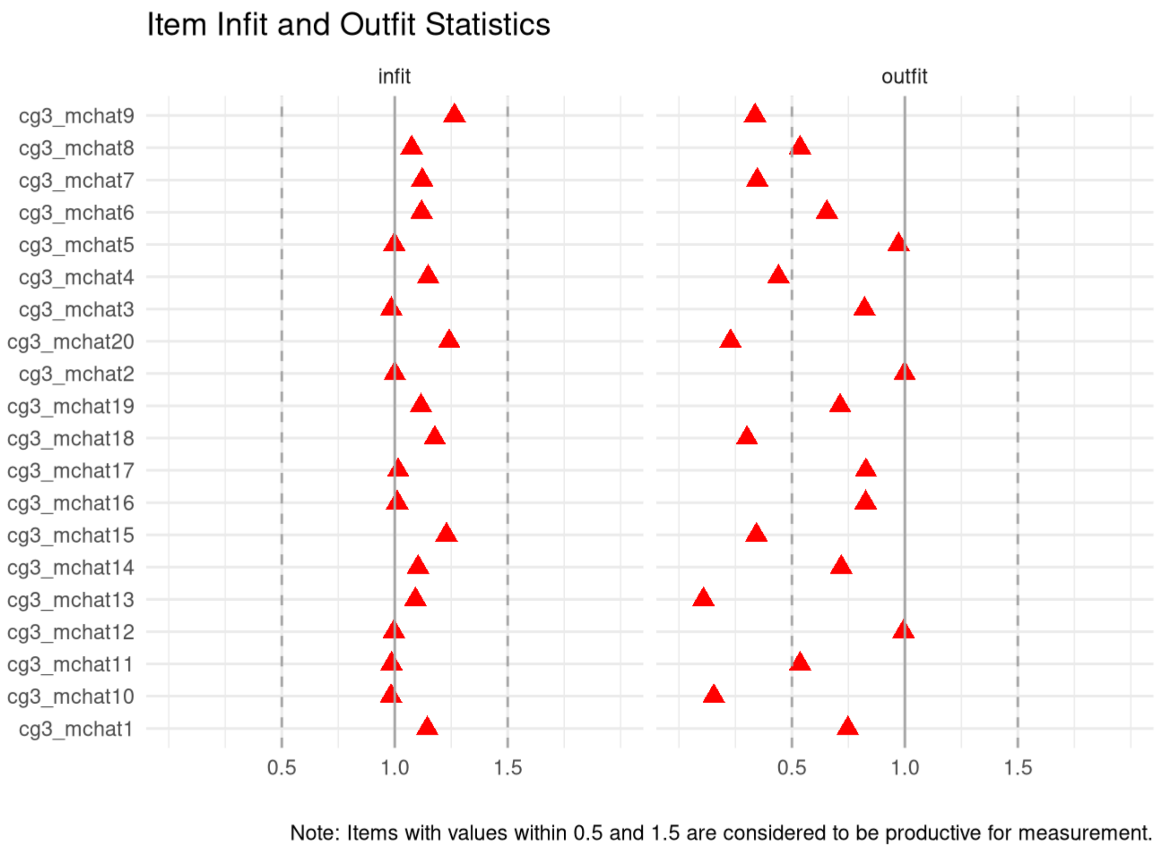

**Supplementary Figure 4.4 -** Modified Checklist for Autism in Toddlers (M-CHAT-R), Caregiver-report: person infit and outfit statistics

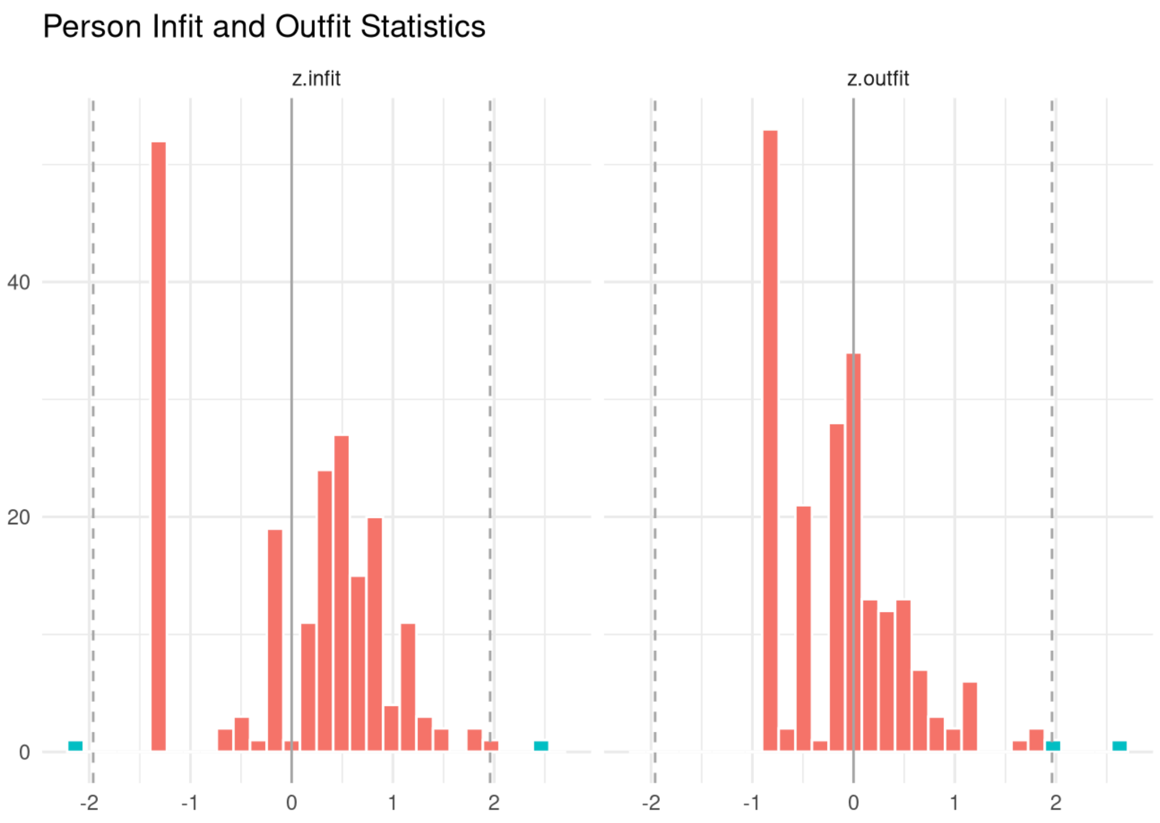

**Supplementary Figure 4.5 -** Modified Checklist for Autism in Toddlers (M-CHAT-R), Caregiver-report: item response functions

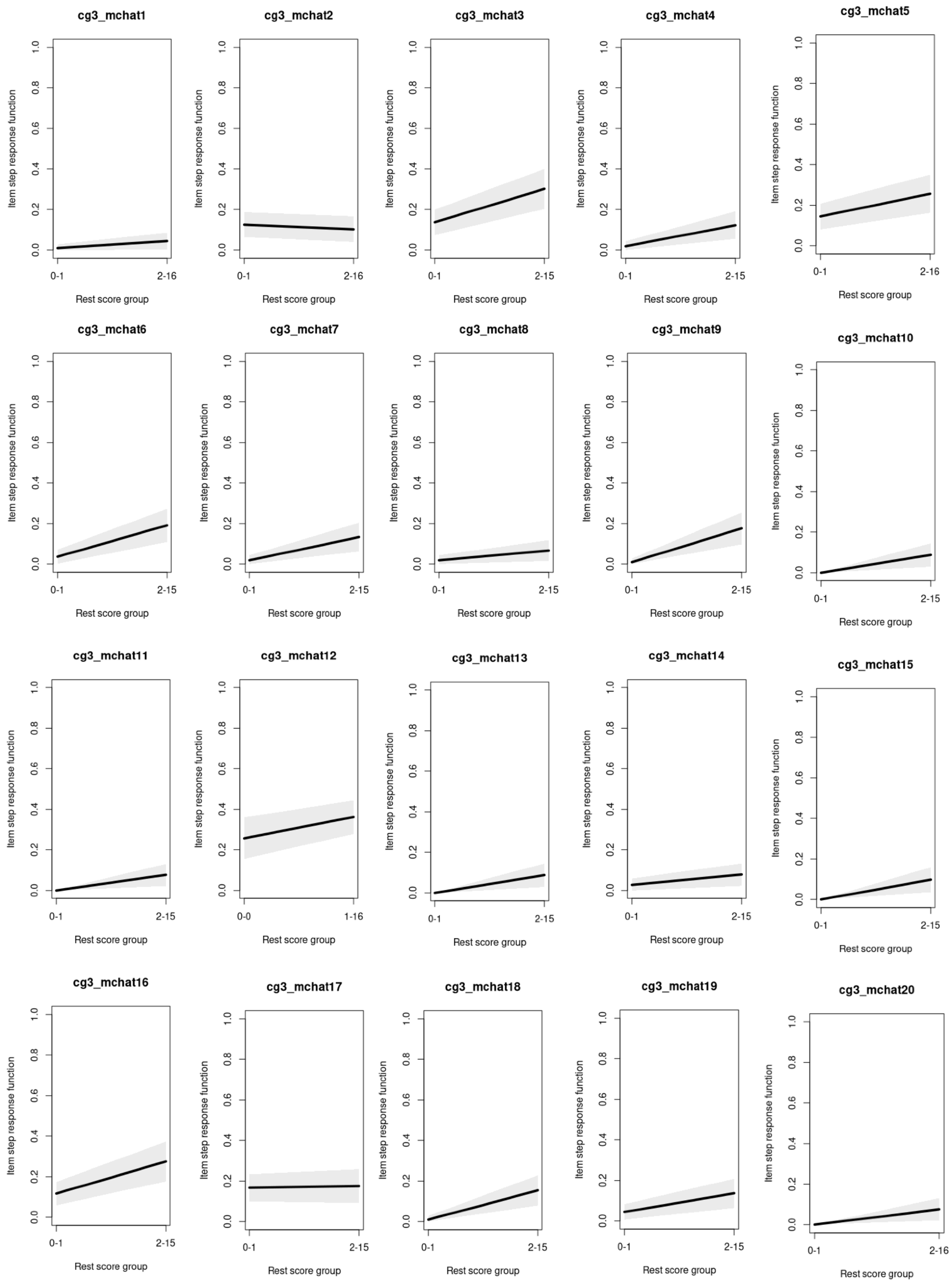

**Supplementary Figure 5.1.1 -** Pediatric Symptom Checklist short version (PSC-17), age under 6 years, Caregiver-report (Attention Scale): test information and expected scores

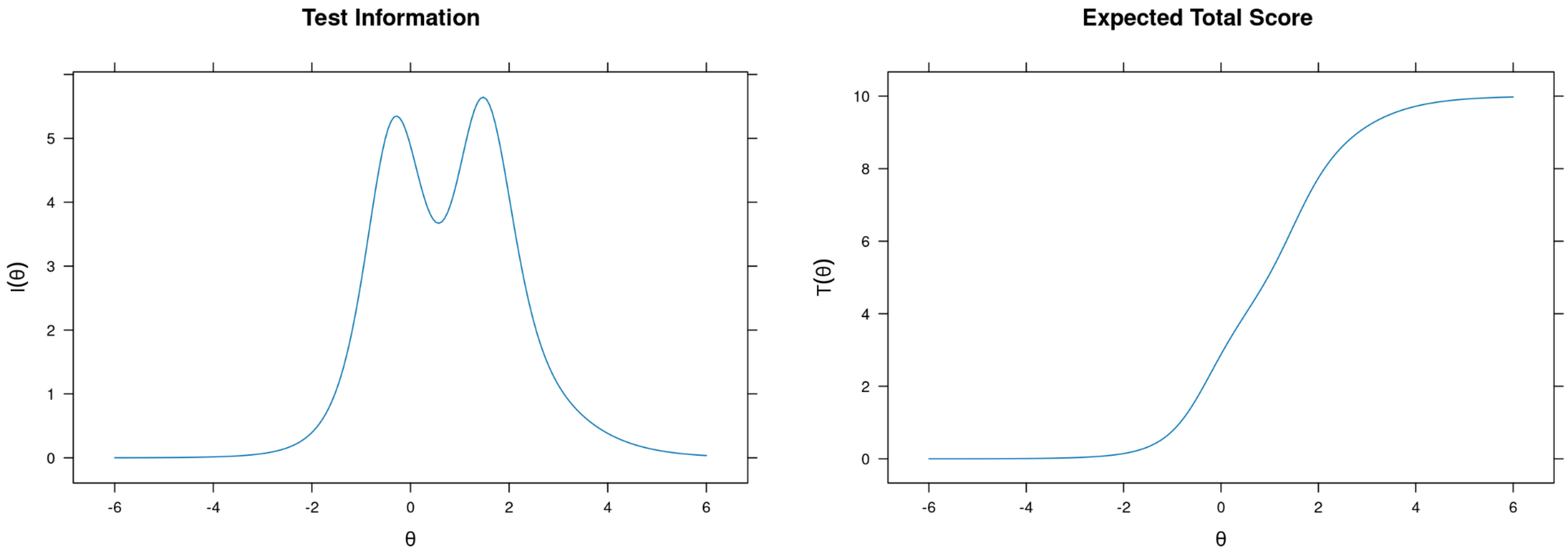

**Supplementary Figure 5.1.2 -** Pediatric Symptom Checklist short version (PSC-17), age under 6 years, Caregiver-report (Attention Scale): item probability functions

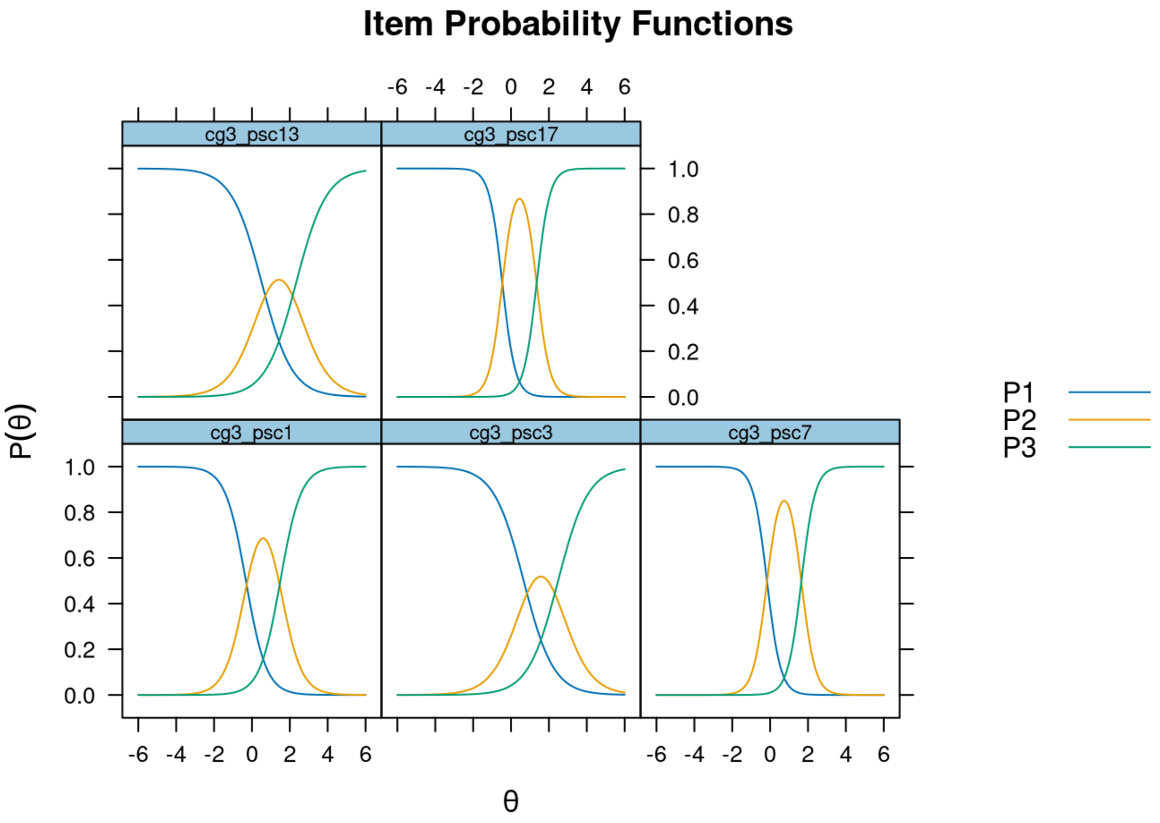

**Supplementary Figure 5.1.3 -** Pediatric Symptom Checklist short version (PSC-17), age under 6 years, Caregiver-report (Attention Scale): item infit and outfit statistics

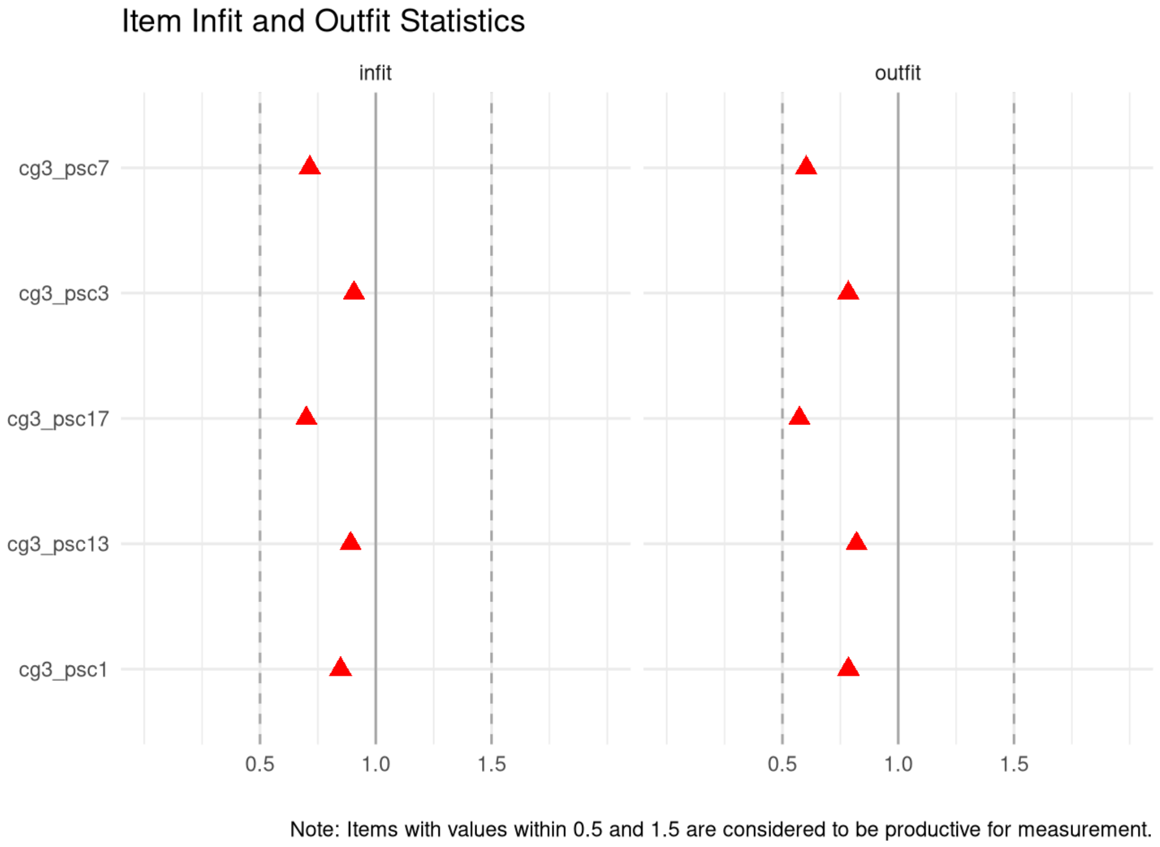

**Supplementary Figure 5.1.4 -** Pediatric Symptom Checklist short version (PSC-17), age under 6 years, Caregiver-report (Attention Scale): person infit and outfit statistics

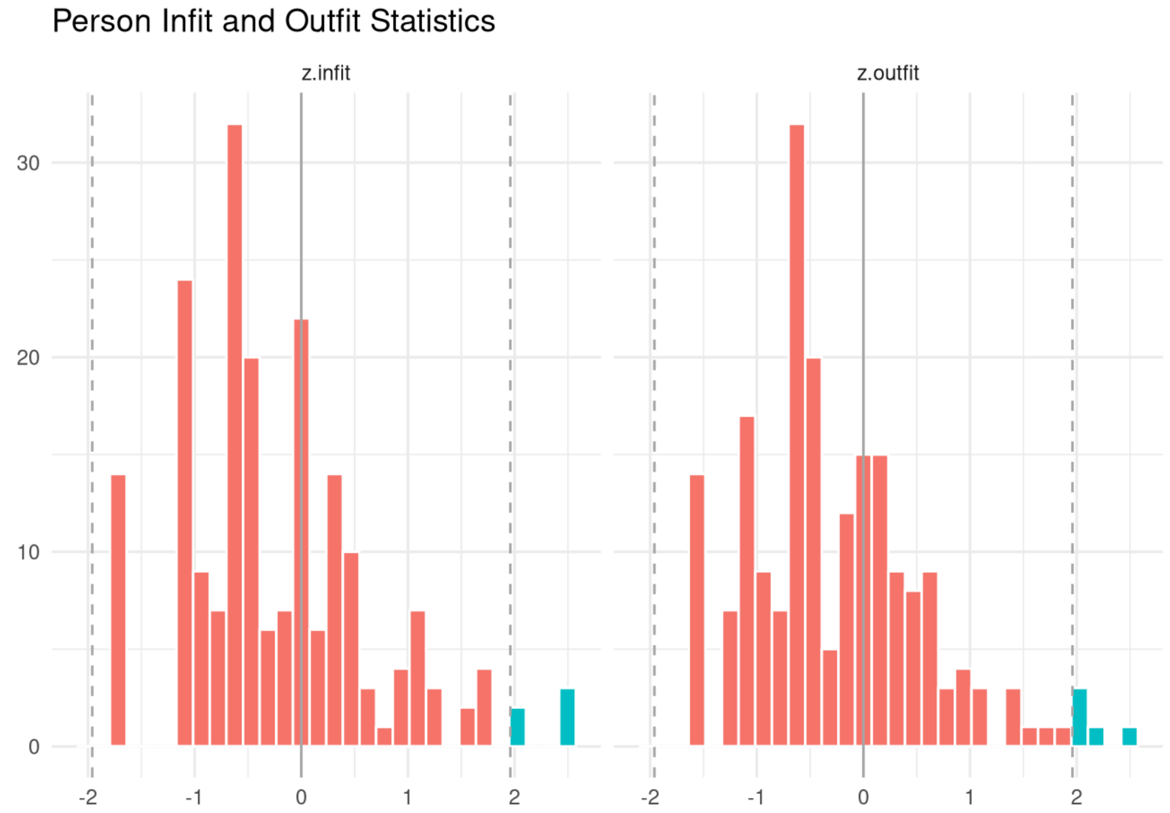

**Supplementary Figure 5.2.1 -** Pediatric Symptom Checklist short version (PSC-17), age under 6 years, Caregiver-report (Externalizing Scale): test information and expected scores

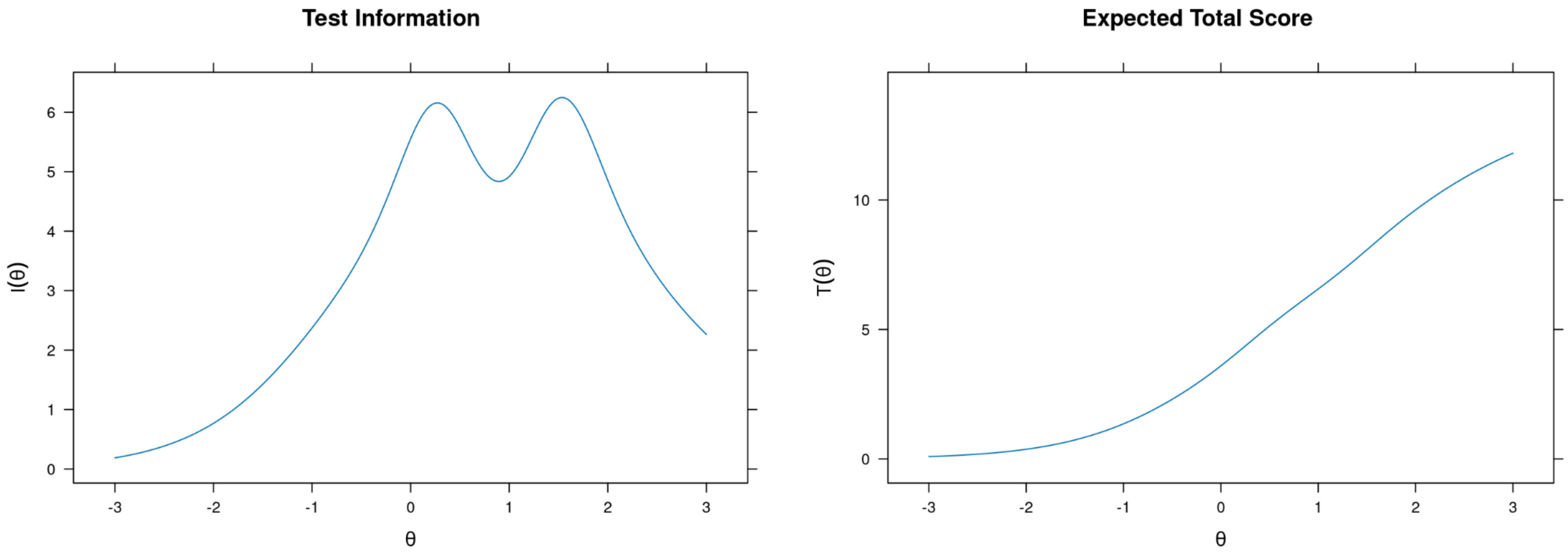

**Supplementary Figure 5.2.2 -** Pediatric Symptom Checklist short version (PSC-17), age under 6 years, Caregiver-report (Externalizing Scale): item probability functions

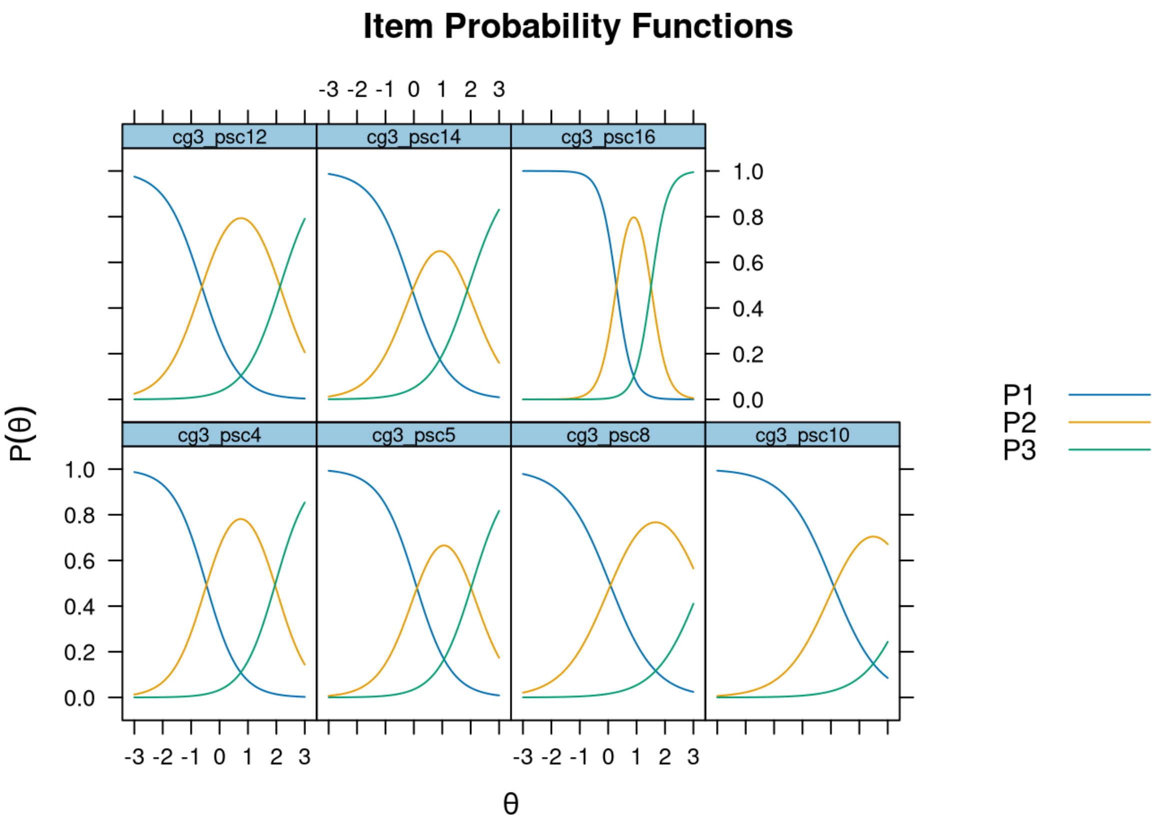

**Supplementary Figure 5.2.3 -** Pediatric Symptom Checklist short version (PSC-17), age under 6 years, Caregiver-report (Externalizing Scale): item infit and outfit statistics

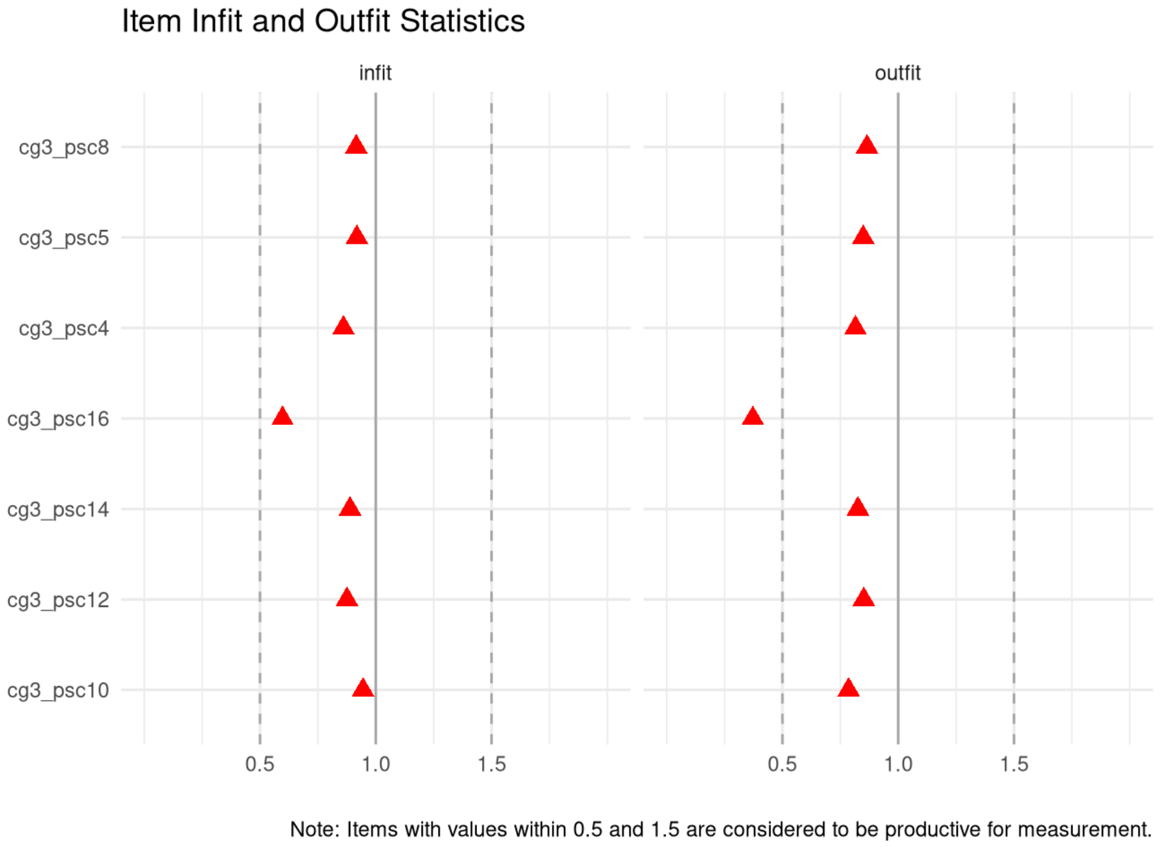

**Supplementary Figure 5.2.4 -** Pediatric Symptom Checklist short version (PSC-17), age under 6 years, Caregiver-report (Externalizing Scale): person infit and outfit statistics

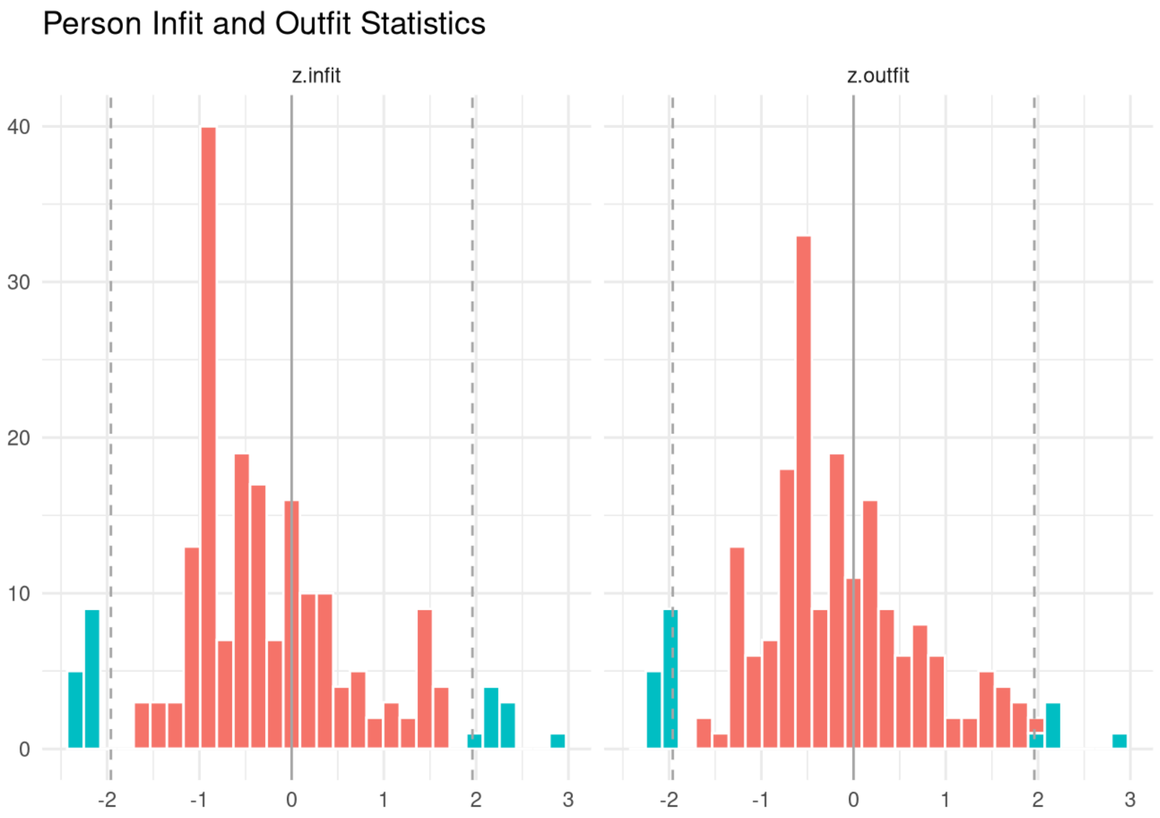

**Supplementary Figure 5.3.1 -** Pediatric Symptom Checklist short version (PSC-17), age under 6 years, Caregiver-report (Internalizing Scale): test information and expected scores

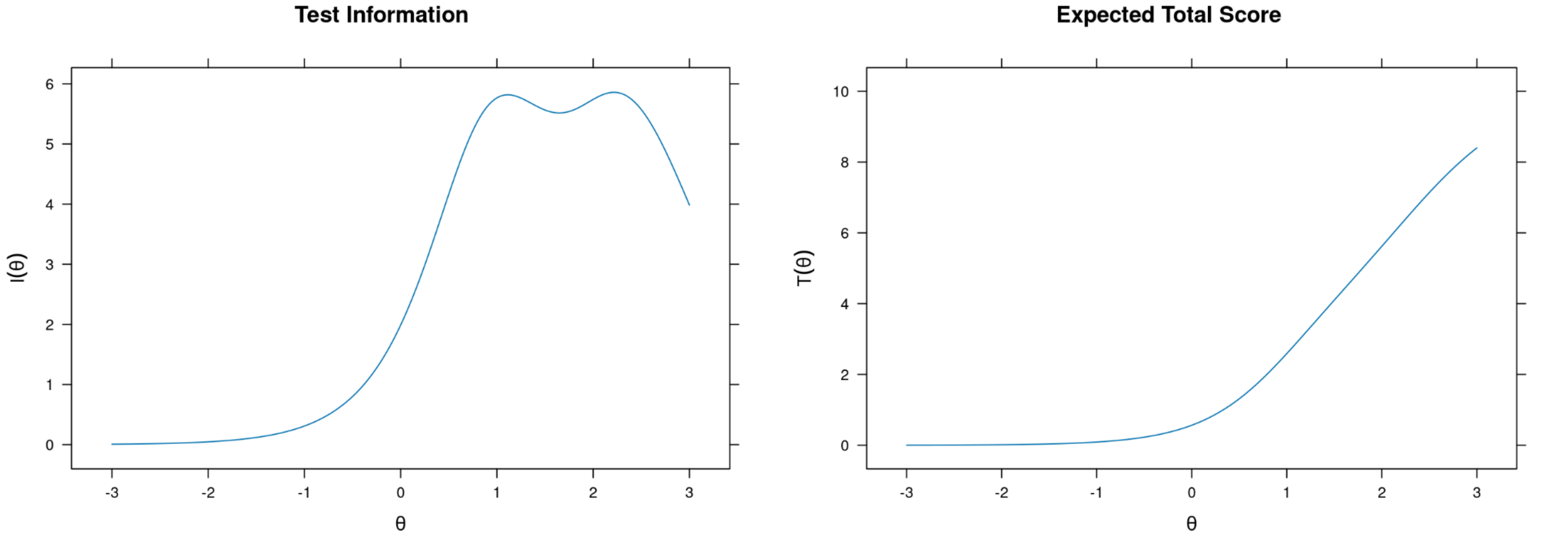

**Supplementary Figure 5.3.2 -** Pediatric Symptom Checklist short version (PSC-17), age under 6 years, Caregiver-report (Internalizing Scale): item probability functions

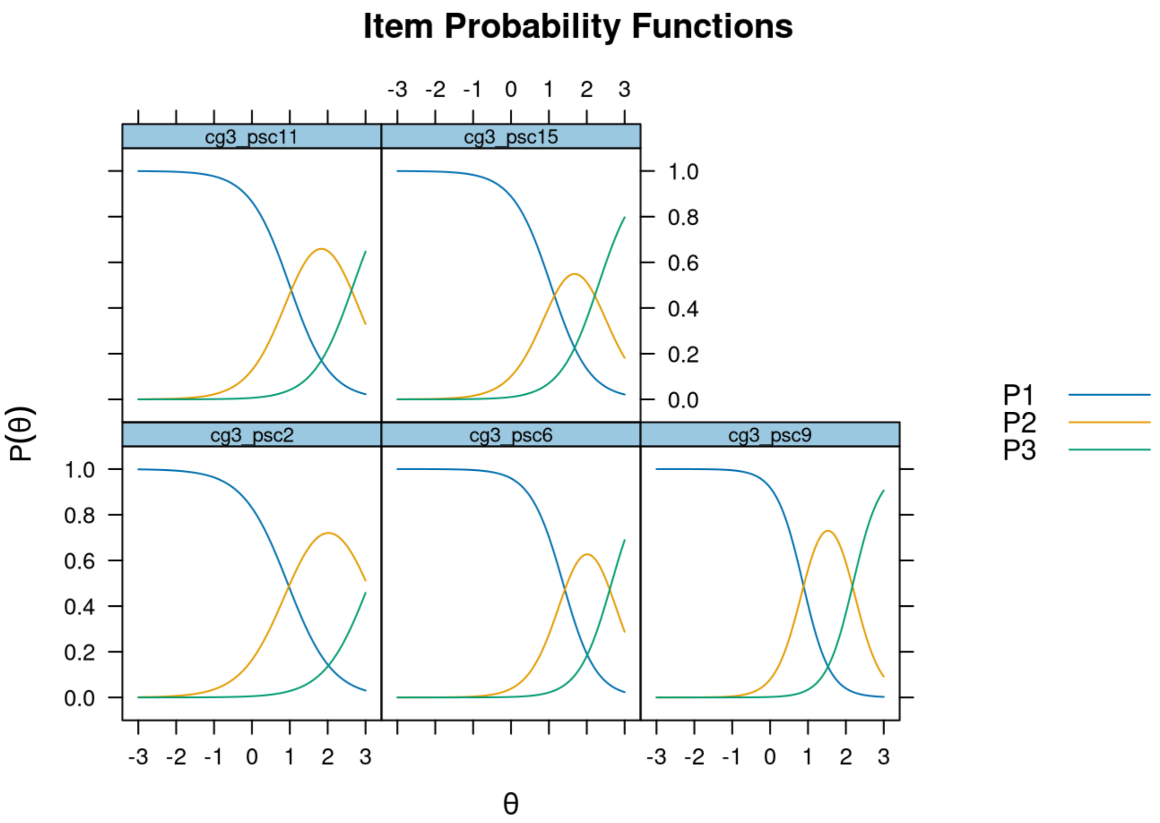

**Supplementary Figure 5.3.3 -** Pediatric Symptom Checklist short version (PSC-17), age under 6 years, Caregiver-report (Internalizing Scale): item infit and outfit statistics

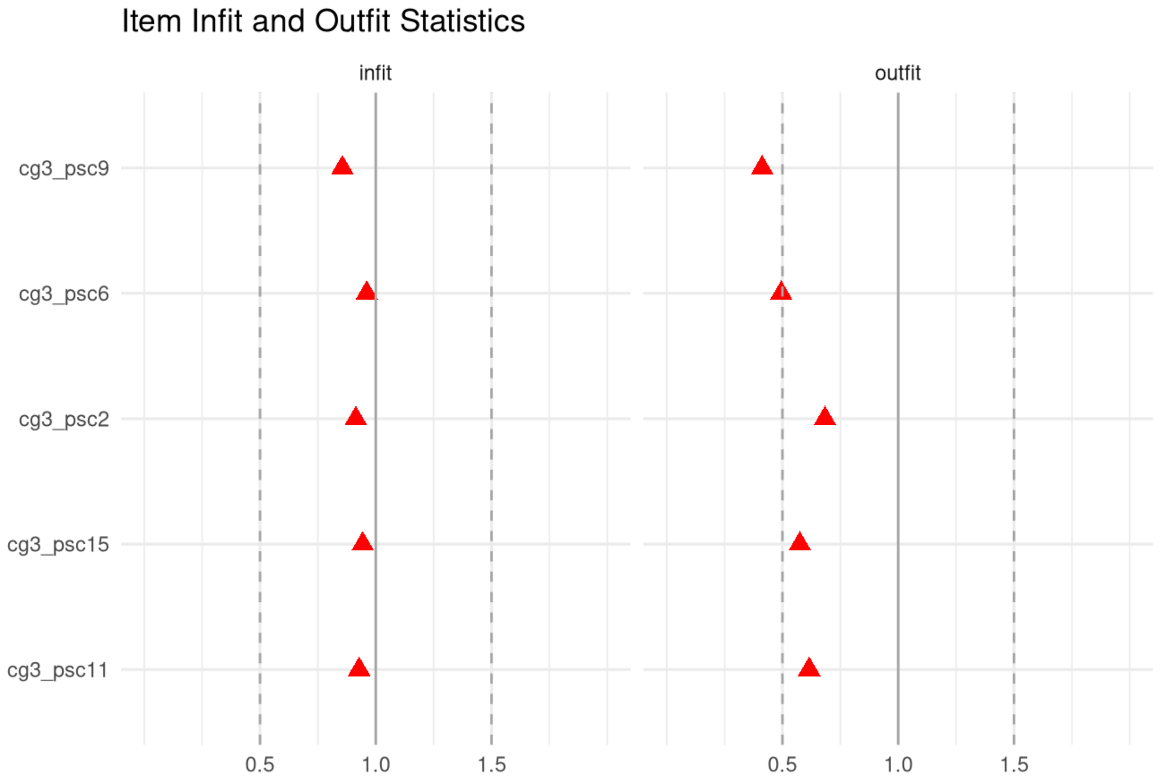

Note: Items with values within 0.5 and 1.5 are considered to be productive for measurement.

**Supplementary Figure 5.3.4 -** Pediatric Symptom Checklist short version (PSC-17), age under 6 years, Caregiver-report (Internalizing Scale): person infit and outfit statistics

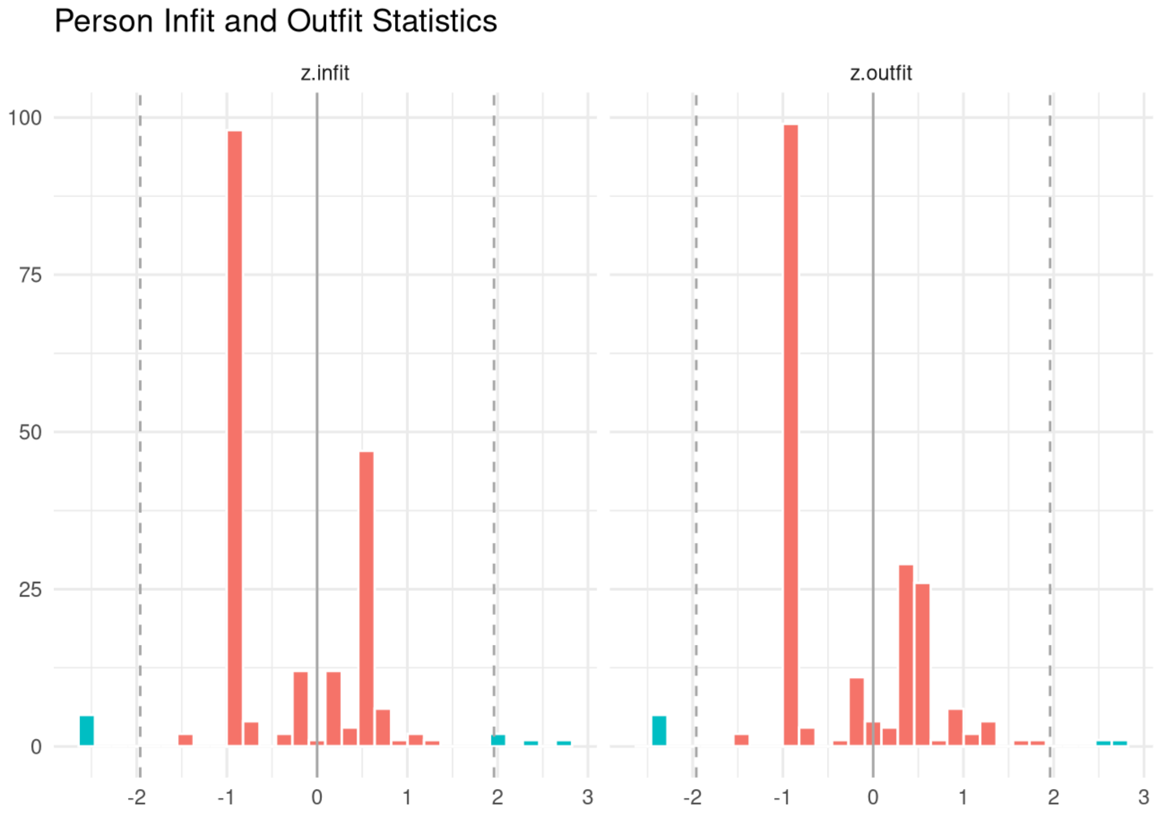

**Supplementary Figure 6.1.1 -** Pediatric Symptom Checklist  Short Version (PSC-17), 6- to 18-year-olds, Caregiver-report (Attention Scale): test information and expected scores

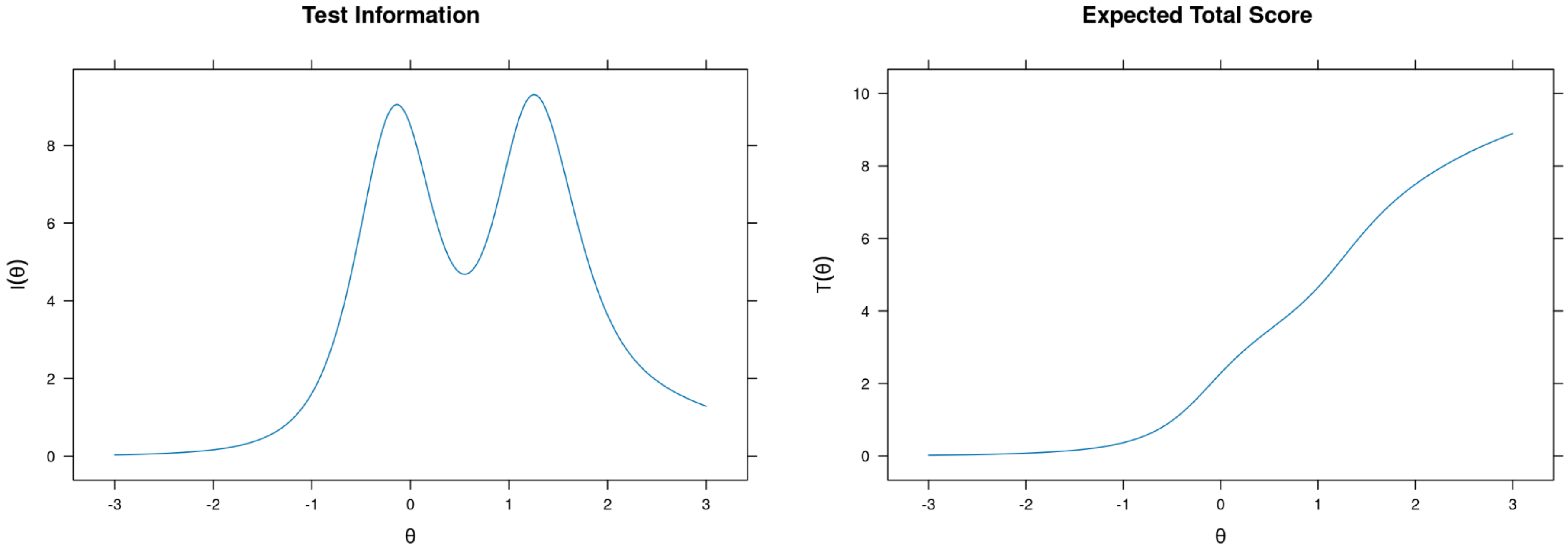

**Supplementary Figure 6.1.2 -** Pediatric Symptom Checklist Short Version (PSC-17), 6- to 18-year-olds, Caregiver-report (Attention Scale): item probability functions

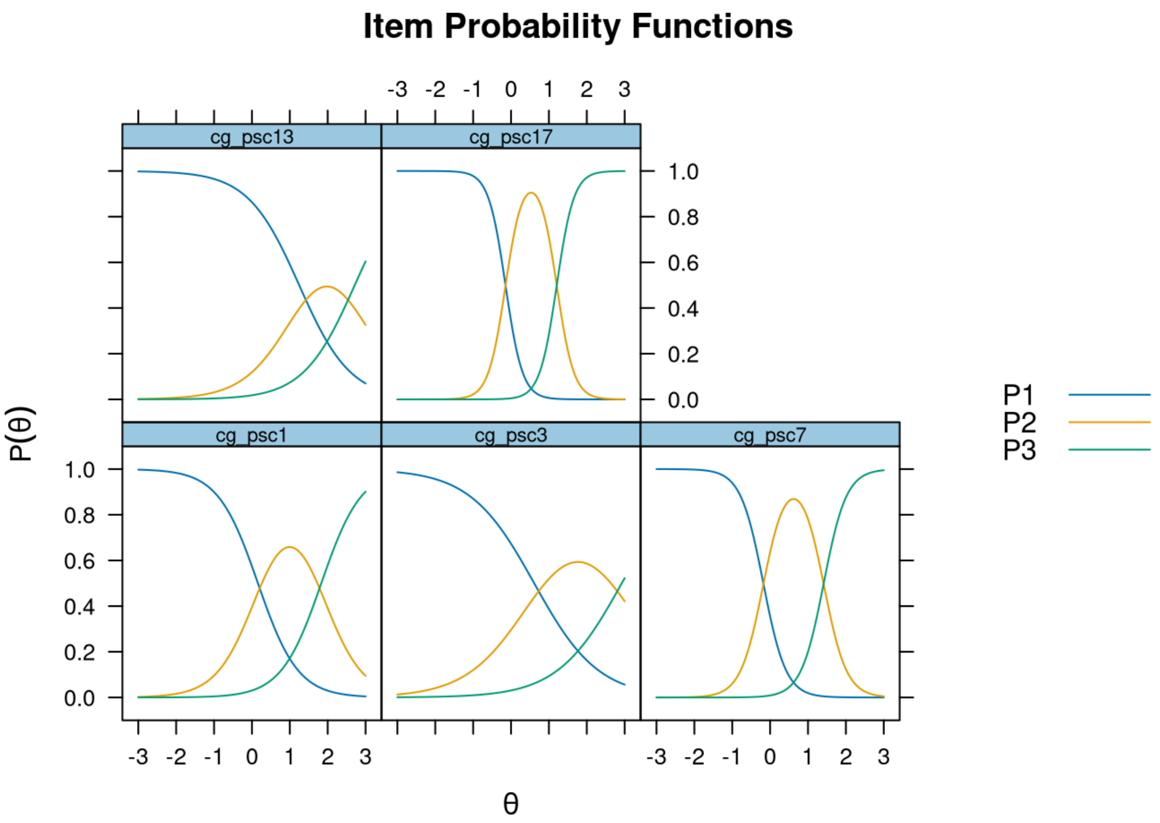

**Supplementary Figure 6.1.3 -** Pediatric Symptom Checklist  Short Version (PSC-17), 6- to 18-year-olds, Caregiver-report (Attention Scale): item infit and outfit statistics

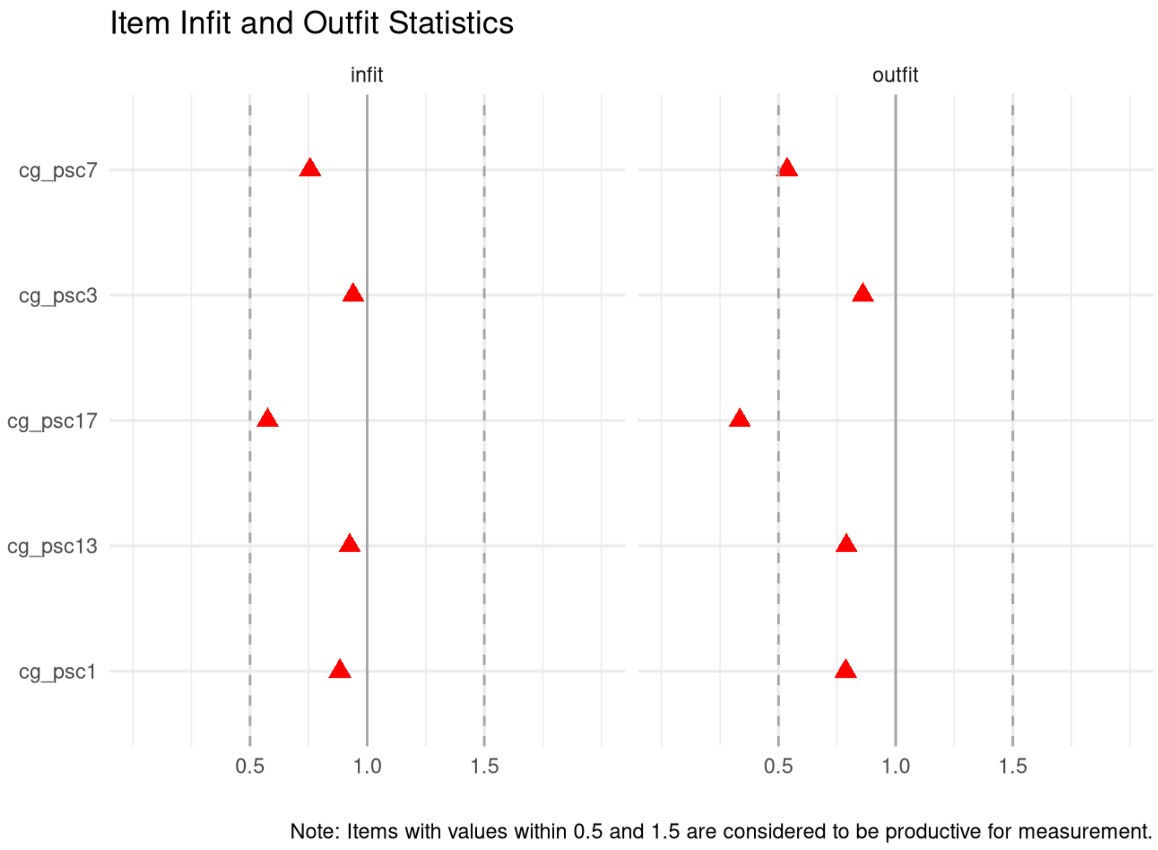

**Supplementary Figure 6.1.4 -** Pediatric Symptom Checklist Short Version (PSC-17), 6- to 18-year-olds, Caregiver-report (Attention Scale): person infit and outfit statistics

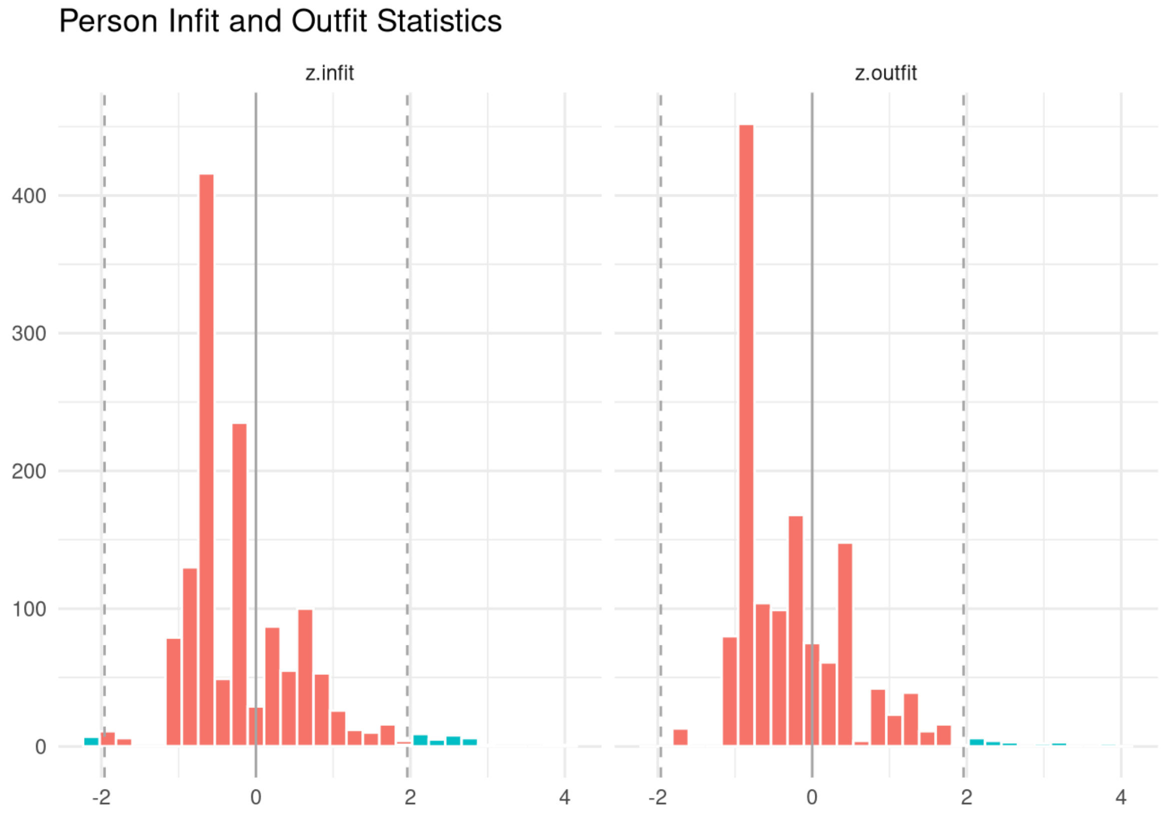

**Supplementary Figure 6.2.1 -** Pediatric Symptom Checklist Short Version (PSC-17), 6- to 18-year-olds, Caregiver-report (Externalizing Scale): test information and expected scores

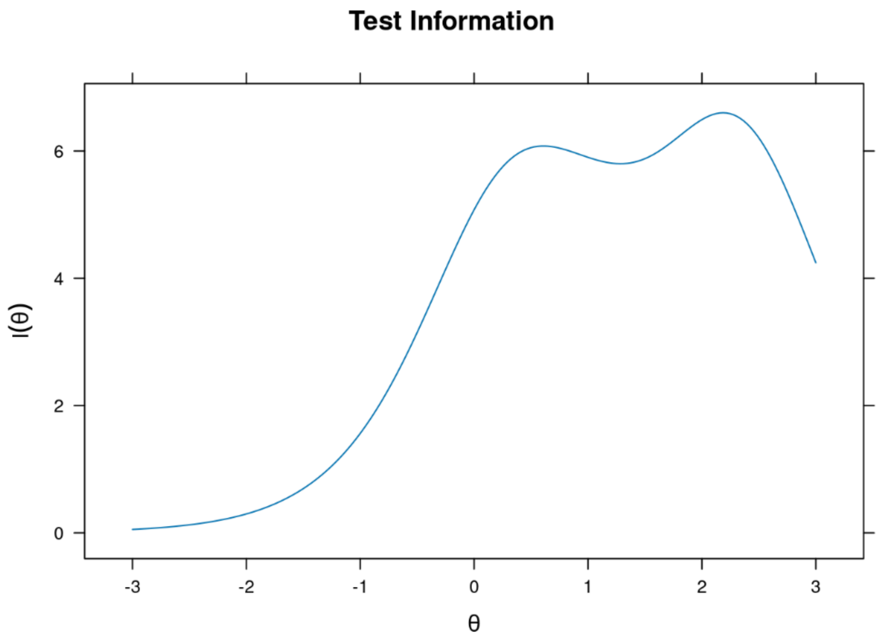

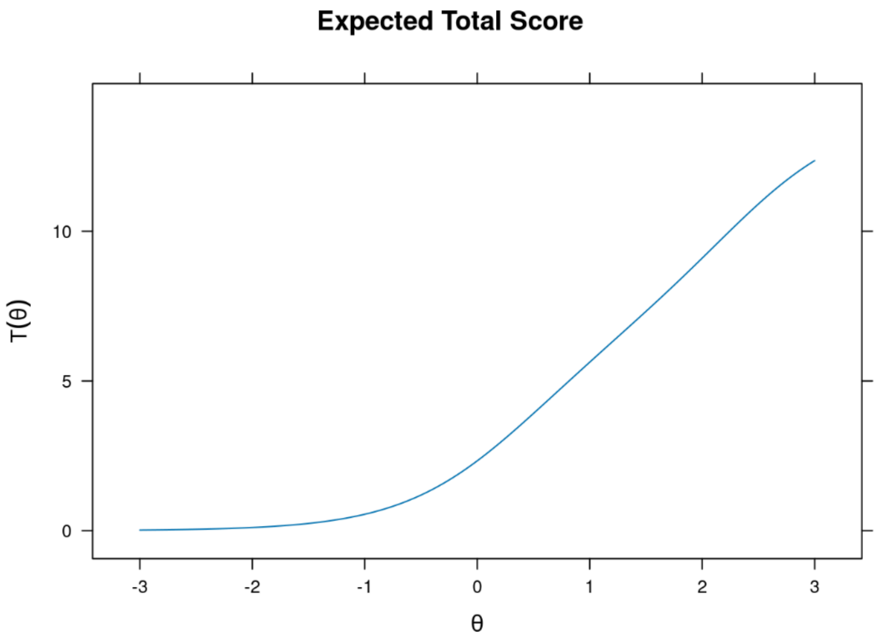

**Supplementary Figure 6.2.2 -** Pediatric Symptom Checklist Short Version (PSC-17), 6- to 18-year-olds, Caregiver-report (Externalizing Scale): item probability functions

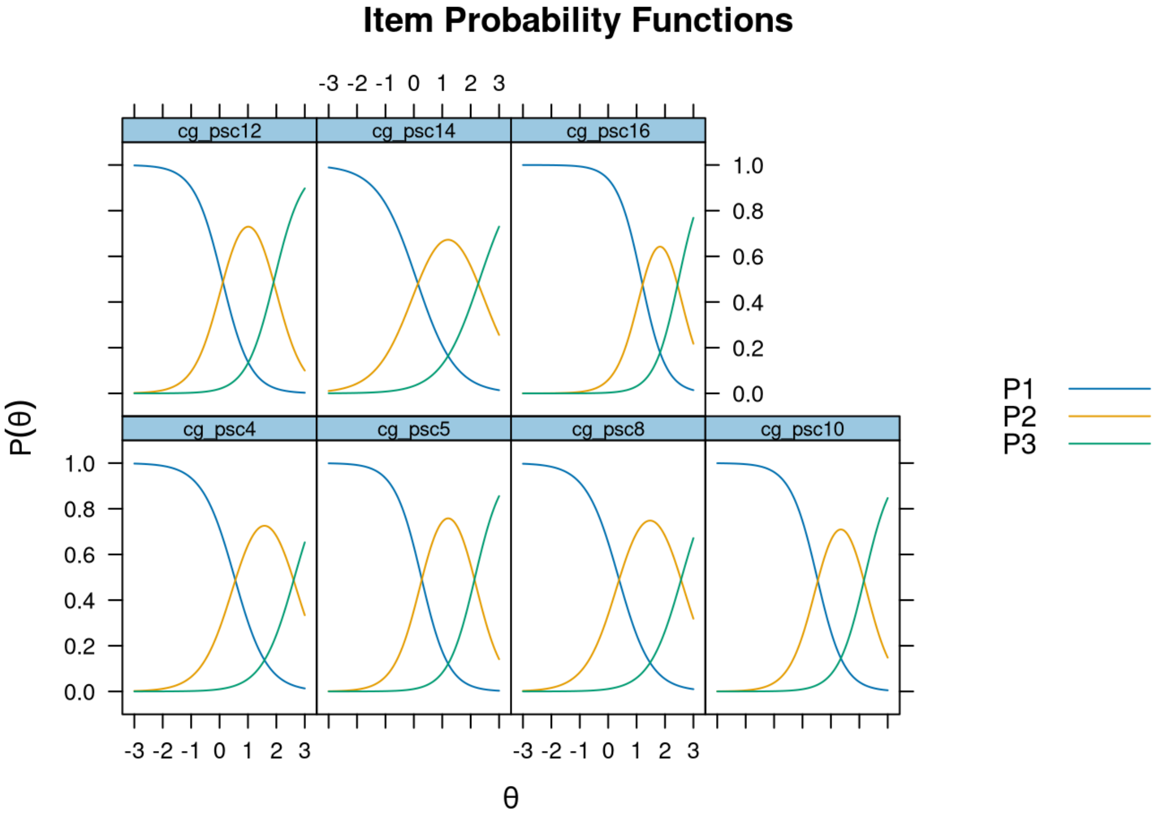

**Supplementary Figure 6.2.3 -** Pediatric Symptom Checklist Short Version (PSC-17), 6- to 18-year-olds, Caregiver-report (Externalizing Scale): item infit and outfit statistics

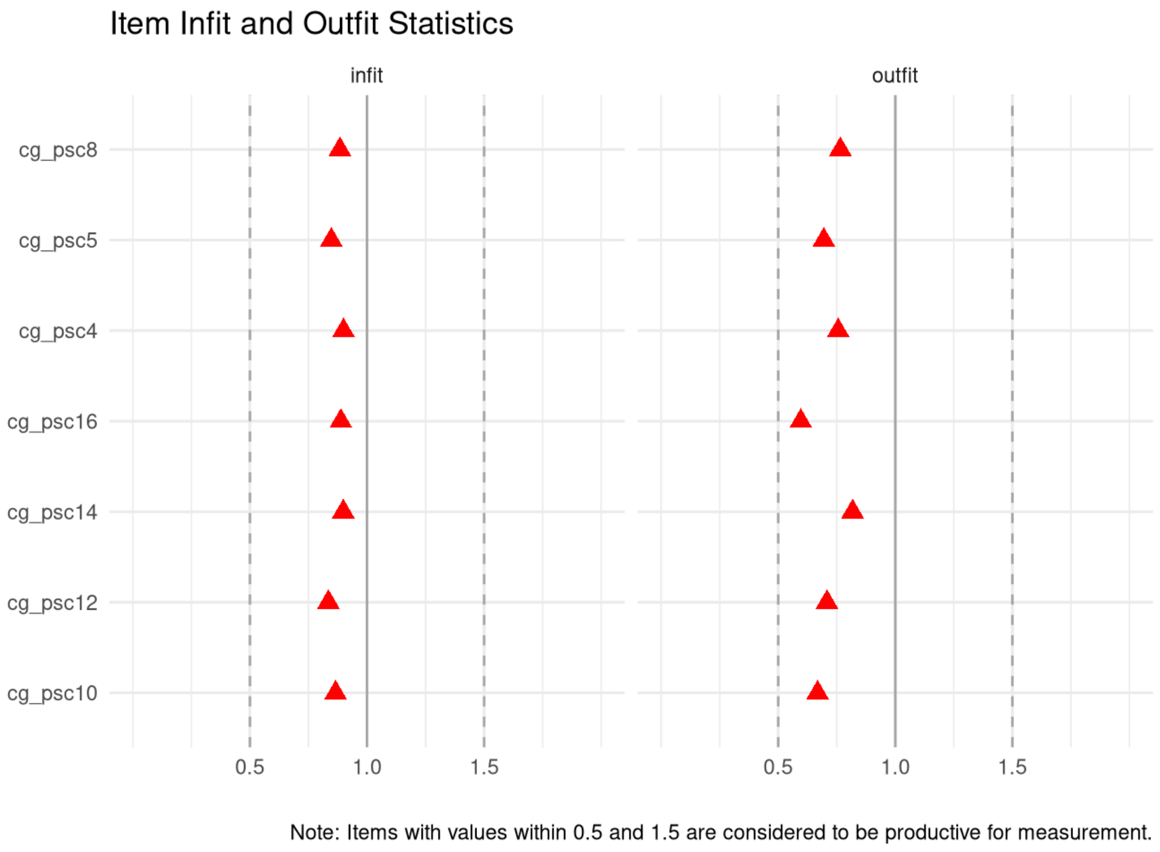

**Supplementary Figure 6.2.4 -** Pediatric Symptom Checklist Short Version (PSC-17), 6- to 18-year-olds, Caregiver-report (Externalizing Scale): person infit and outfit statistics

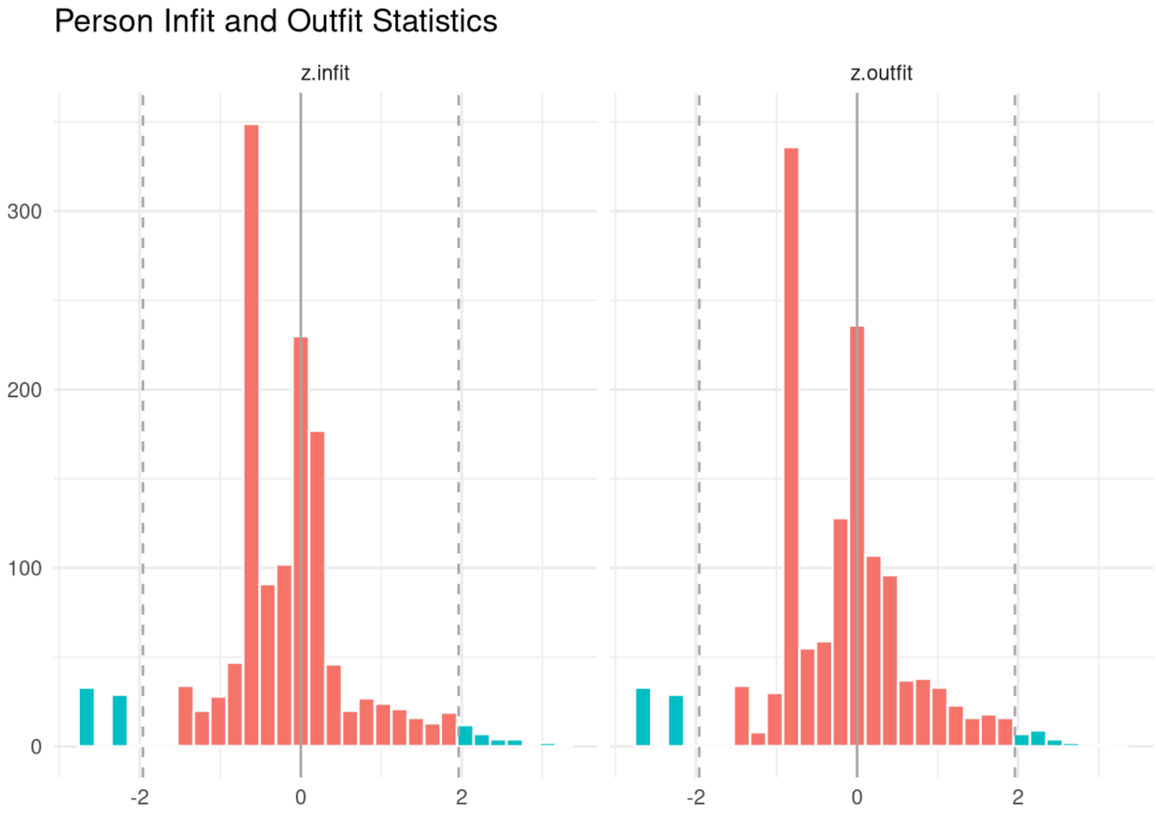

**Supplementary Figure 6.3.1 -** Pediatric Symptom Checklist  Short Version (PSC-17), 6- to 18-year-olds, Caregiver-report (Internalizing Scale): test information and expected scores

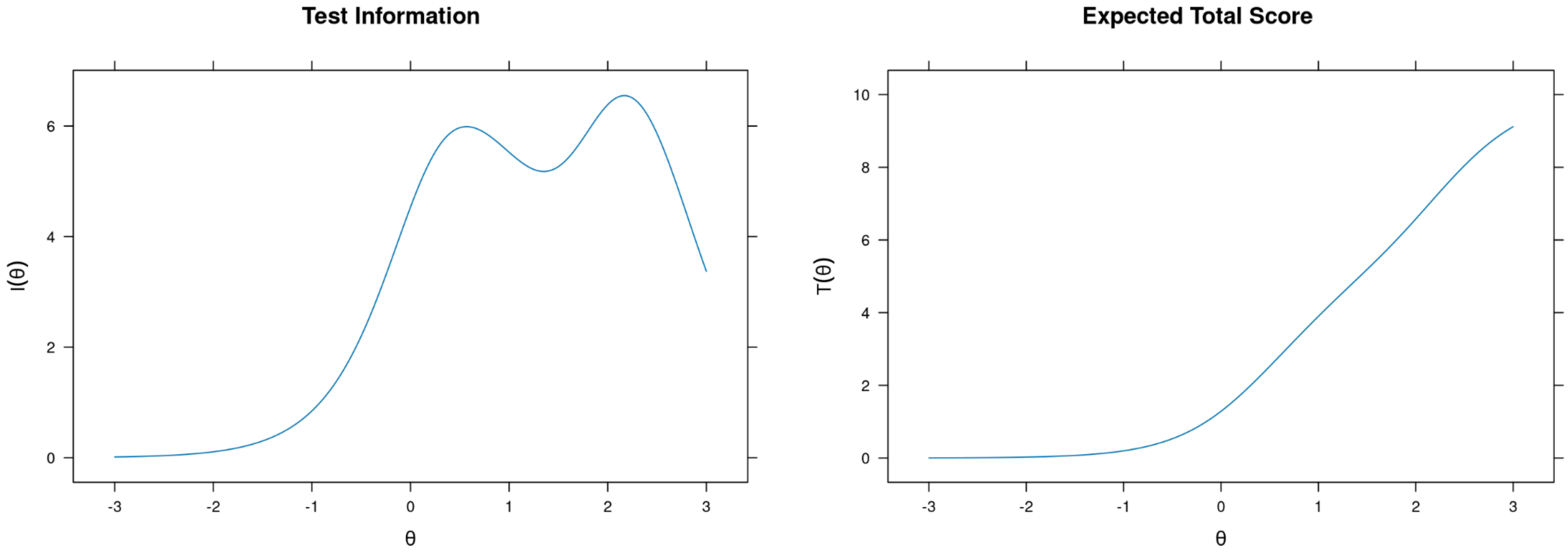

**Supplementary Figure 6.3.2 -** Pediatric Symptom Checklist Short Version (PSC-17), 6- to 18-year-olds, Caregiver-report (Internalizing Scale): item probability functions

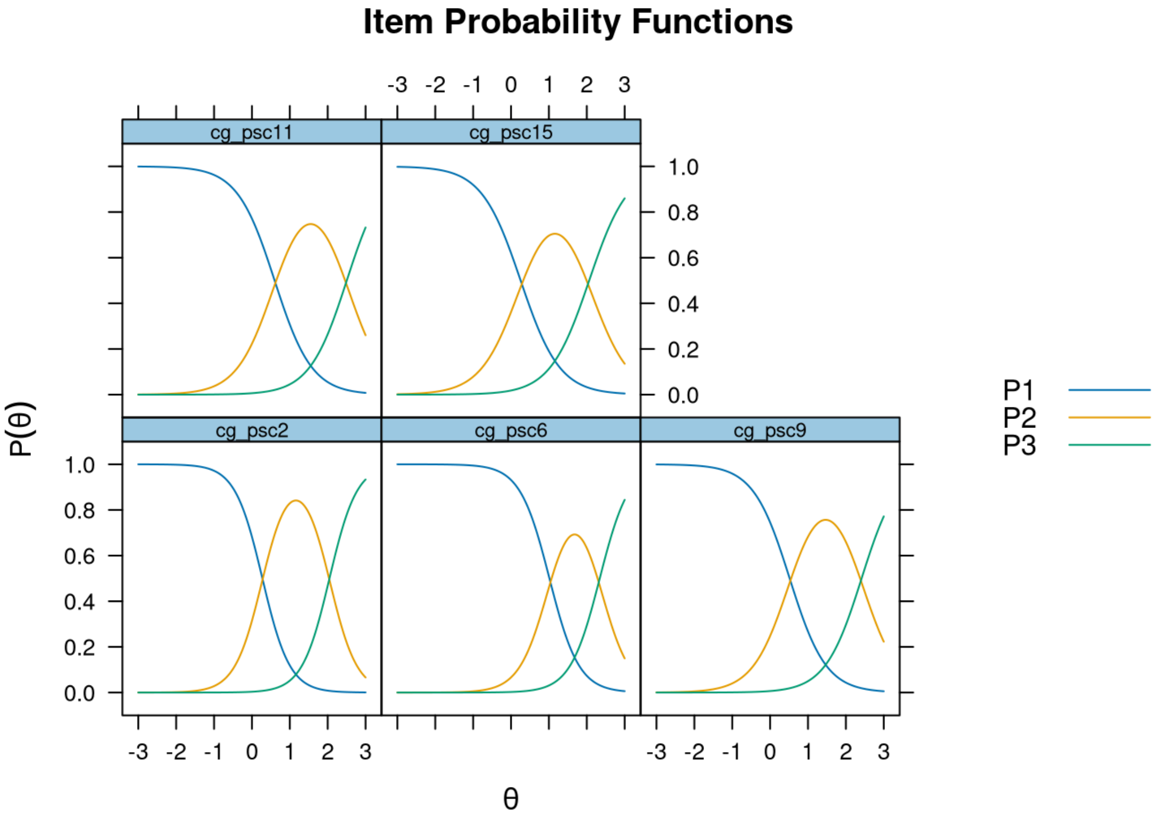

**Supplementary Figure 6.3.3 -** Pediatric Symptom Checklist  Short Version (PSC-17), 6- to 18-year-olds, Caregiver-report (Internalizing Scale): item infit and outfit statistics

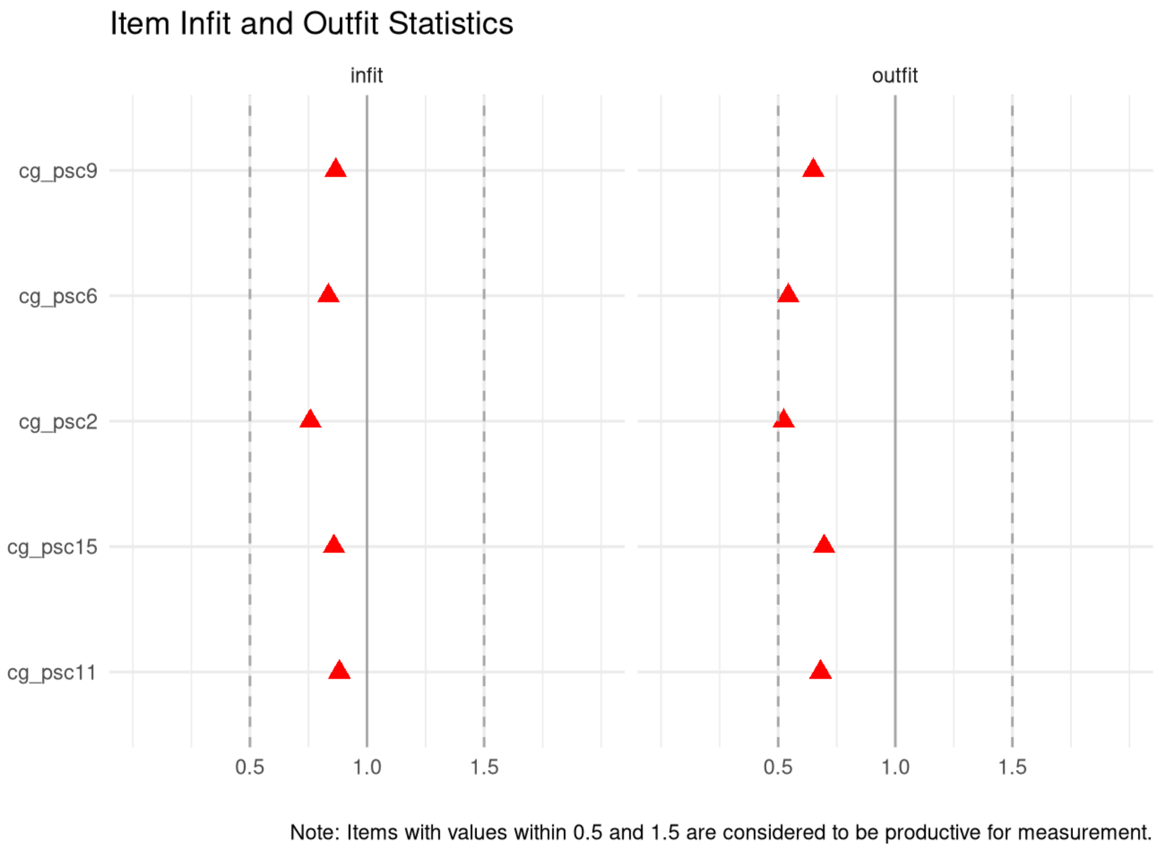

**Supplementary Figure 6.3.4 -** Pediatric Symptom Checklist  Short Version (PSC-17), 6- to 18-year-olds, Self-report (Internalizing Scale): person infit and outfit statistics

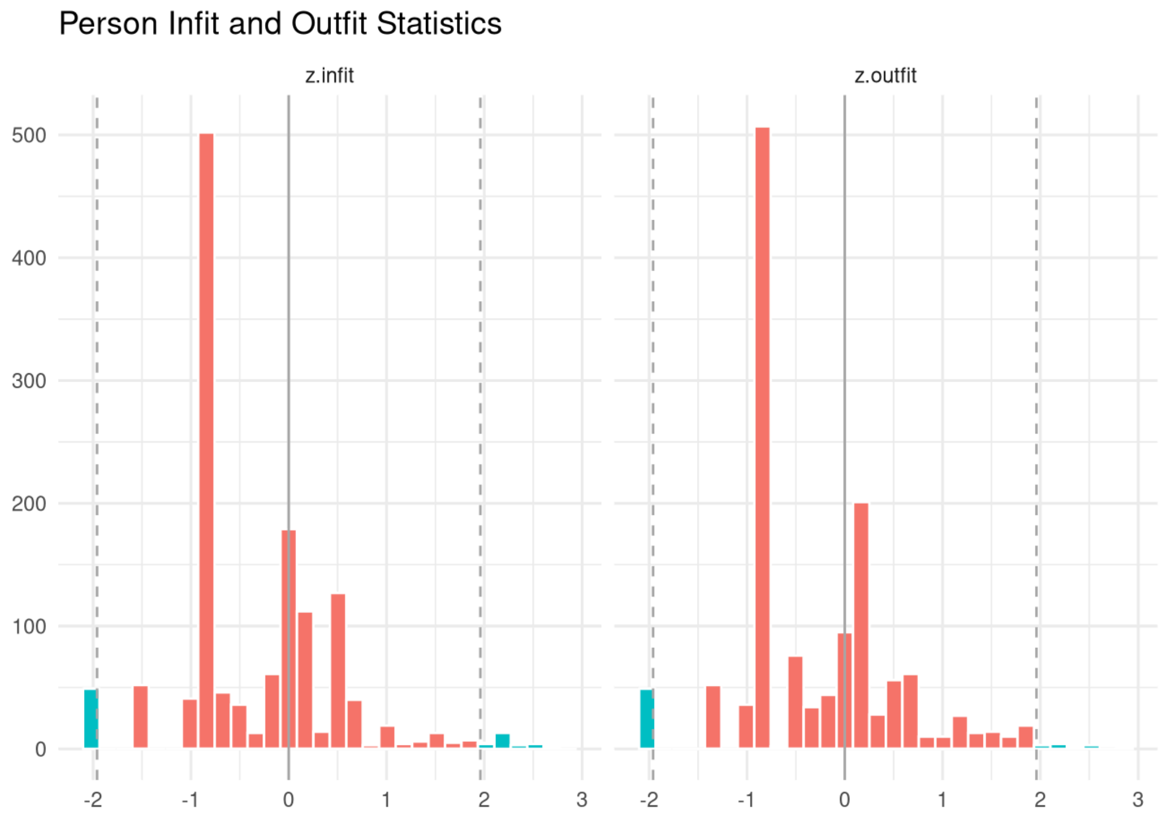

**Supplementary Figure 7.1.1 -** Pediatric Symptom Checklist Short Version (PSC-17), 6- to 18-year-olds, Self-report (Attention Scale): test information and expected scores

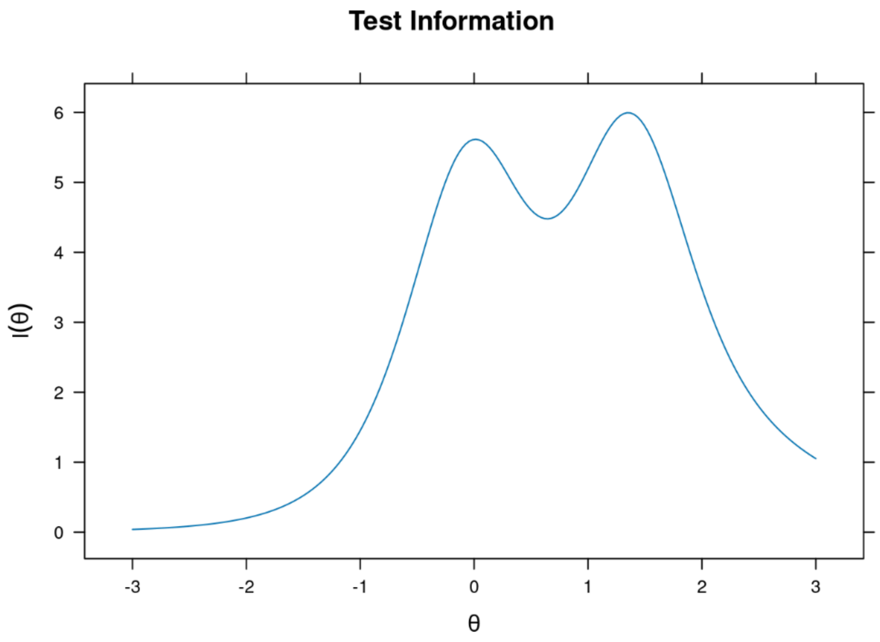

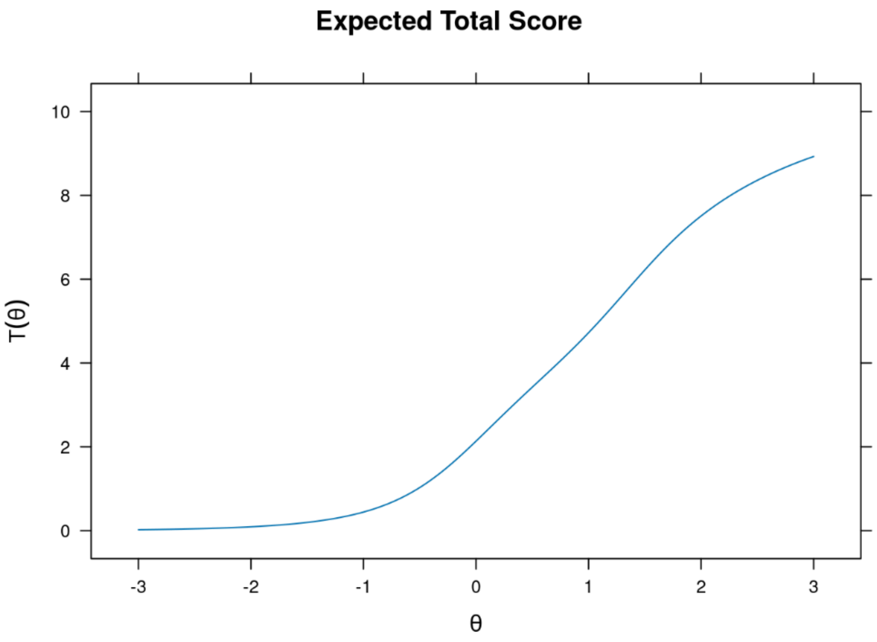

**Supplementary Figure 7.1.2 -** Pediatric Symptom Checklist Short Version (PSC-17), 6- to 18-year-olds, Self-report (Attention Scale): item probability functions

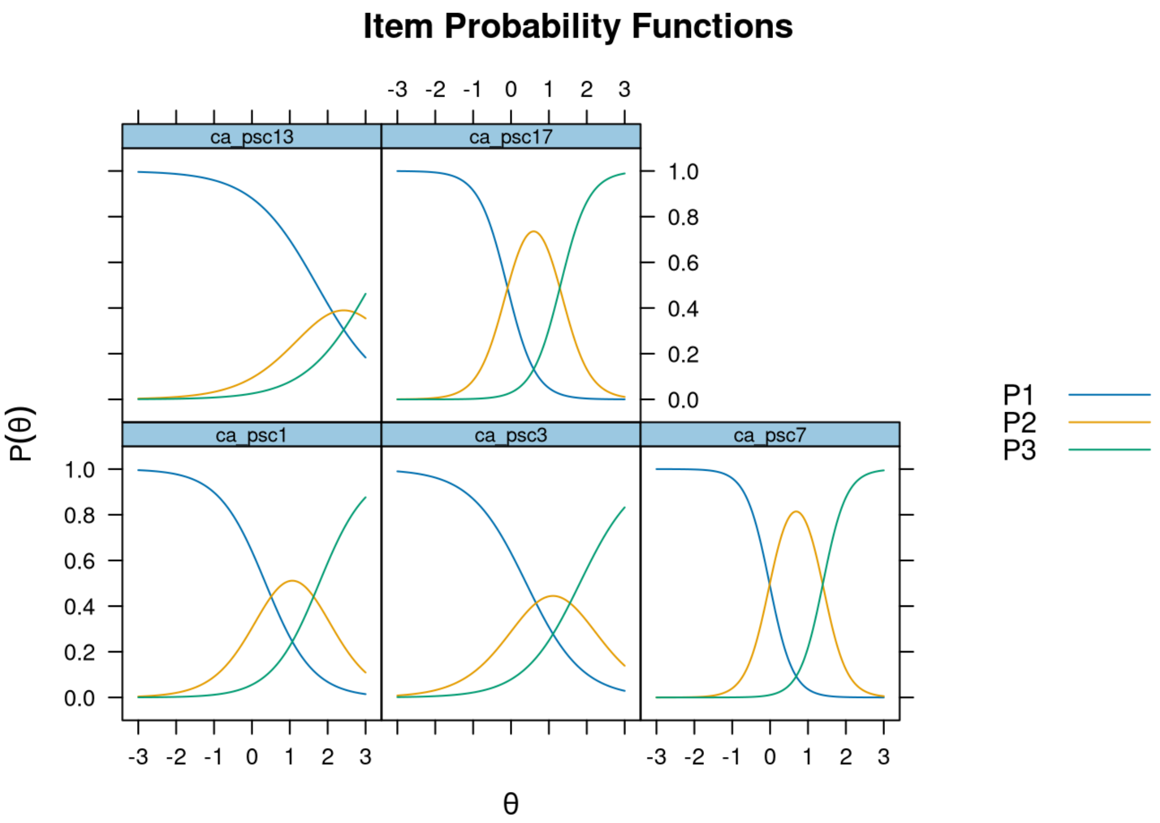

**Supplementary Figure 7.1.3 -** Pediatric Symptom Checklist  Short Version (PSC-17), 6- to 18-year-olds, Self-report (Attention Scale): item infit and outfit statistics

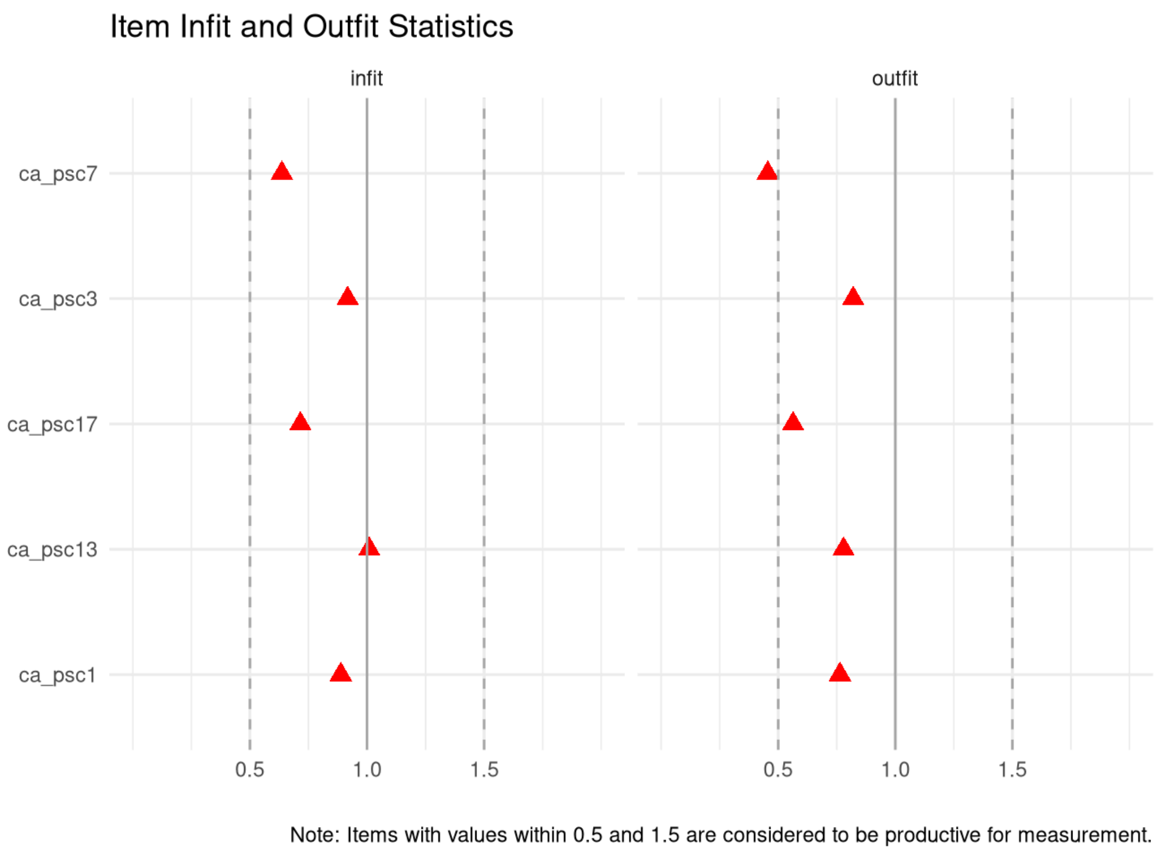

**Supplementary Figure 7.1.4 -** Pediatric Symptom Checklist  Short Version (PSC-17), 6- to 18-year-olds, Self-report (Attention Scale): person infit and outfit statistics

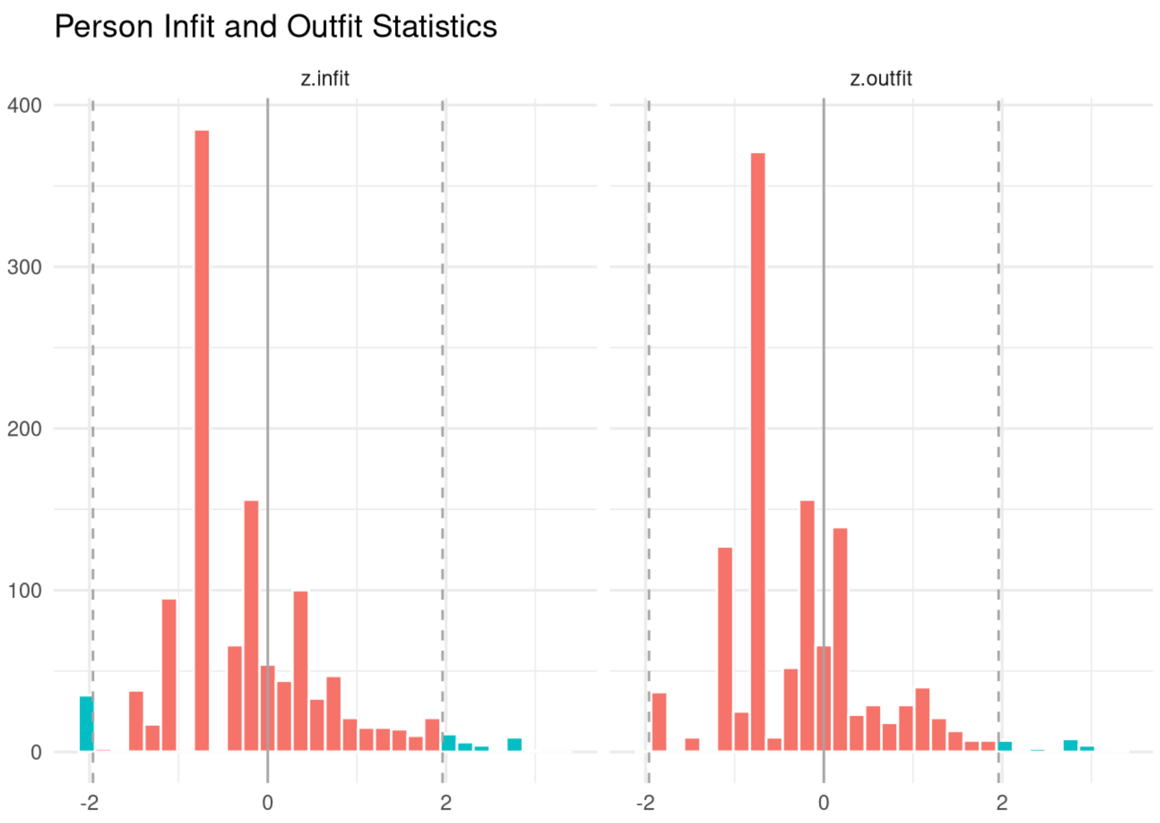

**Supplementary Figure 7.2.1 -** Pediatric Symptom Checklist Short Version (PSC-17), 6- to 18-year-olds, Self-report (Externalizing Scale): test information and expected scores

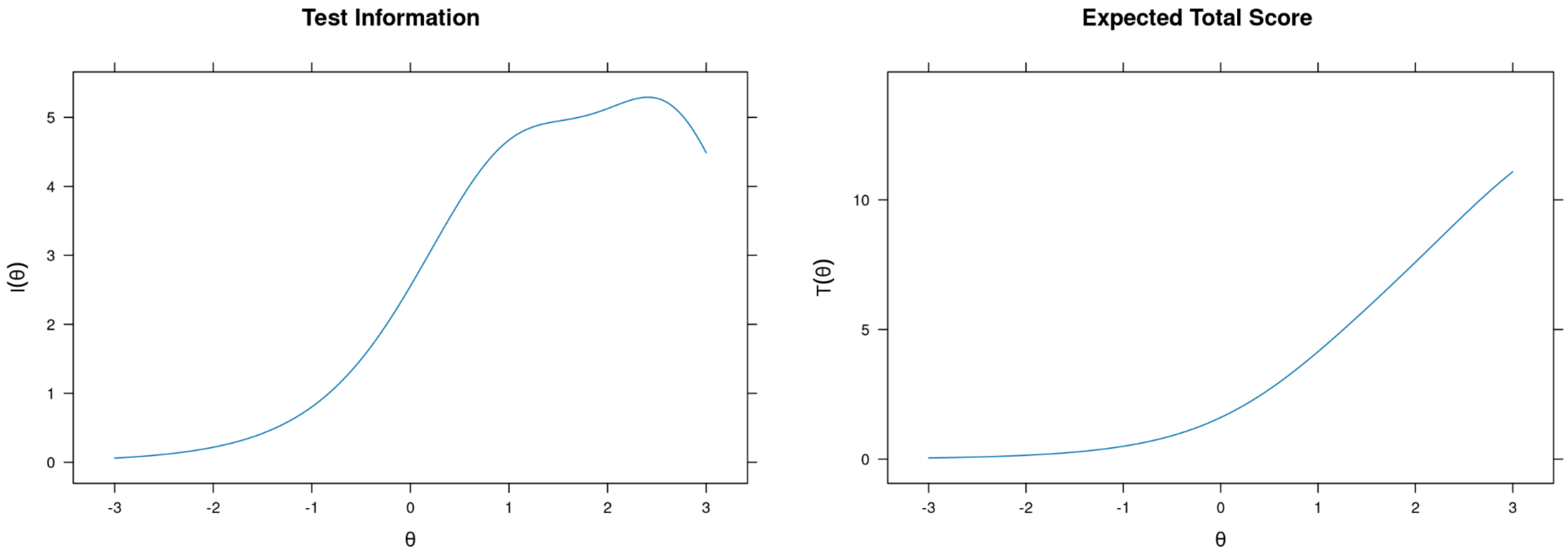

**Supplementary Figure 7.2.2 -** Pediatric Symptom Checklist Short Version (PSC-17), 6- to 18-year-olds, Self-report (Externalizing Scale): item probability functions

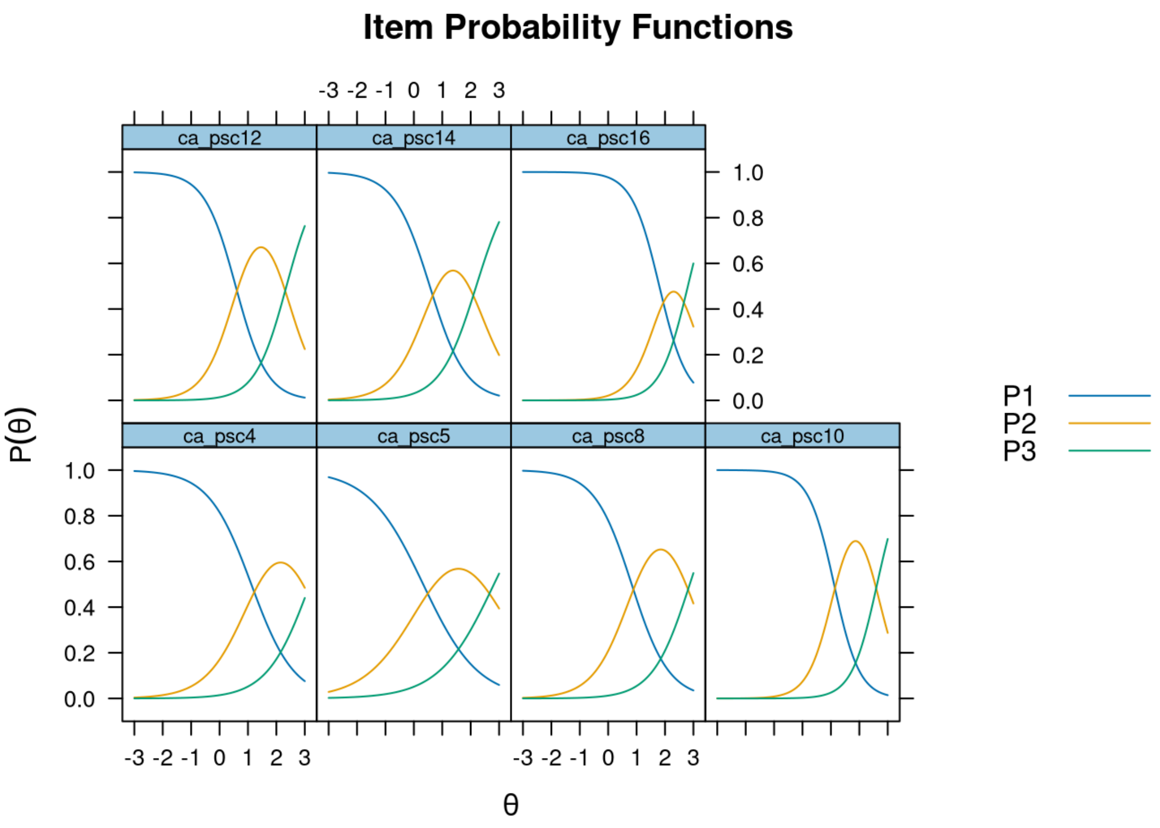

**Supplementary Figure 7.2.3 -** Pediatric Symptom Checklist  Short Version (PSC-17), 6- to 18-year-olds, Self-report (Externalizing Scale): item infit and outfit statistics

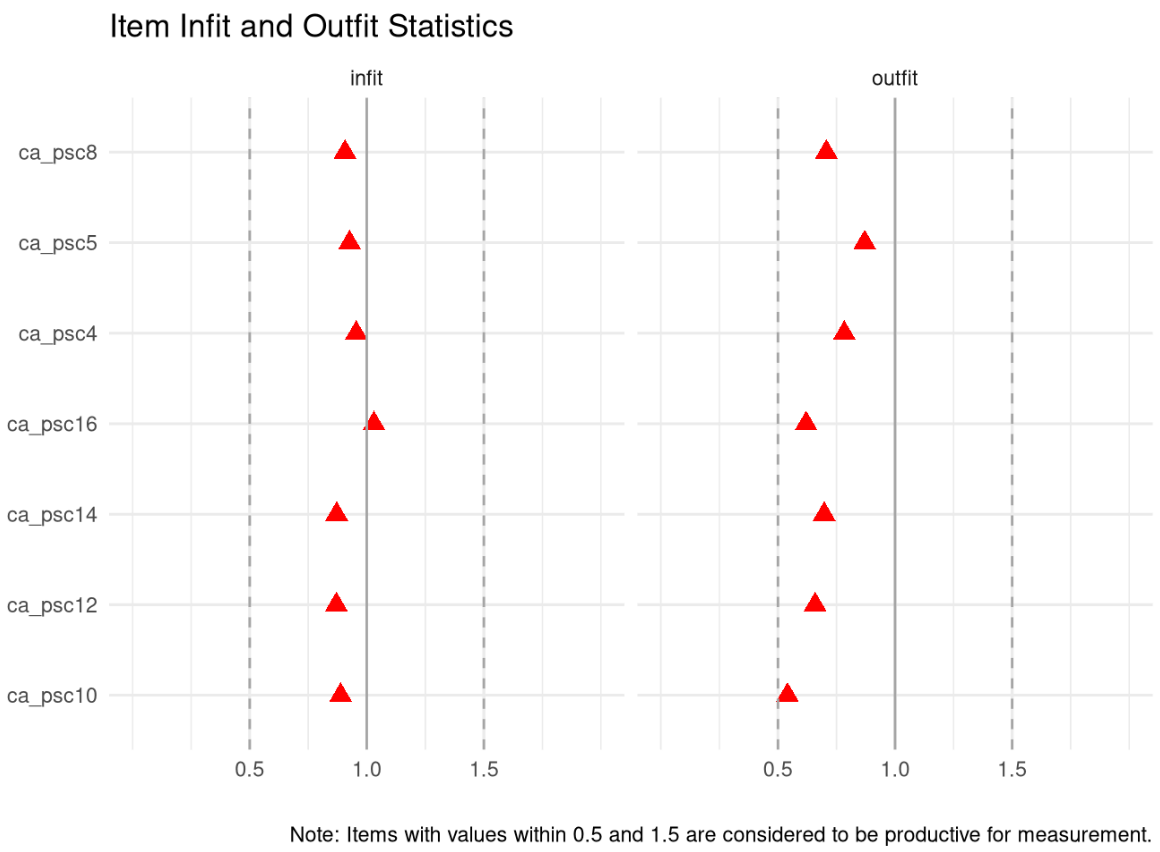

**Supplementary Figure 7.2.4 -** Pediatric Symptom Checklist Short Version (PSC-17), 6- to 18-year-olds, Self-report (Externalizing Scale): person infit and outfit statistics

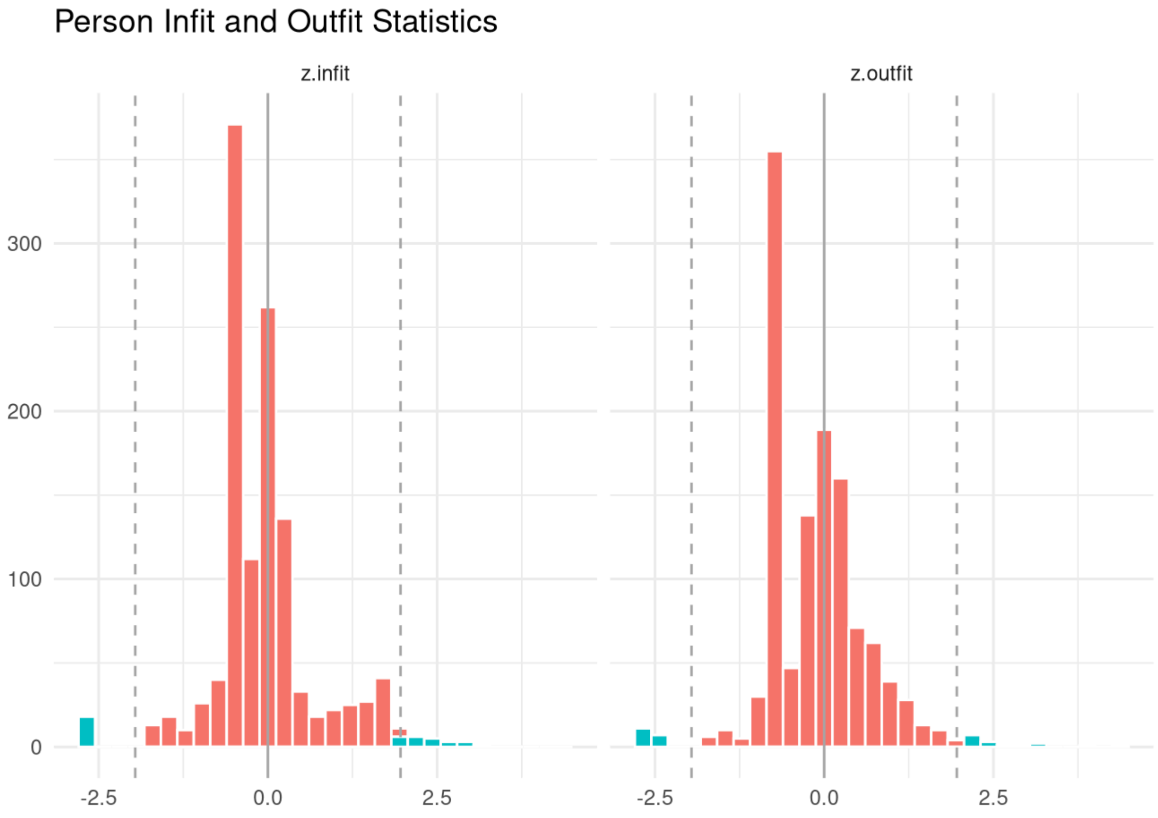

**Supplementary Figure 7.3.1 -** Pediatric Symptom Checklist  Short Version (PSC-17), 6- to 18-year-olds, Self-report (Internalizing Scale): test information and expected scores

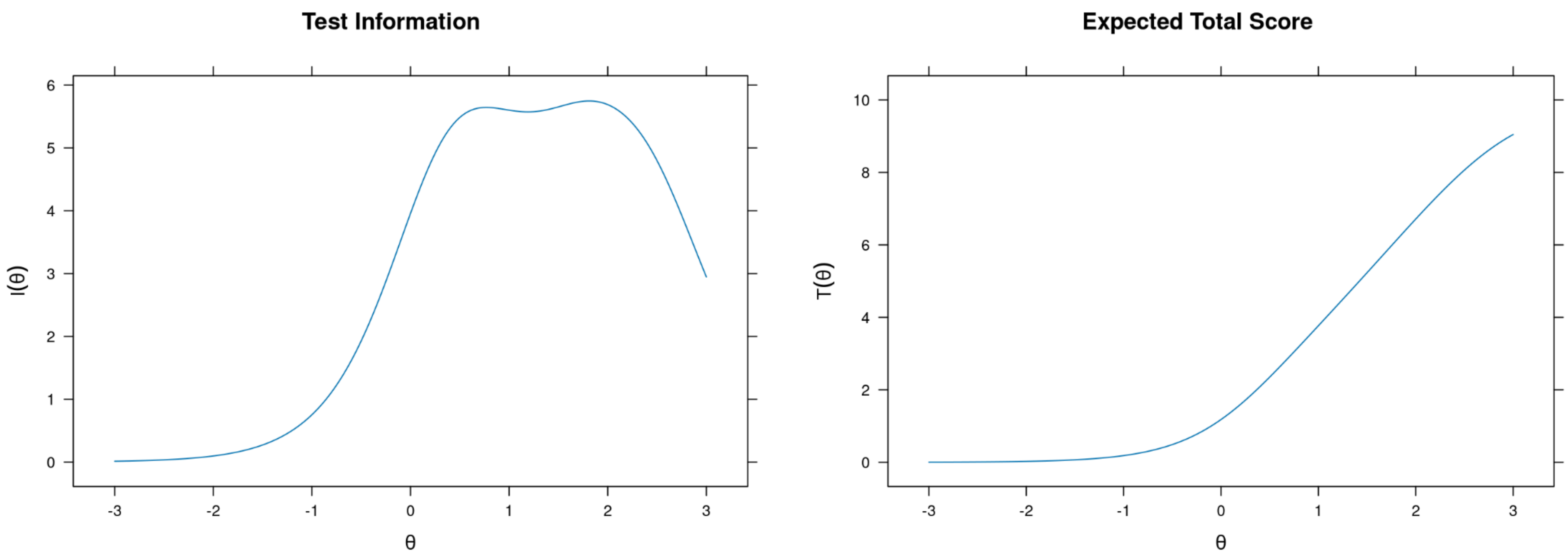

**Supplementary Figure 7.3.2 -** Pediatric Symptom Checklist  Short Version (PSC-17), 6- to 18-year-olds, Self-report (Internalizing Scale): item probability functions

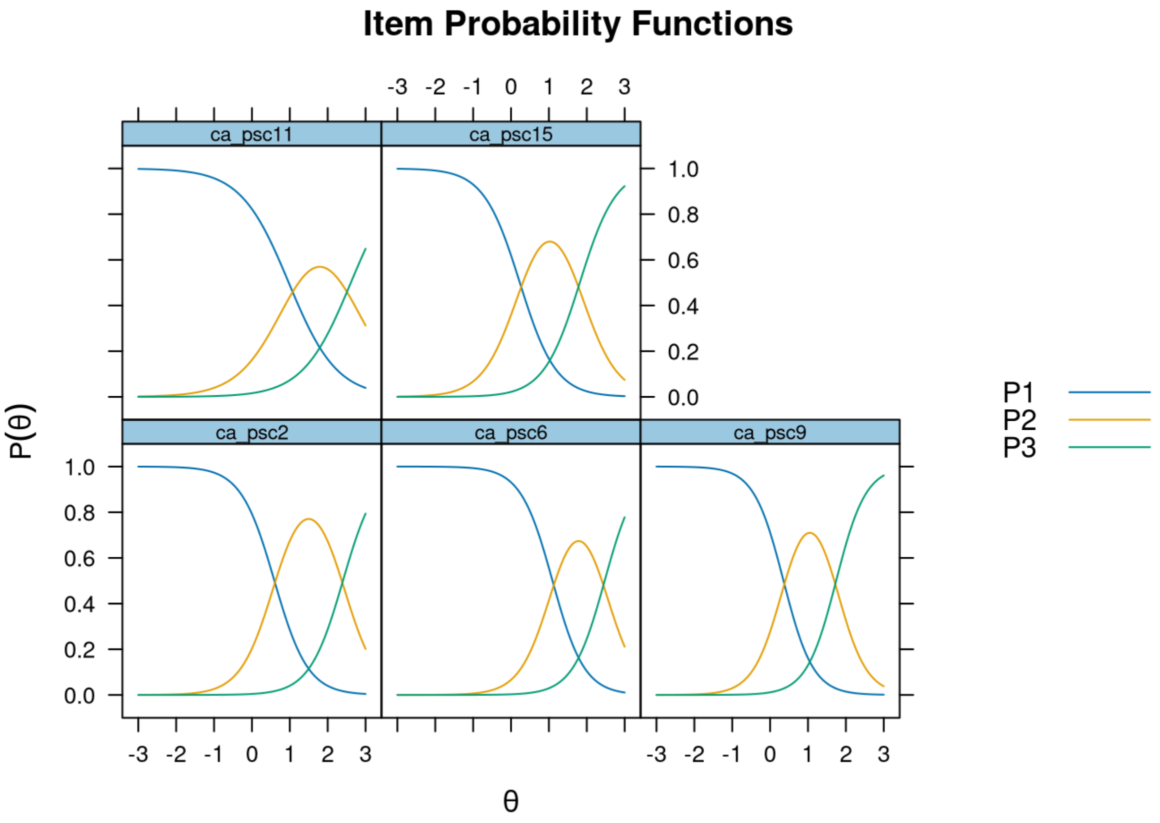

**Supplementary Figure 7.3.3 -** Pediatric Symptom Checklist Short Version (PSC-17), 6- to 18-year-olds, Self-report (Internalizing Scale): item infit and outfit statistics

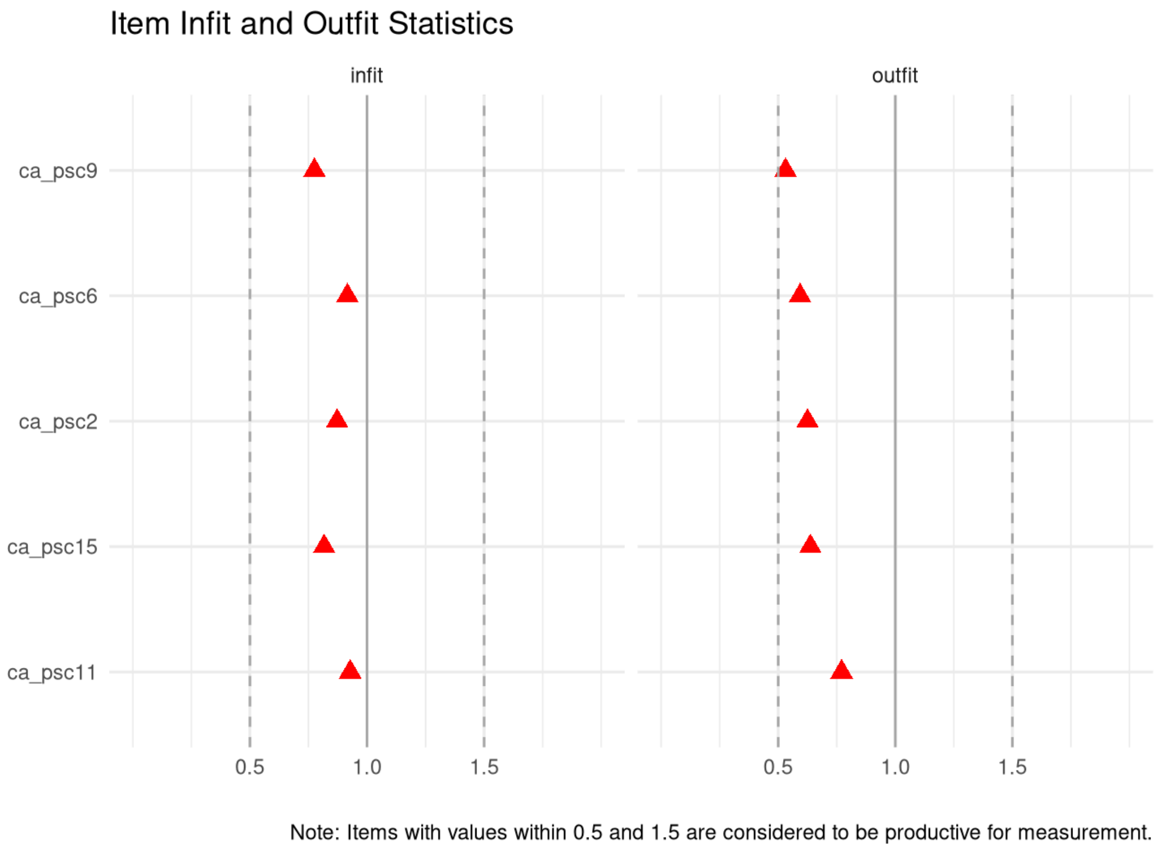

**Supplementary Figure 7.3.4 -** Pediatric Symptom Checklist  Short Version (PSC-17), 6- to 18-year-olds, Self-report (Internalizing Scale): person infit and outfit statistics

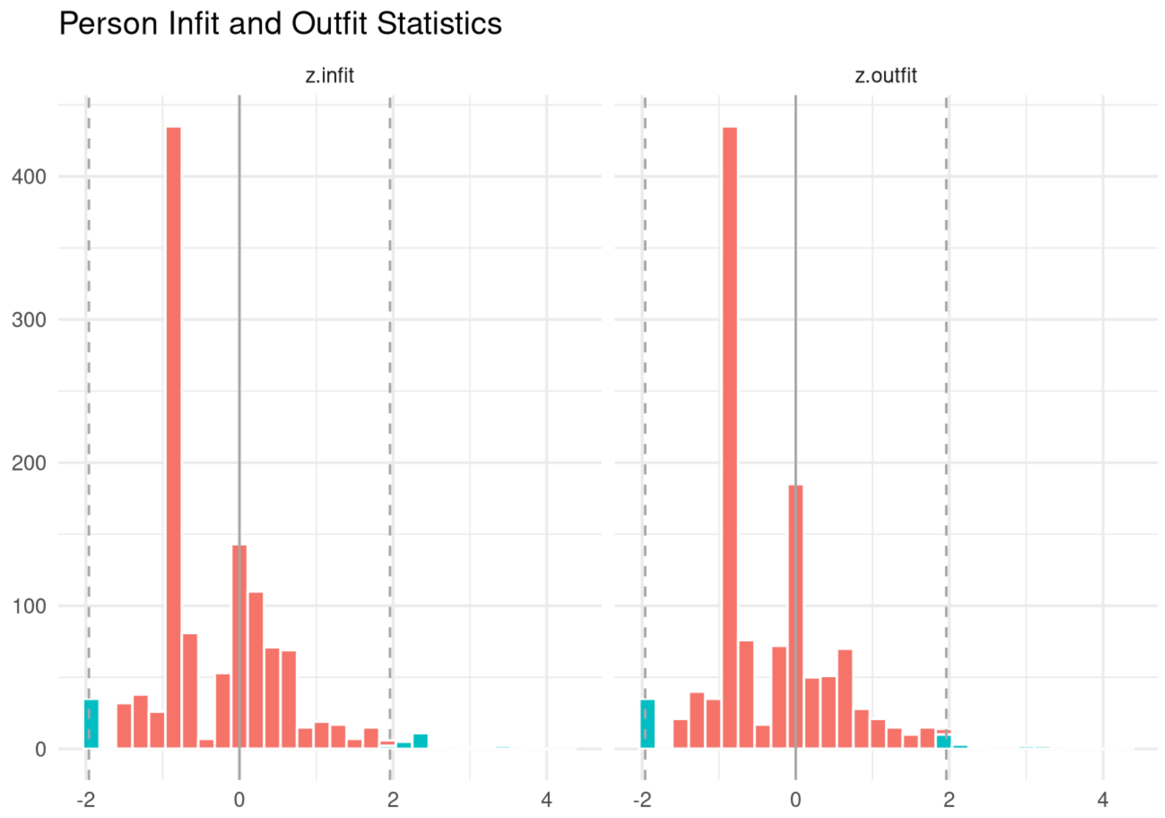

**Supplementary Figure 8.1.1 -** Revised Children's Anxiety and Depression Scale short-version (RCADS-25), Caregiver-report (Anxiety Scale): test information and expected scores

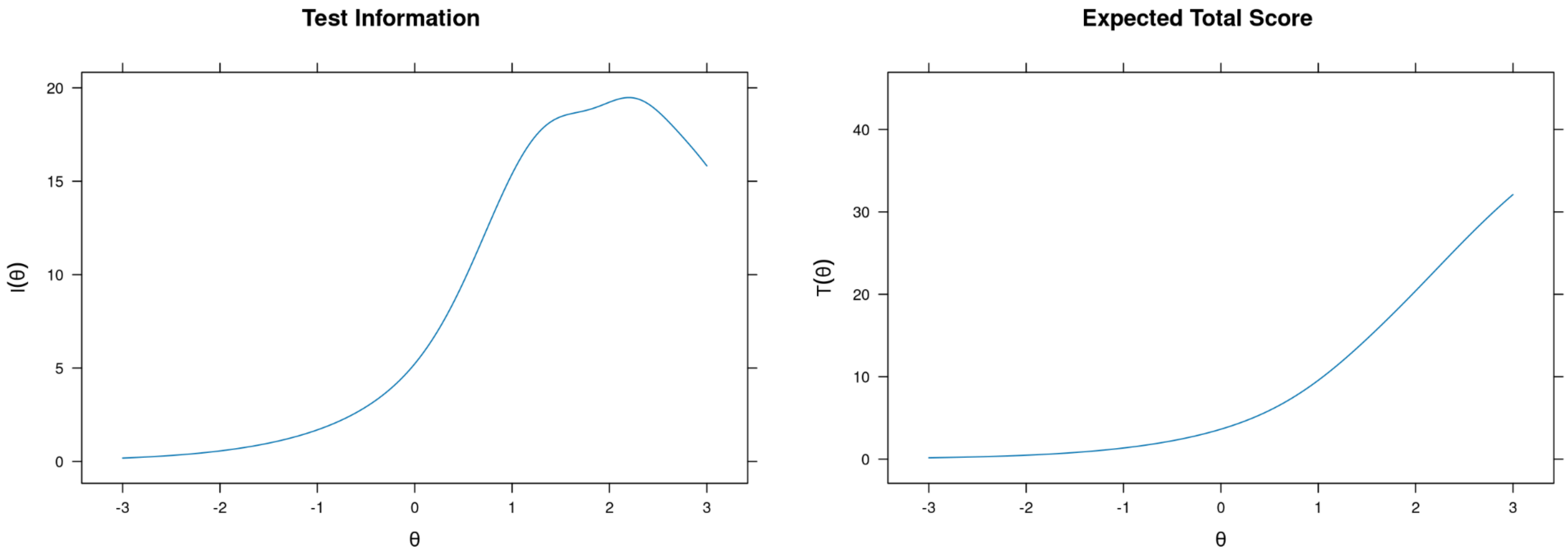

**Supplementary Figure 8.1.2 -** Revised Children's Anxiety and Depression Scale short-version (RCADS-25), Caregiver-report (Anxiety Scale): item probability functions

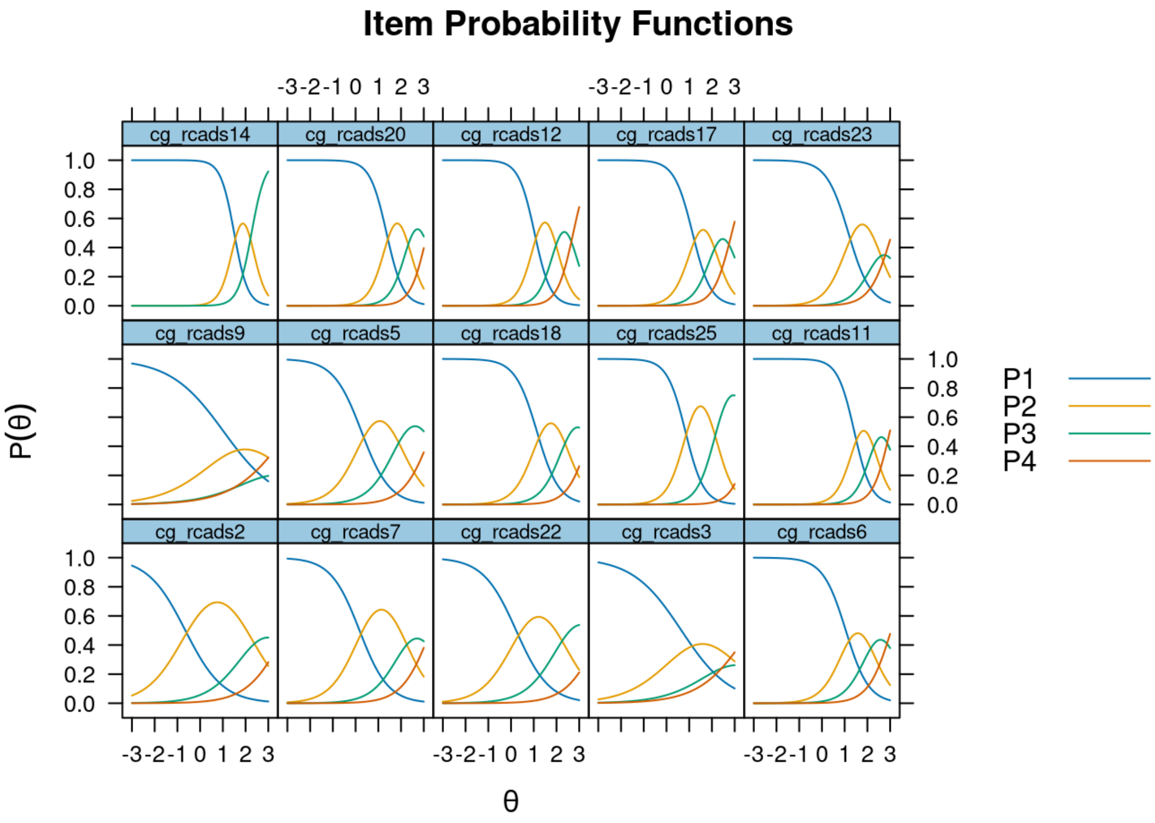

**Supplementary Figure 8.1.3 -** Revised Children's Anxiety and Depression Scale short-version (RCADS-25), Caregiver-report (Anxiety Scale): item infit and outfit statistics

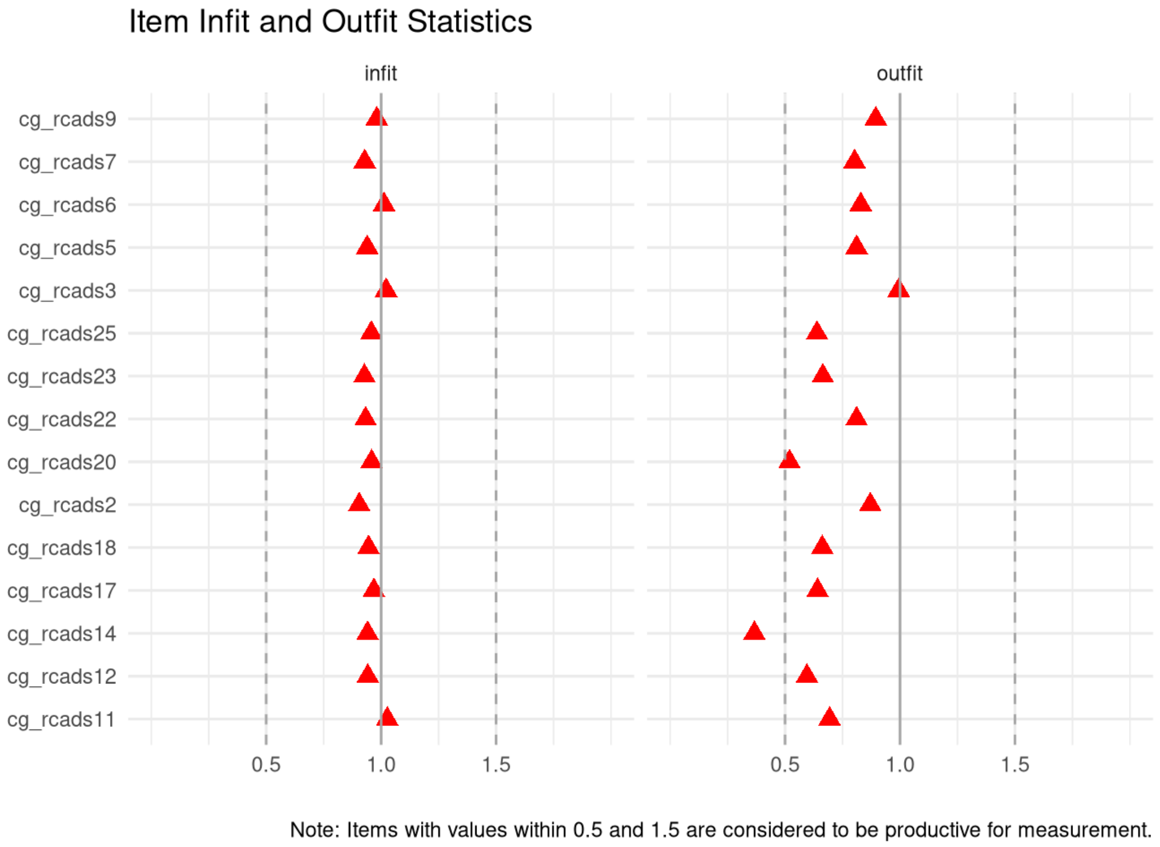

**Supplementary Figure 8.1.4 -** Revised Children's Anxiety and Depression Scale short-version (RCADS-25), Caregiver-report (Anxiety Scale): person infit and outfit statistics

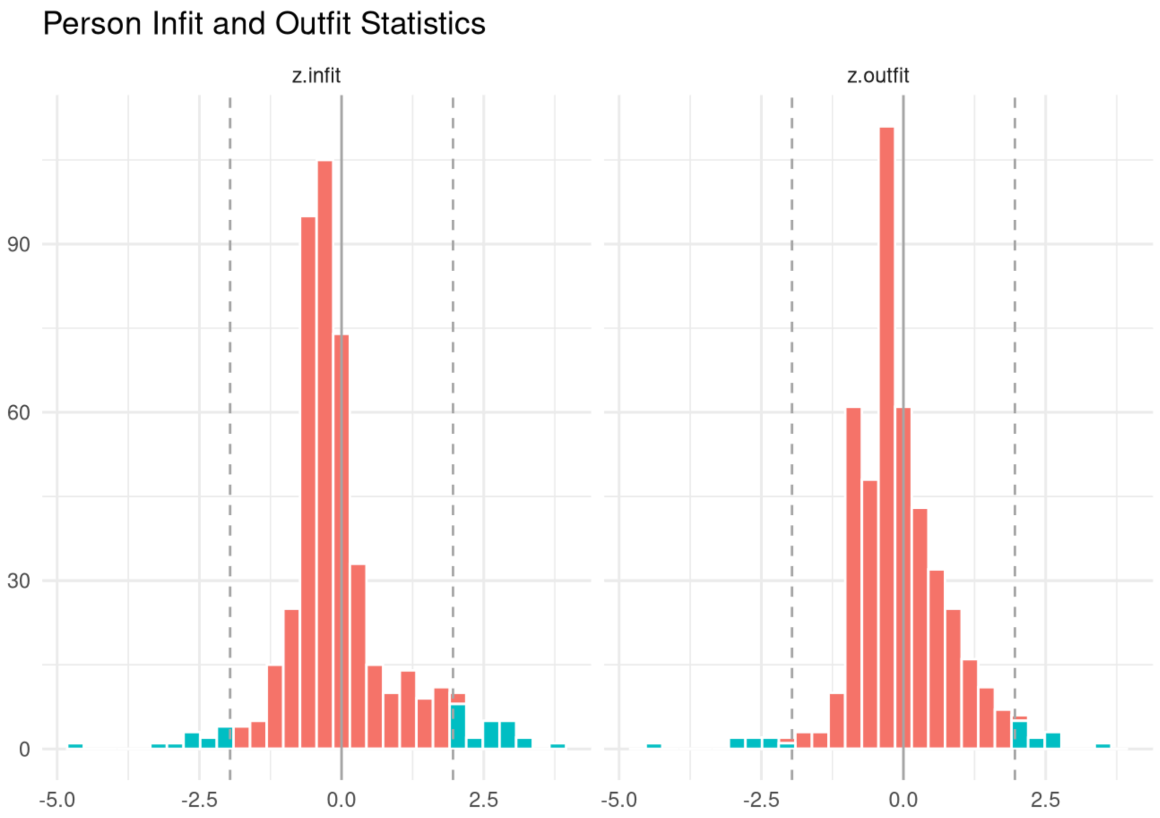

**Supplementary Figure 8.2.1 -** Revised Children's Anxiety and Depression Scale short-version (RCADS-25), Caregiver-report (Depression Scale): test information and expected scores

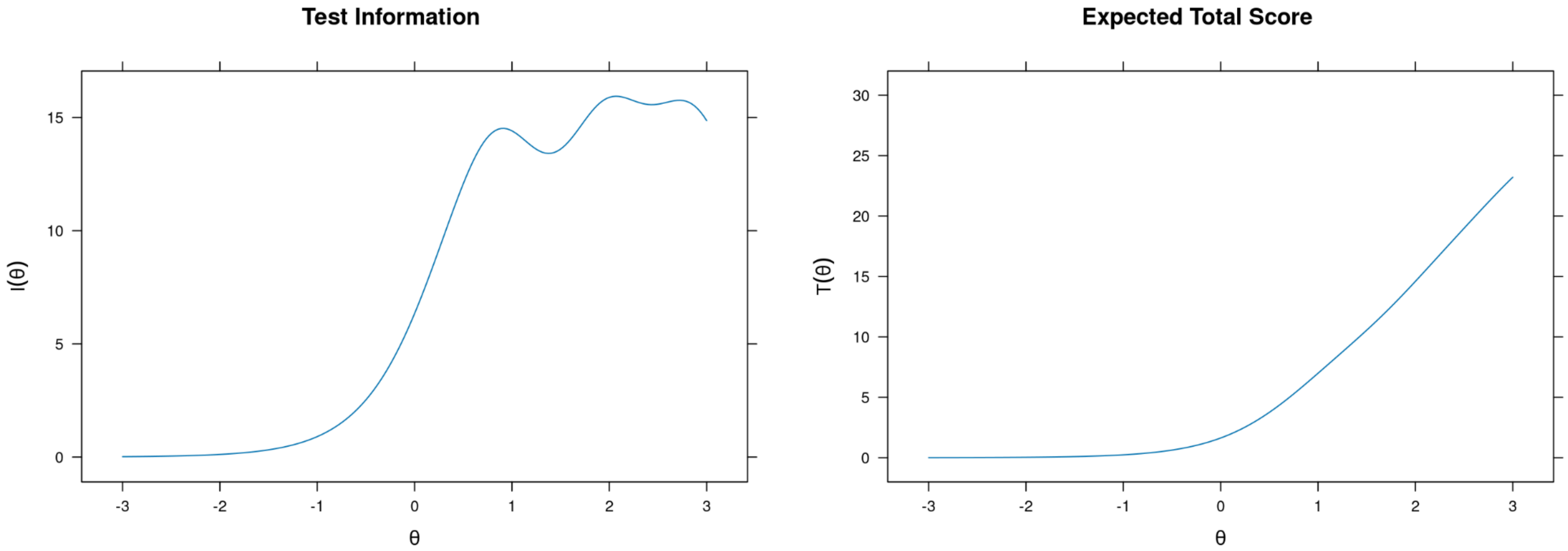

**Supplementary Figure 8.2.2 -** Revised Children's Anxiety and Depression Scale short-version (RCADS-25), Caregiver-report (Depression Scale): item probability functions

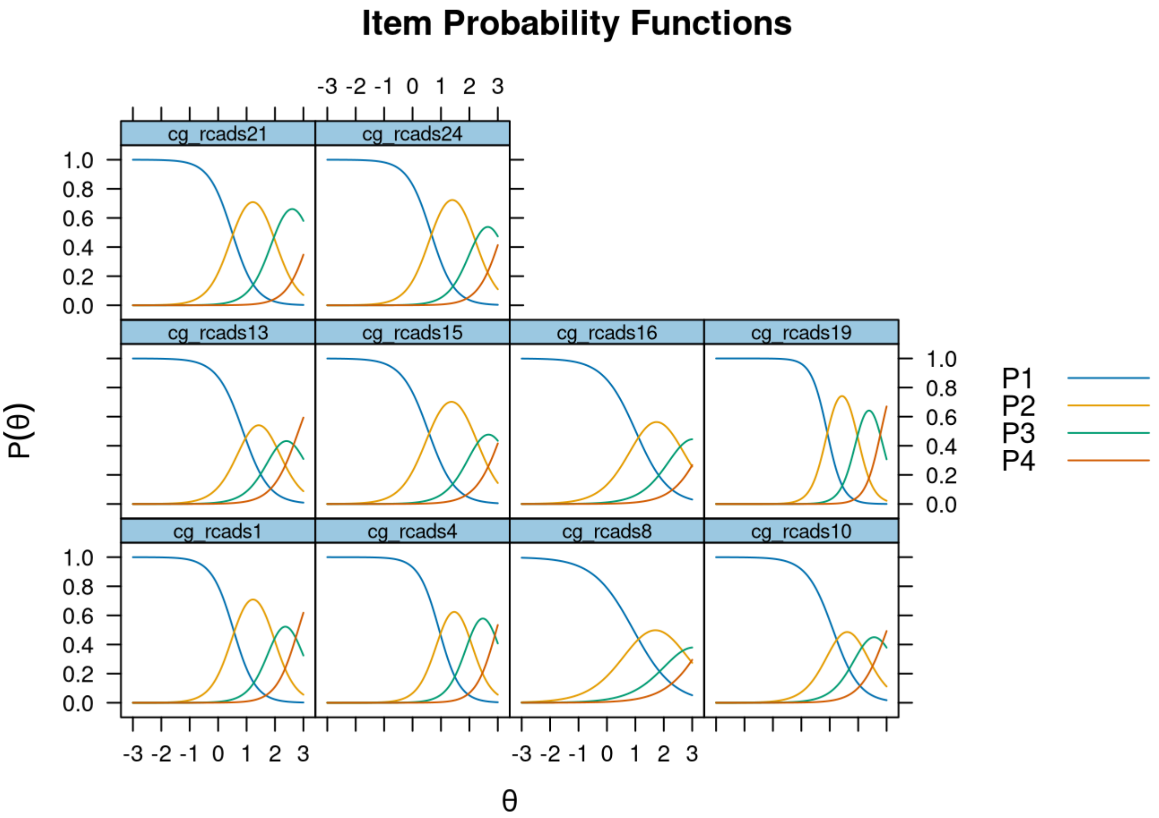

**Supplementary Figure 8.2.3 -** Revised Children's Anxiety and Depression Scale short-version (RCADS-25), Caregiver-report (Depression Scale): item infit and outfit statistics

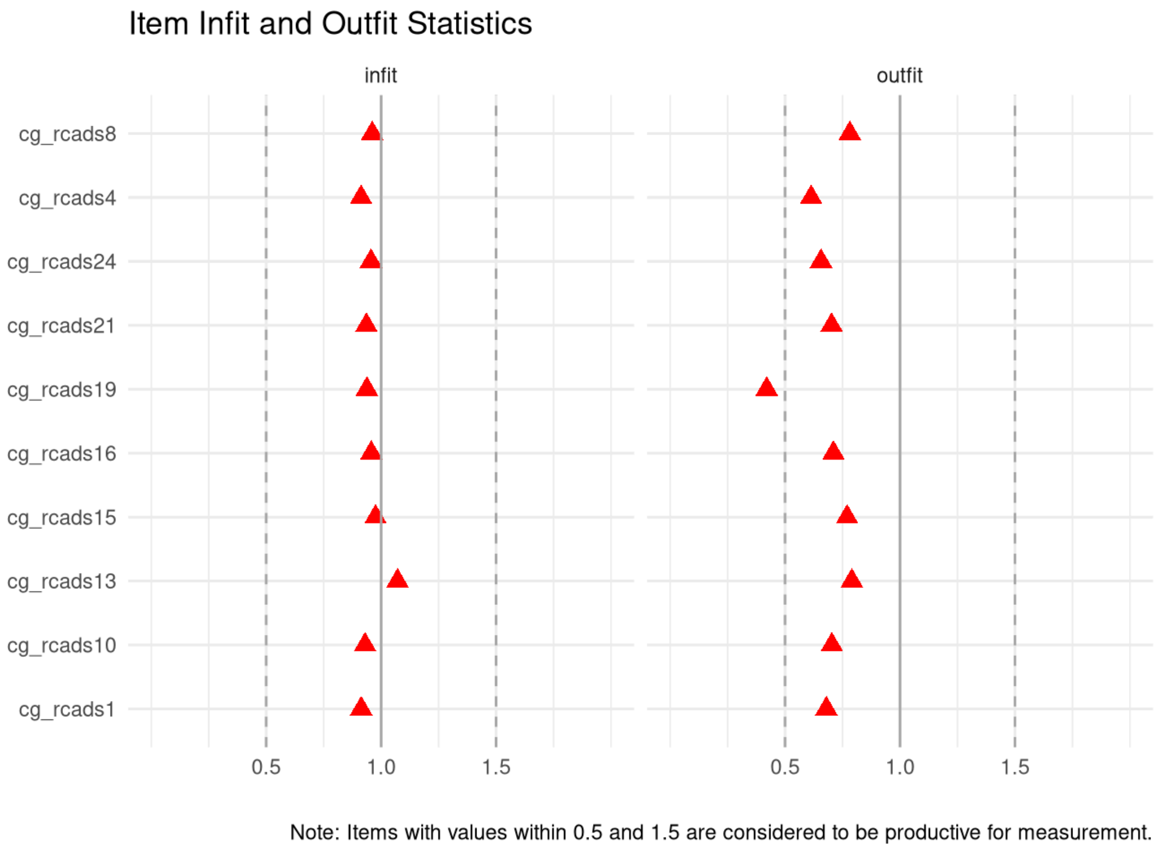

**Supplementary Figure 8.2.4 -** Revised Children's Anxiety and Depression Scale short-version (RCADS-25), Caregiver-report (Depression Scale): person infit and outfit statistics

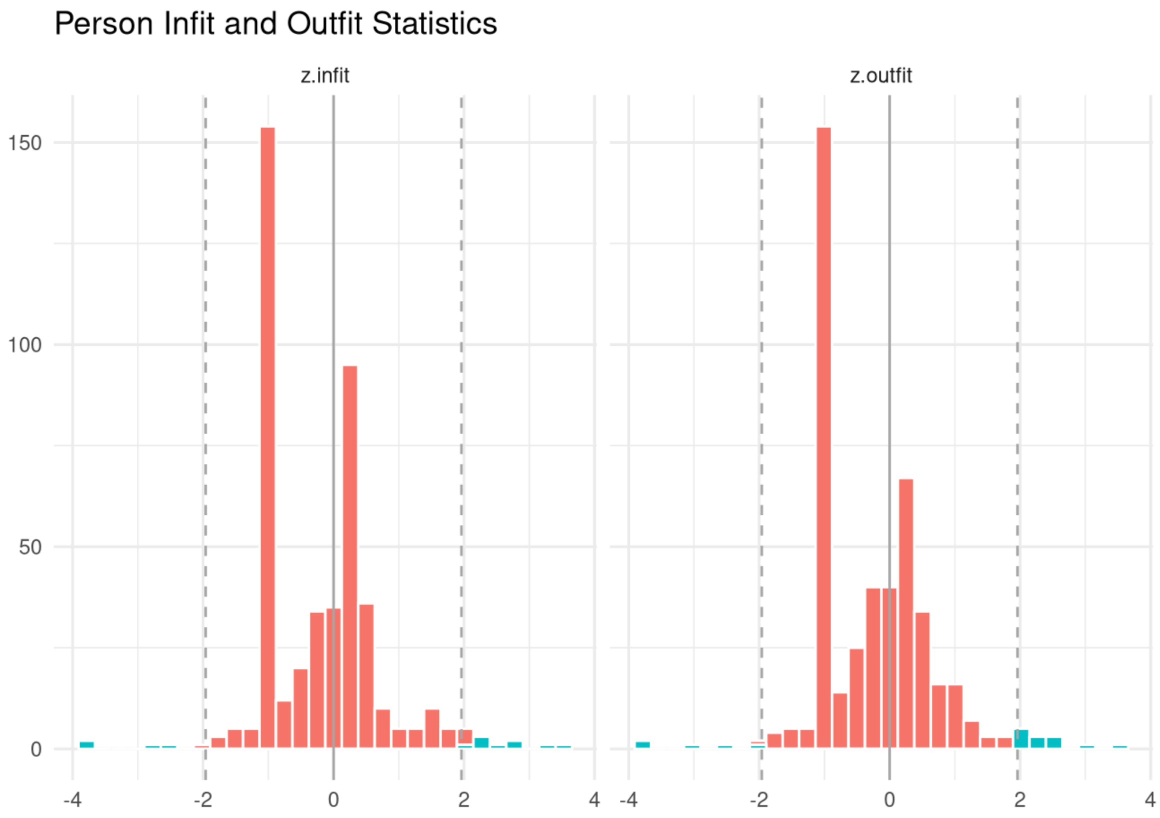

**Supplementary Figure 9.1.1 -** Revised Children's Anxiety and Depression Scale short-version (RCADS-25), Self-report (Anxiety Scale): test information and expected scores

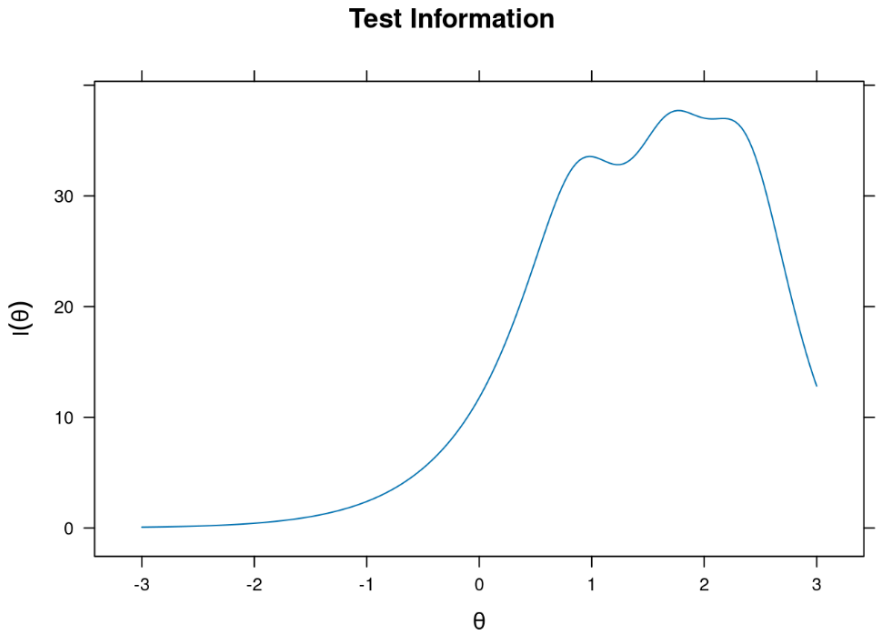

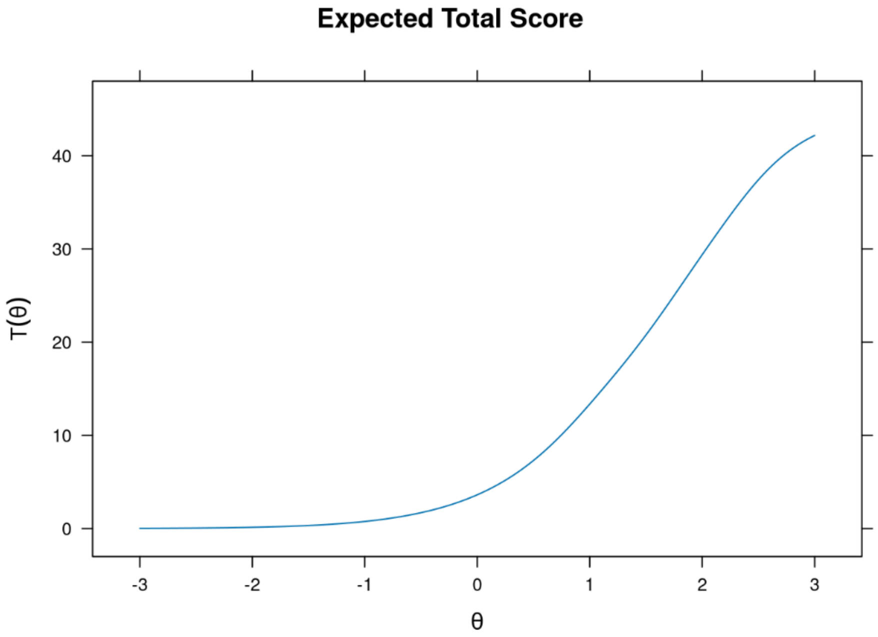

**Supplementary Figure 9.1.2 -** Revised Children's Anxiety and Depression Scale short-version (RCADS-25), Self-report (Anxiety Scale): item probability functions

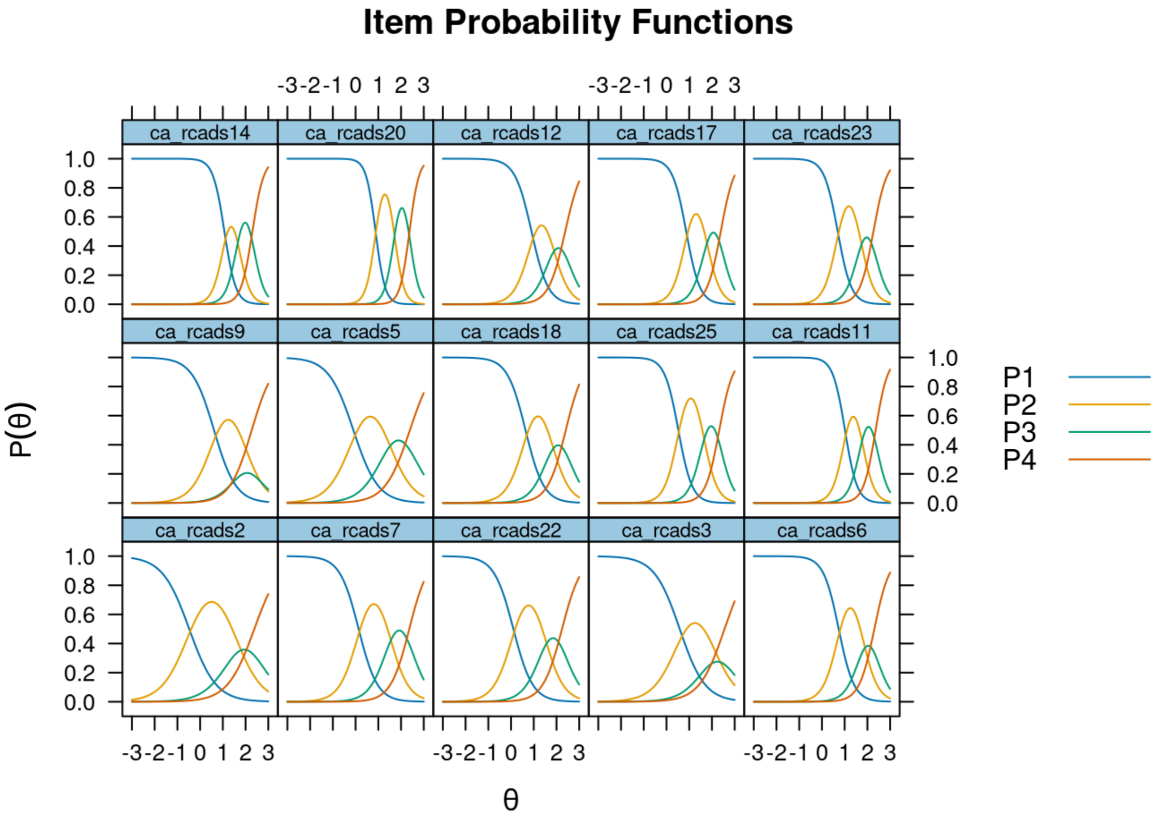

**Supplementary Figure 9.1.3 -** Revised Children's Anxiety and Depression Scale short-version (RCADS-25), Self-report (Anxiety Scale): item infit and outfit statistics

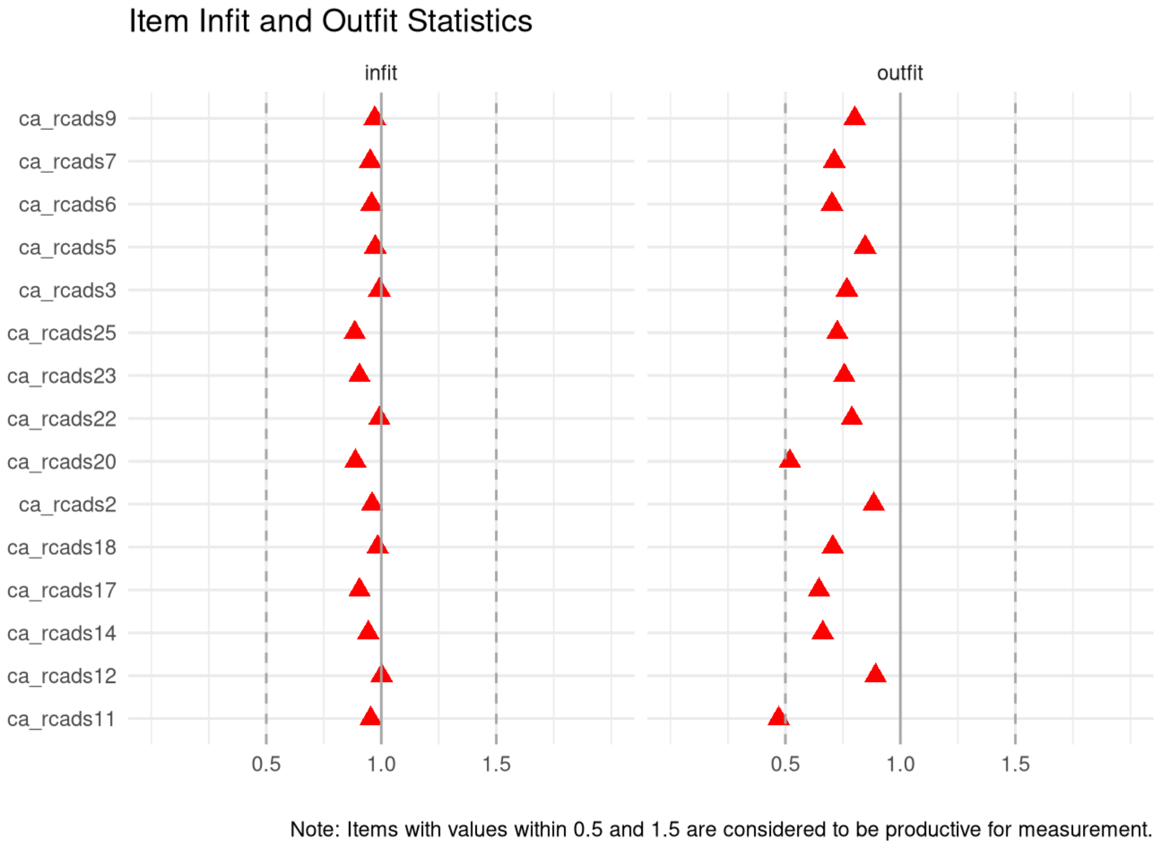

**Supplementary Figure 9.1.4 -** Revised Children's Anxiety and Depression Scale short-version (RCADS-25), Self-report (Anxiety Scale): person infit and outfit statistics

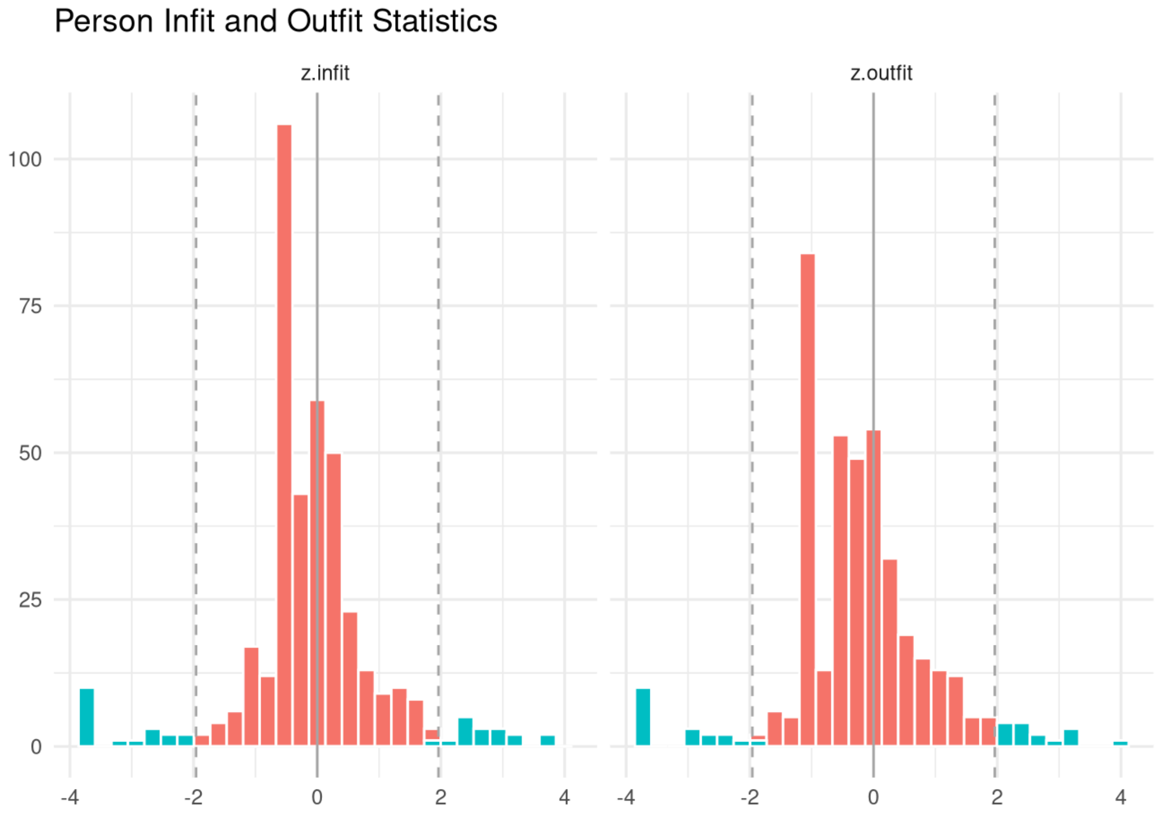

**Supplementary Figure 9.2.1 -** Revised Children's Anxiety and Depression Scale short-version (RCADS-25), Self-report (Depression Scale): test information and expected scores

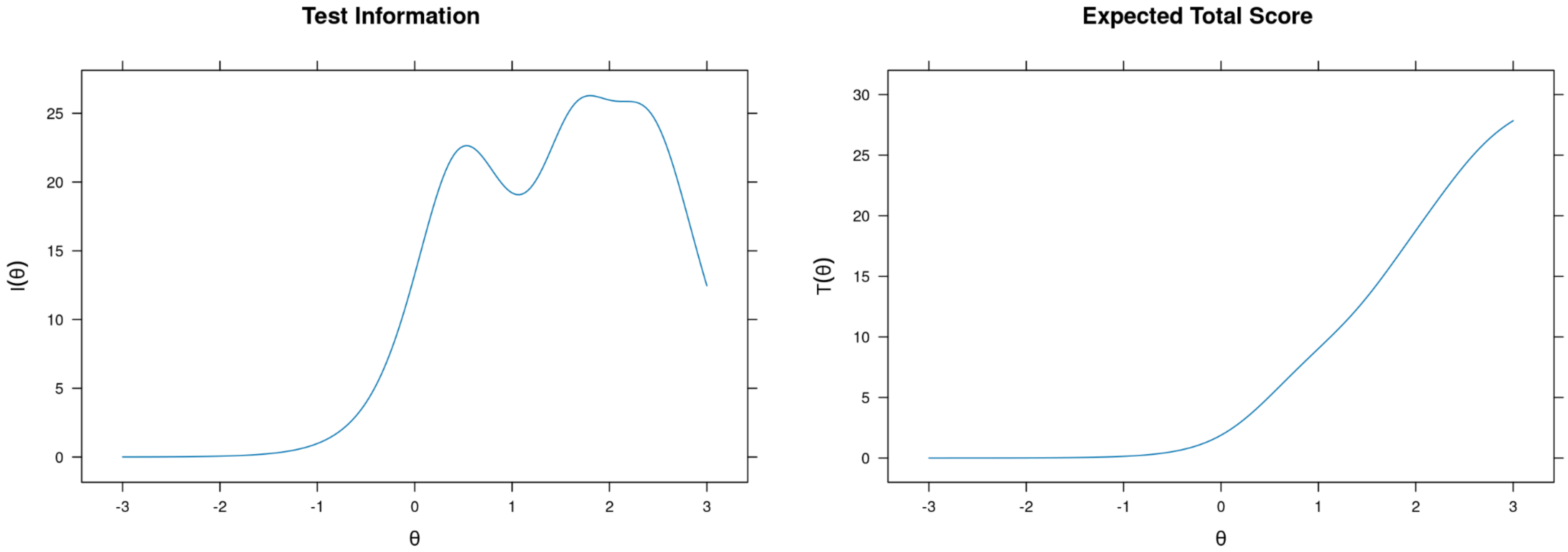

**Supplementary Figure 9.2.2 -** Revised Children's Anxiety and Depression Scale short-version (RCADS-25), Self-report (Depression Scale): item probability functions

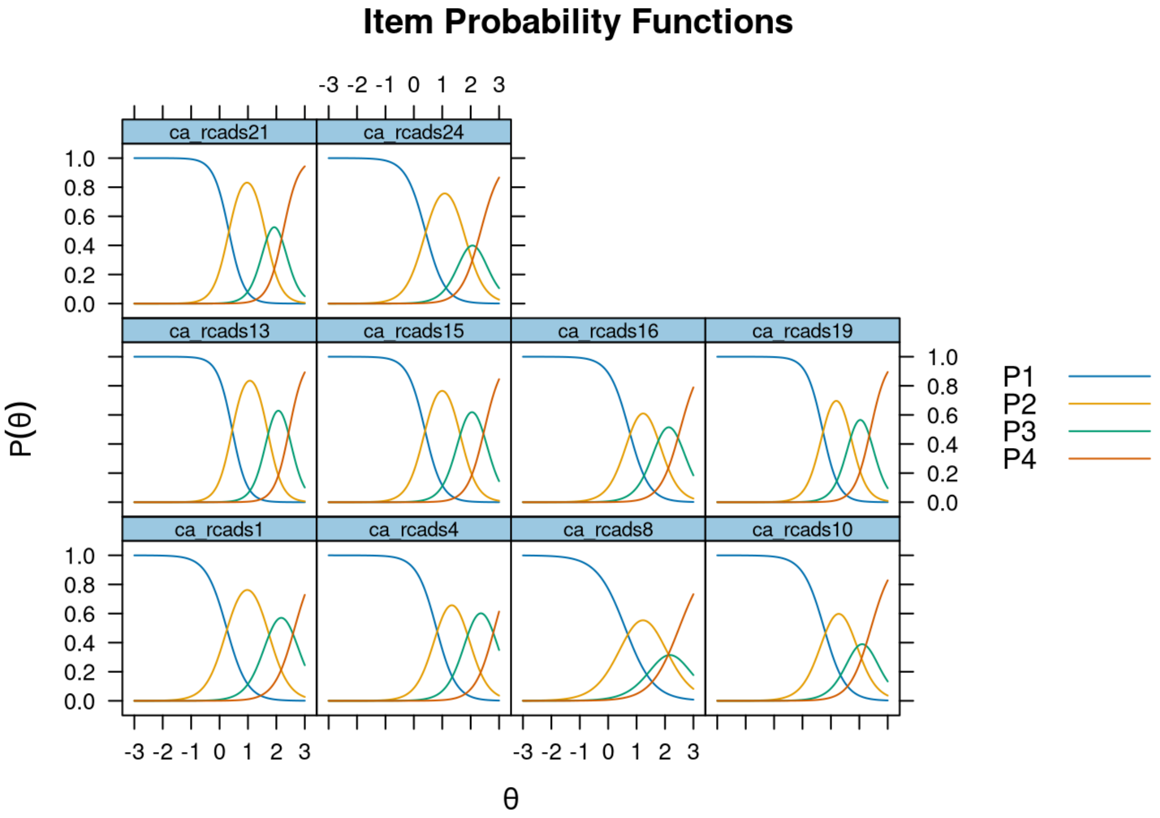

**Supplementary Figure 9.2.3 -** Revised Children's Anxiety and Depression Scale short-version (RCADS-25), Self-report (Depression Scale): item infit and outfit statistics

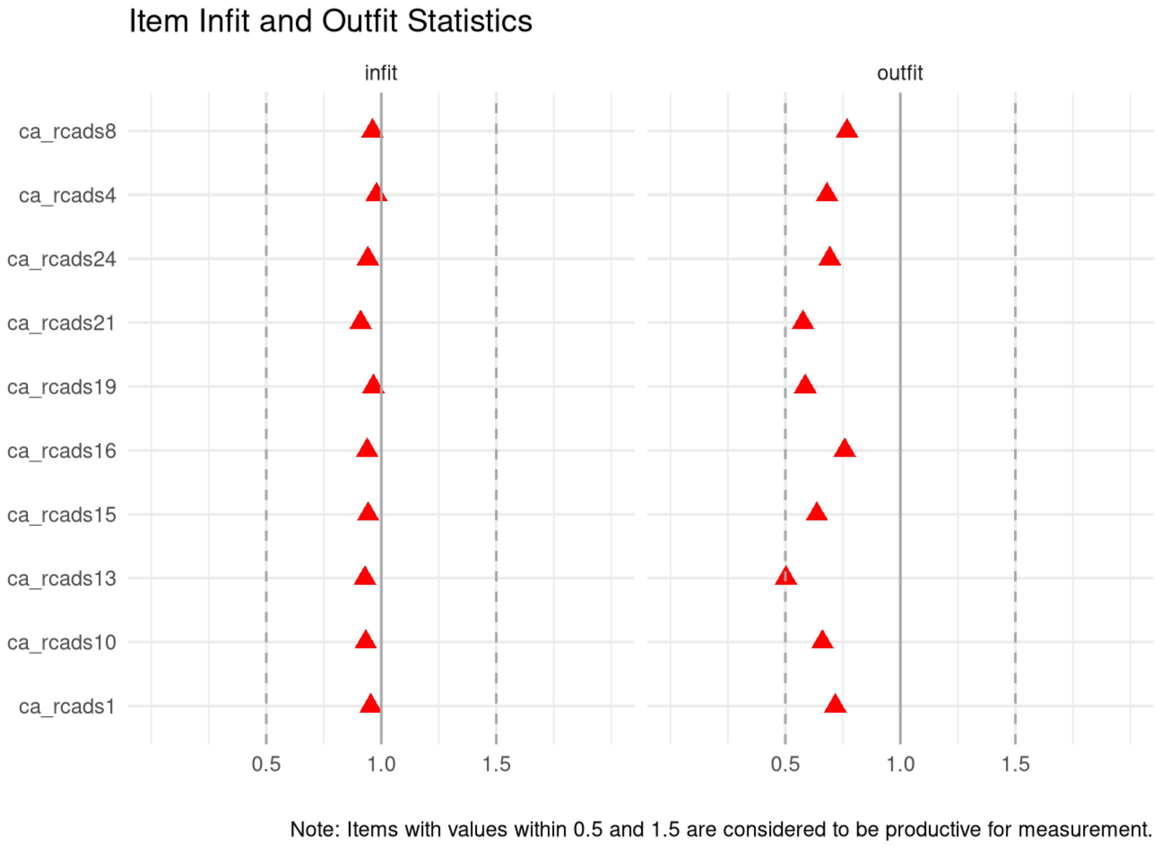

**Supplementary Figure 9.2.4 -** Revised Children's Anxiety and Depression Scale short-version (RCADS-25), Self-report (Depression Scale): person infit and outfit statistics

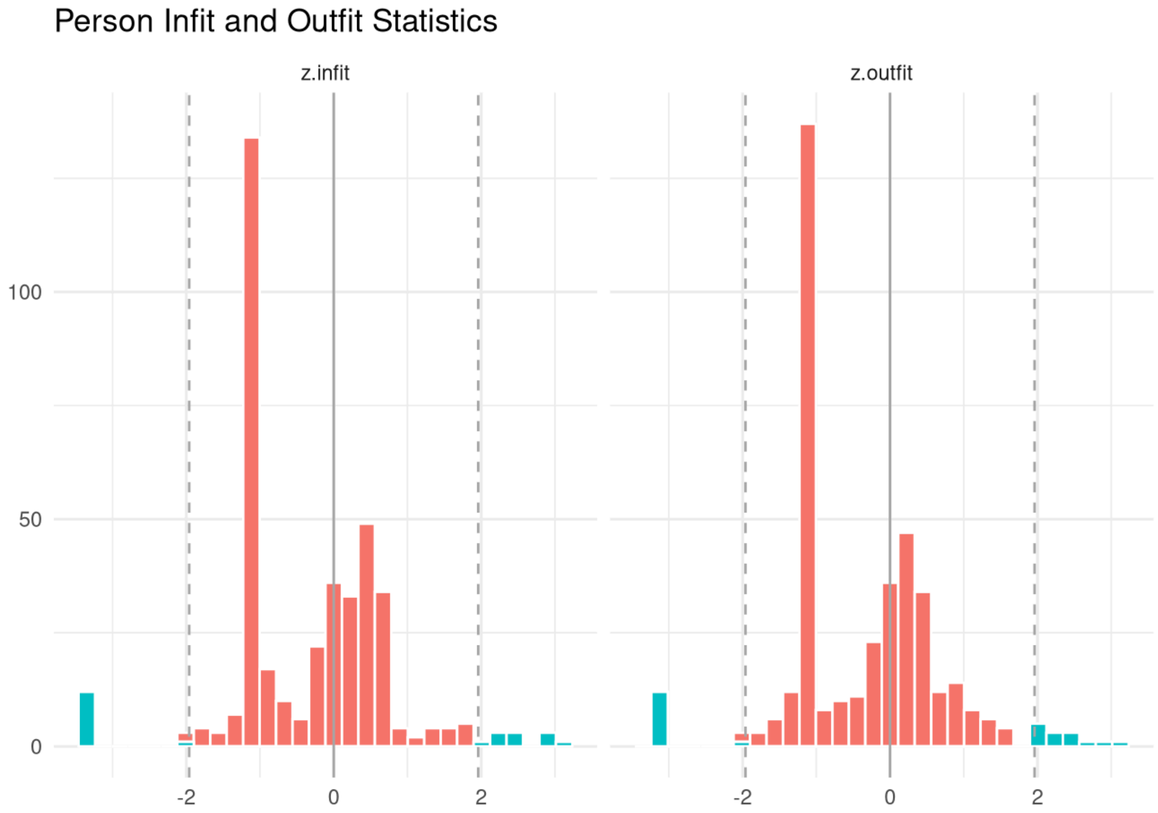

**Supplementary Figure 10.1.1 -** Swanson, Nolan and Pelham Scale (SNAP-IV), Caregiver-report (Hyperactivity Scale): test information and expected scores

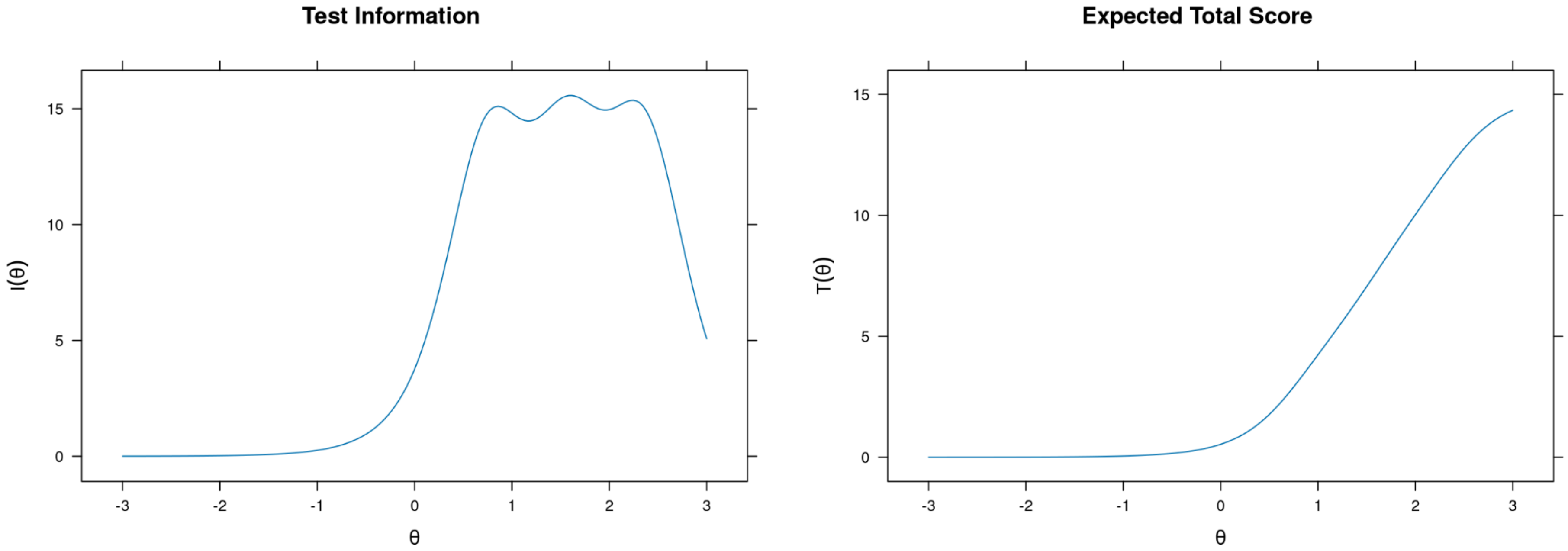

**Supplementary Figure 10.1.2 -** Swanson, Nolan and Pelham Scale (SNAP-IV), Caregiver-report (Hyperactivity Scale): item probability functions

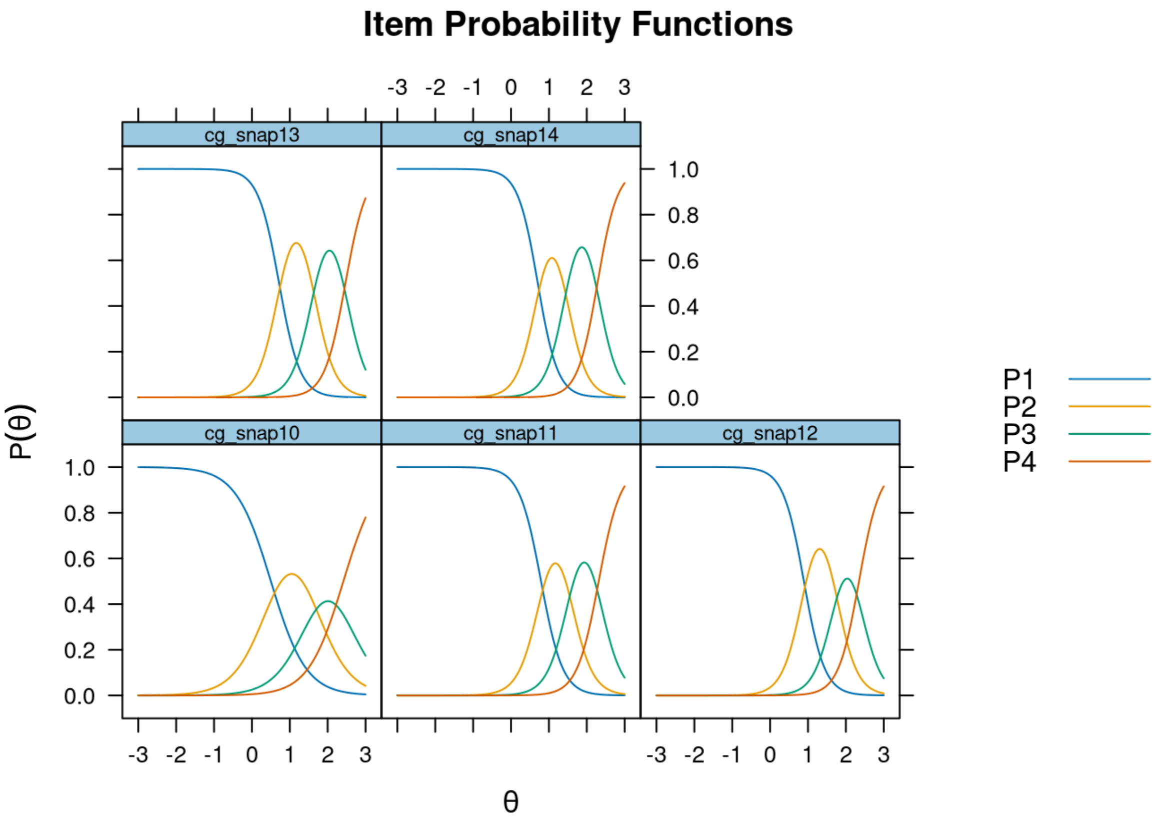

**Supplementary Figure 10.1.3 -** Swanson, Nolan and Pelham Scale (SNAP-IV), Caregiver-report (Hyperactivity Scale): item infit and outfit statistics

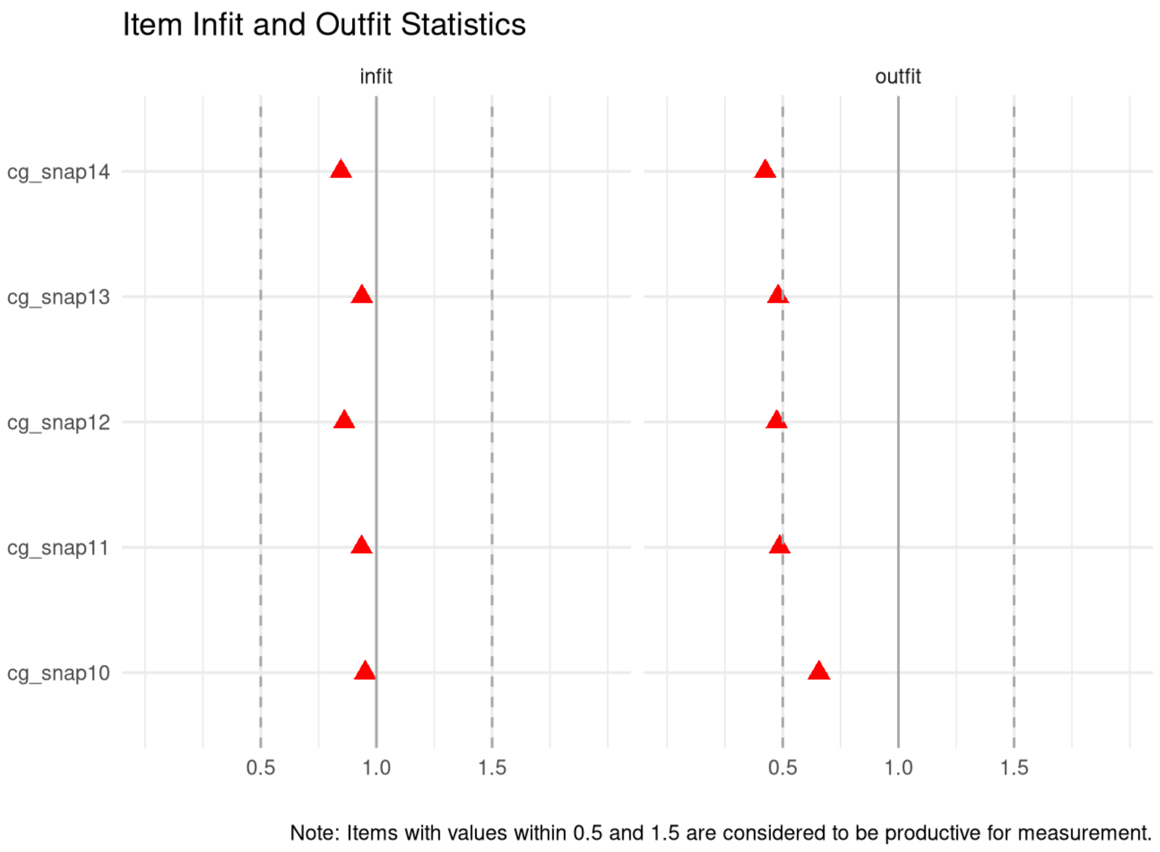

**Supplementary Figure 10.1.4 -** Swanson, Nolan and Pelham Scale (SNAP-IV), Caregiver-report (Hyperactivity Scale): person infit and outfit statistics

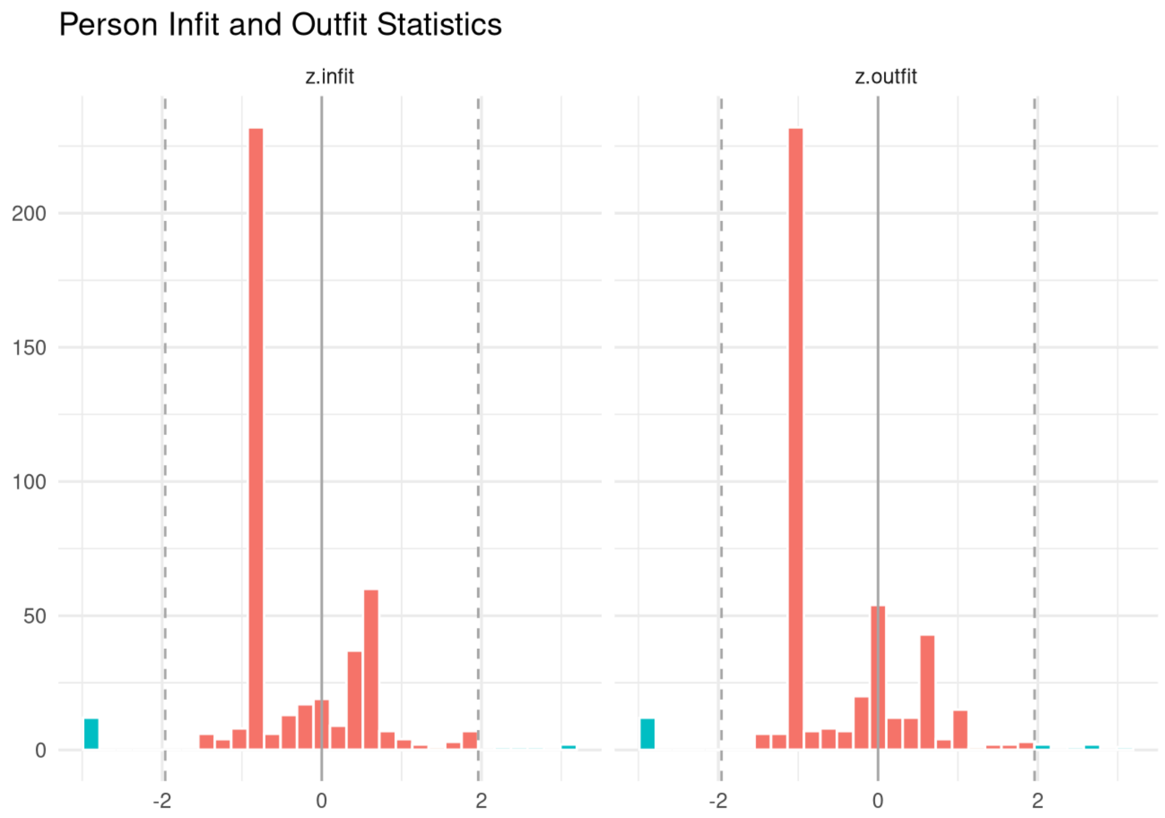

**Supplementary Figure 10.2.1 -** Swanson, Nolan and Pelham Scale (SNAP-IV), Caregiver-report (Impulsivity Scale): test information and expected score

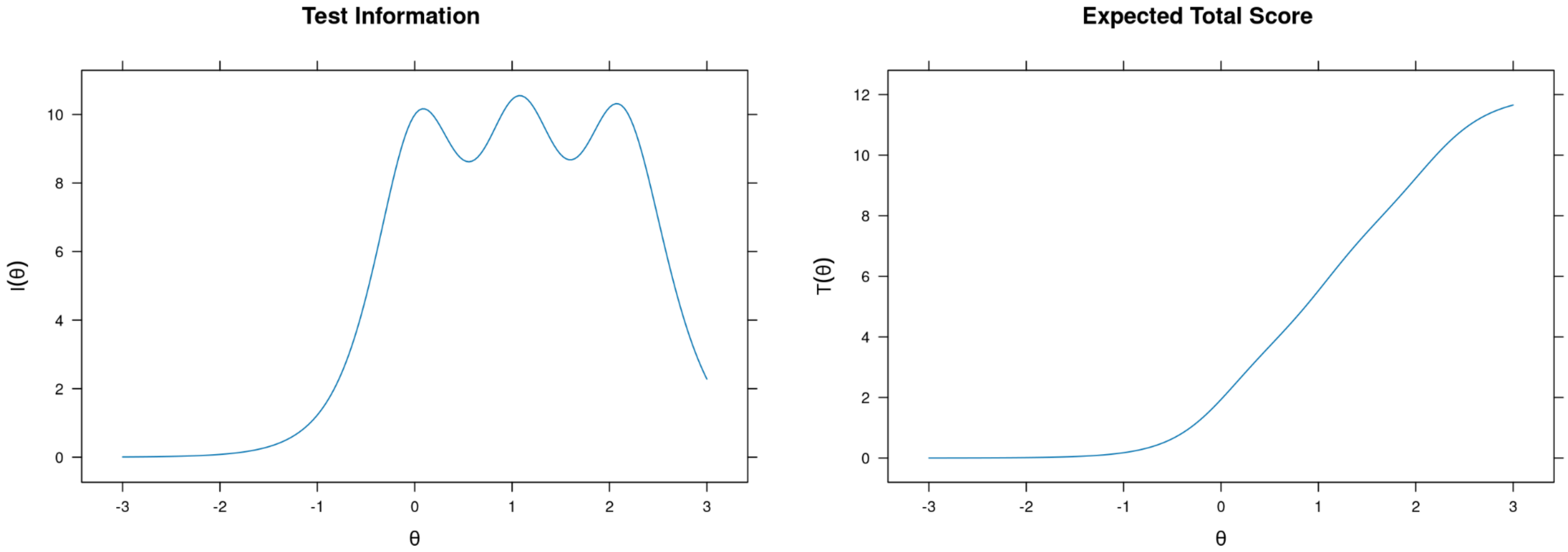

**Supplementary Figure 10.2.2 -** Swanson, Nolan and Pelham Scale (SNAP-IV), Caregiver-report (Impulsivity Scale): item probability functions

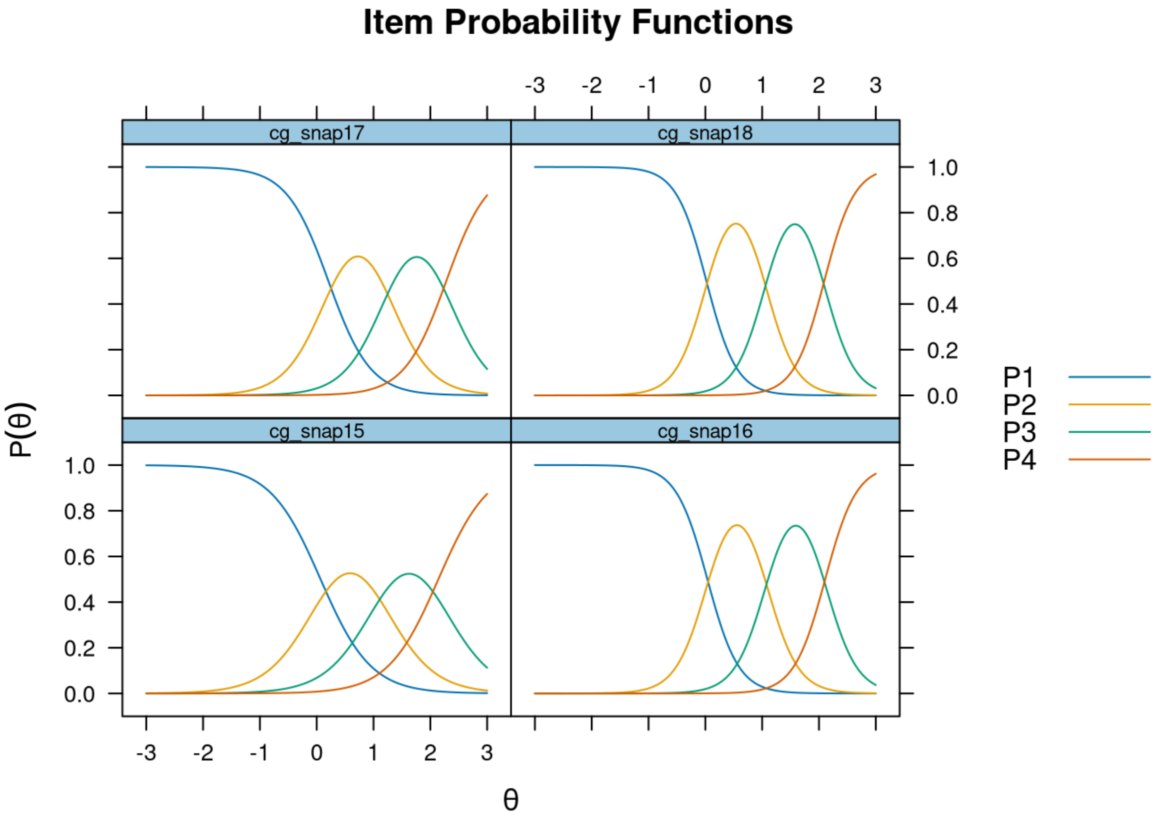

**Supplementary Figure 10.2.3 -** Swanson, Nolan and Pelham Scale (SNAP-IV), Caregiver-report (Impulsivity Scale): item infit and outfit statistics

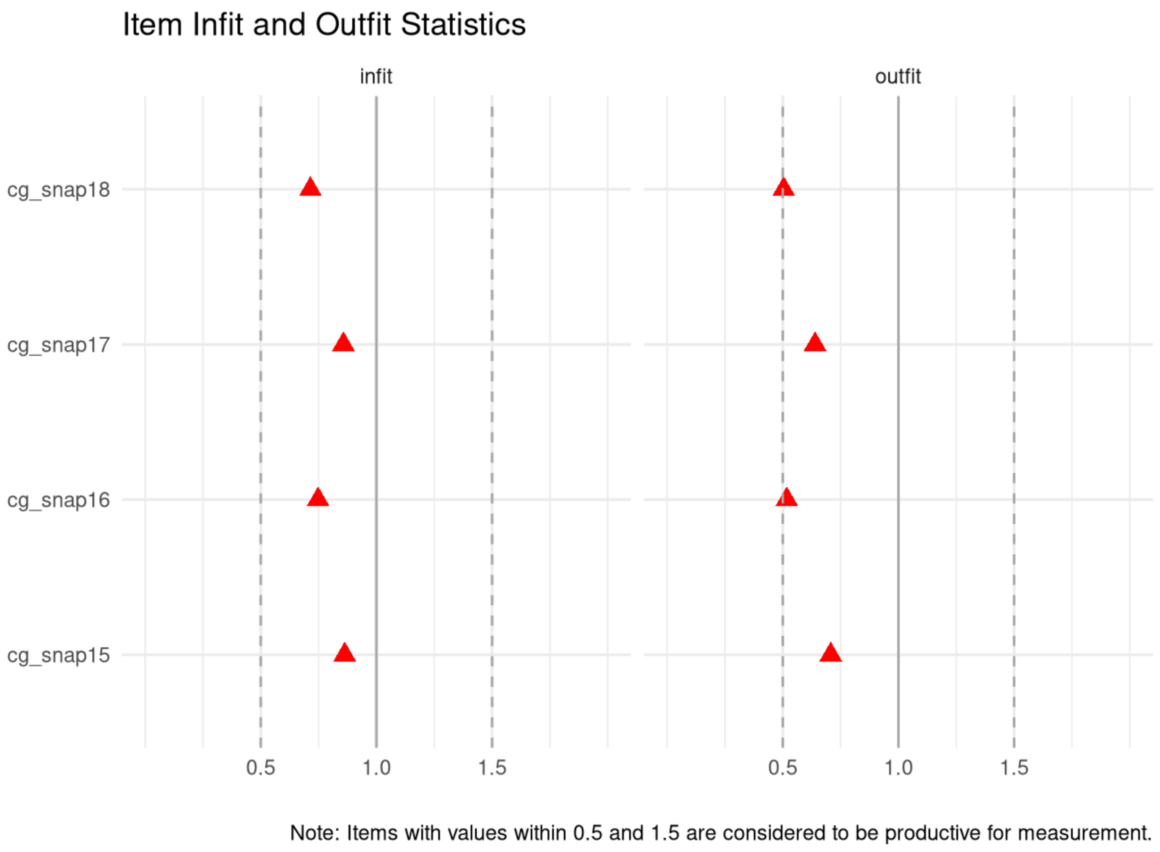

**Supplementary Figure 10.2.4** - Swanson, Nolan and Pelham Scale (SNAP-IV), Caregiver-report (Impulsivity Scale): person infit and outfit statistics

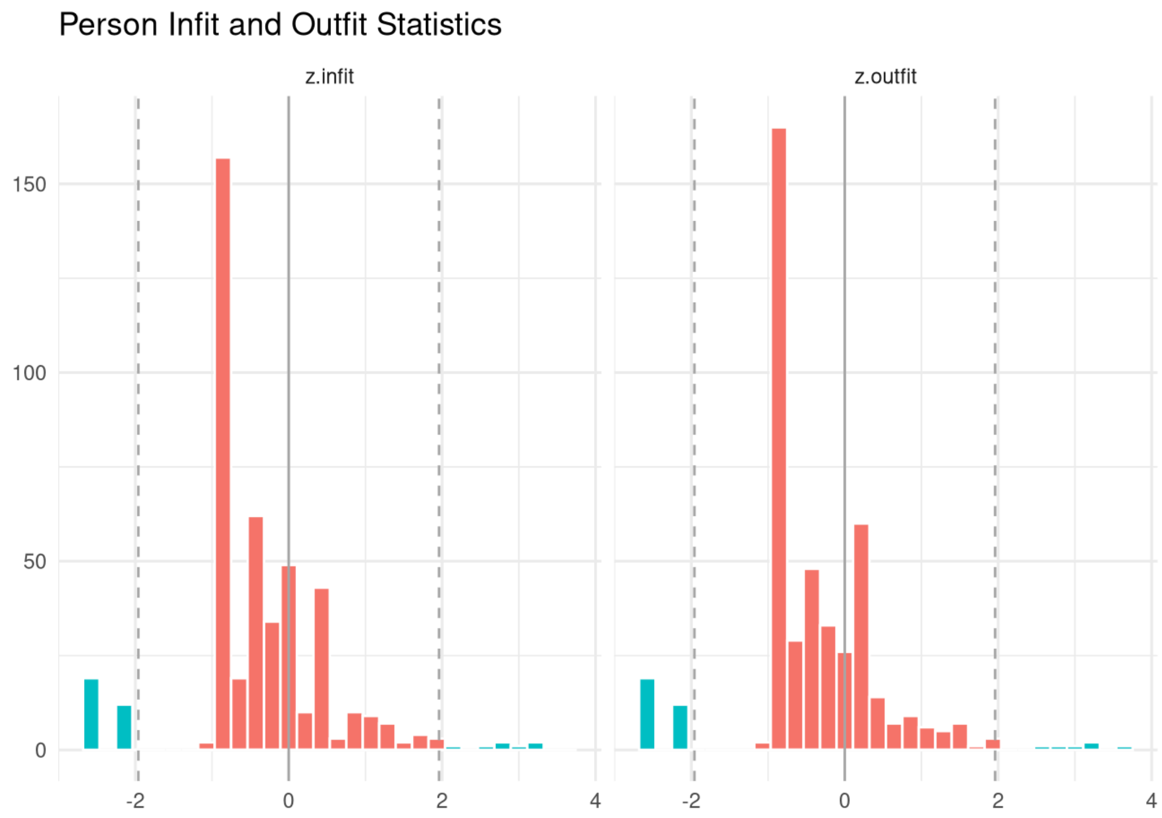

**Supplementary Figure 10.3.1 -** Swanson, Nolan and Pelham Scale (SNAP-IV), Caregiver-report (Inattention Scale): test information and expected scores

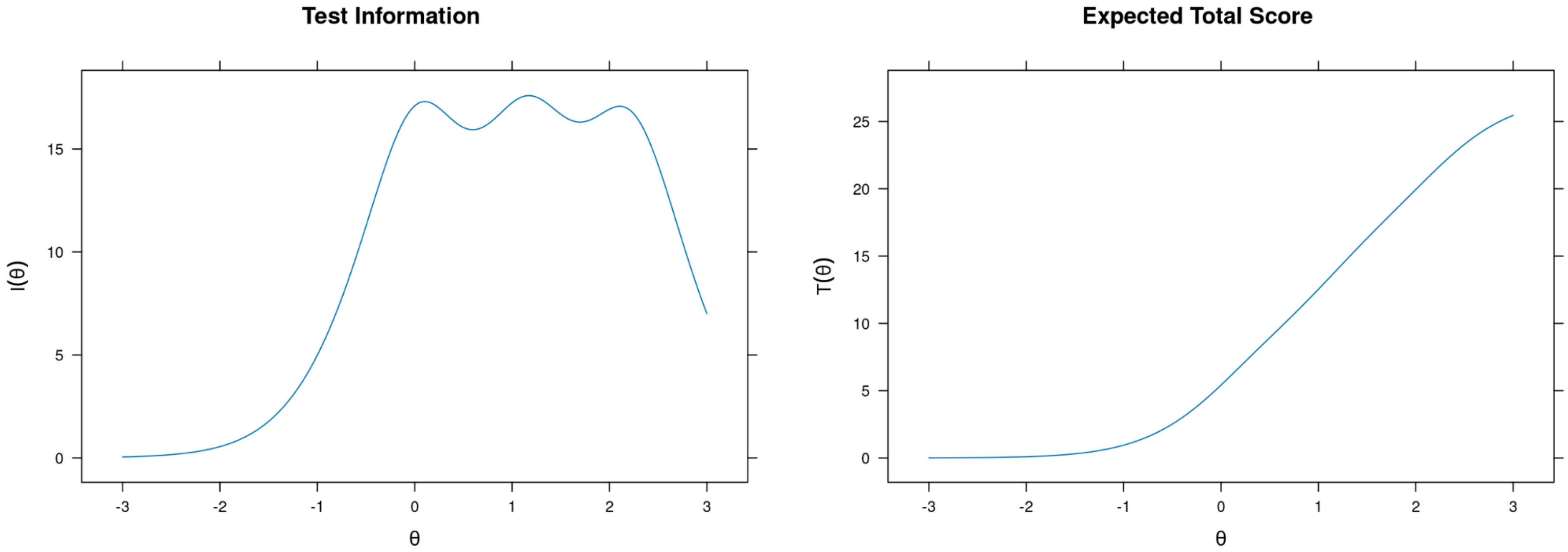

**Supplementary Figure 10.3.2 -** Swanson, Nolan and Pelham Scale (SNAP-IV), Caregiver-report (Inattention Scale): item probability functions

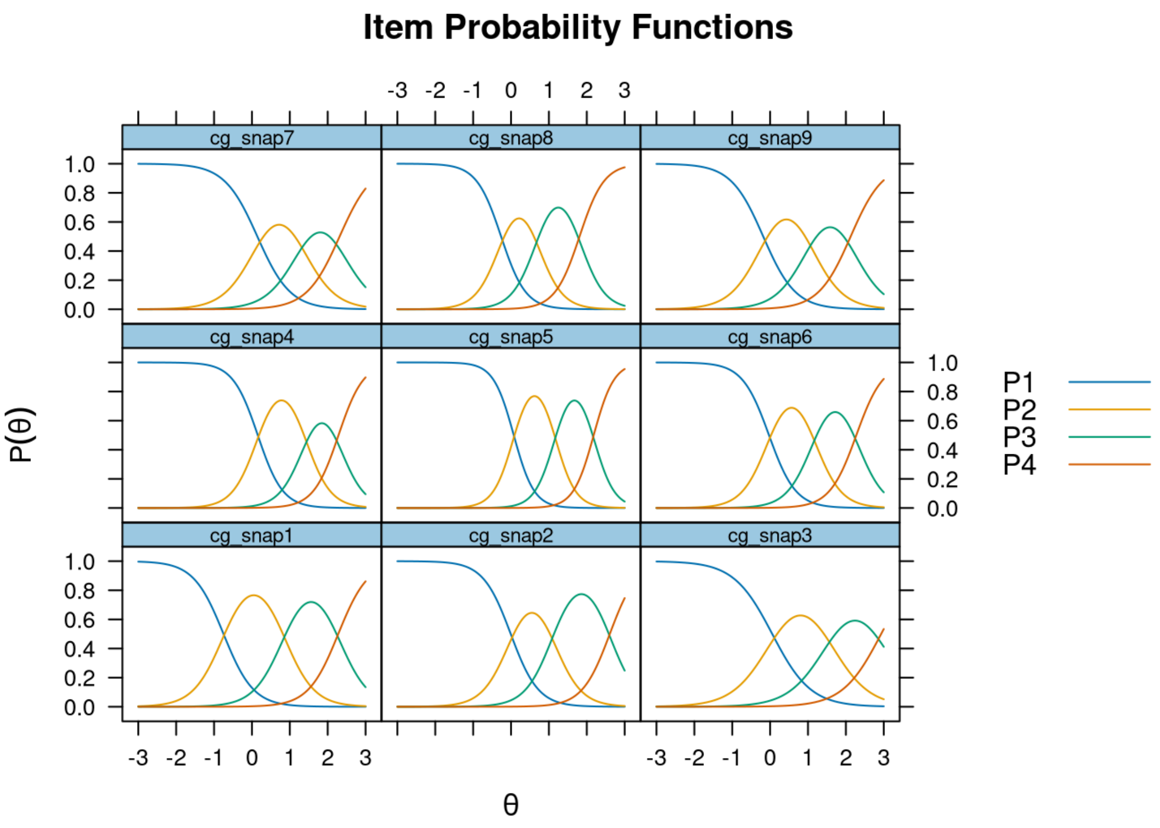

**Supplementary Figure 10.3.3 -** Swanson, Nolan and Pelham Scale (SNAP-IV), Caregiver-report (Inattention Scale): item infit and outfit statistics

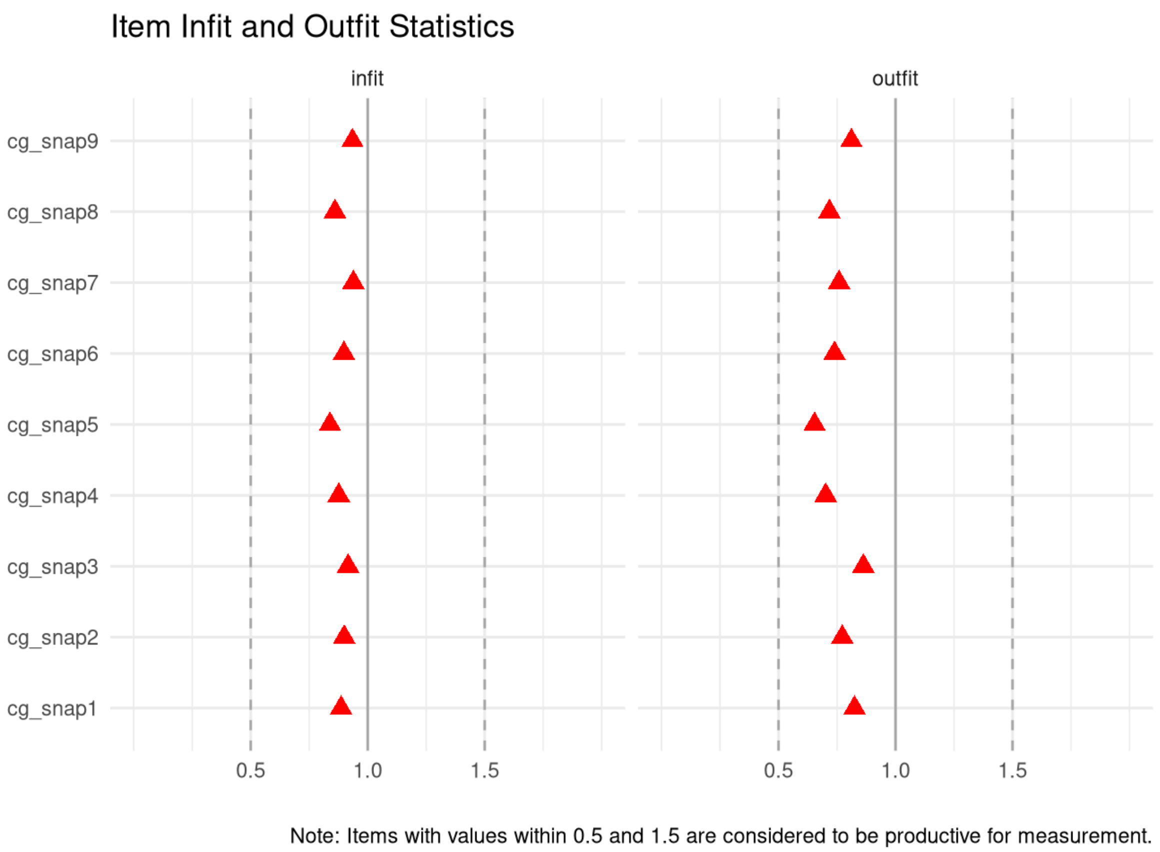

**Supplementary Figure 10.3.4 -** Swanson, Nolan and Pelham Scale (SNAP-IV), Caregiver-report (Inattention Scale): person infit and outfit statistics

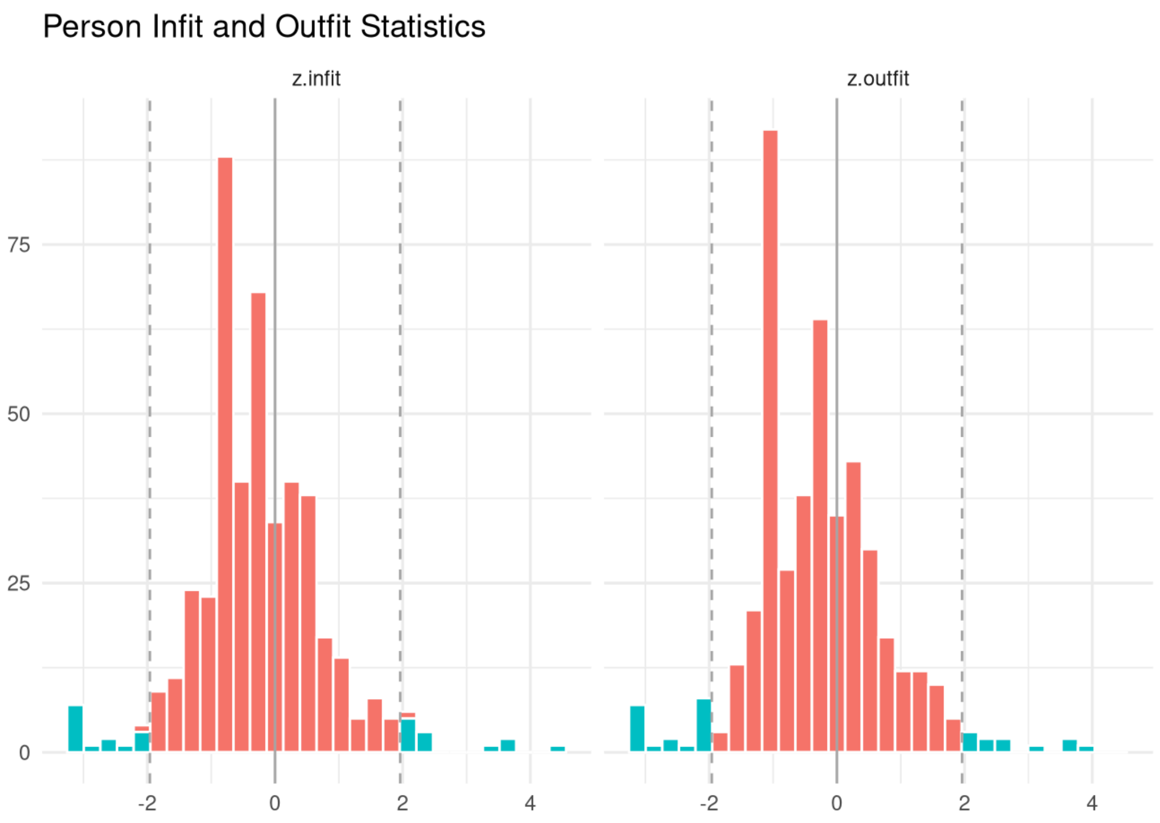

**Supplementary Figure 10.4.1 -** Swanson, Nolan and Pelham Scale (SNAP-IV), Caregiver-report (Oppositionality Scale): test information and expected scores

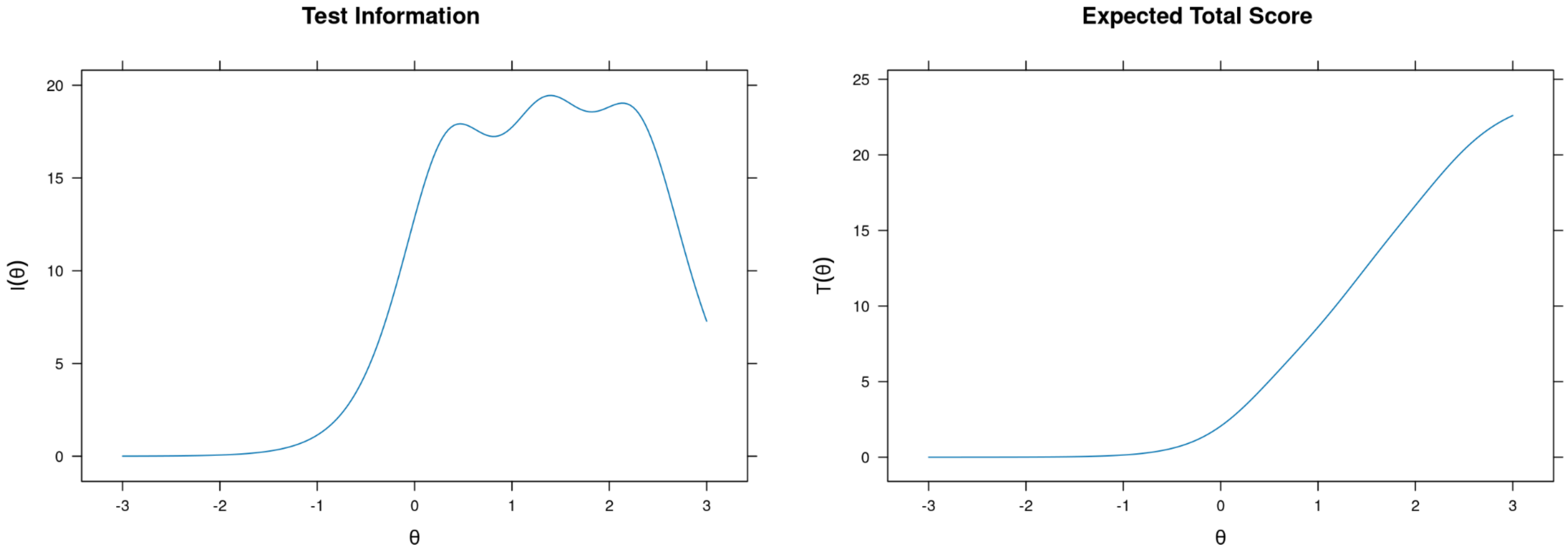

**Supplementary Figure 10.4.2 -** Swanson, Nolan and Pelham Scale (SNAP-IV), Caregiver-report (Oppositionality Scale): item probability functions

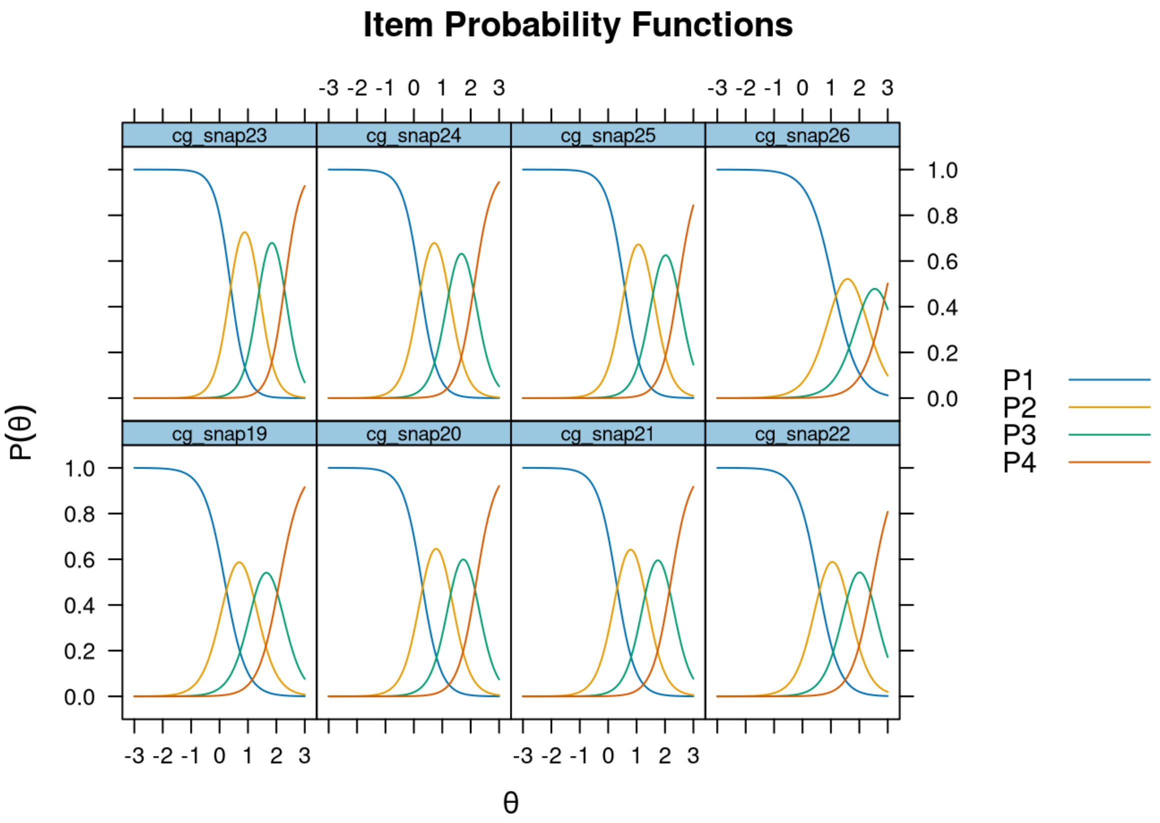

**Supplementary Figure 10.4.3 -** Swanson, Nolan and Pelham Scale (SNAP-IV), Caregiver-report (Oppositionality Scale): item infit and outfit statistics

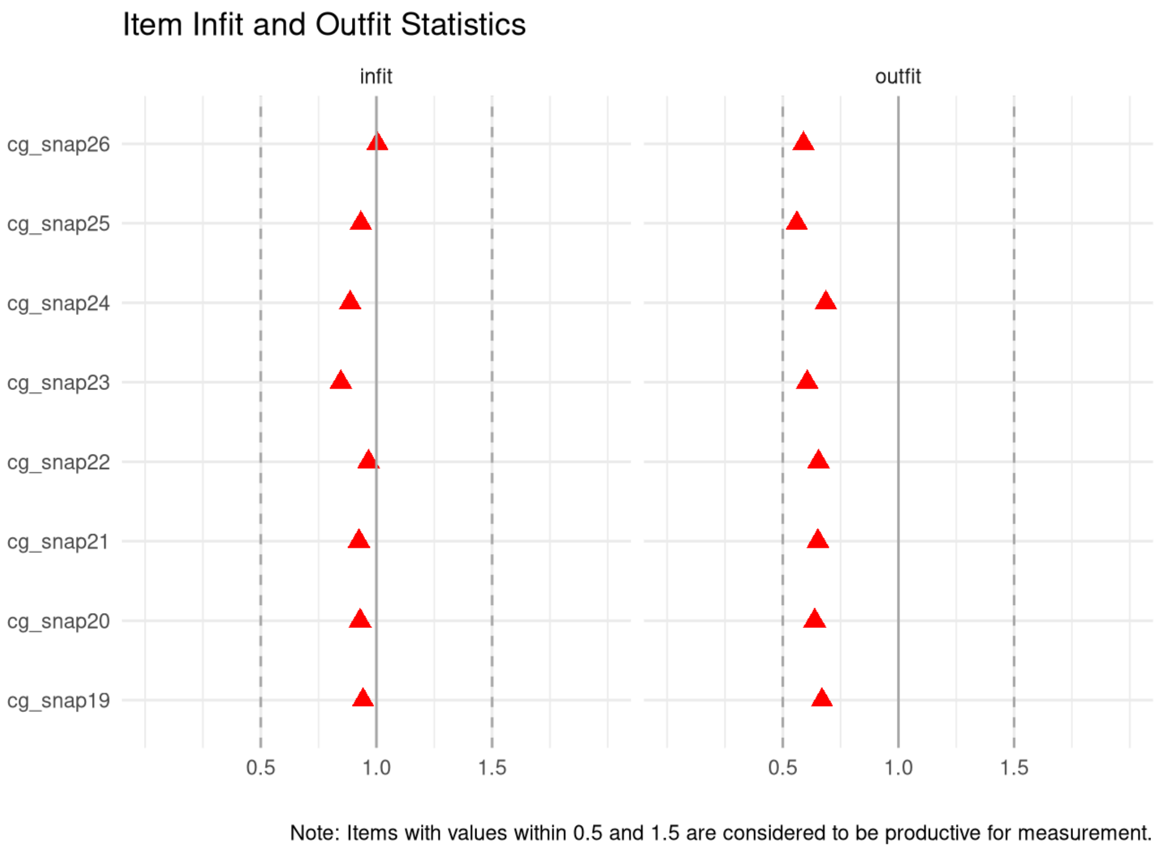

**Supplementary Figure 10.4.4 -** Swanson, Nolan and Pelham Scale (SNAP-IV), Caregiver-report (Oppositionality Scale): person infit and outfit statistics

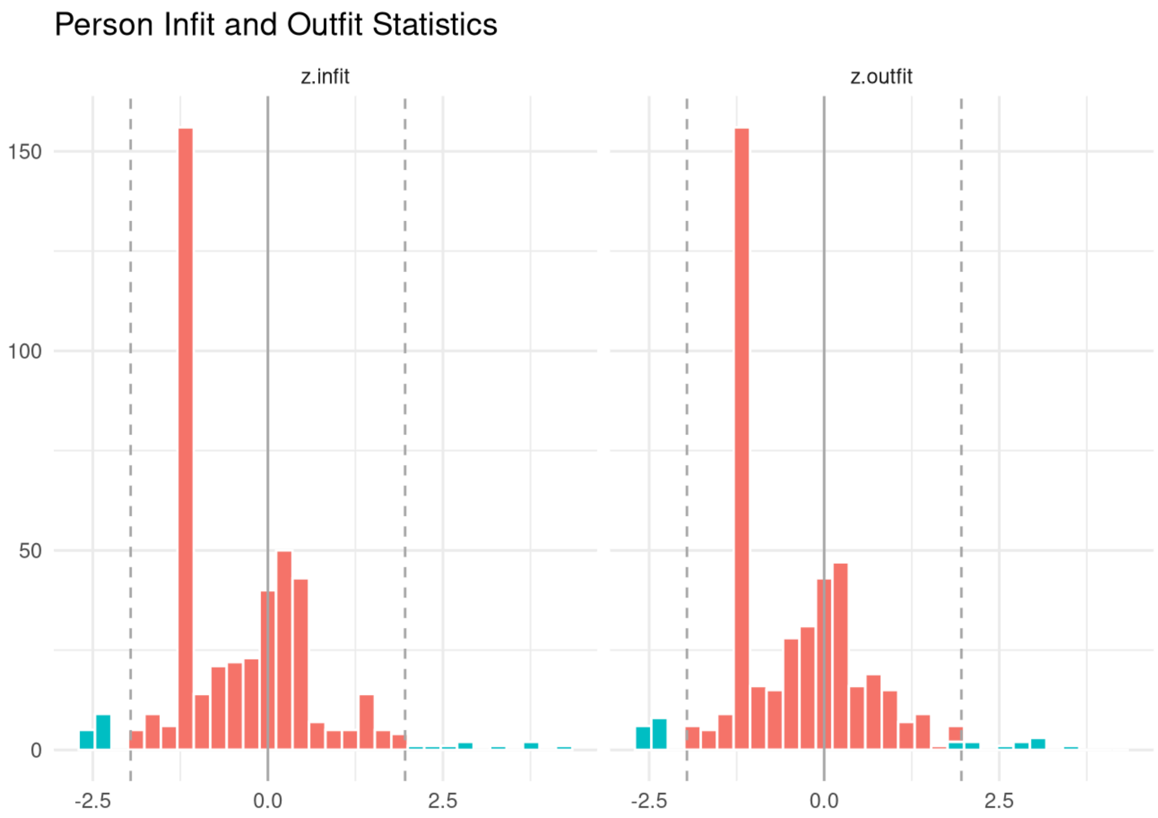
